# Inequalities in mental and social wellbeing during the COVID-19 pandemic: prospective longitudinal observational study of five UK cohorts

**DOI:** 10.1101/2022.02.07.22270588

**Authors:** Darío Moreno-Agostino, Helen L. Fisher, Stephani L. Hatch, Craig Morgan, George B. Ploubidis, Jayati Das-Munshi

## Abstract

**Background:** Research suggests that there have been inequalities in the impact of the COVID-19 pandemic and related non-pharmaceutical interventions on population mental health. We explored these inequalities during the first year of the pandemic using nationally representative cohorts from the UK.

**Methods:** We analysed data from 26,772 participants from five longitudinal cohorts representing generations born between 1946 and 2000, collected in May 2020, September-October 2020, and February-March 2021 across all five cohorts. We used a multilevel growth curve modelling approach to explore sociodemographic and socioeconomic differences in levels of anxiety and depressive symptomatology, loneliness, and life satisfaction over time.

**Results:** Younger generations had worse levels of mental and social wellbeing throughout the first year of the pandemic. Whereas these generational inequalities narrowed between the first and last observation periods for life satisfaction (−0.33 [95% CI: −0.51, −0.15]), they became larger for anxiety (0.22 [0.10, 0.33]). Pre-existing generational inequalities in depression and loneliness did not change, but initial depression levels of the youngest cohort were worse than expected if the generational inequalities had not accelerated. Women and those experiencing financial difficulties had worse initial mental and social wellbeing levels than men and those financially living comfortably, respectively, and these gaps did not substantially differ between the first and last observation periods. Inequalities by additional factors are reported.

**Conclusions:** By March 2021, mental and social wellbeing inequalities persisted in the UK adult population. Pre-existing generational inequalities may have been exacerbated with the pandemic onset. Policies aimed at protecting vulnerable groups are needed.

## Introduction

Evidence from the initial months of the COVID-19 pandemic suggests that the impact of its onset and of the measures to control its spread have been substantially different not only across different measures of mental and social wellbeing, but also across social groups, contexts, and countries.^1,2^ A systematic review of 117 studies from 28 different countries found that, among the most usually reported inequality factors, women, younger people, and those in more disadvantaged socioeconomic situations generally had worse mental and social wellbeing levels in the initial stages of the pandemic.^2^

This is consistent with findings from the UK, where longitudinal evidence comparing mental health outcomes before and after the introduction of the first nationwide lockdown measures on 23 March 2020, has shown that overall levels of distress and anxiety increased in the population, with younger people, women, those in worse financial situations, and those with pre-existing mental health conditions being disproportionately impacted.^3–7^ Differences by region and urbanicity have also been reported, with higher levels of distress reported as more likely in some specific areas such as London or among urban dwellers.^3^ Although depression levels seemed to remain stable in the adult population compared with levels before the pandemic onset,^6^ an increase was found among women aged 50+,^8^ thus pointing at a potential interplay between generational and gender inequalities.

Further studies have focused on monitoring the changes in mental and social wellbeing outcomes during the first months of the pandemic, as control measures were gradually eased.^9^ Stable or improving levels of anxiety and depressive symptomatology, loneliness, and subjective wellbeing were found across the first months after the first lockdown and up to July/August 2020.^10–12^ Yet again, younger people, women, and those in worse socioeconomic situations, as well as people from minority ethnic groups, displayed worse levels or trajectories over time,^10–13^ and even if improvements in depressive and anxiety symptomatology were found to be steeper among some of these disadvantaged subgroups, inequalities were still evident at the end of the study period.^11^

As restrictions were reintroduced around October/November 2020, studies monitoring the UK adult population distress levels have found heterogeneous trajectories across age groups and genders, with women seemingly more impacted by the restrictions.^7,14,15^ Moreover, subgroups within the population with similar distress trajectories have been found using evidence up to May 2021, with younger people, women, those in a worse financial situation, not in a relationship, and those from minority ethnic groups being more likely to show increasing or consistently higher distress trajectories.^15,16^

In summary, the available literature shows the existence of mental and social wellbeing inequalities across generational, gender, and socioeconomic subgroups within the UK population. However, most of this evidence refers to distress levels, with fewer providing evidence on finer grained outcomes such as anxiety or depressive symptomatology, and even fewer on other relevant mental and social wellbeing outcomes such as loneliness or life satisfaction. Moreover, even if inequalities are reported at the early stages of the pandemic, in most cases it remains unknown whether those inequalities have changed over time. Finally, there is very limited evidence on the potential interplay of combined inequality factors on those initial levels or rates of change (e.g., combined generational and sex inequalities).^2^ Hence, this study aims to explore inequalities within the population in the initial levels and rates of change of a wide range of mental and social wellbeing measures during the first year of the COVID-19 pandemic, studying the interplay between generational and other inequality factors.

## Methods

### Sample and procedure

We used the data from the COVID-19 survey (https://cls.ucl.ac.uk/covid-19-survey/) conducted with participants from five UK cohorts representing different generations: National Survey of Health and Development (NSHD, 1946 cohort),^17^ National Child Development Study (NCDS, 1958 cohort),^18^ British Cohort Study (BCS, 1970 cohort),^19^ Next Steps (NS, 1990 cohort),^20^ and Millennium Cohort Study (MCS, 2000-02 cohort).^21^ Detailed information on these cohorts and their designs is available in https://cls.ucl.ac.uk/ for NCDS/1958, BCS/1970, NS/1990, and MCS/2000-02; and in https://www.nshd.mrc.ac.uk/ for NSHD/1946. The COVID-19 survey was designed to collect relevant information around the pandemic impact on the cohort members. Data were collected at three time points: May 2020 (survey wave 1, during the first national lockdown), September-October 2020 (survey wave 2, between the first and second national lockdowns), and February-March 2021 (survey wave 3, during the third national lockdown).^9^ Data collection took place via web interviews, supplemented by telephone interviews in survey wave 3. In this study, we focused on those individuals currently alive and residing in the UK (**Appendix S1**, Supplementary Material). In MCS, only the data from the main cohort members were included, despite in some cases more than one family member (other sibling/s or parent/s) participating in the survey. Overall response rates with respect to the target populations ranged from 20.8% (survey wave 1) to 31.2% (survey wave 3). Non-response weights were used to restore sample representativeness (**Appendix S2**, Supplementary Material). The COVID-19 Survey was approved by the National Health Service (NHS) Research Ethics Committee, and all participants provided informed consent.

### Measures

A set of common instruments assessing multiple mental and social wellbeing outcomes were used across all cohorts in the COVID-19 surveys. Experiences indicative of anxiety and depression were measured with the 2-item General Anxiety Disorder (GAD-2)^22^ and the 2-item Patient Health Questionnaire (PHQ-2),^23^ respectively. Each of these tools include two items on the frequency the respondent has been bothered by experiences of anxiety or depression over the previous two weeks, ranging from 0 (“Not at all”) to 3 (“Nearly every day”), which were summed, ranging from 0 (lowest anxiety/depression) to 6 (highest anxiety/depression). Loneliness was measured with the 3-item University of California Los Angeles (UCLA-3) loneliness scale,^24^ which includes three items on how frequently the respondents felt they lacked companionship, were left out, or were isolated from others, with three response options: 1 (“Hardly ever”), 2 (“Some of the time”), and 3 (“Often”). The total sum score ranged from 3 (lowest loneliness) to 9 (highest loneliness). Subjective wellbeing was measured with the Office for National Statistics single question on life satisfaction: “Overall, how satisfied are you with your life nowadays?”, with response options ranging from 0 (“Not at all”) to 10 (“Completely”).^25^

Inequalities in these measures were explored by cohort (NSHD, NCDS, BCS, NS, MCS) and additional demographic, socioeconomic, and geographical subgrouping variables. These included birth sex (man or woman); self-reported financial situation in the three months prior to the pandemic outbreak (“pre-pandemic financial situation” from here onwards, grouped into “Living comfortably”, “Doing all right”, or “Just about getting by”/“Finding it quite difficult”/“Finding it very difficult”); relationship status (in a relationship or not, regardless of cohabiting with the partner); housing tenure (house owned/partly owned or rented/rent-free/other arrangement); urbanicity (urban or rural dwelling) based on geographic information and classified according to the corresponding governmental recommendations;^26,27^ UK country of residence (England, Northern Ireland, Scotland, or Wales); and self-designated ethnicity grouped into White (all), Mixed, Indian/Pakistani/Bangladeshi, Black Caribbean/Black African, and Other (aggregated due to the small number of cases in some of the individual groups, including all ethnicities not captured by the previous categories).^28^ Information on ethnicity was only available in the two youngest cohorts (NS and MCS) and corresponded to the most recent self-designated ethnicity. This was complemented by the parents’ report in MCS participants in those cases where self-designated information was not available. The earliest available information within the three time-points was used to assign the participants’ relationship status, housing tenure, urbanicity, and UK country of residence. Although we used data on birth sex and refer in the text to “sex inequalities”, differences across the groups (labelled as “men” and “women”) are considered as the result not only of biological sex but also different experiences of socialisation and oppression.

Additional variables were included in the models as ‘a priori’ confounders. These comprised birth sex (in the models by inequality factors different than birth sex); highest academic or vocational qualification level achieved (harmonised into National Vocational Qualification [NVQ] levels,^29^ and corresponding to the parents’ qualification in the case of MCS participants); pre-pandemic self-reported health; existence of psychological distress in the most recent pre-pandemic cohort assessment; and household composition. Further details on these variables can be found in the **Appendix S3** (Supplementary Material).

## Data analyses

We used a multilevel growth curve modelling approach to analyse differences in the initial levels (at the first survey wave) and change over time (throughout the two additional survey waves) across subgroups in the different outcomes under study. After an initial visual exploration of the outcomes, linear (i.e., stable increase/decrease) and quadratic (i.e., accelerated/decelerated change) time terms were included to account for curvilinear trends. Cohort (a), the above-mentioned subgrouping variables (b), and the interaction among cohort and the subgrouping variables (c) were included in the models to explore generational inequalities, subgroup inequalities, and the interplay between generational and subgroup inequalities in the outcomes’ initial levels. In turn, the interaction between each of these three terms (a-c) and each of the two time terms (linear and quadratic) were included in the models to explore generational inequalities, subgroup inequalities, and the interplay between generational and subgroup inequalities in the change over time in the outcomes. Separate sets of models were estimated for each outcome and for each subgrouping variable, starting with a set of models with cohort as the only subgrouping variable. Unadjusted and adjusted (including birth sex, highest qualification achieved, pre-pandemic self-reported health and psychological distress, and household composition as covariates) models were estimated. Marginal mean estimates and 95% CIs of the outcomes were obtained from each of the models (unadjusted and adjusted) and plotted by the different subgroups. Contrasts of marginal predicted levels were performed in some cases to obtain estimates (and 95% CIs) of the differences in the adjusted marginal means (*diff*) by the inequality factors at the beginning of the study, as well as of the change in those initial differences (difference-in-differences, *DID*) by the end of the study period.

To further explore the existence of accelerated generational inequalities in the initial outcome levels, we estimated an additional set of models to answer the counterfactual question of when MCS participants ‘should’ have been born to have their mental and social wellbeing initial levels, provided that generational inequalities had not accelerated (in other words, what birth year more closely resembled the marginal mean levels predicted for the MCS cohort if generational inequalities were linear).

Further details on the analytical approaches used are available in **Appendix S4** (Supplementary Material). To investigate the impact of missing data, unadjusted models were also computed restricting their samples to those of the adjusted models as sensitivity analyses.

All analyses and plots were carried out in Stata MP 17.^30^

## Results

Data from 26,772 survey participants were analysed, comprising 57,048 observations across the three survey waves of the COVID-19 surveys (**Appendix S1**, Supplementary Material). As shown in **Table 1**, most participants were women (52.3%-62.4%), living in England (65.3%-94.6%), and White (73.2% in NS and 83.0% in MCS). Most participants reported having a comfortable financial situation in the months before the pandemic onset (although younger generations were more likely to report being in comparatively worse situations); being in a relationship (except members in the youngest cohort, MCS); owning/partly owning a house (with decreasing proportions of house ownership among younger participants); living in urban areas (with older adults being more likely to live in rural areas); and living with their partners or with their partners and others (except for the two youngest generations, NS and MCS, which were more likely to live in a different arrangement but not living alone, most particularly among MCS cohort members).

**Table 1.** Sociodemographic, economic, and living setting characteristics of the COVID-19 survey participants from the different cohorts.

|  | Cohort's name and birth year |  |  |  |  |
| --- | --- | --- | --- | --- | --- |
|  | **NSHD,**<br>**1946** | **NCDS,**<br>**1958** | **BCS, 1970** | **NS, 1990** | **MCS, 2000** |
| **No. of participants** | 1,567 | 7,691 | 7,042 | 4,971 | 5,501 |
| **No. of observations per survey wave** |  |  |  |  |  |
| Survey wave 1 – May 2020 | 1,170 | 5,119 | 4,132 | 1,876 | 2,607 |
| Survey wave 2 – September/October 2020 | 1,488 | 6,228 | 5,236 | 3,609 | 3,229 |
| Survey wave 3 – February/March 2021 | 1,325 | 6,757 | 5,684 | 4,167 | 4,421 |
| **Women, N (%)** | 820 (52.3) | 4,019 (52.3) | 3,933 (55.9) | 3,101 (62.4) | 3,314 (60.2) |
| **Pre-pandemic financial situation, N (%)** |  |  |  |  |  |
| Just about getting by/Finding it quite/very difficult | 60 (3.8) | 800 (10.4) | 1,144 (16.2) | 932 (18.7) | 964 (17.5) |
| Doing all right | 312 (19.9) | 2,531 (32.9) | 2,719 (38.6) | 2,015 (40.5) | 2,344 (42.6) |
| Living comfortably | 986 (62.9) | 4,055 (52.7) | 2,909 (41.3) | 1,827 (36.8) | 1,825 (33.2) |
| *Missing* | 209 (13.3) | 305 (4.0) | 270 (3.8) | 197 (4.0) | 368 (6.7) |
| **In a relationship, N (%)** |  |  |  |  |  |
| No | 46 (2.9) | 918 (11.9) | 839 (11.9) | 1,068 (21.5) | 3,172 (57.7) |
| Yes | 1,271 (81.1) | 6,314 (82.1) | 5,904 (83.8) | 3,691 (74.3) | 2,169 (39.4) |
| *Missing* | 250 (16.0) | 459 (6.0) | 299 (4.2) | 212 (4.3) | 160 (2.9) |
| **Housing tenure, N (%)** |  |  |  |  |  |
| No (rented/rent-free/other arrangement) | 100 (6.4) | 912 (11.9) | 1,095 (15.5) | 2,049 (41.2) | 2,668 (48.5) |
| Yes (house owned/partly owned) | 1,371 (87.5) | 6,066 (78.9) | 5,007 (71.1) | 2,493 (50.2) | 1,843 (33.5) |
| *Missing* | 96 (6.1) | 713 (9.3) | 940 (13.3) | 429 (8.6) | 990 (18.0) |
| **Urban vs rural dwelling, N (%)** |  |  |  |  |  |
| Urban | 1,021 (65.2) | 5,264 (68.4) | 5,002 (71.0) | 4,059 (81.7) | 3,935 (71.5) |
| Rural | 527 (33.6) | 2,062 (26.8) | 1,684 (23.9) | 703 (14.1) | 951 (17.3) |
| *Missing* | 19 (1.2) | 365 (4.7) | 356 (5.1) | 209 (4.2) | 615 (11.2) |
| **Country, N (%)** |  |  |  |  |  |
| England | 1,365 (87.1) | 6,257 (81.4) | 5,769 (81.9) | 4,701 (94.6) | 3,591 (65.3) |
| Northern Ireland |  | 5 (0.1) | 7 (0.1) | 6 (0.1) | 387 (7.0) |
| Scotland | 121 (7.7) | 670 (8.7) | 566 (8.0) | 28 (0.6) | 631 (11.5) |
| Wales | 62 (4.0) | 399 (5.2) | 350 (5.0) | 32 (0.6) | 639 (11.6) |
| *Missing* | *19 (1.2)* | *360 (4.7)* | *350 (5.0)* | *204 (4.1)* | *253 (4.6)* |
| **Ethnicity, N (%)** |  |  |  |  |  |
| White * |  |  |  | 3,637 (73.2) | 4,461 (81.1) |
| Mixed |  |  |  | 224 (4.5) | 240 (4.4) |
| Indian/Pakistani/Bangladeshi |  |  |  | 778 (15.7) | 511 (9.3) |
| Black Caribbean/Black African |  |  |  | 219 (4.4) | 155 (2.8) |
| Other ** |  |  |  | 113 (2.3) | 130 (2.4) |
| *Missing* |  |  |  |  | *4 (0.1)* |
| **Highest qualification (academic or vocational), N (%)\*\*\*** |  |  |  |  |  |
| None | 325 (20.7) | 493 (6.4) | 507 (7.2) | 145 (2.9) | 513 (9.3) |
| NVQ-1 | 99 (6.3) | 734 (9.5) | 453 (6.4) | 278 (5.6) | 319 (5.8) |
| NVQ-2 | 348 (22.2) | 1,866 (24.3) | 1,702 (24.2) | 890 (17.9) | 1,378 (25.0) |
| NVQ-3 | 496 (31.7) | 1,350 (17.6) | 967 (13.7) | 993 (20.0) | 795 (14.5) |
| NVQ-4 | 190 (12.1) | 2,682 (34.9) | 2,387 (33.9) | 1,295 (26.1) | 1,893 (34.4) |
| NVQ-5 | 20 (1.3) | 393 (5.1) | 543 (7.7) | 815 (16.4) | 276 (5.0) |
| *Missing* | *89 (5.7)* | *173 (2.2)* | *483 (6.9)* | *555 (11.2)* | *327 (5.9)* |
| **Psychological distress in the most recent pre-COVID sweep, N (%)** |  |  |  |  |  |
| No | 1,249 (79.7) | 5,985 (77.8) | 4,717 (67.0) | 3,201 (64.4) | 4,321 (78.5) |
| Yes | 160 (10.2) | 883 (11.5) | 1,030 (14.6) | 1,111 (22.3) | 939 (17.1) |
| *Missing* | *158 (10.1)* | *823 (10.7)* | *1,295 (18.4)* | *659 (13.3)* | *241 (4.4)* |
| **Pre-pandemic self-reported health, N (%)** |  |  |  |  |  |
| Poor | 34 (2.2) | 305 (4.0) | 213 (3.0) | 97 (2.0) | 168 (3.1) |
| Fair | 165 (10.5) | 967 (12.6) | 774 (11.0) | 470 (9.5) | 608 (11.1) |
| Good | 441 (28.1) | 2,536 (33.0) | 2,277 (32.3) | 1,519 (30.6) | 1,722 (31.3) |
| Very good | 535 (34.1) | 2,930 (38.1) | 2,840 (40.3) | 2,114 (42.5) | 2,096 (38.1) |
| Excellent | 154 (9.8) | 912 (11.9) | 905 (12.9) | 756 (15.2) | 875 (15.9) |
| *Missing* | *238 (15.2)* | *41 (0.5)* | *33 (0.5)* | *15 (0.3)* | *32 (0.6)* |
| **Household composition, N (%)** |  |  |  |  |  |
| Living alone | 368 (23.5) | 1,884 (24.5) | 1,047 (14.9) | 633 (12.7) | 109 (2.0) |
| Living with partner | 1,076 (68.7) | 3,569 (46.4) | 1,230 (17.5) | 1,334 (26.8) | 140 (2.5) |
| Living with partner and other(s) | 73 (4.7) | 1,688 (21.9) | 3,928 (55.8) | 1,569 (31.6) | 172 (3.1) |
| Other arrangements | 47 (3.0) | 486 (6.3) | 773 (11.0) | 1,374 (27.6) | 4,975 (90.4) |
| *Missing* | *3 (0.2)* | *64 (0.8)* | *64 (0.9)* | *61 (1.2)* | *105 (1.9)* |

*Note.* BCS: 1970 British Cohort Study; NCDS: 1958 National Child Development Study; NVQ: harmonised qualification categories according to the National Vocational Qualification system (higher numbers represent higher qualification); NS: 1990 Next Steps; NSHD: 1946 National Survey of Health and Development; MCS: 2000 Millennium Cohort Study. \* “White” includes all British and non-British White participants; \*\* “Other” includes all ethnicities not included in the previous categories. \*\*\* In MCS, highest qualification corresponds to the parents’ qualification.

Coefficients, along with the resulting marginal mean estimates and 95% CIs and their corresponding visual depictions, are provided in the **Appendices S5-S12** (Supplementary Material), organised by inequality factors. Overall, mental and social wellbeing outcomes worsened throughout the first year of the pandemic. Compared with May 2020, anxiety, depression, loneliness, and life satisfaction (LS) adjusted marginal mean levels were, on average, *diff*_GAD-2_=0.14 [0.11, 0.17], *diff*_PHQ-2_=0.10 [0.07, 0.13], *diff*_UCLA-3_=0.15 [0.12, 0.18], and *diff*_LS_=−0.41 [−0.45, −0.36] points worse, respectively, by February/March 2021, relatively small changes considering the scale ranges. However, the change was not predominantly linear (**Figure 1**): increases in anxiety symptomatology were most pronounced between May 2020 (during the first lockdown) and September/October 2020 (between the first and second lockdowns). By contrast, levels of depressive symptomatology, loneliness, and life satisfaction seemed to improve by September/October 2020, further worsening by February/March 2021 (during the third lockdown).

**Figure 1.**
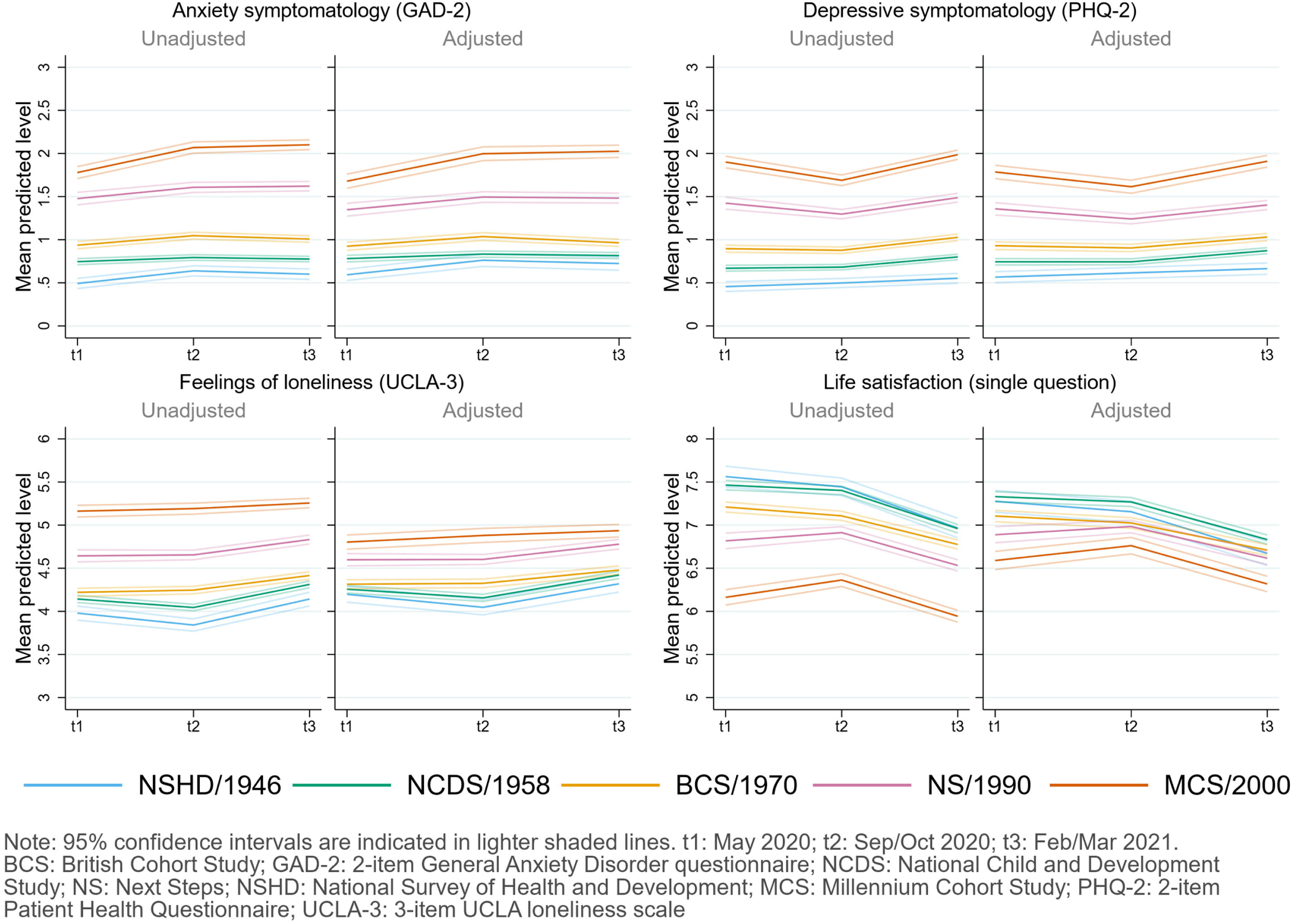
Unadjusted and adjusted marginal mean estimates and 95% CIs estimated for each outcome at each time-point and cohort. Adjusted models included birth sex, highest qualification achieved, pre-pandemic self-reported health and psychological distress, and household composition as covariates.

### Inequalities by generation

Younger cohorts had higher initial levels of anxiety and depressive symptomatology and were more likely to report loneliness and lower life satisfaction compared to older cohorts (**Figure 1**). These generational inequalities were particularly salient in the two youngest cohorts (NS/1990, MCS/2000-02), and remained evident after adjustment and throughout the first year of the pandemic (**Appendix S5**). The differences in the initial levels between the youngest (MCS) and oldest (NSHD) cohorts were, on average, 1.09 [0.97, 1.20] for anxiety symptomatology; 1.21 [1.11, 1.33] for depressive symptomatology; 0.60 [0.47, 0.73] for loneliness; and −0.69 [−0.86, −0.52] for life satisfaction. By the end of the study period, that difference had become *DID*_GAD-2_=0.22 [0.10, 0.33] points wider for anxiety symptomatology, mainly driven by a greater increase in anxiety symptoms among the youngest cohort between the first and second time-points. The gaps between the youngest and oldest cohorts remained stable for loneliness (*DID*_PHQ-2_=0.01 [−0.11, 0.13]) and for depressive symptomatology (*DID*_UCLA-3_=0.03 [−0.07, 0.13]), despite a temporary improvement in the depressive symptomatology levels among the two youngest cohorts (NS and MCS) by the second time-point. Generational inequalities in life satisfaction narrowed by *DID*_LS_=−0.33 [−0.51, −0.15] points when comparing the oldest and youngest cohorts. Importantly, the adjusted life satisfaction initial levels were not the highest among the oldest (NSHD, *M*=7.28 [7.15, 7.40]) but among the immediately younger generation (NCDS, *M*=7.33 [7.27, 7.39]). Nevertheless, the narrowing of the life satisfaction generational inequalities was also observed when comparing the cohorts with the highest (NCDS) and lowest (MCS) levels, with a reduction in the gap of *DID*=−0.23 [−0.34, −0.11] points over the first year of the pandemic.

Figure 2. shows the comparison between the adjusted predicted initial levels for each cohort, obtained from the abovementioned models, and those predicted by birth year from the models estimated excluding MCS data to explore whether the generational inequalities in the initial levels were accelerating. Although the predictions by the models by birth year for anxiety and depressive symptomatology broadly overlapped with those by cohort for the generations born between 1946 and 1990, the youngest cohort’s levels were substantially higher than expected if the increase had been linear over time. This was most noticeable for depressive symptomatology, where the point estimate (*M*_MCS_=1.79 [1.71, 1.86]) corresponded to that of those born 15 years later, in 2015 (*M*_2015_=1.78 [1.67, 1.89]). These results were robust to the inclusion of additional pre-pandemic characteristics (**Figure S5.2**, Supplementary Material). The adjusted models by birth year did not seem to adequately capture the marginal initial levels of loneliness (which seemed to follow an accelerated increasing trend early on, with levels for those born in 1990 higher than expected from a linear trend) and life satisfaction levels (which seemed to follow a more complex trend).

**Figure 2.**
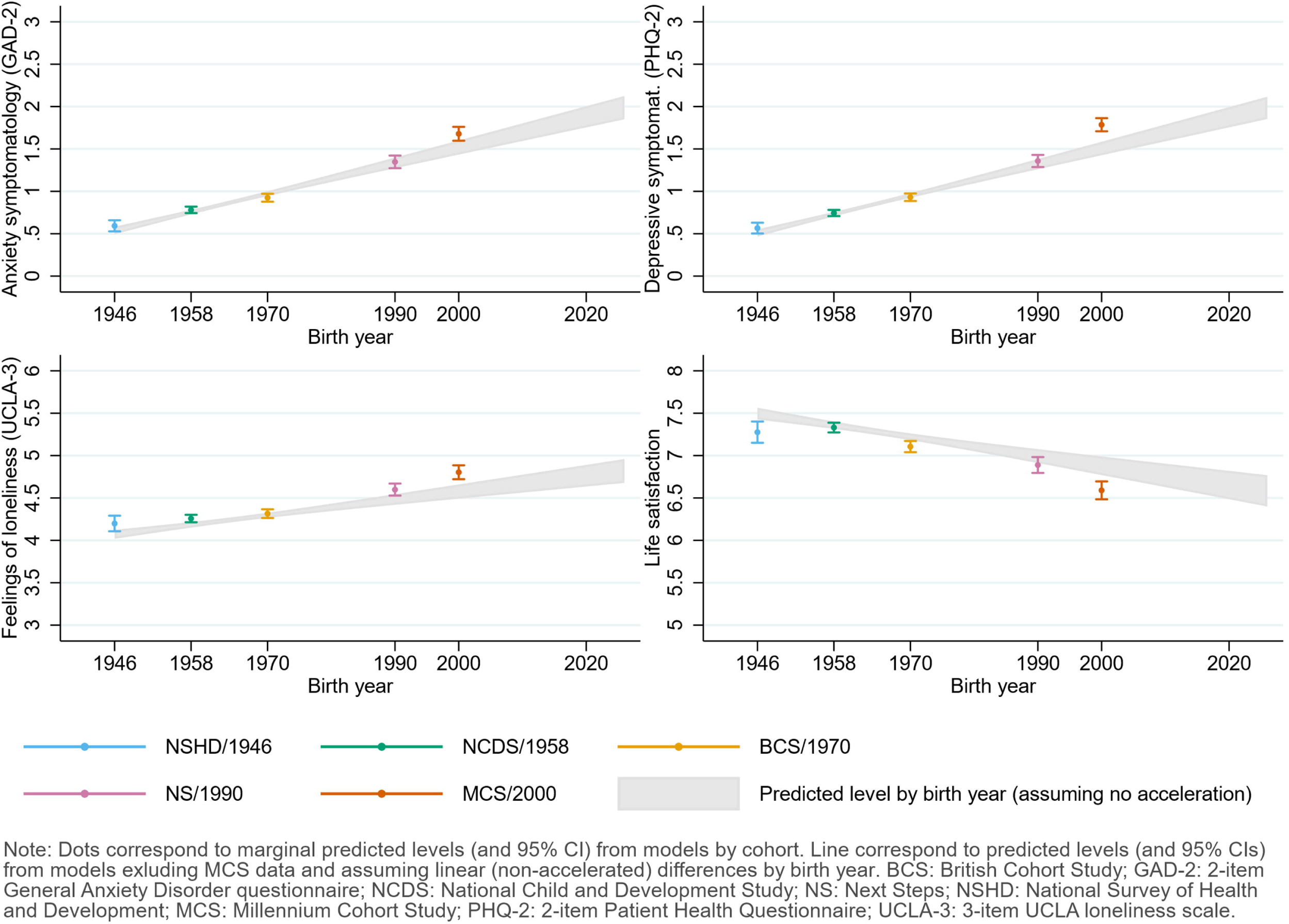
Comparison between marginal predicted initial levels by cohort and marginal predicted initial levels by birth year assuming no acceleration in the differences by birth year and excluding data from the youngest cohort (MCS). Models adjusted by birth sex, highest qualification achieved, pre-pandemic self-reported health and psychological distress, and household composition.

### Inequalities by birth sex

Women had, on average, worse initial anxiety symptomatology (*diff*_GAD-2_=0.48 [0.43, 0.54]), depressive symptomatology (*diff*_PHQ-2_=0.23 [0.18, 0.28]), loneliness (*diff*_UCLA-3_=0.19 [0.13, 0.25]), and life satisfaction (*diff*_LS_=−0.11 [−0.19, −0.03]) levels than men (**Figure 3**). However, there was generational variation: women from younger cohorts had higher-than-expected initial levels of anxiety (adjusted unstandardised regression coefficients B_NS*woman_=0.22 [0.06, 0.38]; B_MCS*woman_=0.56 [0.40, 0.71]) and depressive symptomatology (B_MCS*woman_=0.36 [0.21, 0.52]), and lower-than-expected initial levels of loneliness (B_NS*woman_=−0.17 [−0.33, −0.002]; B_MCS*woman_=−0.19 [−0.36, −0.02]) (**Appendix S6**). Transient improvements by the second time-point in depressive symptomatology, previously observed among the youngest cohorts, were observed mainly among women (B_NS*woman*t2_=−0.26 [−0.45, −0.06]; B_MCS*woman*t2_=−0.29 [−0.50, −0.08]). Nevertheless, by the end of the study period, sex inequalities were not substantially different than at the beginning (**Appendix S6**).

**Figure 3.**
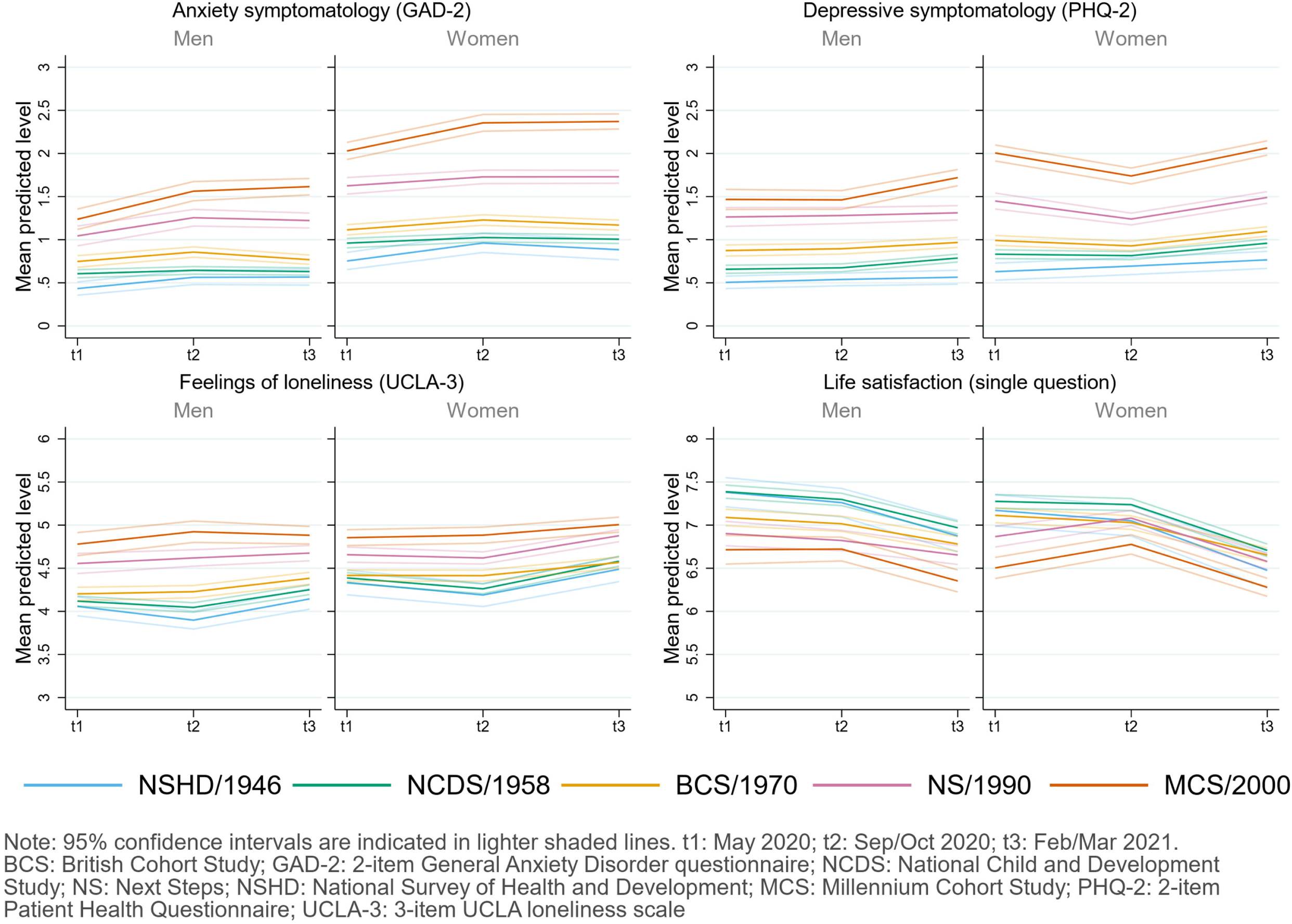
Adjusted marginal mean estimates and 95% CIs estimated for each outcome at each time-point and cohort by birth sex. Models adjusted by birth sex, highest qualification achieved, pre-pandemic self-reported health and psychological distress, and household composition.

### Inequalities by pre-pandemic financial situation

Pre-pandemic financial situation drove some of the largest differences in the initial levels in all outcomes, with those in worse situations showing worse initial levels in all outcomes (**Figure 4**). The average adjusted difference in the initial levels between those in the worst-off and best-off pre-pandemic financial situations was *diff*_GAD-2_=0.54 [0.40, 0.68], *diff*_PHQ-2_=0.68 [0.54, 0.84], *diff*_UCLA-3_=0.68 [0.54, 0.82], and *diff*_LS_=−1.21 [−1.41, −1.01]. Differences between the “doing all right” and “living comfortably” subgroups among the oldest adults were smaller than in the other cohorts, resulting in these other cohorts overtaking the oldest in the financially best-off group; however, oldest adults in the worst-off financial situation subgroup had substantially worse levels in all outcomes (**Appendix S7**). Overall, there was no substantial change in the differences between those in the worst-off and best-off financial situations by the end of the study period (**Appendix S7**).

**Figure 4.**
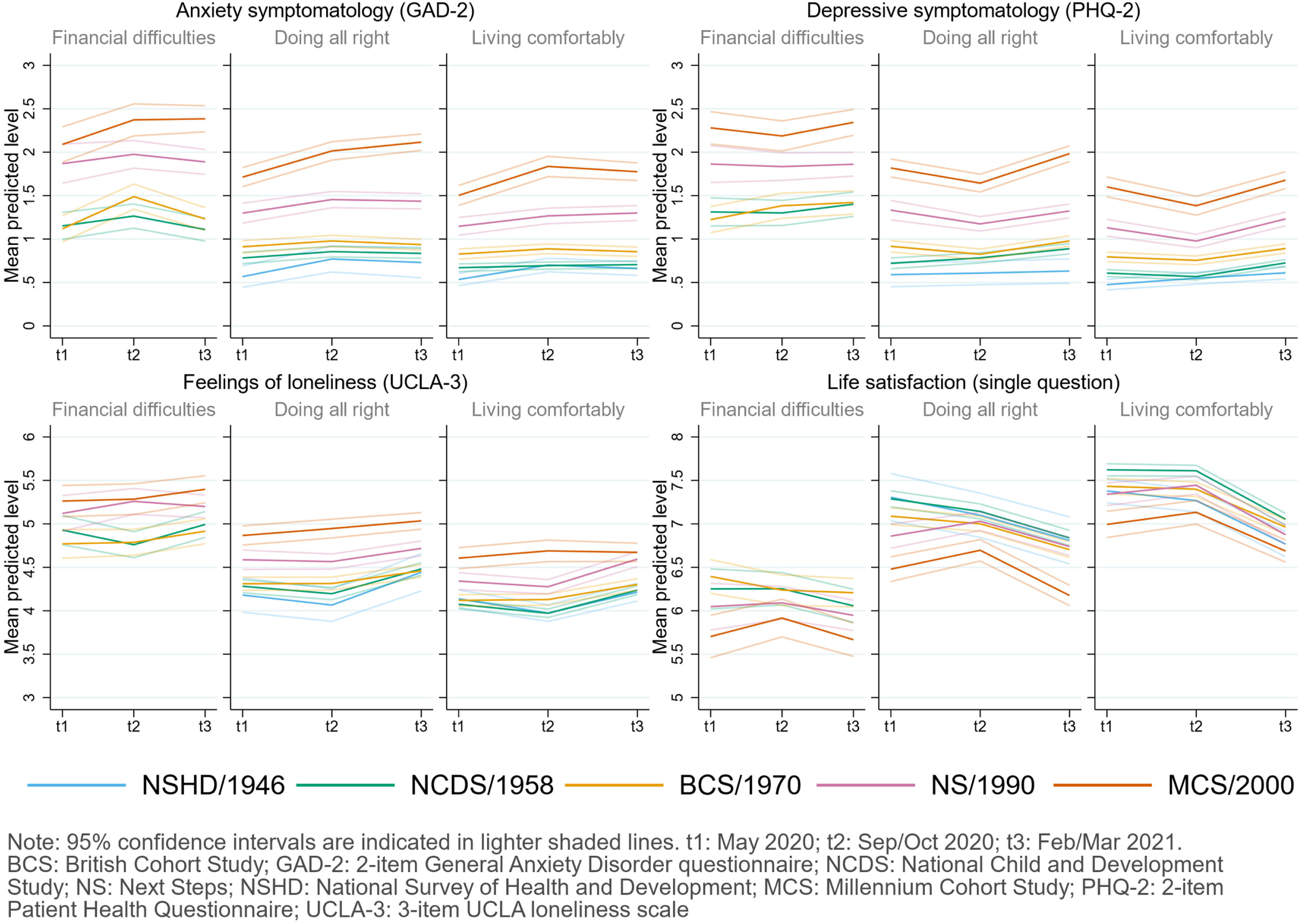
Adjusted marginal mean estimates and 95% CIs estimated for each outcome at each time-point and cohort by pre-pandemic financial situation. NSHD data for the “financial difficulties” group is not shown due to the small number of cases.

### Inequalities by additional factors

Those in a relationship had, in general, better initial levels in all outcomes (**Appendix S8**). These differences varied substantially across generations, particularly among the youngest, where the differences were smaller or, as in the case of anxiety symptomatology, went in the opposite direction, with those in a relationship showing higher anxiety levels. The initial increase in anxiety symptomatology among the youngest cohort took place mainly among those not in a relationship. In turn, the increase in depressive symptomatology between the second and third time-points in the NS/1990 cohort only occurred among those in a relationship, who still had better levels than those not in a relationship by the end of the study period.

Participants owning/partly owning a house showed better initial levels in all outcomes (**Appendix S9**), and the difference was again less pronounced among the youngest. Change over time in the outcomes was very similar by housing tenure.

People living in rural areas showed, overall, slightly better initial levels in all outcomes than those living in urban areas (*diff*_GAD-2_=−0.04 [−0.10, 0.03], *diff*_PHQ-2_=−0.06 [−0.12, −0.003], *diff*_UCLA-3_=−0.02 [−0.09, 0.05], *diff*_LS_=0.015 [0.06, 0.24]). There were differences in the change over time in the outcomes (**Appendix S10**), with the abovementioned temporary improvements in the two youngest cohorts’ depressive symptomatology occurring mainly among those living in urban areas, and a temporary increase in the loneliness levels happening among those from the youngest cohort living in rural areas. Despite these differences, gaps between people living in rural and urban areas in the initial levels were similar at the end of the study period.

Participants living in England had the lowest initial levels of life satisfaction (**Appendix S11**), although the difference with Scotland was no longer apparent after adjusting for confounders; and initial levels of depressive symptomatology and life satisfaction among the youngest were best for those living in Northern Ireland. Several differences could be observed in the change in the outcomes over time by country, particularly in the degree to which some of those outcomes changed by the second time-point, matching with periods of relatively eased restrictions, although these patterns were not homogeneous across generations living in the same UK countries.

Inequalities in the initial levels could be observed across ethnic groups (**Appendix S12**). Increases in the levels of anxiety and depressive symptomatology were larger among Black African/Black Caribbean participants from the youngest cohort, whose levels at the end of the study period were the highest of all groups with adjusted marginal means of *M*_GAD-2_=2.11 [1.72, 2.49] and *M*_PHQ-2_= 2.14 [1.69, 2.60].

### Sensitivity analyses

The sensitivity analyses (non-fully adjusted models performed with the same sample as the fully adjusted models) showed very similar results as those with unrestricted samples (**Appendices S5-S12**).

## Discussion

This study provides evidence on inequalities in initial levels and change over time of several mental and social wellbeing outcomes in the UK population during the first year of the COVID-19 pandemic. These inequalities were largely observed by generation, birth sex, and financial situation before the pandemic onset, but also by relationship status, housing tenure, urbanicity, country of residence, and ethnicity. We also found evidence of interactions between generational and other inequality factors like birth sex, relationship status, housing tenure, and urbanicity, suggesting that some inequalities do not occur equivalently across generations.

Younger cohorts systematically showed worse levels in most outcomes at most time-points. These inequalities were still evident after accounting for relevant variables (such as pre-existing psychological distress and health levels) and disaggregating the data by the different subgrouping variables studied. Our results also suggest that the generational inequalities have narrowed in some cases (by about 0.33 points in life satisfaction, a small difference considering the 10-point range of the measure) and became wider in others (by about 1.09 points in anxiety, a large difference considering the 6-point range of the measure) during the first year of the pandemic. The inconsistencies of our results, compared with a previous study showing improving anxiety levels among UK young adults ^11^ may be explained by differences between the periods covered in that study (which spanned up to August 2020, when the reinstatement of restrictions by mid-October 2020 had not yet been announced), and ours (with the second time-point already taking place after that announcement by mid-September 2020).^9^ We found evidence of accelerated generational inequalities in initial anxiety and, most notably, depression levels, with the youngest generation’s initial levels being substantially worse than expected, if the generational inequalities had followed a linear trend. Thus, and considering that the initial levels in this study correspond to the levels in the early stages of the pandemic, our findings are not only consistent with the idea that the pandemic onset had a disproportionate impact among the youngest,^3,4,6^ but suggest that this impact was even larger than expected if generational inequalities had followed a linear trend. All outcomes remained substantially worse for the youngest cohort by the end of the study period, indicating that, regardless of the source of those inequalities, these have not reduced. Thus, younger generations (i.e., those in their late teenage years, early twenties) may be the most vulnerable age groups during these challenging times.

A puzzling exception to this general trend was found in the life satisfaction levels of those born in 1946, which were lower than expected, especially among women. Although life satisfaction levels have been found to increase with age in cross-sectional studies,^31^ the longitudinal evidence shows that, with age, a steeper decline in these levels is expected.^32^ However, our study suggests that the oldest adults’ (NSHD/1946) life satisfaction levels were already lower than those of the immediately younger generation (NCDS/1958) at the beginning of the study. Altogether, this may suggest that the pandemic onset had a larger impact among the UK’s older adults’ (i.e., those in their seventies) life satisfaction levels. Further research is needed to analyse the differential impact of the pandemic onset on life satisfaction trajectories in the UK’s older populations.

Women showed worse levels than men in all outcomes at most time-points, and generational inequalities in anxiety and depressive symptomatology seemed to be substantially larger among women, highlighting the interplay between generational and sex disparities. In line with previous evidence,^11^ we found that sex inequalities in anxiety and depressive symptomatology narrowed over time during the initial months of the pandemic; however, those inequalities widened once again by the end of the study period. Very similar results have been reported on distress for young adults (18-29) between April and November 2020.^14^ Altogether, these findings may suggest a differential impact of the policies put in place to control the pandemic and highlight the importance of continually monitoring the levels of mental and social wellbeing over time in the population, as trajectories may not follow a linear trend, particularly under the rapidly changing scenarios which took place after the pandemic onset.

Inequalities by pre-pandemic financial situation were observed across all generations. The large, cross-cutting, and relatively homogeneous better mental and social wellbeing levels among those doing financially “all right” compared to those in the worst-off financial situation suggests that this may be an optimal target for public policies aimed at enhancing mental and social wellbeing.

We explored inequalities by urbanicity and country of residence, a gap in knowledge highlighted by previous literature.^11^ People living in rural areas showed slightly better outcomes than those living in urban areas, thus shedding some light on the existing mixed evidence.^2^ However, by explicitly exploring the interplay between generational and the subgrouping variables, we could qualify previous evidence showing an association between urban settings and higher loneliness,^33^ suggesting that, among the youngest, the opposite relationship seemed to be occurring over time. Additionally, initial life satisfaction levels were generally lowest among people living in England. Nevertheless, the differences found by these geographical aspects were the smallest among those investigated in the present study.

Our study also shows that people in a relationship and owning/partly owning a house had, in general, better mental and social wellbeing than those not. The fact that a great majority of the youngest participants were living with their parents^34^ may explain why differences by these factors were smaller than in older generations, as very few were cohabiting with their partners and the financial pressures of housing expenditure may fall on older family members.

Finally, we also found differences by ethnicity in most outcomes. Studies from the initial stages of the pandemic suggest a higher impact of the pandemic onset on Black, Asian, and minority ethnic people’s distress levels in high-income countries including the UK,^2,5^ whereas recent evidence from 11 UK longitudinal studies did not find such evidence.^7^ Although the small sample sizes of minority ethnic groups made it hard to provide more solid evidence in our study, the results suggest that there is a high heterogeneity both between and within ethnic minority groups (for instance, by generation) and, therefore, grouping them together into a single group may not be adequate, as this may obscure underlying differences.

By using data from five probability samples of the UK population representing generations born in different years (i.e., 1946, 1958, 1970, 1990, and 2000-2002), and using weights to account for both the survey designs and the probability of participating in each of the three COVID-19 survey waves, this study provides evidence that is nationally representative and generalisable to the UK adult population. Our study provides evidence on a wide range of mental and social wellbeing outcomes experienced by people during the different phases of the pandemic. Unlike most available evidence, this study provides nuanced evidence on the mental and social wellbeing inequalities in both the initial levels and changes over time. We also accounted for the interplay between generational inequalities and other inequality factors. Moreover, by using data from cohort studies already existing prior to the pandemic onset we could control for pre-pandemic characteristics measured prospectively instead of retrospectively.

However, this study has several limitations. First, the tools used to assess anxiety and depressive symptomatology included only core symptoms, thus providing a relevant but relatively limited picture of that symptomatology. Extended assessment tools such as the GAD-7 or the PHQ-9 could not be included due to logistic limitations and to avoid increasing respondent burden. Second, although our study covers an extended period up to March 2021, the reduced number of repeated assessments limits the granularity of the identified trajectories. Therefore, we acknowledge that there may be additional dynamics taking place between the time-points covered in this study.^16^ Third, although the inclusion of multiple interaction terms allowed the trajectories to vary in both their initial levels and growth parameters by the subgrouping variables under study, this resulted in a substantial reduction of power to assess these differences. Future research may use alternative analytical approaches that allow accounting for multiple intersecting social identities (e.g., ethnicity, gender, sexual orientation) tied to social power developed for their use from an intersectional approach.^35–37^ It is also important to note that the use of self-reported information to characterise the mental and social wellbeing of participants may have led to the underestimation of emotional difficulties in cases where reporting such experiences may be potentially stigmatised, such as among men.^2^ Finally, although the analyses were adjusted for relevant pre-pandemic characteristics, the influence of unmeasured confounding cannot be ruled out, thus limiting the causal interpretation of the findings.

Overall, our study builds upon previous evidence showing generational, sex, and financial inequalities in mental and social wellbeing outcomes,^2-6,10,11,13-16,33^ by showing that, in the UK adult population, these inequalities persisted one year after the first national lockdown. Our study provides crucial evidence on the acceleration of generational mental health inequalities with the pandemic onset, with the younger cohorts not only being more impacted than the older cohorts, but also beyond what would have been expected if that impact was similar to pre-pandemic generational differences. Moreover, it shows that generational inequalities in anxiety have widened, whereas those in life satisfaction have narrowed, although with all generations showing substantially worse levels at the end of the first year of the pandemic than at its early stages. Crucially, our study highlights the importance of exploring the interplay between generational inequalities and those posed by other characteristics such as birth sex, relationship status, or urbanicity, as some combinations of inequality factors show even worse results than expected by simply summing them (e.g., anxiety and depressive symptomatology among younger women). As the COVID-19 pandemic continues, it is critical to keep monitoring mental and social wellbeing levels with an appropriate level of granularity. In particular, measures to support the mental health of the most vulnerable groups in the population may be needed, with a focus on reducing existing gaps and preventing new gaps from appearing.

## Data Availability

Deidentified data and documentation on NCDS/1958, BCS/1970, NS/1990, and MCS/2000-02 are available from the UK Data Service: https://ukdataservice.ac.uk/. Deidentified data and documentation on NSHD/1946 are available under request from https://www.nshd.mrc.ac.uk/data.

https://www.nshd.mrc.ac.uk/data

## Acknowledgements

We would like to thank all individuals who participated in these five cohort studies (NSHD, NCDS, BCS, NS, and MCS) for so generously giving up their time over so many years, and all the study team members for their tremendous efforts in collecting and managing the data.

## References

1. Prati G, Mancini AD. The psychological impact of COVID-19 pandemic lockdowns: a review and meta-analysis of longitudinal studies and natural experiments. Psychol Med. Jan 2021;51(2):201–211. doi:10.1017/S0033291721000015

2. Gibson B, Schneider J, Talamonti D, Forshaw M. The impact of inequality on mental health outcomes during the COVID-19 pandemic: A systematic review. Canadian Psychology/Psychologie canadienne. 2021;62(1):101–126. doi:10.1037/cap0000272

3. Pierce M, Hope H, Ford T, et al. Mental health before and during the COVID-19 pandemic: a longitudinal probability sample survey of the UK population. Lancet Psychiatry. Oct 2020;7(10):883–892. doi:10.1016/S2215-0366(20)30308-4

4. Niedzwiedz CL, Green MJ, Benzeval M, et al. Mental health and health behaviours before and during the initial phase of the COVID-19 lockdown: longitudinal analyses of the UK Household Longitudinal Study. J Epidemiol Community Health. Mar 2021;75(3):224–231. doi:10.1136/jech-2020-215060

5. Proto E, Quintana-Domeque C. COVID-19 and mental health deterioration by ethnicity and gender in the UK. PloS one. 2021;16(1):e0244419. doi:10.1371/journal.pone.0244419

6. Kwong ASF, Pearson RM, Adams MJ, et al. Mental health before and during the COVID-19 pandemic in two longitudinal UK population cohorts. The British journal of psychiatry : the journal of mental science. Nov 24 2020:1–10. doi:10.1192/bjp.2020.242

7. Patel K, Robertson E, Kwong ASF, et al. Psychological Distress Before and During the COVID-19 Pandemic: Sociodemographic Inequalities in 11 UK Longitudinal Studies. medRxiv. 2021:2021.10.22.21265368. doi:10.1101/2021.10.22.21265368

8. Creese B, Khan Z, Henley W, et al. Loneliness, physical activity, and mental health during COVID-19: a longitudinal analysis of depression and anxiety in adults over the age of 50 between 2015 and 2020. Int Psychogeriatr. May 2021;33(5):505–514. doi:10.1017/S1041610220004135

9. Institute for Government. Timeline of UK government coronavirus lockdowns. Accessed 4 November 2021, 2021. https://www.instituteforgovernment.org.uk/charts/uk-government-coronavirus-lockdowns

10. O’Connor RC, Wetherall K, Cleare S, et al. Mental health and well-being during the COVID-19 pandemic: longitudinal analyses of adults in the UK COVID-19 Mental Health & Wellbeing study. The British journal of psychiatry : the journal of mental science. Oct 21 2020:1–8. doi:10.1192/bjp.2020.212

11. Fancourt D, Steptoe A, Bu F. Trajectories of anxiety and depressive symptoms during enforced isolation due to COVID-19 in England: a longitudinal observational study. Lancet Psychiatry. Feb 2021;8(2):141–149. doi:10.1016/S2215-0366(20)30482-X

12. Varga TV, Bu F, Dissing AS, et al. Loneliness, worries, anxiety, and precautionary behaviours in response to the COVID-19 pandemic: A longitudinal analysis of 200,000 Western and Northern Europeans. Lancet Reg Health Eur. Mar 2021;2:100020. doi:10.1016/j.lanepe.2020.100020

13. Saunders R, Buckman JEJ, Fonagy P, Fancourt D. Understanding different trajectories of mental health across the general population during the COVID-19 pandemic. Psychol Med. Mar 3 2021:1–9. doi:10.1017/S0033291721000957

14. Stroud I, Gutman LM. Longitudinal changes in the mental health of UK young male and female adults during the COVID-19 pandemic. Psychiatry Res. Sep 2021;303:114074. doi:10.1016/j.psychres.2021.114074

15. Pierce M, McManus S, Hope H, et al. Mental health responses to the COVID-19 pandemic: a latent class trajectory analysis using longitudinal UK data. Lancet Psychiatry. Jul 2021;8(7):610–619. doi:10.1016/S2215-0366(21)00151-6

16. Ellwardt L, Prag P. Heterogeneous mental health development during the COVID-19 pandemic in the United Kingdom. Sci Rep. Aug 5 2021;11(1):15958. doi:10.1038/s41598-021-95490-w

17. Wadsworth M, Kuh D, Richards M, Hardy R. Cohort Profile: The 1946 National Birth Cohort (MRC National Survey of Health and Development). Int J Epidemiol. Feb 2006;35(1):49–54. doi:10.1093/ije/dyi201

18. Power C, Elliott J. Cohort profile: 1958 British birth cohort (National Child Development Study). Int J Epidemiol. Feb 2006;35(1):34–41. doi:10.1093/ije/dyi183

19. Elliott J, Shepherd P. Cohort profile: 1970 British Birth Cohort (BCS70). Int J Epidemiol. Aug 2006;35(4):836–43. doi:10.1093/ije/dyl174

20. Calderwood L, Sánchez C. Next Steps (formerly known as the Longitudinal Study of Young People in England). Open Health Data. 2016;4(1)doi:10.5334/ohd.16

21. Connelly R, Platt L. Cohort profile: UK Millennium Cohort Study (MCS). Int J Epidemiol.Dec 2014;43(6):1719–25. doi:10.1093/ije/dyu001

22. Kroenke K, Spitzer RL, Williams JB, Monahan PO, Lowe B. Anxiety disorders in primary care: prevalence, impairment, comorbidity, and detection. Ann Intern Med. Mar 6 2007;146(5):317–25. doi:10.7326/0003-4819-146-5-200703060-00004

23. Kroenke K, Spitzer RL, Williams JB. The Patient Health Questionnaire-2: validity of a two-item depression screener. Med Care. Nov 2003;41(11):1284–92. doi:10.1097/01.MLR.0000093487.78664.3C

24. Hughes ME, Waite LJ, Hawkley LC, Cacioppo JT. A Short Scale for Measuring Loneliness in Large Surveys: Results From Two Population-Based Studies. Res Aging. 2004;26(6):655–672. doi:10.1177/0164027504268574

25. Office for National Statistics. Personal well-being user guidance. Accessed 28 September 2021, https://www.ons.gov.uk/peoplepopulationandcommunity/wellbeing/methodologies/personalwellbeingsurveyuserguide

26. Rural and Environment Science and Analytical Services Division. Scottish Government Urban Rural Classification 2016. Scottish Government; 2018.

27. Bibby P, Brindley P. The 2011 Rural-Urban Classification For Small Area Geographies: A User Guide and Frequently Asked Questions (v1.0). Government Statistical Service; 2013.

28. Office for National Statistics. Ethnic group, national identity and religion. Accessed 02/02/2022, 2022. https://www.ons.gov.uk/methodology/classificationsandstandards/measuringequality/ethnicgroupnationalidentityandreligion

29. Dodgeon B, Parsons S. Deriving highest qualification in NCDS and BCS70. Centre for Longitudinal Studies, Institute of Education; 2011.

30. Stata Statistical Software: Release 17. StataCorp LLC; 2021.

31. Steptoe A, Deaton A, Stone AA. Subjective wellbeing, health, and ageing. Lancet. Feb 14 2015;385(9968):640–648. doi:10.1016/S0140-6736(13)61489-0

32. Jivraj S, Nazroo J, Vanhoutte B, Chandola T. Aging and subjective well-being in later life. J Gerontol B Psychol Sci Soc Sci. Nov 2014;69(6):930–41. doi:10.1093/geronb/gbu006

33. Bu F, Steptoe A, Fancourt D. Who is lonely in lockdown? Cross-cohort analyses of predictors of loneliness before and during the COVID-19 pandemic. Public Health. Sep 2020;186:31–34. doi:10.1016/j.puhe.2020.06.036

34. Zilanawala A, Chanfreau J, Sironi M, Palma M. Household composition, couples’ relationship quality, and social support during lockdown - Initial findings from the COVID-19 Survey in Five National Longitudinal Studies. UCL Centre for Longitudinal Studies; 2020.

35. Bauer GR, Churchill SM, Mahendran M, Walwyn C, Lizotte D, Villa-Rueda AA. Intersectionality in quantitative research: A systematic review of its emergence and applications of theory and methods. SSM Popul Health. Jun 2021;14:100798. doi:10.1016/j.ssmph.2021.100798

36. Evans CR, Williams DR, Onnela JP, Subramanian SV. A multilevel approach to modeling health inequalities at the intersection of multiple social identities. Soc Sci Med. Apr 2018;203:64–73. doi:10.1016/j.socscimed.2017.11.011

37. Merlo J. Multilevel analysis of individual heterogeneity and discriminatory accuracy (MAIHDA) within an intersectional framework. Soc Sci Med. Apr 2018;203:74–80. doi:10.1016/j.socscimed.2017.12.026

