## Supplementary Material for "Inequalities in mental and social wellbeing during the COVID-19 pandemic: prospective longitudinal observational study of five UK cohorts"

### Appendix S1. Flow diagram of the study sample.

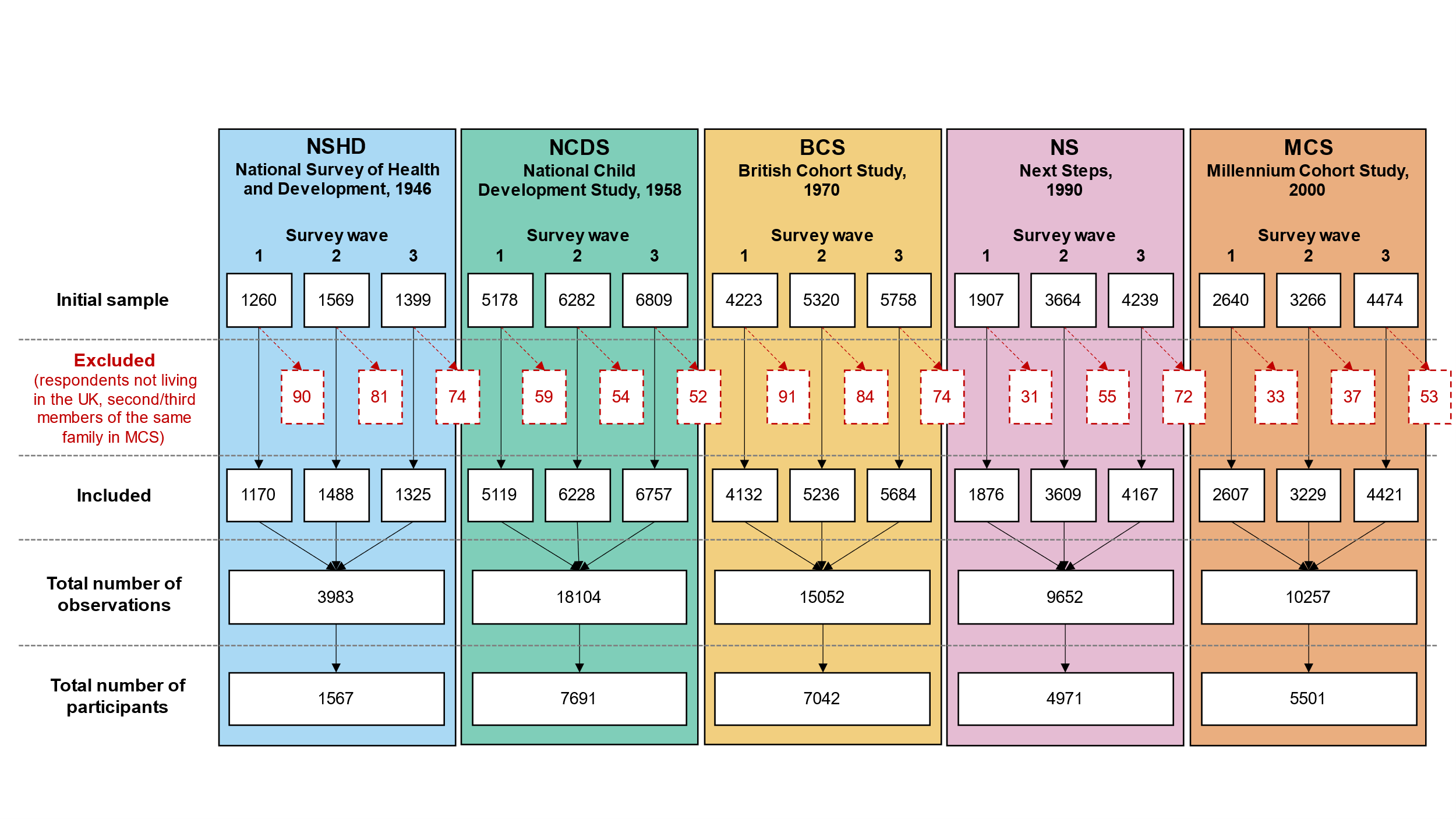

### Appendix S2. Details on the inverse probability weighting approach used.

To restore representativeness to the respective target populations due to the differential probability of participating in the COVID surveys, all models were estimated using an inverse probability weighting (IPW) approach. Where appropriate, non-response weights were combined with the corresponding survey design weights. Participants’ internet access was accounted for when deriving the weights to adjust for the potential sample selection effects induced by the survey administration mode (web survey).

Further information on the derivation and effectiveness of the weights can be found in the COVID-19 Survey User Guide.^1^

### Appendix S3. Details on variables included in the models as confounders.

The adjusted models included birth sex, highest qualification achieved (either academic or vocational, corresponding to the parents’ qualification in the case of MCS participants), pre-pandemic self-reported health, existence of psychological distress in the most recent pre-pandemic cohort assessment, and household composition as ‘a priori’ confounders.

Highest qualification was recoded into National Vocational Qualifications (NVQs) equivalents as reported by Dodgeon and Parsons.^2^ Data from the youngest cohort (MCS) corresponded to the parents’ highest qualification at the cohort members’ birth, as the degree of variability among cohort members was very small due to their age at the last pre-pandemic data collection (2018, 17 years old).

Self-reported health level prior to the pandemic onset was assessed as part of the COVID-19 survey study with the question “In general, would you say your health is 1) excellent, 2) very good, 3) good, 4) fair, or 5) poor?”. Responses were recoded so higher scores represented higher self-reported health levels. This variable was included as a continuous variable as this helped to simplify the estimation of the models without altering the results.

A binary variable representing the existence of pre-pandemic psychological distress was created using the measures available in the different cohort studies. These measures corresponded to the most recent pre-pandemic available assessment:

- NSHD, 2015, age 69, 28-item General Health Questionnaire (GHQ-28) score equal or greater than 5.
- NCDS, 2008, age 50, Malaise inventory score equal or greater than 4.
- BCS, 2016-2018, age 46, Malaise inventory score equal or greater than 4.
- NS, 2015, age 25/26, 12-item General Health Questionnaire (GHQ-12) score equal or greater than 4.
- MCS, 2018, age 17, 6-item Kessler Psychological Distress Scale (K-6) score equal or greater than 5.

Household composition was classified based on the people living with the respondent at the time of their first participation in the COVID-19 survey study. The four created categories were:

- Alone: if the person reported not living with anyone else.
- Partner: if the person reported living only with their partner.
- Partner and others: if the person reported living with their partner as well as with their children and/or parents.
- Other: if the person reported living in other kind of arrangement (including only with their children and/or parents and with other people.

To further explore the potential acceleration of the generational inequalities with the pandemic onset, models including an extended set of covariates (those from the adjusted models and early life cognitive ability, parental social class during childhood, and the existence of psychological distress in the second to most recent pre-pandemic cohort assessment) were estimated. These additional covariates were included to account for further sources of differences in the initial levels in the mental and social wellbeing outcomes, which were the focus of these analyses. These models did not include Next Steps (NS), as information on psychological distress was not available earlier than in the most recent pre-pandemic assessment for that cohort.

Early life cognitive ability was operationalised using the results from crystallised ability (verbal ability) at age 10-11 across all four cohorts included in this sub-analysis. The reason for choosing this subdomain of cognitive ability was the availability of assessments of a comparable construct at similar ages across the largest number of cohorts (see McElroy et al., 2021, pages 33 and 35).^3^ The measures used corresponded to the verbal test from the National Foundation for Educational Research (NFER) for NSHD and NCDS at age 11, the word similarities test from the British Ability Scales (BAS) for BCS at age 10, and the verbal similarities test from the BAS II for MCS at age 11. The scores were transformed into a common metric ranging from 0 (lowest verbal ability) to 50 (highest verbal ability). Further details on these measures are available in McElroy et al.^3^

Parental social class was assessed at ages 10/11 across the four cohorts included in this sub-analysis, representing the highest parental social class from “Professional” to “Unskilled”. Further details on the harmonisation of this variable is available in Dodgeon et al.^4^

A binary variable representing the existence of psychological distress in the second to most recent pre-pandemic assessment was obtained for each of the four cohorts included in this sub-analysis:

- NSHD, 2009, age 63, GHQ-28 score equal or greater than 5.
- NCDS, 2000, age 42, Malaise inventory score equal or greater than 4.
- BCS, 2012, age 42, Malaise inventory score equal or greater than 4.
- MCS, 2015, age 14, Short mood and Feelings Questionnaire (SMFQ) score equal or greater than 12.

### Appendix S4. Details on the analytical approach used.

#### Appendix S4.1. Multilevel growth curve models.

We used a multilevel growth curve modelling approach to analyse differences in the initial levels (at the first survey wave) and change over time (throughout the two additional survey waves) across subgroups in the different outcomes under study. In this approach, repeated observations of the outcomes (level 1) are nested within the corresponding individuals (level 2), allowing to simultaneously explore variables related to within- and between-person variability while adjusting for potential confounders.

As predictors of the between-person variability, we used the cohort/birth year to account for generational differences in the initial levels; the additional subgrouping variables (birth sex, pre-pandemic financial situation, relationship status, housing tenure, urbanicity, UK country of residence, or ethnicity) to account for between-groups differences in the initial levels, and the interaction among cohort/birth year and the subgrouping variables to account for the interplay between generational effects and the corresponding subgroups in the initial levels.

As predictors of the within-person variability, we used linear and quadratic time terms in order to account for potential curvilinear trends, as well as the interaction between the time terms and the cohort/birth year (to account for generational differences in the change over time), the time terms and the subgrouping variable under study (to account for between-groups differences in the change over time), and the time terms and the interaction between cohort/birth year and the subgrouping variable under study (to account for the interplay between generational effects and the corresponding subgroups in the change over time).

In other words, models studying a subgrouping variable included the three-way interaction terms “time (linear and quadratic) * cohort/birth year * subgrouping variable”.

Separate sets of models were estimated for each outcome and for each subgrouping variable, starting with a set of models with cohort/birth year as the only subgrouping variable. Unadjusted and adjusted (including birth sex, highest qualification, pre-pandemic self-reported health and psychological distress, and household composition as covariates) models were estimated.

All these variables were included in the fixed-effects part of the multilevel models. The random-effects part included the variability in the initial levels (random intercepts) and in the linear change over time (random slopes). These two random effects were allowed to covary freely. Random quadratic slopes were not modelled as their inclusion led to estimation problems.

The overall statistical significance of the main effects of cohort/birth year and the subgrouping variables with more than two levels (i.e., pre-pandemic financial situation and ethnicity), as well as the interaction terms they were part of, were tested by means of the Wald chi-squared test. Marginal mean estimates and 95% CIs of the outcomes were obtained from each of the models (unadjusted and adjusted) and plotted by the different subgroups.

Some subgroups within the NSHD data (i.e., pre-pandemic financial difficulties, not in a relationship, and rented/rent-free/other) were not included in the plots due to the small number of cases in those combinations and subsequent extremely large CIs. Similarly, NS data from all countries but England, data on participants living in Northern Ireland except for those from the MCS cohort, and NSHD data from Wales were not included in the models to safeguard anonymity.

#### Appendix S4.2. Exploration of accelerated generational inequalities in the initial outcome levels.

To further explore the existence of accelerated generational inequalities in the initial outcome levels, we estimated an additional set of models to answer the counterfactual question of when MCS participants ‘should’ have been born to have their mental and social wellbeing initial levels, provided that generational inequalities had not accelerated (in other words, what birth year more closely resembled the marginal mean levels predicted for the MCS cohort if generational inequalities were linear).

To do so, these models were estimated excluding MCS data and using birth year (centred at 1946, the birth year of NSHD cohort) as a continuous variable to explore linear generational inequalities instead of cohort as a categorical variable. Marginal predictions were obtained from these models for every birth year from 1946 to 2020 and were then compared to those from the original models to identify the birth year that provided the closest match to each of the cohorts. Both unadjusted and adjusted models were computed. Models including an extended set of covariates (early life cognitive ability, parental social class during childhood, and the existence of psychological distress in the second to most recent pre-pandemic cohort assessment) were estimated to account for further sources of differences in the initial levels. These latter models did not include NS, as information on psychological distress was not available earlier than in the most recent pre-pandemic assessment. Further details on these variables can be found in the **Appendix S3**.

### Appendix S5. Results by cohort/birth year.

#### Table S5.1. Results of multilevel growth curve models by cohort/birth year.

|  | **Anxiety symptomatology (GAD-2)** | | | **Depressive symptomatology (PHQ-2)** | | | **Feelings of loneliness (UCLA-3)** | | | **Life satisfaction (ONS single question)** | | |
| --- | --- | --- | --- | --- | --- | --- | --- | --- | --- | --- | --- | --- |
| **Unadjusted models** |  |  |  |  |  |  |  |  |  |  |  |  |
| N participants | 25,635 |  |  | 25,633 |  |  | 25,664 |  |  | 25,722 |  |  |
| N observations | 54,140 |  |  | 54,123 |  |  | 54,208 |  |  | 54,444 |  |  |
|  | ***B* (95% CI)** | **χ2** | ***p*** | ***B* (95% CI)** | **χ2** | ***p*** | ***B* (95% CI)** | **χ2** | ***p*** | ***B* (95% CI)** | **χ2** | ***p*** |
| Time (linear) | 0.08 (0.02, 0.14) |  | 0.005 | -0.04 (-0.09, 0.01) |  | 0.139 | -0.28 (-0.34, -0.22) |  | <0.001 | 0.13 (0.04, 0.21) |  | 0.003 |
| Time (quadratic) | -0.03 (-0.06, -0.01) |  | 0.014 | 0.05 (0.03, 0.08) |  | <0.001 | 0.18 (0.15, 0.21) |  | <0.001 | -0.19 (-0.23, -0.15) |  | <0.001 |
| Cohort (ref. NCDS) |  | 1107.6 (4) | <0.001 |  | 1472.1 (4) | <0.001 |  | 823.3 (4) | <0.001 |  | 698.7 (4) | <0.001 |
| NSHD | -0.25 (-0.32, -0.18) |  |  | -0.21 (-0.28, -0.15) |  |  | -0.16 (-0.26, -0.07) |  |  | 0.10 (-0.03, 0.23) |  |  |
| BCS | 0.19 (0.14, 0.25) |  |  | 0.23 (0.17, 0.28) |  |  | 0.08 (0.02, 0.14) |  |  | -0.25 (-0.33, -0.17) |  |  |
| NS | 0.73 (0.65, 0.81) |  |  | 0.75 (0.68, 0.83) |  |  | 0.50 (0.42, 0.58) |  |  | -0.65 (-0.75, -0.54) |  |  |
| MCS | 1.03 (0.95, 1.11) |  |  | 1.23 (1.16, 1.31) |  |  | 1.02 (0.94, 1.10) |  |  | -1.30 (-1.41, -1.20) |  |  |
| Linear change * cohort (ref. NCDS) |  | 24.7 (4) | <0.001 |  | 50.5 (4) | <0.001 |  | 41 (4) | <0.001 |  | 32.7 (4) | <0.001 |
| NSHD | 0.16 (0.04, 0.28) |  |  | 0.07 (-0.04, 0.19) |  |  | -0.08 (-0.22, 0.07) |  |  | -0.07 (-0.28, 0.15) |  |  |
| BCS | 0.10 (0.01, 0.20) |  |  | -0.06 (-0.15, 0.02) |  |  | 0.24 (0.14, 0.34) |  |  | -0.12 (-0.25, 0.01) |  |  |
| NS | 0.11 (-0.03, 0.25) |  |  | -0.25 (-0.38, -0.12) |  |  | 0.21 (0.08, 0.34) |  |  | 0.21 (0.03, 0.38) |  |  |
| MCS | 0.34 (0.19, 0.48) |  |  | -0.43 (-0.57, -0.28) |  |  | 0.29 (0.15, 0.44) |  |  | 0.38 (0.20, 0.57) |  |  |
| Quadratic change * cohort (ref. NCDS) |  | 11.3 (4) | 0.024 |  | 53.7 (4) | <0.001 |  | 49.2 (4) | <0.001 |  | 23.0 (4) | <0.001 |
| NSHD | -0.06 (-0.11, 0.00) |  |  | -0.05 (-0.10, 0.01) |  |  | 0.04 (-0.03, 0.10) |  |  | 0.01 (-0.09, 0.11) |  |  |
| BCS | -0.04 (-0.08, 0.00) |  |  | 0.03 (-0.01, 0.07) |  |  | -0.11 (-0.16, -0.07) |  |  | 0.08 (0.02, 0.14) |  |  |
| NS | -0.03 (-0.09, 0.04) |  |  | 0.11 (0.05, 0.16) |  |  | -0.10 (-0.16, -0.04) |  |  | -0.05 (-0.12, 0.03) |  |  |
| MCS | -0.10 (-0.16, -0.03) |  |  | 0.20 (0.14, 0.27) |  |  | -0.17 (-0.23, -0.10) |  |  | -0.12 (-0.20, -0.04) |  |  |
| **Adjusted models** |  |  |  |  |  |  |  |  |  |  |  |  |
| N participants | 21,773 |  |  | 21,770 |  |  | 21,798 |  |  | 21,844 |  |  |
| N observations | 47,034 |  |  | 47,022 |  |  | 47,087 |  |  | 47,288 |  |  |
|  | ***B* (95% CI)** | **χ2** | ***p*** | ***B* (95% CI)** | **χ2** | ***p*** | ***B* (95% CI)** | **χ2** | ***p*** | ***B* (95% CI)** | **χ2** | ***p*** |
| Time (linear) | 0.09 (0.03, 0.15) |  | 0.005 | -0.07 (-0.12, -0.01) |  | 0.026 | -0.28 (-0.35, -0.22) |  | <0.001 | 0.12 (0.04, 0.21) |  | 0.006 |
| Time (quadratic) | -0.03 (-0.06, -0.01) |  | 0.017 | 0.06 (0.04, 0.09) |  | <0.001 | 0.18 (0.15, 0.21) |  | <0.001 | -0.19 (-0.23, -0.15) |  | <0.001 |
| Cohort (ref. NCDS) |  | 515.4 (4) | <0.001 |  | 708.2 (4) | <0.001 |  | 164.5 (4) | <0.001 |  | 158.5 (4) | <0.001 |
| NSHD | -0.19 (-0.26, -0.12) |  |  | -0.18 (-0.25, -0.11) |  |  | -0.06 (-0.16, 0.04) |  |  | -0.05 (-0.19, 0.08) |  |  |
| BCS | 0.14 (0.09, 0.20) |  |  | 0.19 (0.13, 0.24) |  |  | 0.06 (-0.01, 0.12) |  |  | -0.22 (-0.31, -0.14) |  |  |
| NS | 0.57 (0.48, 0.65) |  |  | 0.61 (0.53, 0.69) |  |  | 0.34 (0.26, 0.42) |  |  | -0.44 (-0.55, -0.33) |  |  |
| MCS | 0.90 (0.80, 0.99) |  |  | 1.04 (0.95, 1.13) |  |  | 0.55 (0.45, 0.65) |  |  | -0.74 (-0.87, -0.61) |  |  |
| Linear change * cohort (ref. NCDS) |  | 27.2 (4) | <0.001 |  | 31.1 (4) | <0.001 |  | 38.8 (4) | <0.001 |  | 23.1 (4) | <0.001 |
| NSHD | 0.19 (0.05, 0.32) |  |  | 0.11 (-0.01, 0.24) |  |  | -0.08 (-0.24, 0.08) |  |  | -0.06 (-0.31, 0.18) |  |  |
| BCS | 0.12 (0.01, 0.22) |  |  | -0.04 (-0.14, 0.06) |  |  | 0.22 (0.11, 0.33) |  |  | -0.09 (-0.23, 0.05) |  |  |
| NS | 0.14 (-0.01, 0.29) |  |  | -0.19 (-0.33, -0.05) |  |  | 0.20 (0.06, 0.34) |  |  | 0.20 (0.02, 0.39) |  |  |
| MCS | 0.38 (0.22, 0.53) |  |  | -0.34 (-0.49, -0.18) |  |  | 0.37 (0.22, 0.53) |  |  | 0.36 (0.16, 0.55) |  |  |
| Quadratic change * cohort (ref. NCDS) |  | 13.6 (4) | 0.008 |  | 34.0 (4) | <0.001 |  | 44.5 (4) | <0.001 |  | 17.7 (4) | 0.001 |
| NSHD | -0.07 (-0.14, 0.00) |  |  | -0.06 (-0.12, 0.00) |  |  | 0.03 (-0.05, 0.11) |  |  | 0.01 (-0.12, 0.13) |  |  |
| BCS | -0.06 (-0.11, -0.01) |  |  | 0.01 (-0.03, 0.06) |  |  | -0.11 (-0.16, -0.06) |  |  | 0.07 (0.00, 0.14) |  |  |
| NS | -0.05 (-0.11, 0.02) |  |  | 0.07 (0.01, 0.14) |  |  | -0.10 (-0.16, -0.03) |  |  | -0.04 (-0.13, 0.04) |  |  |
| MCS | -0.11 (-0.18, -0.04) |  |  | 0.17 (0.10, 0.24) |  |  | -0.20 (-0.27, -0.12) |  |  | -0.12 (-0.21, -0.03) |  |  |
| **Sensitivity models** |  |  |  |  |  |  |  |  |  |  |  |  |
| N participants | 21,773 |  |  | 21,770 |  |  | 21,798 |  |  | 21,844 |  |  |
| N observations | 47,034 |  |  | 47,022 |  |  | 47,087 |  |  | 47,288 |  |  |
|  | ***B* (95% CI)** | **χ2** | ***p*** | ***B* (95% CI)** | **χ2** | ***p*** | ***B* (95% CI)** | **χ2** | ***p*** | ***B* (95% CI)** | **χ2** | ***p*** |
| Time (linear) | 0.09 (0.03, 0.15) |  | 0.003 | -0.06 (-0.12, 0.00) |  | 0.050 | -0.27 (-0.34, -0.21) |  | <0.001 | 0.11 (0.02, 0.20) |  | 0.014 |
| Time (quadratic) | -0.04 (-0.06, -0.01) |  | 0.012 | 0.06 (0.04, 0.09) |  | <0.001 | 0.18 (0.15, 0.21) |  | <0.001 | -0.18 (-0.23, -0.14) |  | <0.001 |
| Cohort (ref. NCDS) |  | 973.4 (4) | <0.001 |  | 1254.2 (4) | <0.001 |  | 701.0 (4) | <0.001 |  | 598.3 (4) | <0.001 |
| NSHD | -0.24 (-0.31, -0.16) |  |  | -0.21 (-0.28, -0.14) |  |  | -0.14 (-0.24, -0.04) |  |  | 0.07 (-0.07, 0.21) |  |  |
| BCS | 0.19 (0.13, 0.25) |  |  | 0.21 (0.15, 0.26) |  |  | 0.05 (-0.01, 0.12) |  |  | -0.25 (-0.33, -0.16) |  |  |
| NS | 0.73 (0.64, 0.81) |  |  | 0.74 (0.65, 0.82) |  |  | 0.50 (0.41, 0.58) |  |  | -0.64 (-0.75, -0.53) |  |  |
| MCS | 1.04 (0.96, 1.12) |  |  | 1.20 (1.12, 1.28) |  |  | 1.00 (0.92, 1.08) |  |  | -1.26 (-1.37, -1.15) |  |  |
| Linear change * cohort (ref. NCDS) |  | 24.4 (4) | <0.001 |  | 31.0 (4) | <0.001 |  | 36.7 (4) | <0.001 |  | 21.8 (4) | <0.001 |
| NSHD | 0.18 (0.04, 0.31) |  |  | 0.11 (-0.02, 0.23) |  |  | -0.09 (-0.25, 0.08) |  |  | -0.05 (-0.30, 0.20) |  |  |
| BCS | 0.11 (0.00, 0.21) |  |  | -0.05 (-0.15, 0.05) |  |  | 0.21 (0.10, 0.32) |  |  | -0.07 (-0.21, 0.07) |  |  |
| NS | 0.13 (-0.02, 0.28) |  |  | -0.20 (-0.34, -0.06) |  |  | 0.21 (0.07, 0.35) |  |  | 0.21 (0.02, 0.39) |  |  |
| MCS | 0.36 (0.21, 0.52) |  |  | -0.35 (-0.51, -0.19) |  |  | 0.36 (0.20, 0.52) |  |  | 0.36 (0.16, 0.56) |  |  |
| Quadratic change * cohort (ref. NCDS) |  | 12.9 (4) | 0.012 |  | 32.6 (4) | <0.001 |  | 43.9 (4) | <0.001 |  | 15.2 (4) | 0.004 |
| NSHD | -0.07 (-0.13, 0.00) |  |  | -0.06 (-0.12, 0.00) |  |  | 0.03 (-0.05, 0.11) |  |  | 0.00 (-0.12, 0.13) |  |  |
| BCS | -0.05 (-0.10, -0.01) |  |  | 0.02 (-0.03, 0.06) |  |  | -0.11 (-0.16, -0.06) |  |  | 0.06 (0.00, 0.13) |  |  |
| NS | -0.04 (-0.11, 0.02) |  |  | 0.08 (0.01, 0.14) |  |  | -0.10 (-0.16, -0.04) |  |  | -0.05 (-0.13, 0.04) |  |  |
| MCS | -0.11 (-0.19, -0.04) |  |  | 0.17 (0.09, 0.24) |  |  | -0.20 (-0.27, -0.12) |  |  | -0.11 (-0.21, -0.02) |  |  |

*Note.* Adjusted models included birth sex, highest qualification achieved, pre-pandemic self-reported health, pre-pandemic psychological distress, and household composition as covariates. Sensitivity models correspond to the unadjusted models after restricting the analytical sample to that of the adjusted models. BCS: British Cohort Study, 1970 birth cohort; GAD-2: 2-item General Anxiety Disorder questionnaire; MCS: Millennium Cohort Study, 2000 birth cohort; NCDS: National Child and Development Study, 1958 birth cohort; NS: Next Steps, 1990 cohort; NSHD: National Survey of Health and Development, 1946 birth cohort; ONS: UK Office for National Statistics; PHQ-2: 2-item Patient Health Questionnaire; UCLA-3: 3-item UCLA loneliness scale. χ2: Wald test performed to assess the overall statistical significance of the interaction terms; all χ2 statistics in this table have 4 degrees of freedom.

#### Table S5.2. Unadjusted and adjusted marginal mean estimates and 95% confidence intervals by cohort/birth year.

|  |  | **Anxiety symptomatology (GAD-2)** | **Depressive symptomatology (PHQ-2)** | **Feelings of loneliness (UCLA-3)** | **Life satisfaction (ONS single question)** |
| --- | --- | --- | --- | --- | --- |
| **Cohort** | **Survey wave** | **Unadjusted marginal mean (95% CI)** | **Unadjusted marginal mean (95% CI)** | **Unadjusted marginal mean (95% CI)** | **Unadjusted marginal mean (95% CI)** |
| NSHD | 1 | 0.49 (0.43, 0.55) | 0.46 (0.40, 0.51) | 3.98 (3.90, 4.06) | 7.56 (7.44, 7.68) |
| NSHD | 2 | 0.64 (0.58, 0.70) | 0.50 (0.45, 0.55) | 3.84 (3.77, 3.91) | 7.44 (7.34, 7.55) |
| NSHD | 3 | 0.60 (0.54, 0.66) | 0.55 (0.50, 0.61) | 4.14 (4.06, 4.22) | 6.97 (6.85, 7.08) |
| NCDS | 1 | 0.75 (0.71, 0.78) | 0.67 (0.64, 0.70) | 4.14 (4.10, 4.18) | 7.46 (7.41, 7.52) |
| NCDS | 2 | 0.79 (0.76, 0.83) | 0.68 (0.65, 0.71) | 4.04 (4.01, 4.08) | 7.40 (7.35, 7.45) |
| NCDS | 3 | 0.78 (0.74, 0.81) | 0.80 (0.77, 0.83) | 4.31 (4.27, 4.35) | 6.96 (6.91, 7.01) |
| BCS | 1 | 0.94 (0.89, 0.98) | 0.90 (0.86, 0.94) | 4.22 (4.17, 4.27) | 7.21 (7.15, 7.27) |
| BCS | 2 | 1.05 (1.01, 1.09) | 0.88 (0.84, 0.92) | 4.25 (4.20, 4.29) | 7.11 (7.06, 7.16) |
| BCS | 3 | 1.01 (0.97, 1.05) | 1.03 (0.99, 1.07) | 4.42 (4.37, 4.46) | 6.78 (6.72, 6.83) |
| NS | 1 | 1.48 (1.40, 1.55) | 1.42 (1.35, 1.49) | 4.64 (4.57, 4.71) | 6.82 (6.73, 6.91) |
| NS | 2 | 1.61 (1.55, 1.67) | 1.30 (1.24, 1.35) | 4.65 (4.60, 4.71) | 6.91 (6.85, 6.98) |
| NS | 3 | 1.62 (1.57, 1.68) | 1.49 (1.44, 1.54) | 4.83 (4.78, 4.89) | 6.53 (6.47, 6.60) |
| MCS | 1 | 1.78 (1.71, 1.85) | 1.90 (1.83, 1.97) | 5.16 (5.09, 5.23) | 6.16 (6.07, 6.25) |
| MCS | 2 | 2.07 (2.00, 2.13) | 1.69 (1.63, 1.75) | 5.19 (5.13, 5.26) | 6.36 (6.29, 6.44) |
| MCS | 3 | 2.10 (2.05, 2.16) | 1.99 (1.93, 2.04) | 5.26 (5.20, 5.31) | 5.94 (5.88, 6.01) |
| **Cohort** | **Survey wave** | **Adjusted marginal mean (95% CI)** | **Adjusted marginal mean (95% CI)** | **Adjusted marginal mean (95% CI)** | **Adjusted marginal mean (95% CI)** |
| NSHD | 1 | 0.59 (0.53, 0.66) | 0.57 (0.50, 0.63) | 4.20 (4.11, 4.29) | 7.28 (7.15, 7.40) |
| NSHD | 2 | 0.76 (0.69, 0.83) | 0.62 (0.55, 0.68) | 4.05 (3.96, 4.13) | 7.16 (7.03, 7.28) |
| NSHD | 3 | 0.72 (0.65, 0.80) | 0.66 (0.60, 0.73) | 4.32 (4.22, 4.42) | 6.67 (6.53, 6.81) |
| NCDS | 1 | 0.78 (0.74, 0.82) | 0.74 (0.71, 0.78) | 4.26 (4.21, 4.30) | 7.33 (7.27, 7.39) |
| NCDS | 2 | 0.83 (0.80, 0.87) | 0.74 (0.71, 0.78) | 4.16 (4.12, 4.20) | 7.27 (7.22, 7.32) |
| NCDS | 3 | 0.82 (0.78, 0.85) | 0.87 (0.84, 0.91) | 4.42 (4.38, 4.47) | 6.83 (6.78, 6.89) |
| BCS | 1 | 0.92 (0.88, 0.97) | 0.93 (0.89, 0.98) | 4.32 (4.26, 4.37) | 7.11 (7.04, 7.17) |
| BCS | 2 | 1.04 (0.99, 1.08) | 0.90 (0.86, 0.95) | 4.32 (4.28, 4.37) | 7.03 (6.96, 7.09) |
| BCS | 3 | 0.96 (0.92, 1.01) | 1.03 (0.99, 1.07) | 4.48 (4.43, 4.53) | 6.71 (6.65, 6.77) |
| NS | 1 | 1.35 (1.27, 1.42) | 1.36 (1.29, 1.43) | 4.60 (4.53, 4.67) | 6.89 (6.80, 6.98) |
| NS | 2 | 1.49 (1.43, 1.56) | 1.24 (1.18, 1.30) | 4.60 (4.54, 4.66) | 6.98 (6.91, 7.06) |
| NS | 3 | 1.48 (1.43, 1.54) | 1.40 (1.35, 1.46) | 4.78 (4.72, 4.83) | 6.62 (6.55, 6.69) |
| MCS | 1 | 1.68 (1.60, 1.76) | 1.79 (1.71, 1.86) | 4.80 (4.72, 4.89) | 6.59 (6.48, 6.70) |
| MCS | 2 | 2.00 (1.92, 2.08) | 1.61 (1.54, 1.69) | 4.88 (4.80, 4.96) | 6.76 (6.67, 6.86) |
| MCS | 3 | 2.03 (1.95, 2.10) | 1.91 (1.84, 1.98) | 4.94 (4.86, 5.01) | 6.32 (6.23, 6.41) |

*Note.* Adjusted models included birth sex, highest qualification achieved, pre-pandemic self-reported health, pre-pandemic psychological distress, and household composition as covariates. BCS: British Cohort Study, 1970 birth cohort; GAD-2: 2-item General Anxiety Disorder questionnaire; MCS: Millennium Cohort Study, 2000 birth cohort; NCDS: National Child and Development Study, 1958 birth cohort; NS: Next Steps, 1990 cohort; NSHD: National Survey of Health and Development, 1946 birth cohort; ONS: UK Office for National Statistics; PHQ-2: 2-item Patient Health Questionnaire; UCLA-3: 3-item UCLA loneliness scale. Survey wave 1: May 2020; survey wave 2: September/October 2020; survey wave 3: February/March 2021.

#### Figure S5.1. Comparison between marginal predicted initial levels by cohort and marginal predicted initial levels by birth year assuming no acceleration in the differences by birth year and excluding data from the youngest cohort (MCS). Unadjusted models.

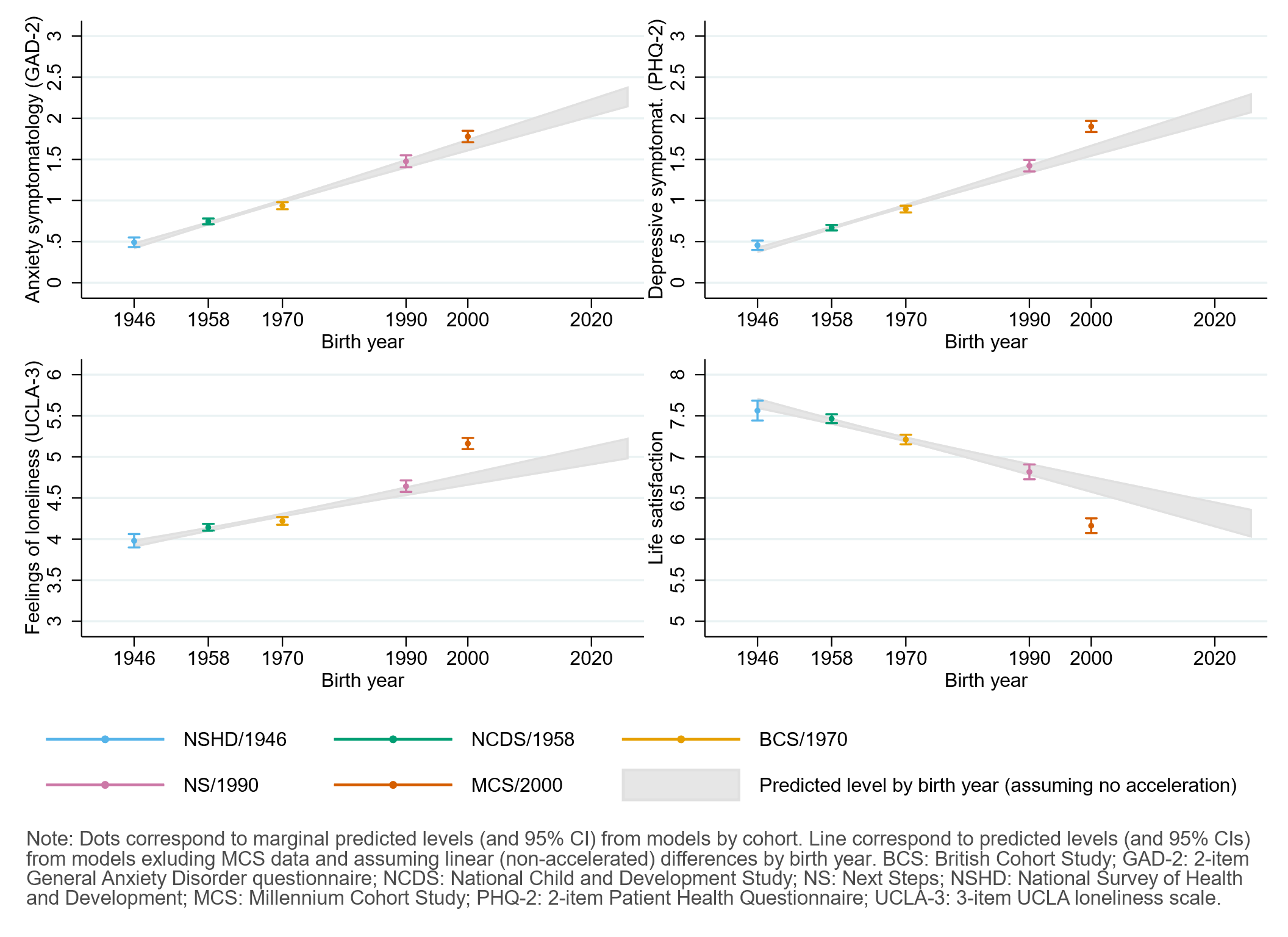

#### Figure S5.2. Comparison between marginal predicted initial levels by cohort and marginal predicted initial levels by birth year assuming no acceleration in the differences by birth year and excluding data from the youngest cohort (MCS). Models adjusted by birth sex, highest qualification achieved, pre-pandemic self-reported health, household composition, early life cognitive ability (verbal), parental social class at childhood, and both most and second-to-most recent pre-pandemic assessments of psychological distress.

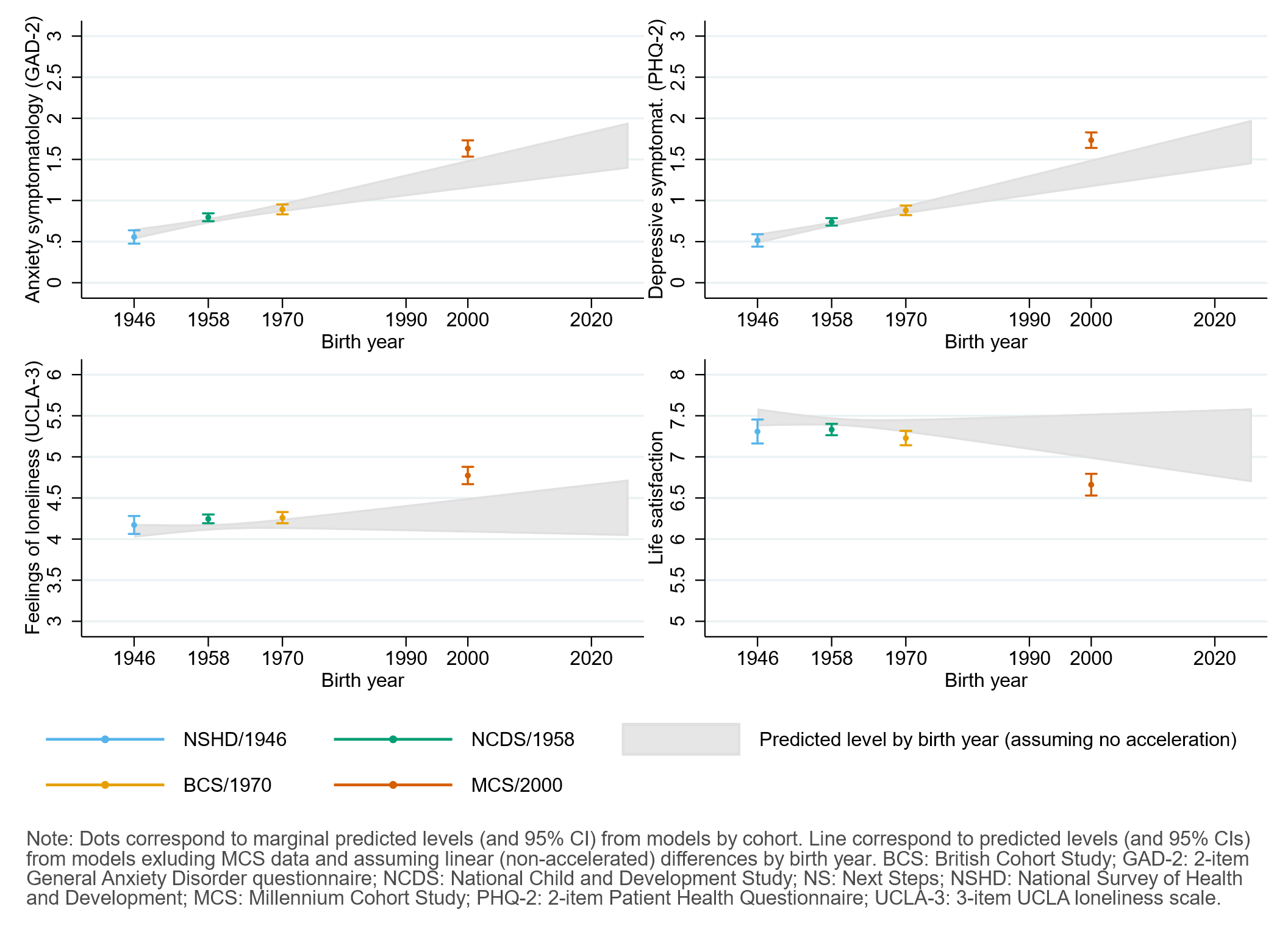

### Appendix S6. Results by birth sex.

#### Table S6.1. Results of multilevel growth curve models by birth sex.

|  | **Anxiety symptomatology (GAD-2)** | | | **Depressive symptomatology (PHQ-2)** | | | **Feelings of loneliness (UCLA-3)** | | | **Life satisfaction (ONS single question)** | | |
| --- | --- | --- | --- | --- | --- | --- | --- | --- | --- | --- | --- | --- |
| **Unadjusted models** |  |  |  |  |  |  |  |  |  |  |  |  |
| N participants | 25,635 |  |  | 25,633 |  |  | 25,664 |  |  | 25,722 |  |  |
| N observations | 54,140 |  |  | 54,123 |  |  | 54,208 |  |  | 54,444 |  |  |
|  | ***B* (95% CI)** | **χ2** | ***p*** | ***B* (95% CI)** | **χ2** | ***p*** | ***B* (95% CI)** | **χ2** | ***p*** | ***B* (95% CI)** | **χ2** | ***p*** |
| Time (linear) | 0.05 (-0.02, 0.12) |  | 0.157 | -0.01 (-0.08, 0.06) |  | 0.719 | -0.23 (-0.31, -0.14) |  | <0.001 | 0.09 (-0.02, 0.21) |  | 0.113 |
| Time (quadratic) | -0.02 (-0.06, 0.01) |  | 0.192 | 0.04 (0.01, 0.07) |  | 0.020 | 0.15 (0.11, 0.19) |  | <0.001 | -0.15 (-0.21, -0.10) |  | <0.001 |
| Cohort (ref. NCDS) |  | 278.7 | <0.001 |  | 456.5 | <0.001 |  | 343.2 | <0.001 |  | 220.2 | <0.001 |
| NSHD | -0.23 (-0.31, -0.15) |  | <0.001 | -0.21 (-0.29, -0.14) |  | <0.001 | -0.19 (-0.31, -0.08) |  | 0.001 | 0.17 (-0.02, 0.36) |  | 0.073 |
| BCS | 0.21 (0.13, 0.28) |  | <0.001 | 0.27 (0.19, 0.34) |  | <0.001 | 0.09 (0.00, 0.18) |  | 0.053 | -0.31 (-0.43, -0.19) |  | <0.001 |
| NS | 0.55 (0.43, 0.67) |  | <0.001 | 0.72 (0.60, 0.84) |  | <0.001 | 0.60 (0.46, 0.73) |  | <0.001 | -0.68 (-0.85, -0.52) |  | <0.001 |
| MCS | 0.65 (0.53, 0.76) |  | <0.001 | 0.89 (0.78, 1.01) |  | <0.001 | 1.05 (0.92, 1.19) |  | <0.001 | -1.11 (-1.29, -0.94) |  | <0.001 |
| Linear change * cohort (ref. NCDS) |  | 17.4 | 0.002 |  | 1.6 | 0.809 |  | 25.1 | <0.001 |  | 2.9 | 0.569 |
| NSHD | 0.15 (-0.01, 0.30) |  | 0.058 | 0.02 (-0.11, 0.15) |  | 0.758 | -0.10 (-0.28, 0.07) |  | 0.249 | 0.00 (-0.31, 0.31) |  | 0.986 |
| BCS | 0.10 (-0.04, 0.23) |  | 0.174 | 0.00 (-0.13, 0.12) |  | 0.961 | 0.23 (0.09, 0.37) |  | 0.002 | -0.12 (-0.31, 0.06) |  | 0.197 |
| NS | 0.25 (0.03, 0.46) |  | 0.022 | 0.01 (-0.19, 0.22) |  | 0.894 | 0.30 (0.08, 0.52) |  | 0.006 | -0.10 (-0.36, 0.16) |  | 0.447 |
| MCS | 0.40 (0.18, 0.63) |  | <0.001 | -0.13 (-0.35, 0.09) |  | 0.252 | 0.34 (0.10, 0.59) |  | 0.005 | 0.10 (-0.21, 0.40) |  | 0.532 |
| Quadratic change * cohort (ref. NCDS) |  | 7.9 | 0.1 |  | 4.2 | 0.384 |  | 27 | <0.001 |  | 5.9 | 0.209 |
| NSHD | -0.05 (-0.13, 0.02) |  | 0.135 | -0.02 (-0.08, 0.04) |  | 0.449 | 0.05 (-0.03, 0.13) |  | 0.249 | -0.03 (-0.17, 0.12) |  | 0.727 |
| BCS | -0.04 (-0.10, 0.02) |  | 0.201 | 0.00 (-0.06, 0.06) |  | 0.943 | -0.09 (-0.15, -0.02) |  | 0.008 | 0.07 (-0.01, 0.16) |  | 0.087 |
| NS | -0.08 (-0.17, 0.01) |  | 0.092 | -0.02 (-0.11, 0.07) |  | 0.651 | -0.15 (-0.25, -0.06) |  | 0.002 | 0.09 (-0.02, 0.21) |  | 0.113 |
| MCS | -0.11 (-0.22, -0.01) |  | 0.033 | 0.09 (-0.01, 0.19) |  | 0.093 | -0.18 (-0.30, -0.07) |  | 0.001 | -0.03 (-0.16, 0.11) |  | 0.699 |
| Birth sex (ref. Men) | 0.46 (0.39, 0.53) |  | <0.001 | 0.28 (0.21, 0.35) |  | <0.001 | 0.39 (0.30, 0.47) |  | <0.001 | -0.22 (-0.33, -0.11) |  | <0.001 |
| Linear change * birth sex (ref. Men) | 0.05 (-0.06, 0.17) |  | 0.357 | -0.05 (-0.16, 0.05) |  | 0.323 | -0.10 (-0.23, 0.02) |  | 0.106 | 0.07 (-0.10, 0.24) |  | 0.430 |
| Quadratic change * birth sex (ref. Men) | -0.02 (-0.07, 0.03) |  | 0.437 | 0.03 (-0.02, 0.08) |  | 0.250 | 0.07 (0.01, 0.13) |  | 0.018 | -0.07 (-0.15, 0.01) |  | 0.069 |
| Cohort (ref. NCDS) * birth sex (ref. Men) |  | 67.8 | <0.001 |  | 52.1 | <0.001 |  | 8.7 | 0.07 |  | 12.7 | 0.013 |
| NSHD | -0.05 (-0.18, 0.09) |  | 0.485 | 0.00 (-0.13, 0.13) |  | 0.980 | 0.06 (-0.12, 0.24) |  | 0.542 | -0.14 (-0.40, 0.12) |  | 0.302 |
| BCS | -0.06 (-0.17, 0.05) |  | 0.316 | -0.09 (-0.20, 0.02) |  | 0.094 | -0.05 (-0.17, 0.08) |  | 0.471 | 0.11 (-0.05, 0.27) |  | 0.160 |
| NS | 0.22 (0.06, 0.38) |  | 0.008 | 0.01 (-0.15, 0.17) |  | 0.867 | -0.21 (-0.38, -0.05) |  | 0.013 | 0.09 (-0.12, 0.31) |  | 0.401 |
| MCS | 0.56 (0.40, 0.71) |  | <0.001 | 0.49 (0.34, 0.64) |  | <0.001 | -0.12 (-0.28, 0.05) |  | 0.170 | -0.25 (-0.47, -0.04) |  | 0.023 |
| Linear change * cohort (ref. NCDS) *  birth sex (ref. Men) |  | 3.0 | 0.555 |  | 18.8 | <0.001 |  | 1.6 | 0.817 |  | 13.1 | 0.011 |
| NSHD | 0.01 (-0.22, 0.25) |  | 0.904 | 0.10 (-0.12, 0.32) |  | 0.364 | 0.05 (-0.23, 0.33) |  | 0.738 | -0.12 (-0.55, 0.31) |  | 0.584 |
| BCS | 0.01 (-0.17, 0.20) |  | 0.880 | -0.10 (-0.28, 0.08) |  | 0.263 | 0.02 (-0.18, 0.22) |  | 0.822 | 0.00 (-0.25, 0.26) |  | 0.973 |
| NS | -0.22 (-0.50, 0.05) |  | 0.115 | -0.41 (-0.68, -0.15) |  | 0.002 | -0.14 (-0.41, 0.14) |  | 0.326 | 0.49 (0.15, 0.83) |  | 0.005 |
| MCS | -0.07 (-0.36, 0.22) |  | 0.645 | -0.42 (-0.71, -0.13) |  | 0.004 | -0.06 (-0.36, 0.25) |  | 0.718 | 0.42 (0.03, 0.80) |  | 0.034 |
| Quadratic change * cohort (ref. NCDS) *  birth sex (ref. Men) |  | 2.4 | 0.671 |  | 18.7 | <0.001 |  | 3.9 | 0.416 |  | 12 | 0.017 |
| NSHD | -0.01 (-0.12, 0.10) |  | 0.887 | -0.04 (-0.15, 0.06) |  | 0.410 | -0.02 (-0.15, 0.11) |  | 0.752 | 0.07 (-0.13, 0.27) |  | 0.505 |
| BCS | 0.00 (-0.09, 0.08) |  | 0.961 | 0.05 (-0.03, 0.13) |  | 0.227 | -0.05 (-0.14, 0.05) |  | 0.323 | 0.01 (-0.11, 0.12) |  | 0.922 |
| NS | 0.09 (-0.03, 0.22) |  | 0.155 | 0.20 (0.08, 0.32) |  | 0.001 | 0.08 (-0.04, 0.20) |  | 0.201 | -0.22 (-0.37, -0.07) |  | 0.004 |
| MCS | 0.02 (-0.11, 0.16) |  | 0.757 | 0.17 (0.03, 0.30) |  | 0.014 | 0.02 (-0.12, 0.16) |  | 0.769 | -0.13 (-0.31, 0.04) |  | 0.134 |
| **Adjusted models** |  |  |  |  |  |  |  |  |  |  |  |  |
| N participants | 21,773 |  |  | 21,770 |  |  | 21,798 |  |  | 21,844 |  |  |
| N observations | 47,034 |  |  | 47,022 |  |  | 47,087 |  |  | 47,288 |  |  |
|  | ***B* (95% CI)** | **χ2** | ***p*** | ***B* (95% CI)** | **χ2** | ***p*** | ***B* (95% CI)** | **χ2** | ***p*** | ***B* (95% CI)** | **χ2** | ***p*** |
| Time (linear) | 0.07 (-0.01, 0.14) |  | 0.101 | -0.03 (-0.11, 0.04) |  | 0.424 | -0.21 (-0.30, -0.13) |  | <0.001 | 0.03 (-0.09, 0.15) |  | 0.642 |
| Time (quadratic) | -0.03 (-0.06, 0.01) |  | 0.156 | 0.05 (0.01, 0.08) |  | 0.008 | 0.14 (0.10, 0.18) |  | <0.001 | -0.12 (-0.18, -0.06) |  | <0.001 |
| Cohort (ref. NCDS) |  | 164.6 | <0.001 |  | 279.8 | <0.001 |  | 111.2 | <0.001 |  | 79.7 | <0.001 |
| NSHD | -0.17 (-0.26, -0.08) |  | <0.001 | -0.15 (-0.23, -0.07) |  | <0.001 | -0.06 (-0.18, 0.06) |  | 0.318 | -0.01 (-0.19, 0.18) |  | 0.944 |
| BCS | 0.14 (0.06, 0.22) |  | <0.001 | 0.22 (0.14, 0.30) |  | <0.001 | 0.08 (-0.01, 0.18) |  | 0.078 | -0.30 (-0.42, -0.18) |  | <0.001 |
| NS | 0.44 (0.32, 0.56) |  | <0.001 | 0.61 (0.49, 0.72) |  | <0.001 | 0.44 (0.31, 0.57) |  | <0.001 | -0.49 (-0.64, -0.33) |  | <0.001 |
| MCS | 0.63 (0.50, 0.76) |  | <0.001 | 0.81 (0.68, 0.94) |  | <0.001 | 0.66 (0.51, 0.81) |  | <0.001 | -0.67 (-0.86, -0.48) |  | <0.001 |
| Linear change * cohort (ref. NCDS) |  | 15.9 | 0.003 |  | 2.2 | 0.691 |  | 22.7 | <0.001 |  | 1.6 | 0.816 |
| NSHD | 0.13 (-0.05, 0.31) |  | 0.165 | 0.07 (-0.06, 0.20) |  | 0.306 | -0.15 (-0.35, 0.06) |  | 0.152 | -0.02 (-0.34, 0.31) |  | 0.920 |
| BCS | 0.14 (-0.01, 0.29) |  | 0.062 | 0.03 (-0.12, 0.17) |  | 0.734 | 0.17 (0.02, 0.33) |  | 0.032 | -0.02 (-0.23, 0.18) |  | 0.832 |
| NS | 0.27 (0.04, 0.49) |  | 0.020 | 0.04 (-0.18, 0.26) |  | 0.715 | 0.28 (0.05, 0.51) |  | 0.015 | -0.07 (-0.34, 0.21) |  | 0.639 |
| MCS | 0.40 (0.16, 0.63) |  | 0.001 | -0.11 (-0.35, 0.14) |  | 0.389 | 0.45 (0.18, 0.72) |  | 0.001 | 0.17 (-0.15, 0.49) |  | 0.310 |
| Quadratic change * cohort (ref. NCDS) |  | 8.9 | 0.065 |  | 6.4 | 0.172 |  | 24.8 | <0.001 |  | 3.4 | 0.488 |
| NSHD | -0.04 (-0.13, 0.05) |  | 0.416 | -0.05 (-0.11, 0.01) |  | 0.097 | 0.06 (-0.04, 0.17) |  | 0.231 | -0.01 (-0.17, 0.14) |  | 0.861 |
| BCS | -0.07 (-0.14, -0.01) |  | 0.035 | -0.02 (-0.09, 0.05) |  | 0.527 | -0.07 (-0.15, 0.00) |  | 0.046 | 0.04 (-0.06, 0.13) |  | 0.428 |
| NS | -0.10 (-0.20, 0.01) |  | 0.064 | -0.04 (-0.14, 0.06) |  | 0.417 | -0.14 (-0.25, -0.04) |  | 0.006 | 0.08 (-0.05, 0.20) |  | 0.240 |
| MCS | -0.11 (-0.22, 0.00) |  | 0.050 | 0.08 (-0.03, 0.20) |  | 0.145 | -0.23 (-0.35, -0.11) |  | <0.001 | -0.07 (-0.21, 0.08) |  | 0.349 |
| Birth sex (ref. Men) | 0.36 (0.29, 0.42) |  | <0.001 | 0.18 (0.11, 0.24) |  | <0.001 | 0.27 (0.19, 0.35) |  | <0.001 | -0.11 (-0.22, -0.01) |  | 0.037 |
| Linear change * birth sex (ref. Men) | 0.04 (-0.08, 0.16) |  | 0.498 | -0.07 (-0.18, 0.05) |  | 0.256 | -0.14 (-0.27, 0.00) |  | 0.042 | 0.18 (0.00, 0.36) |  | 0.045 |
| Quadratic change * birth sex (ref. Men) | -0.02 (-0.07, 0.04) |  | 0.573 | 0.03 (-0.02, 0.09) |  | 0.236 | 0.08 (0.02, 0.14) |  | 0.007 | -0.13 (-0.21, -0.04) |  | 0.003 |
| Cohort (ref. NCDS) * birth sex (ref. Men) |  | 38.4 | <0.001 |  | 29 | <0.001 |  | 7.9 | 0.095 |  | 6.3 | 0.178 |
| NSHD | -0.04 (-0.18, 0.10) |  | 0.614 | -0.05 (-0.19, 0.08) |  | 0.458 | 0.01 (-0.18, 0.20) |  | 0.930 | -0.10 (-0.36, 0.17) |  | 0.470 |
| BCS | 0.01 (-0.10, 0.12) |  | 0.837 | -0.06 (-0.16, 0.05) |  | 0.282 | -0.05 (-0.18, 0.07) |  | 0.389 | 0.14 (-0.02, 0.30) |  | 0.097 |
| NS | 0.23 (0.06, 0.39) |  | 0.007 | 0.01 (-0.15, 0.17) |  | 0.896 | -0.17 (-0.33, 0.00) |  | 0.048 | 0.08 (-0.13, 0.29) |  | 0.476 |
| MCS | 0.44 (0.28, 0.59) |  | <0.001 | 0.36 (0.21, 0.52) |  | <0.001 | -0.19 (-0.36, -0.02) |  | 0.025 | -0.10 (-0.32, 0.12) |  | 0.375 |
| Linear change * cohort (ref. NCDS) *  birth sex (ref. Men) |  | 3.5 | 0.483 |  | 12.8 | 0.012 |  | 3.2 | 0.5222 |  | 10 | 0.041 |
| NSHD | 0.12 (-0.16, 0.39) |  | 0.404 | 0.09 (-0.16, 0.34) |  | 0.494 | 0.13 (-0.19, 0.45) |  | 0.431 | -0.09 (-0.58, 0.40) |  | 0.727 |
| BCS | -0.05 (-0.26, 0.16) |  | 0.644 | -0.11 (-0.31, 0.09) |  | 0.282 | 0.09 (-0.12, 0.31) |  | 0.392 | -0.13 (-0.41, 0.16) |  | 0.380 |
| NS | -0.22 (-0.52, 0.08) |  | 0.150 | -0.38 (-0.67, -0.10) |  | 0.008 | -0.12 (-0.41, 0.17) |  | 0.419 | 0.43 (0.06, 0.80) |  | 0.022 |
| MCS | -0.02 (-0.34, 0.29) |  | 0.887 | -0.36 (-0.67, -0.04) |  | 0.026 | -0.12 (-0.45, 0.21) |  | 0.485 | 0.28 (-0.13, 0.69) |  | 0.177 |
| Quadratic change * cohort (ref. NCDS) *  birth sex (ref. Men) |  | 3.3 | 0.514 |  | 12 | 0.017 |  | 5.5 | 0.3 |  | 8.4 | 0.077 |
| NSHD | -0.06 (-0.20, 0.07) |  | 0.354 | -0.02 (-0.14, 0.09) |  | 0.694 | -0.06 (-0.22, 0.09) |  | 0.421 | 0.03 (-0.21, 0.28) |  | 0.789 |
| BCS | 0.03 (-0.07, 0.12) |  | 0.562 | 0.06 (-0.03, 0.15) |  | 0.218 | -0.07 (-0.17, 0.03) |  | 0.170 | 0.06 (-0.07, 0.19) |  | 0.352 |
| NS | 0.09 (-0.05, 0.22) |  | 0.207 | 0.19 (0.06, 0.32) |  | 0.003 | 0.07 (-0.06, 0.20) |  | 0.295 | -0.19 (-0.35, -0.02) |  | 0.027 |
| MCS | 0.00 (-0.15, 0.14) |  | 0.972 | 0.13 (-0.02, 0.28) |  | 0.080 | 0.06 (-0.10, 0.21) |  | 0.479 | -0.07 (-0.26, 0.12) |  | 0.463 |
| **Sensitivity models** |  |  |  |  |  |  |  |  |  |  |  |  |
| N participants | 21,773 |  |  | 21,770 |  |  | 21,798 |  |  | 21,844 |  |  |
| N observations | 47,034 |  |  | 47,022 |  |  | 47,087 |  |  | 47,288 |  |  |
|  | ***B* (95% CI)** | **χ2** | ***p*** | ***B* (95% CI)** | **χ2** | ***p*** | ***B* (95% CI)** | **χ2** | ***p*** | ***B* (95% CI)** | **χ2** | ***p*** |
| Time (linear) | 0.07 (-0.01, 0.15) |  | 0.075 | -0.02 (-0.10, 0.05) |  | 0.558 | -0.20 (-0.29, -0.11) |  | <0.001 | 0.01 (-0.11, 0.13) |  | 0.834 |
| Time (quadratic) | -0.03 (-0.06, 0.01) |  | 0.136 | 0.05 (0.01, 0.08) |  | 0.011 | 0.14 (0.09, 0.18) |  | <0.001 | -0.12 (-0.17, -0.06) |  | <0.001 |
| Cohort (ref. NCDS) |  | 258.1 | <0.001 |  | 401.9 | <0.001 |  | 299 | <0.001 |  | 208.4 | <0.001 |
| NSHD | -0.21 (-0.29, -0.12) |  | <0.001 | -0.19 (-0.27, -0.11) |  | <0.001 | -0.17 (-0.29, -0.05) |  | 0.006 | 0.14 (-0.05, 0.33) |  | 0.162 |
| BCS | 0.19 (0.11, 0.27) |  | <0.001 | 0.26 (0.18, 0.34) |  | <0.001 | 0.10 (0.00, 0.19) |  | 0.048 | -0.34 (-0.46, -0.21) |  | <0.001 |
| NS | 0.56 (0.43, 0.69) |  | <0.001 | 0.71 (0.59, 0.84) |  | <0.001 | 0.60 (0.46, 0.74) |  | <0.001 | -0.68 (-0.85, -0.51) |  | <0.001 |
| MCS | 0.69 (0.57, 0.82) |  | <0.001 | 0.90 (0.78, 1.02) |  | <0.001 | 1.06 (0.92, 1.20) |  | <0.001 | -1.13 (-1.31, -0.95) |  | <0.001 |
| Linear change * cohort (ref. NCDS) |  | 15.7 | 0.004 |  | 1.7 | 0.788 |  | 22.3 | <0.001 |  | 1.3 | 0.857 |
| NSHD | 0.11 (-0.07, 0.29) |  | 0.232 | 0.06 (-0.07, 0.19) |  | 0.400 | -0.16 (-0.37, 0.04) |  | 0.116 | 0.00 (-0.32, 0.33) |  | 0.984 |
| BCS | 0.12 (-0.03, 0.27) |  | 0.112 | 0.00 (-0.15, 0.14) |  | 0.974 | 0.14 (-0.02, 0.30) |  | 0.080 | 0.03 (-0.18, 0.23) |  | 0.804 |
| NS | 0.27 (0.04, 0.50) |  | 0.019 | 0.05 (-0.17, 0.27) |  | 0.657 | 0.30 (0.07, 0.53) |  | 0.011 | -0.06 (-0.34, 0.22) |  | 0.668 |
| MCS | 0.41 (0.17, 0.65) |  | 0.001 | -0.10 (-0.35, 0.15) |  | 0.451 | 0.45 (0.17, 0.72) |  | 0.001 | 0.16 (-0.16, 0.49) |  | 0.332 |
| Quadratic change * cohort (ref. NCDS) |  | 8.9 | 0.064 |  | 5.6 | 0.235 |  | 24.9 | <0.001 |  | 2.7 | 0.614 |
| NSHD | -0.03 (-0.12, 0.06) |  | 0.494 | -0.05 (-0.11, 0.01) |  | 0.110 | 0.07 (-0.04, 0.17) |  | 0.205 | -0.02 (-0.17, 0.14) |  | 0.823 |
| BCS | -0.06 (-0.13, 0.00) |  | 0.061 | -0.01 (-0.08, 0.06) |  | 0.752 | -0.06 (-0.14, 0.01) |  | 0.094 | 0.02 (-0.08, 0.12) |  | 0.689 |
| NS | -0.10 (-0.20, 0.00) |  | 0.054 | -0.05 (-0.15, 0.05) |  | 0.352 | -0.15 (-0.26, -0.05) |  | 0.004 | 0.08 (-0.05, 0.20) |  | 0.240 |
| MCS | -0.12 (-0.23, -0.01) |  | 0.035 | 0.07 (-0.04, 0.19) |  | 0.207 | -0.23 (-0.36, -0.11) |  | <0.001 | -0.06 (-0.21, 0.09) |  | 0.420 |
| Birth sex (ref. Men) | 0.47 (0.40, 0.54) |  | <0.001 | 0.29 (0.22, 0.36) |  | <0.001 | 0.39 (0.31, 0.48) |  | <0.001 | -0.24 (-0.35, -0.13) |  | <0.001 |
| Linear change * birth sex (ref. Men) | 0.04 (-0.08, 0.16) |  | 0.524 | -0.07 (-0.18, 0.04) |  | 0.233 | -0.14 (-0.27, -0.01) |  | 0.037 | 0.19 (0.01, 0.37) |  | 0.039 |
| Quadratic change * birth sex (ref. Men) | -0.02 (-0.07, 0.04) |  | 0.588 | 0.03 (-0.02, 0.09) |  | 0.222 | 0.09 (0.02, 0.15) |  | 0.006 | -0.13 (-0.21, -0.05) |  | 0.002 |
| Cohort (ref. NCDS) * birth sex (ref. Men) |  | 50.1 | <0.001 |  | 41.7 | <0.001 |  | 10.8 | 0 |  | 11.7 | 0.02 |
| NSHD | -0.04 (-0.18, 0.11) |  | 0.599 | -0.02 (-0.16, 0.12) |  | 0.777 | 0.08 (-0.12, 0.28) |  | 0.406 | -0.15 (-0.42, 0.13) |  | 0.305 |
| BCS | -0.03 (-0.15, 0.08) |  | 0.583 | -0.11 (-0.22, 0.01) |  | 0.066 | -0.11 (-0.24, 0.03) |  | 0.114 | 0.17 (0.00, 0.35) |  | 0.044 |
| NS | 0.22 (0.05, 0.38) |  | 0.013 | 0.00 (-0.17, 0.16) |  | 0.960 | -0.22 (-0.40, -0.05) |  | 0.013 | 0.10 (-0.12, 0.33) |  | 0.357 |
| MCS | 0.52 (0.36, 0.68) |  | <0.001 | 0.45 (0.29, 0.61) |  | <0.001 | -0.15 (-0.33, 0.02) |  | 0.087 | -0.17 (-0.40, 0.05) |  | 0.131 |
| Linear change * cohort (ref. NCDS) *  birth sex (ref. Men) |  | 4 | 0.413 |  | 13.7 | 0.008 |  | 4.6 | 0.336 |  | 11.4 | 0.022 |
| NSHD | 0.13 (-0.14, 0.40) |  | 0.355 | 0.10 (-0.15, 0.35) |  | 0.443 | 0.14 (-0.18, 0.47) |  | 0.383 | -0.10 (-0.59, 0.40) |  | 0.695 |
| BCS | -0.02 (-0.23, 0.19) |  | 0.847 | -0.07 (-0.27, 0.13) |  | 0.484 | 0.13 (-0.08, 0.35) |  | 0.224 | -0.18 (-0.47, 0.10) |  | 0.204 |
| NS | -0.24 (-0.54, 0.06) |  | 0.120 | -0.40 (-0.69, -0.11) |  | 0.006 | -0.14 (-0.43, 0.16) |  | 0.356 | 0.43 (0.05, 0.80) |  | 0.025 |
| MCS | -0.04 (-0.36, 0.28) |  | 0.813 | -0.38 (-0.70, -0.06) |  | 0.021 | -0.11 (-0.45, 0.22) |  | 0.507 | 0.29 (-0.13, 0.70) |  | 0.176 |
| Quadratic change * cohort (ref. NCDS) *  birth sex (ref. Men) |  | 3.7 | 0.446 |  | 12.9 | 0.012 |  | 7.3 | 0.123 |  | 9.9 | 0.042 |
| NSHD | -0.07 (-0.20, 0.06) |  | 0.310 | -0.03 (-0.15, 0.09) |  | 0.648 | -0.07 (-0.23, 0.09) |  | 0.376 | 0.04 (-0.21, 0.29) |  | 0.758 |
| BCS | 0.02 (-0.08, 0.11) |  | 0.707 | 0.04 (-0.05, 0.14) |  | 0.353 | -0.09 (-0.19, 0.01) |  | 0.092 | 0.09 (-0.04, 0.22) |  | 0.196 |
| NS | 0.10 (-0.04, 0.23) |  | 0.160 | 0.20 (0.07, 0.33) |  | 0.002 | 0.08 (-0.05, 0.21) |  | 0.232 | -0.19 (-0.36, -0.02) |  | 0.028 |
| MCS | 0.00 (-0.14, 0.15) |  | 0.956 | 0.14 (-0.01, 0.29) |  | 0.066 | 0.05 (-0.11, 0.20) |  | 0.530 | -0.07 (-0.26, 0.12) |  | 0.481 |

*Note.* Adjusted models included highest qualification achieved, pre-pandemic self-reported health, pre-pandemic psychological distress, and household composition as covariates. Sensitivity models correspond to the unadjusted models after restricting the analytical sample to that of the adjusted models. BCS: British Cohort Study, 1970 birth cohort; GAD-2: 2-item General Anxiety Disorder questionnaire; MCS: Millennium Cohort Study, 2000 birth cohort; NCDS: National Child and Development Study, 1958 birth cohort; NS: Next Steps, 1990 cohort; NSHD: National Survey of Health and Development, 1946 birth cohort; ONS: UK Office for National Statistics; PHQ-2: 2-item Patient Health Questionnaire; UCLA-3: 3-item UCLA loneliness scale. χ2: Wald test performed to assess the overall statistical significance of the interaction terms; all χ2 statistics in this table have 4 degrees of freedom.

#### Table S6.2. Unadjusted and adjusted marginal mean estimates and 95% confidence intervals by birth sex.

|  |  |  | **Anxiety symptomatology (GAD-2)** | **Depressive symptomatology (PHQ-2)** | **Feelings of loneliness (UCLA-3)** | **Life satisfaction (ONS single question)** |
| --- | --- | --- | --- | --- | --- | --- |
| **Cohort** | **Birth sex** | **Survey wave** | **Unadjusted marginal mean (95% CI)** | **Unadjusted marginal mean (95% CI)** | **Unadjusted marginal mean (95% CI)** | **Unadjusted marginal mean (95% CI)** |
| NSHD | Men | 1 | 0.28 (0.21, 0.34) | 0.31 (0.25, 0.37) | 3.75 (3.65, 3.85) | 7.75 (7.58, 7.92) |
| NSHD | Men | 2 | 0.40 (0.33, 0.47) | 0.33 (0.27, 0.39) | 3.61 (3.53, 3.70) | 7.66 (7.52, 7.80) |
| NSHD | Men | 3 | 0.37 (0.30, 0.44) | 0.38 (0.32, 0.45) | 3.87 (3.77, 3.97) | 7.22 (7.05, 7.38) |
| NSHD | Women | 1 | 0.69 (0.60, 0.78) | 0.59 (0.50, 0.68) | 4.19 (4.07, 4.32) | 7.39 (7.22, 7.56) |
| NSHD | Women | 2 | 0.85 (0.76, 0.95) | 0.65 (0.56, 0.73) | 4.05 (3.94, 4.16) | 7.25 (7.11, 7.39) |
| NSHD | Women | 3 | 0.81 (0.71, 0.90) | 0.70 (0.62, 0.79) | 4.39 (4.27, 4.51) | 6.74 (6.59, 6.89) |
| NCDS | Men | 1 | 0.50 (0.46, 0.55) | 0.52 (0.48, 0.57) | 3.94 (3.89, 4.00) | 7.58 (7.50, 7.65) |
| NCDS | Men | 2 | 0.53 (0.49, 0.57) | 0.55 (0.51, 0.59) | 3.86 (3.81, 3.91) | 7.52 (7.45, 7.59) |
| NCDS | Men | 3 | 0.52 (0.48, 0.56) | 0.65 (0.61, 0.69) | 4.07 (4.02, 4.12) | 7.16 (7.09, 7.22) |
| NCDS | Women | 1 | 0.97 (0.91, 1.02) | 0.80 (0.75, 0.85) | 4.33 (4.27, 4.39) | 7.36 (7.28, 7.44) |
| NCDS | Women | 2 | 1.03 (0.98, 1.08) | 0.80 (0.76, 0.85) | 4.21 (4.16, 4.27) | 7.30 (7.23, 7.36) |
| NCDS | Women | 3 | 1.01 (0.96, 1.06) | 0.94 (0.89, 0.98) | 4.53 (4.47, 4.58) | 6.78 (6.71, 6.85) |
| BCS | Men | 1 | 0.71 (0.65, 0.77) | 0.79 (0.73, 0.85) | 4.03 (3.96, 4.10) | 7.27 (7.18, 7.35) |
| BCS | Men | 2 | 0.79 (0.74, 0.85) | 0.81 (0.76, 0.87) | 4.09 (4.02, 4.15) | 7.16 (7.08, 7.24) |
| BCS | Men | 3 | 0.76 (0.71, 0.81) | 0.92 (0.86, 0.97) | 4.26 (4.20, 4.33) | 6.90 (6.82, 6.98) |
| BCS | Women | 1 | 1.12 (1.06, 1.17) | 0.98 (0.92, 1.03) | 4.37 (4.31, 4.43) | 7.17 (7.09, 7.24) |
| BCS | Women | 2 | 1.25 (1.19, 1.30) | 0.93 (0.88, 0.98) | 4.37 (4.31, 4.43) | 7.07 (7.00, 7.14) |
| BCS | Women | 3 | 1.21 (1.15, 1.26) | 1.11 (1.06, 1.17) | 4.54 (4.48, 4.60) | 6.68 (6.61, 6.75) |
| NS | Men | 1 | 1.05 (0.94, 1.17) | 1.24 (1.13, 1.36) | 4.54 (4.42, 4.66) | 6.90 (6.75, 7.04) |
| NS | Men | 2 | 1.25 (1.16, 1.34) | 1.26 (1.17, 1.35) | 4.61 (4.51, 4.70) | 6.83 (6.71, 6.94) |
| NS | Men | 3 | 1.24 (1.16, 1.32) | 1.31 (1.23, 1.39) | 4.66 (4.58, 4.75) | 6.64 (6.54, 6.75) |
| NS | Women | 1 | 1.74 (1.65, 1.83) | 1.53 (1.45, 1.62) | 4.71 (4.62, 4.79) | 6.77 (6.66, 6.89) |
| NS | Women | 2 | 1.83 (1.76, 1.91) | 1.32 (1.25, 1.39) | 4.68 (4.62, 4.75) | 6.97 (6.88, 7.05) |
| NS | Women | 3 | 1.86 (1.79, 1.93) | 1.60 (1.54, 1.67) | 4.94 (4.88, 5.01) | 6.46 (6.38, 6.55) |
| MCS | Men | 1 | 1.15 (1.04, 1.26) | 1.42 (1.31, 1.52) | 5.00 (4.88, 5.12) | 6.46 (6.31, 6.62) |
| MCS | Men | 2 | 1.47 (1.37, 1.57) | 1.40 (1.30, 1.49) | 5.07 (4.97, 5.18) | 6.47 (6.35, 6.60) |
| MCS | Men | 3 | 1.52 (1.44, 1.60) | 1.63 (1.55, 1.71) | 5.07 (4.99, 5.16) | 6.13 (6.02, 6.24) |
| MCS | Women | 1 | 2.17 (2.08, 2.26) | 2.19 (2.10, 2.27) | 5.26 (5.18, 5.35) | 5.99 (5.88, 6.10) |
| MCS | Women | 2 | 2.48 (2.39, 2.56) | 1.89 (1.81, 1.97) | 5.27 (5.19, 5.35) | 6.29 (6.19, 6.38) |
| MCS | Women | 3 | 2.51 (2.44, 2.59) | 2.24 (2.17, 2.31) | 5.39 (5.31, 5.46) | 5.81 (5.72, 5.90) |
| **Cohort** | **Birth sex** | **Survey wave** | **Adjusted marginal mean (95% CI)** | **Adjusted marginal mean (95% CI)** | **Adjusted marginal mean (95% CI)** | **Adjusted marginal mean (95% CI)** |
| NSHD | Men | 1 | 0.43 (0.36, 0.51) | 0.51 (0.43, 0.58) | 4.06 (3.95, 4.17) | 7.38 (7.21, 7.55) |
| NSHD | Men | 2 | 0.56 (0.48, 0.65) | 0.54 (0.47, 0.61) | 3.90 (3.79, 4.00) | 7.26 (7.10, 7.43) |
| NSHD | Men | 3 | 0.57 (0.47, 0.66) | 0.57 (0.48, 0.65) | 4.14 (4.03, 4.26) | 6.87 (6.69, 7.06) |
| NSHD | Women | 1 | 0.75 (0.65, 0.85) | 0.63 (0.53, 0.73) | 4.34 (4.19, 4.48) | 7.17 (6.99, 7.35) |
| NSHD | Women | 2 | 0.96 (0.85, 1.07) | 0.69 (0.60, 0.79) | 4.19 (4.06, 4.32) | 7.05 (6.87, 7.23) |
| NSHD | Women | 3 | 0.88 (0.77, 1.00) | 0.77 (0.67, 0.86) | 4.49 (4.35, 4.63) | 6.48 (6.27, 6.68) |
| NCDS | Men | 1 | 0.61 (0.56, 0.65) | 0.66 (0.61, 0.70) | 4.12 (4.06, 4.18) | 7.39 (7.31, 7.46) |
| NCDS | Men | 2 | 0.64 (0.60, 0.69) | 0.67 (0.63, 0.72) | 4.05 (3.99, 4.10) | 7.30 (7.23, 7.37) |
| NCDS | Men | 3 | 0.63 (0.59, 0.67) | 0.79 (0.74, 0.83) | 4.25 (4.19, 4.31) | 6.97 (6.90, 7.04) |
| NCDS | Women | 1 | 0.96 (0.91, 1.02) | 0.83 (0.78, 0.88) | 4.39 (4.33, 4.45) | 7.28 (7.20, 7.36) |
| NCDS | Women | 2 | 1.03 (0.97, 1.08) | 0.82 (0.77, 0.86) | 4.26 (4.21, 4.32) | 7.24 (7.17, 7.31) |
| NCDS | Women | 3 | 1.01 (0.96, 1.06) | 0.96 (0.91, 1.01) | 4.58 (4.52, 4.64) | 6.71 (6.63, 6.78) |
| BCS | Men | 1 | 0.75 (0.68, 0.82) | 0.87 (0.81, 0.94) | 4.20 (4.13, 4.28) | 7.09 (6.99, 7.18) |
| BCS | Men | 2 | 0.86 (0.80, 0.92) | 0.90 (0.83, 0.96) | 4.23 (4.16, 4.30) | 7.02 (6.93, 7.10) |
| BCS | Men | 3 | 0.77 (0.71, 0.82) | 0.97 (0.91, 1.03) | 4.38 (4.31, 4.45) | 6.78 (6.69, 6.87) |
| BCS | Women | 1 | 1.11 (1.05, 1.17) | 0.99 (0.93, 1.05) | 4.42 (4.35, 4.48) | 7.11 (7.03, 7.20) |
| BCS | Women | 2 | 1.23 (1.17, 1.29) | 0.93 (0.87, 0.98) | 4.41 (4.35, 4.48) | 7.03 (6.95, 7.11) |
| BCS | Women | 3 | 1.17 (1.11, 1.23) | 1.10 (1.04, 1.15) | 4.57 (4.50, 4.63) | 6.65 (6.57, 6.73) |
| NS | Men | 1 | 1.04 (0.93, 1.16) | 1.26 (1.15, 1.37) | 4.56 (4.44, 4.67) | 6.90 (6.76, 7.04) |
| NS | Men | 2 | 1.26 (1.16, 1.35) | 1.28 (1.19, 1.37) | 4.62 (4.52, 4.72) | 6.82 (6.70, 6.94) |
| NS | Men | 3 | 1.22 (1.14, 1.31) | 1.31 (1.23, 1.39) | 4.68 (4.59, 4.76) | 6.65 (6.54, 6.76) |
| NS | Women | 1 | 1.63 (1.53, 1.72) | 1.45 (1.36, 1.54) | 4.66 (4.57, 4.74) | 6.87 (6.75, 6.99) |
| NS | Women | 2 | 1.73 (1.65, 1.81) | 1.24 (1.17, 1.31) | 4.62 (4.55, 4.69) | 7.08 (6.99, 7.17) |
| NS | Women | 3 | 1.73 (1.65, 1.80) | 1.49 (1.42, 1.56) | 4.88 (4.81, 4.95) | 6.58 (6.49, 6.67) |
| MCS | Men | 1 | 1.24 (1.12, 1.36) | 1.47 (1.35, 1.58) | 4.78 (4.65, 4.91) | 6.72 (6.55, 6.88) |
| MCS | Men | 2 | 1.56 (1.45, 1.67) | 1.46 (1.35, 1.57) | 4.92 (4.80, 5.05) | 6.72 (6.58, 6.86) |
| MCS | Men | 3 | 1.62 (1.52, 1.71) | 1.72 (1.63, 1.81) | 4.88 (4.78, 4.98) | 6.35 (6.23, 6.48) |
| MCS | Women | 1 | 2.03 (1.93, 2.13) | 2.01 (1.91, 2.10) | 4.85 (4.76, 4.95) | 6.50 (6.38, 6.63) |
| MCS | Women | 2 | 2.36 (2.26, 2.45) | 1.74 (1.65, 1.83) | 4.88 (4.79, 4.98) | 6.78 (6.66, 6.89) |
| MCS | Women | 3 | 2.37 (2.28, 2.46) | 2.06 (1.98, 2.15) | 5.01 (4.92, 5.09) | 6.28 (6.17, 6.38) |

*Note.* Adjusted models included highest qualification achieved, pre-pandemic self-reported health, pre-pandemic psychological distress, and household composition as covariates. BCS: British Cohort Study, 1970 birth cohort; GAD-2: 2-item General Anxiety Disorder questionnaire; MCS: Millennium Cohort Study, 2000 birth cohort; NCDS: National Child and Development Study, 1958 birth cohort; NS: Next Steps, 1990 cohort; NSHD: National Survey of Health and Development, 1946 birth cohort; ONS: UK Office for National Statistics; PHQ-2: 2-item Patient Health Questionnaire; UCLA-3: 3-item UCLA loneliness scale. Survey wave 1: May 2020; survey wave 2: September/October 2020; survey wave 3: February/March 2021.

#### Table S6.3. Difference-in-differences (change in difference between men and women between first vs last time point) estimates and 95% confidence intervals.

|  | *DID* (95% CI) | *p* |
| --- | --- | --- |
| GAD-2 | -0.01 (-0.07, 0.05) | 0.705 |
| PHQ-2 | -0.02 (-0.08, 0.03) | 0.413 |
| UCLA-3 | 0.05 (-0.01, 0.11) | 0.130 |
| Life satisfaction | -0.08 (-0.17, 0.01) | 0.085 |

*Note*. DID: difference-in-differences; GAD-2: 2-item General Anxiety Disorder questionnaire; PHQ-2: 2-item Patient Health Questionnaire; UCLA-3: 3-item UCLA loneliness scale.

#### Figure S6.1. Unadjusted and adjusted (by highest qualification achieved, pre-pandemic self-reported health, pre-pandemic psychological distress, and household composition) anxiety symptomatology (GAD-2) marginal mean estimates and 95% confidence intervals by birth sex.

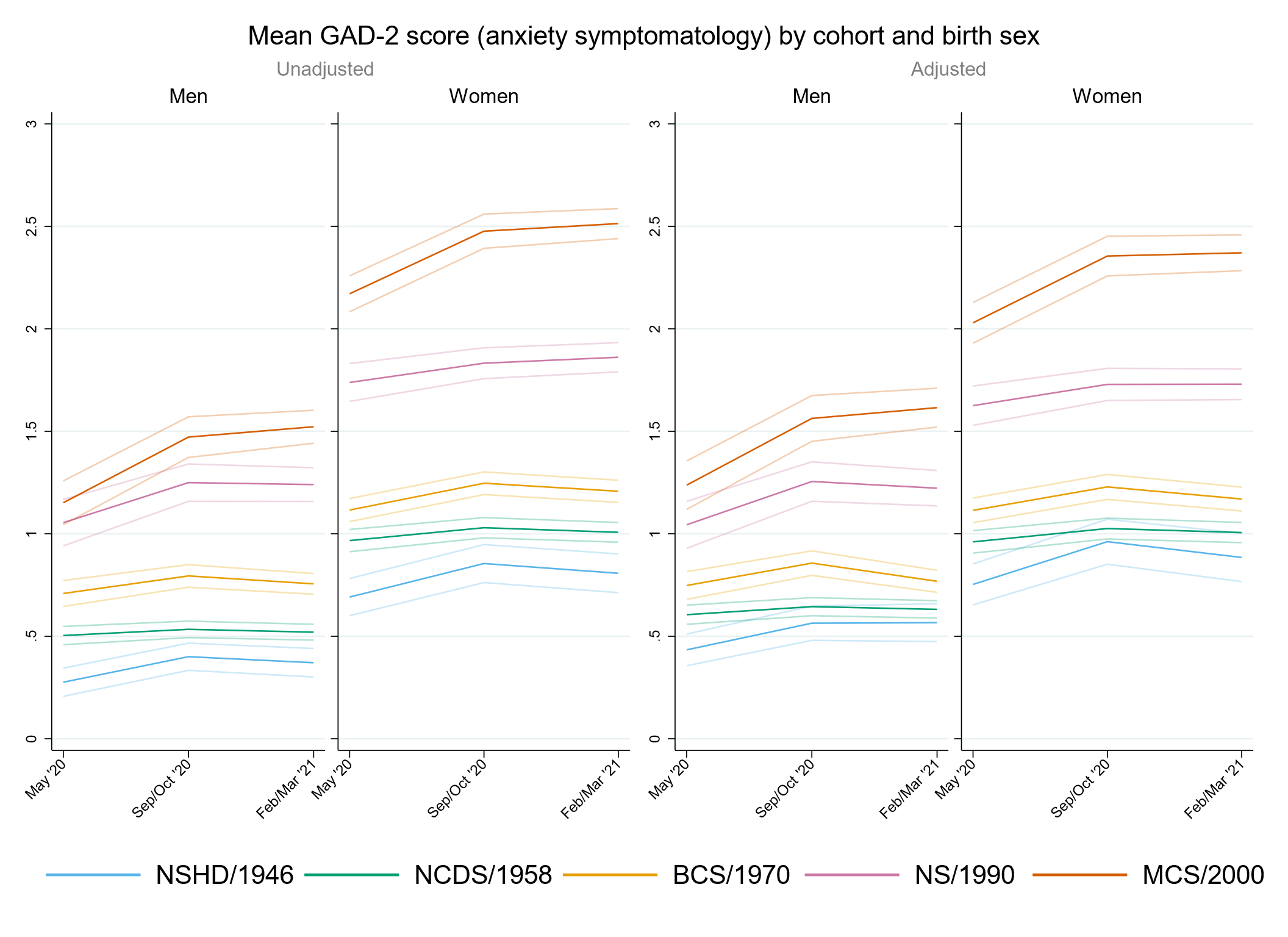

#### Figure S6.2. Unadjusted and adjusted (by highest qualification achieved, pre-pandemic self-reported health, pre-pandemic psychological distress, and household composition) depressive symptomatology (PHQ-2) marginal mean estimates and 95% confidence intervals by birth sex.

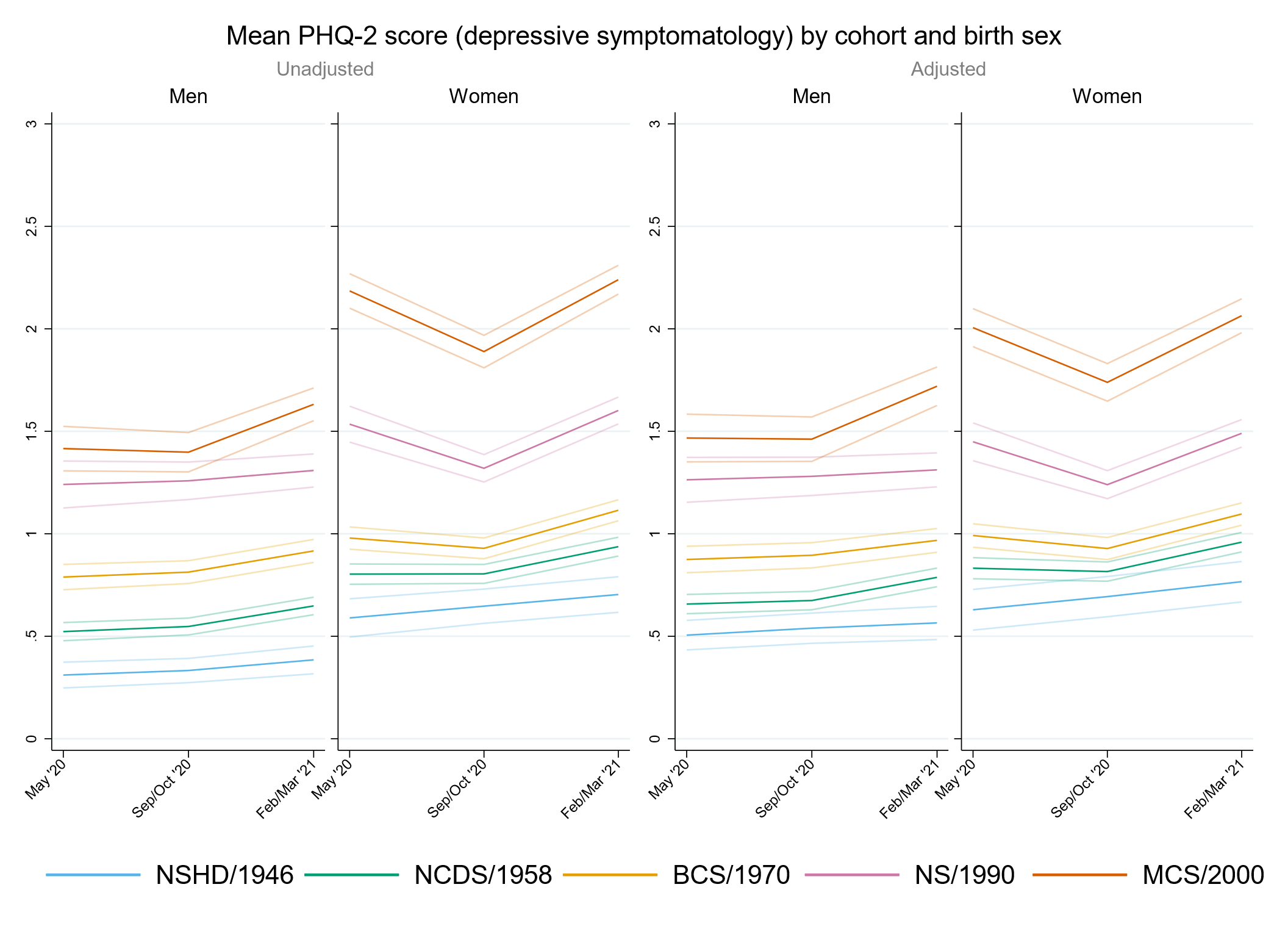

#### Figure S6.3. Unadjusted and adjusted (by highest qualification achieved, pre-pandemic self-reported health, pre-pandemic psychological distress, and household composition) loneliness (UCLA-3) marginal mean estimates and 95% confidence intervals by birth sex.

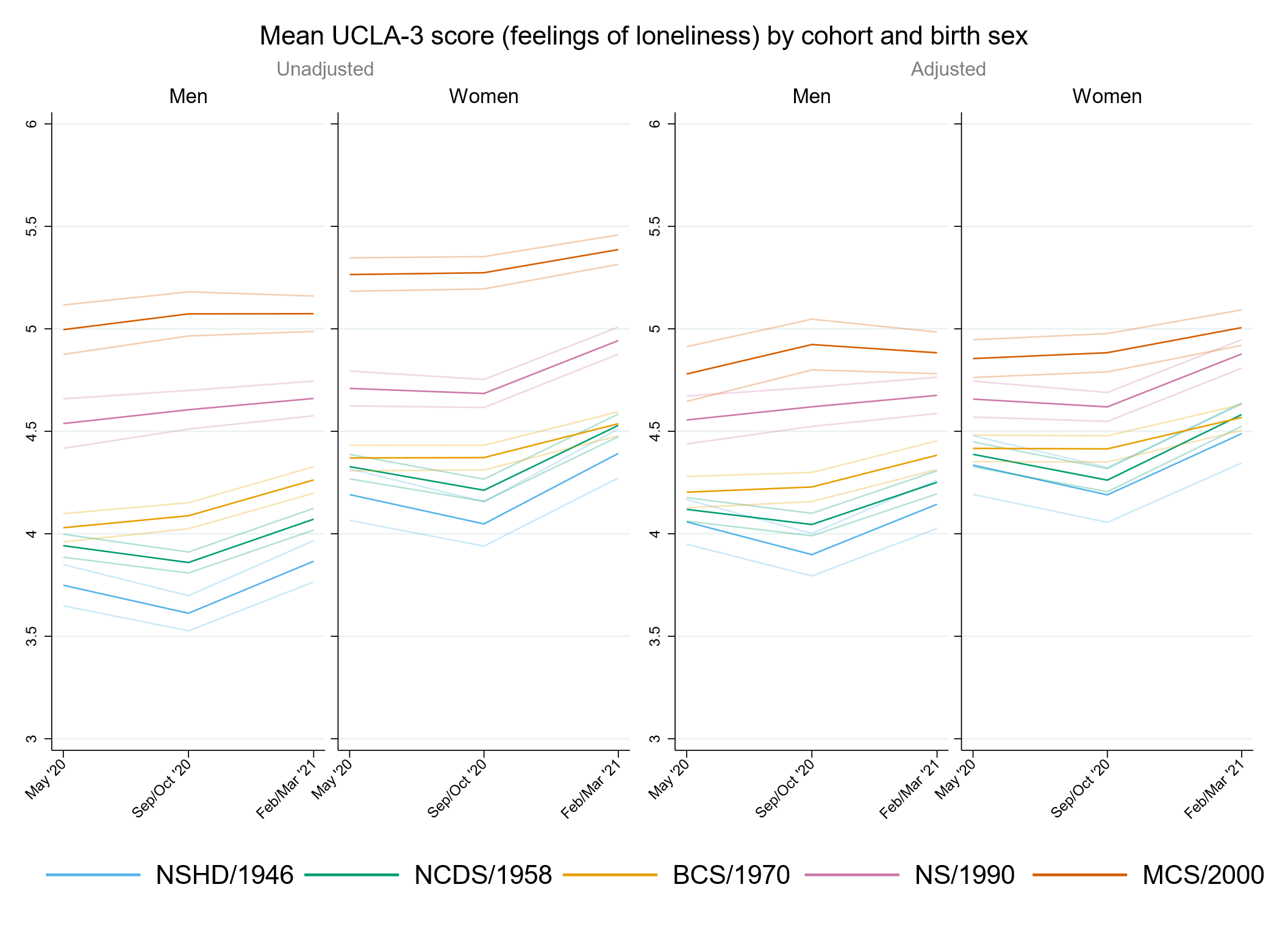

#### Figure S6.4. Unadjusted and adjusted (by highest qualification achieved, pre-pandemic self-reported health, pre-pandemic psychological distress, and household composition) life satisfaction marginal mean estimates and 95% confidence intervals by birth sex.

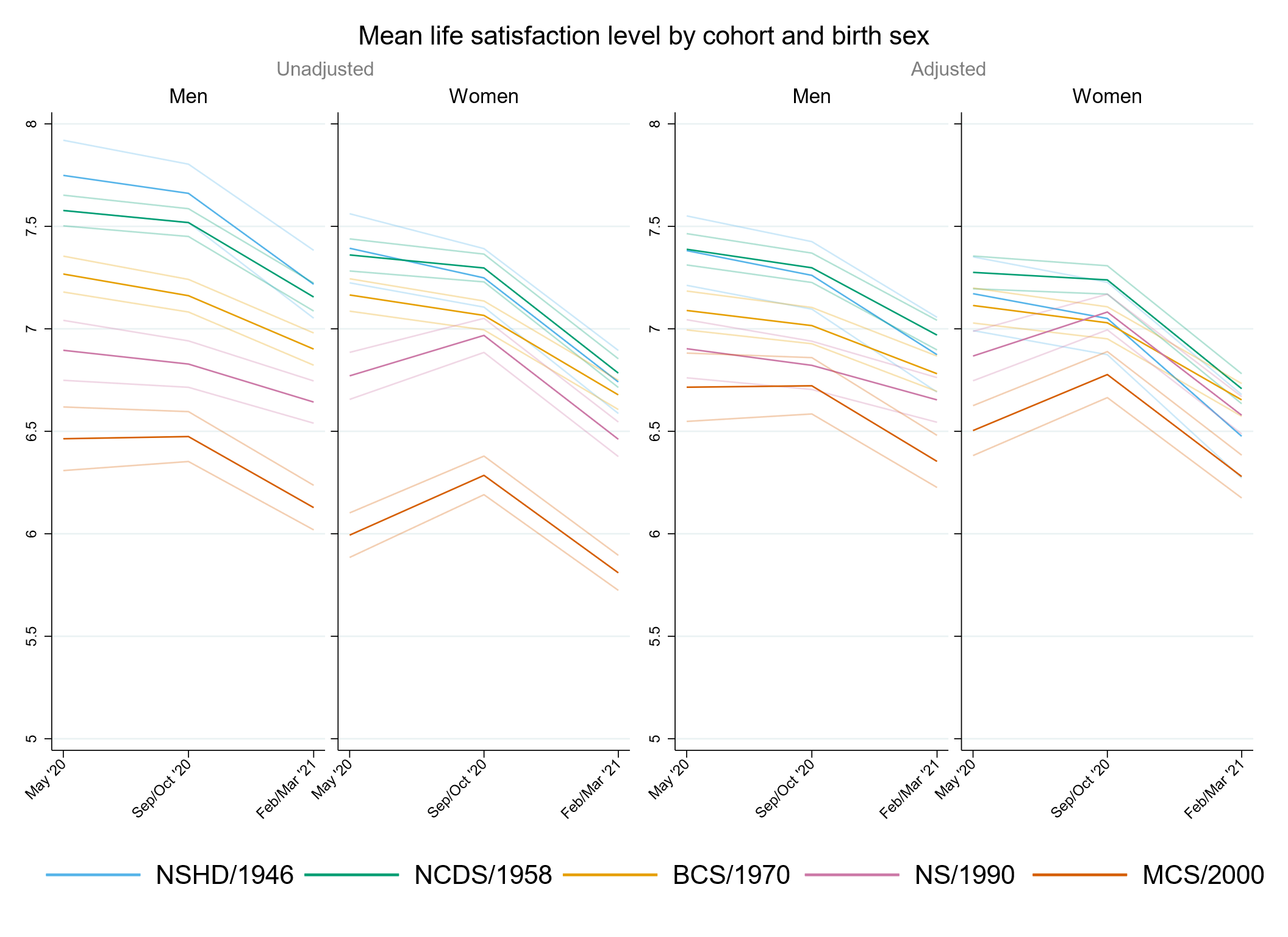

### Appendix S5. Results by pre-pandemic financial situation.

#### Table S7.1. Results of multilevel growth curve models by pre-pandemic financial situation.

|  | **Anxiety symptomatology (GAD-2)** | | | **Depressive symptomatology (PHQ-2)** | | | **Feelings of loneliness (UCLA-3)** | | | **Life satisfaction (ONS single question)** | | |
| --- | --- | --- | --- | --- | --- | --- | --- | --- | --- | --- | --- | --- |
| **Unadjusted models** |  | | |  | | |  | | |  | | |
| N participants | 24,874 |  |  | 24,873 |  |  | 24,913 |  |  | 24,963 |  |  |
| N observations | 52,962 |  |  | 52,947 |  |  | 53,039 |  |  | 53,262 |  |  |
|  | ***B* (95% CI)** | **χ2** | ***p*** | ***B* (95% CI)** | **χ2** | ***p*** | ***B* (95% CI)** | **χ2** | ***p*** | ***B* (95% CI)** | **χ2** | ***p*** |
| Time (linear) | 0.09 (-0.02, 0.20) |  | 0.113 | 0.05 (-0.05, 0.15) |  | 0.310 | -0.27 (-0.39, -0.15) |  | <0.001 | -0.01 (-0.16, 0.14) |  | 0.890 |
| Time (quadratic) | -0.04 (-0.09, 0.02) |  | 0.167 | 0.02 (-0.03, 0.06) |  | 0.445 | 0.18 (0.13, 0.24) |  | <0.001 | -0.11 (-0.18, -0.04) |  | 0.003 |
| Cohort (ref. NCDS) |  | 375.9 | <0.001 |  | 518.6 | <0.001 |  | 319.7 | <0.001 |  | 268 | <0.001 |
| NSHD | -0.20 (-0.34, -0.05) |  | 0.007 | -0.06 (-0.21, 0.09) |  | 0.433 | -0.08 (-0.28, 0.12) |  | 0.431 | 0.03 (-0.24, 0.30) |  | 0.822 |
| BCS | 0.11 (0.01, 0.20) |  | 0.025 | 0.17 (0.09, 0.26) |  | <0.001 | -0.04 (-0.15, 0.06) |  | 0.404 | -0.11 (-0.24, 0.01) |  | 0.077 |
| NS | 0.65 (0.52, 0.78) |  | <0.001 | 0.69 (0.56, 0.81) |  | <0.001 | 0.40 (0.27, 0.54) |  | <0.001 | -0.55 (-0.71, -0.39) |  | <0.001 |
| MCS | 0.98 (0.86, 1.10) |  | <0.001 | 1.20 (1.09, 1.32) |  | <0.001 | 0.96 (0.83, 1.08) |  | <0.001 | -1.20 (-1.36, -1.04) |  | <0.001 |
| Linear change * cohort (ref. NCDS) |  | 6.7 | 0.15 |  | 36.9 | <0.001 |  | 12.3 | 0.015 |  | 31.7 | <0.001 |
| NSHD | 0.20 (-0.04, 0.43) |  | 0.103 | -0.08 (-0.31, 0.15) |  | 0.498 | -0.12 (-0.45, 0.22) |  | 0.495 | -0.05 (-0.49, 0.38) |  | 0.812 |
| BCS | 0.04 (-0.12, 0.19) |  | 0.646 | -0.21 (-0.35, -0.06) |  | 0.005 | 0.25 (0.08, 0.42) |  | 0.003 | -0.01 (-0.22, 0.21) |  | 0.947 |
| NS | 0.09 (-0.14, 0.31) |  | 0.449 | -0.40 (-0.61, -0.20) |  | <0.001 | 0.16 (-0.06, 0.38) |  | 0.144 | 0.44 (0.17, 0.70) |  | 0.001 |
| MCS | 0.26 (0.03, 0.48) |  | 0.026 | -0.60 (-0.82, -0.38) |  | <0.001 | 0.24 (0.01, 0.47) |  | 0.038 | 0.64 (0.36, 0.93) |  | <0.001 |
| Quadratic change * cohort (ref. NCDS) |  | 1.7 | 0.792 |  | 35.1 | <0.001 |  | 17.3 | 0.002 |  | 24.6 | <0.001 |
| NSHD | -0.06 (-0.18, 0.05) |  | 0.283 | 0.01 (-0.10, 0.13) |  | 0.804 | 0.08 (-0.08, 0.23) |  | 0.328 | 0.00 (-0.21, 0.20) |  | 0.981 |
| BCS | -0.01 (-0.08, 0.06) |  | 0.735 | 0.08 (0.01, 0.15) |  | 0.017 | -0.12 (-0.20, -0.05) |  | 0.002 | 0.01 (-0.09, 0.11) |  | 0.906 |
| NS | -0.02 (-0.12, 0.08) |  | 0.743 | 0.16 (0.07, 0.26) |  | <0.001 | -0.09 (-0.19, 0.00) |  | 0.056 | -0.14 (-0.26, -0.03) |  | 0.018 |
| MCS | -0.05 (-0.15, 0.06) |  | 0.360 | 0.28 (0.17, 0.38) |  | <0.001 | -0.15 (-0.25, -0.04) |  | 0.005 | -0.27 (-0.40, -0.14) |  | <0.001 |
| Pre-pandemic financial situation (ref. Doing all right) |  | 132 | <0.001 |  | 217.1 | <0.001 |  | 252.4 | <0.001 |  | 322.6 | <0.001 |
| Experiencing difficulties | 0.61 (0.45, 0.78) |  | <0.001 | 0.87 (0.70, 1.04) |  | <0.001 | 0.95 (0.77, 1.14) |  | <0.001 | -1.40 (-1.64, -1.16) |  | <0.001 |
| Living comfortably | -0.24 (-0.32, -0.16) |  | <0.001 | -0.26 (-0.33, -0.19) |  | <0.001 | -0.37 (-0.46, -0.28) |  | <0.001 | 0.54 (0.43, 0.66) |  | <0.001 |
| Linear change *  pre-pandemic financial situation (ref. Doing all right) |  | 4 | 0.134 |  | 8.7 | 0.013 |  | 0.5 | 0.771 |  | 7.6 | 0.022 |
| Experiencing difficulties | 0.18 (-0.09, 0.44) |  | 0.184 | -0.03 (-0.30, 0.23) |  | 0.807 | -0.10 (-0.38, 0.18) |  | 0.474 | 0.11 (-0.25, 0.48) |  | 0.541 |
| Living comfortably | -0.06 (-0.19, 0.07) |  | 0.363 | -0.17 (-0.28, -0.05) |  | 0.004 | -0.02 (-0.17, 0.12) |  | 0.732 | 0.26 (0.07, 0.44) |  | 0.006 |
| Quadratic change *  pre-pandemic financial situation (ref. Doing all right) |  | 6 | 0.051 |  | 7 | 0.03 |  | 0.2 | 0.93 |  | 15.7 | <0.001 |
| Experiencing difficulties | -0.11 (-0.23, 0.01) |  | 0.081 | -0.01 (-0.13, 0.12) |  | 0.916 | 0.02 (-0.10, 0.15) |  | 0.711 | 0.01 (-0.16, 0.18) |  | 0.872 |
| Living comfortably | 0.03 (-0.03, 0.09) |  | 0.341 | 0.07 (0.01, 0.12) |  | 0.013 | 0.00 (-0.06, 0.07) |  | 0.961 | -0.16 (-0.25, -0.07) |  | <0.001 |
| Cohort (ref. NCDS) *  pre-pandemic financial situation (ref. Doing all right) |  | 4.5 | 0.812 |  | 5.6 | 0.694 |  | 21.2 | 0.007 |  | 13 | 0.113 |
| NSHD * Experiencing difficulties | -0.13 (-0.62, 0.36) |  | 0.606 | -0.20 (-0.79, 0.39) |  | 0.509 | -0.50 (-1.06, 0.06) |  | 0.079 | 0.32 (-0.52, 1.15) |  | 0.459 |
| NSHD * Living comfortably | 0.05 (-0.11, 0.21) |  | 0.562 | -0.04 (-0.21, 0.12) |  | 0.604 | 0.06 (-0.16, 0.28) |  | 0.603 | -0.18 (-0.49, 0.13) |  | 0.255 |
| BCS * Experiencing difficulties | 0.07 (-0.16, 0.30) |  | 0.560 | -0.10 (-0.34, 0.13) |  | 0.377 | -0.01 (-0.26, 0.24) |  | 0.961 | 0.18 (-0.13, 0.50) |  | 0.248 |
| BCS * Living comfortably | 0.01 (-0.10, 0.13) |  | 0.813 | -0.04 (-0.14, 0.07) |  | 0.470 | 0.03 (-0.09, 0.16) |  | 0.606 | -0.03 (-0.19, 0.13) |  | 0.721 |
| NS * Experiencing difficulties | 0.09 (-0.21, 0.38) |  | 0.561 | -0.15 (-0.44, 0.13) |  | 0.299 | -0.19 (-0.49, 0.10) |  | 0.197 | 0.43 (0.05, 0.81) |  | 0.025 |
| NS * Living comfortably | -0.05 (-0.22, 0.12) |  | 0.584 | -0.05 (-0.21, 0.11) |  | 0.536 | -0.02 (-0.19, 0.15) |  | 0.853 | 0.07 (-0.15, 0.28) |  | 0.542 |
| MCS * Experiencing difficulties | -0.09 (-0.37, 0.18) |  | 0.501 | -0.29 (-0.55, -0.02) |  | 0.034 | -0.50 (-0.77, -0.22) |  | <0.001 | 0.53 (0.18, 0.89) |  | 0.003 |
| MCS * Living comfortably | -0.09 (-0.26, 0.08) |  | 0.288 | -0.12 (-0.28, 0.04) |  | 0.130 | 0.00 (-0.17, 0.17) |  | 0.978 | 0.09 (-0.13, 0.31) |  | 0.447 |
| Linear change * cohort (ref. NCDS) *  pre-pandemic financial situation (ref. Doing all right) |  | 5.9 | 0.657 |  | 10.3 | 0.242 |  | 6.12 | 0.634 |  | 11.3 | 0.184 |
| NSHD * Experiencing difficulties | -0.31 (-1.04, 0.43) |  | 0.409 | -0.11 (-1.12, 0.89) |  | 0.827 | 0.41 (-0.21, 1.04) |  | 0.195 | 0.85 (-0.36, 2.06) |  | 0.169 |
| NSHD * Living comfortably | 0.00 (-0.27, 0.28) |  | 0.982 | 0.27 (0.00, 0.53) |  | 0.050 | 0.04 (-0.33, 0.41) |  | 0.840 | -0.13 (-0.64, 0.38) |  | 0.617 |
| BCS * Experiencing difficulties | 0.16 (-0.24, 0.57) |  | 0.432 | 0.26 (-0.14, 0.66) |  | 0.206 | 0.03 (-0.37, 0.44) |  | 0.880 | -0.30 (-0.81, 0.21) |  | 0.252 |
| BCS * Living comfortably | 0.05 (-0.15, 0.24) |  | 0.638 | 0.20 (0.02, 0.37) |  | 0.030 | -0.02 (-0.23, 0.19) |  | 0.832 | -0.09 (-0.36, 0.18) |  | 0.518 |
| NS * Experiencing difficulties | -0.16 (-0.65, 0.33) |  | 0.527 | 0.39 (-0.08, 0.87) |  | 0.106 | 0.36 (-0.12, 0.84) |  | 0.140 | -0.51 (-1.12, 0.11) |  | 0.105 |
| NS * Living comfortably | 0.04 (-0.26, 0.33) |  | 0.805 | 0.12 (-0.16, 0.39) |  | 0.403 | -0.08 (-0.36, 0.20) |  | 0.592 | -0.22 (-0.57, 0.14) |  | 0.239 |
| MCS * Experiencing difficulties | -0.17 (-0.66, 0.31) |  | 0.489 | 0.36 (-0.12, 0.84) |  | 0.146 | 0.10 (-0.38, 0.58) |  | 0.681 | -0.35 (-0.97, 0.26) |  | 0.259 |
| MCS * Living comfortably | 0.22 (-0.10, 0.54) |  | 0.183 | 0.24 (-0.07, 0.55) |  | 0.132 | 0.13 (-0.18, 0.45) |  | 0.408 | -0.47 (-0.87, -0.06) |  | 0.023 |
| Quadratic change * cohort (ref. NCDS) *  pre-pandemic financial situation (ref. Doing all right) |  | 6.6 | 0.582 |  | 7.9 | 0.442 |  | 9.2 | 0.324 |  | 12.3 | 0.138 |
| NSHD * Experiencing difficulties | 0.13 (-0.21, 0.47) |  | 0.464 | 0.01 (-0.45, 0.48) |  | 0.960 | -0.22 (-0.55, 0.11) |  | 0.194 | -0.53 (-1.24, 0.19) |  | 0.149 |
| NSHD * Living comfortably | -0.02 (-0.15, 0.12) |  | 0.811 | -0.10 (-0.23, 0.03) |  | 0.118 | -0.05 (-0.22, 0.13) |  | 0.602 | 0.09 (-0.15, 0.33) |  | 0.451 |
| BCS * Experiencing difficulties | -0.05 (-0.24, 0.13) |  | 0.588 | -0.07 (-0.26, 0.11) |  | 0.454 | -0.02 (-0.21, 0.17) |  | 0.831 | 0.14 (-0.09, 0.37) |  | 0.243 |
| BCS * Living comfortably | -0.02 (-0.11, 0.07) |  | 0.635 | -0.07 (-0.15, 0.01) |  | 0.103 | 0.03 (-0.07, 0.12) |  | 0.608 | 0.06 (-0.07, 0.19) |  | 0.347 |
| NS * Experiencing difficulties | 0.08 (-0.14, 0.30) |  | 0.475 | -0.17 (-0.38, 0.04) |  | 0.122 | -0.17 (-0.38, 0.04) |  | 0.122 | 0.18 (-0.09, 0.45) |  | 0.204 |
| NS * Living comfortably | -0.01 (-0.15, 0.12) |  | 0.829 | -0.03 (-0.15, 0.10) |  | 0.673 | 0.07 (-0.05, 0.20) |  | 0.246 | 0.07 (-0.09, 0.23) |  | 0.417 |
| MCS * Experiencing difficulties | 0.08 (-0.14, 0.31) |  | 0.467 | -0.17 (-0.40, 0.05) |  | 0.132 | -0.01 (-0.24, 0.21) |  | 0.914 | 0.18 (-0.11, 0.46) |  | 0.219 |
| MCS * Living comfortably | -0.13 (-0.27, 0.02) |  | 0.096 | -0.10 (-0.25, 0.04) |  | 0.172 | -0.06 (-0.21, 0.09) |  | 0.415 | 0.25 (0.07, 0.44) |  | 0.007 |
| **Adjusted models** |  | | |  | | |  | | |  | | |
| N participants | 21,312 |  |  | 21,309 |  |  | 21,343 |  |  | 21,381 |  |  |
| N observations | 46,337 |  |  | 46,326 |  |  | 46,395 |  |  | 46,585 |  |  |
|  | ***B* (95% CI)** | **χ2** | ***p*** | ***B* (95% CI)** | **χ2** | ***p*** | ***B* (95% CI)** | **χ2** | ***p*** | ***B* (95% CI)** | **χ2** | ***p*** |
| Time (linear) | 0.12 (0.01, 0.24) |  | 0.040 | 0.04 (-0.06, 0.15) |  | 0.434 | -0.27 (-0.40, -0.14) |  | <0.001 | -0.06 (-0.22, 0.09) |  | 0.425 |
| Time (quadratic) | -0.05 (-0.10, 0.01) |  | 0.083 | 0.02 (-0.03, 0.07) |  | 0.407 | 0.19 (0.13, 0.24) |  | <0.001 | -0.08 (-0.15, -0.01) |  | 0.034 |
| Cohort (ref. NCDS) |  | 260.7 | <0.001 |  | 362.3 | <0.001 |  | 91 | <0.001 |  | 92.9 | <0.001 |
| NSHD | -0.21 (-0.35, -0.08) |  | 0.002 | -0.13 (-0.28, 0.02) |  | 0.092 | -0.10 (-0.31, 0.11) |  | 0.347 | 0.02 (-0.27, 0.30) |  | 0.900 |
| BCS | 0.13 (0.03, 0.22) |  | 0.008 | 0.20 (0.11, 0.28) |  | <0.001 | 0.03 (-0.08, 0.14) |  | 0.592 | -0.20 (-0.33, -0.07) |  | 0.003 |
| NS | 0.52 (0.38, 0.65) |  | <0.001 | 0.61 (0.48, 0.74) |  | <0.001 | 0.31 (0.17, 0.44) |  | <0.001 | -0.43 (-0.60, -0.26) |  | <0.001 |
| MCS | 0.93 (0.80, 1.06) |  | <0.001 | 1.10 (0.97, 1.22) |  | <0.001 | 0.58 (0.45, 0.72) |  | <0.001 | -0.81 (-0.98, -0.64) |  | <0.001 |
| Linear change * cohort (ref. NCDS) |  | 7.9 | 0.094 |  | 24.7 | <0.001 |  | 11.1 | 0.025 |  | 26.1 | <0.001 |
| NSHD | 0.20 (-0.06, 0.47) |  | 0.132 | -0.03 (-0.28, 0.23) |  | 0.826 | -0.09 (-0.45, 0.26) |  | 0.609 | -0.10 (-0.58, 0.38) |  | 0.680 |
| BCS | 0.00 (-0.17, 0.17) |  | 0.990 | -0.26 (-0.42, -0.09) |  | 0.002 | 0.20 (0.02, 0.39) |  | 0.032 | 0.08 (-0.15, 0.31) |  | 0.489 |
| NS | 0.12 (-0.11, 0.36) |  | 0.310 | -0.35 (-0.58, -0.13) |  | 0.002 | 0.16 (-0.07, 0.40) |  | 0.173 | 0.47 (0.18, 0.76) |  | 0.002 |
| MCS | 0.28 (0.04, 0.52) |  | 0.021 | -0.47 (-0.71, -0.24) |  | <0.001 | 0.35 (0.10, 0.59) |  | 0.005 | 0.65 (0.35, 0.95) |  | <0.001 |
| Quadratic change * cohort (ref. NCDS) |  | 2.1 | 0.712 |  | 24.5 | <0.001 |  | 15.7 | 0.004 |  | 21.9 | <0.001 |
| NSHD | -0.07 (-0.21, 0.06) |  | 0.277 | -0.02 (-0.14, 0.10) |  | 0.772 | 0.06 (-0.10, 0.23) |  | 0.468 | 0.04 (-0.19, 0.27) |  | 0.737 |
| BCS | -0.01 (-0.09, 0.07) |  | 0.848 | 0.10 (0.03, 0.18) |  | 0.008 | -0.11 (-0.20, -0.03) |  | 0.009 | -0.02 (-0.13, 0.08) |  | 0.653 |
| NS | -0.04 (-0.15, 0.07) |  | 0.457 | 0.13 (0.03, 0.23) |  | 0.008 | -0.10 (-0.20, 0.01) |  | 0.063 | -0.15 (-0.28, -0.02) |  | 0.024 |
| MCS | -0.05 (-0.16, 0.06) |  | 0.339 | 0.24 (0.13, 0.35) |  | <0.001 | -0.18 (-0.29, -0.07) |  | 0.002 | -0.29 (-0.42, -0.15) |  | <0.001 |
| Pre-pandemic financial situation (ref. Doing all right) |  | 39.2 | <0.001 |  | 73.7 | <0.001 |  | 101 | <0.001 |  | 142.4 | <0.001 |
| Experiencing difficulties | 0.37 (0.20, 0.54) |  | <0.001 | 0.59 (0.42, 0.76) |  | <0.001 | 0.65 (0.46, 0.83) |  | <0.001 | -1.04 (-1.28, -0.79) |  | <0.001 |
| Living comfortably | -0.11 (-0.19, -0.04) |  | 0.004 | -0.11 (-0.18, -0.04) |  | 0.002 | -0.21 (-0.30, -0.12) |  | <0.001 | 0.33 (0.22, 0.45) |  | <0.001 |
| Linear change *  pre-pandemic financial situation (ref. Doing all right) |  | 3.6 | 0.165 |  | 8.6 | 0.014 |  | 0.5 | 0.797 |  | 11.3 | 0.004 |
| Experiencing difficulties | 0.13 (-0.17, 0.42) |  | 0.404 | -0.11 (-0.41, 0.19) |  | 0.468 | -0.10 (-0.39, 0.19) |  | 0.501 | 0.16 (-0.24, 0.57) |  | 0.424 |
| Living comfortably | -0.09 (-0.23, 0.04) |  | 0.186 | -0.18 (-0.31, -0.06) |  | 0.003 | -0.02 (-0.16, 0.13) |  | 0.823 | 0.33 (0.14, 0.52) |  | 0.001 |
| Quadratic change *  pre-pandemic financial situation (ref. Doing all right) |  | 4.9 | 0.085 |  | 7.3 | 0.026 |  | 0.1 | 0.968 |  | 19.4 | <0.001 |
| Experiencing difficulties | -0.09 (-0.22, 0.05) |  | 0.207 | 0.04 (-0.10, 0.17) |  | 0.618 | 0.02 (-0.12, 0.15) |  | 0.818 | -0.02 (-0.21, 0.17) |  | 0.848 |
| Living comfortably | 0.04 (-0.02, 0.10) |  | 0.197 | 0.08 (0.02, 0.14) |  | 0.007 | 0.00 (-0.07, 0.07) |  | 0.984 | -0.19 (-0.28, -0.10) |  | <0.001 |
| Cohort (ref. NCDS) *  pre-pandemic financial situation (ref. Doing all right) |  | 12.7 | 0.123 |  | 10.6 | 0.223 |  | 7.6 | 0.476 |  | 12.5 | 0.132 |
| NSHD * Experiencing difficulties | 0.23 (-0.37, 0.82) |  | 0.451 | 0.19 (-0.47, 0.86) |  | 0.565 | -0.22 (-0.83, 0.38) |  | 0.470 | 0.06 (-0.85, 0.97) |  | 0.897 |
| NSHD * Living comfortably | 0.08 (-0.08, 0.24) |  | 0.328 | 0.00 (-0.17, 0.16) |  | 0.969 | 0.16 (-0.07, 0.40) |  | 0.171 | -0.26 (-0.58, 0.06) |  | 0.110 |
| BCS * Experiencing difficulties | -0.16 (-0.40, 0.07) |  | 0.173 | -0.29 (-0.52, -0.05) |  | 0.019 | -0.19 (-0.44, 0.07) |  | 0.153 | 0.34 (0.02, 0.67) |  | 0.039 |
| BCS * Living comfortably | 0.03 (-0.09, 0.14) |  | 0.632 | -0.01 (-0.12, 0.10) |  | 0.870 | 0.02 (-0.11, 0.15) |  | 0.804 | 0.01 (-0.16, 0.18) |  | 0.912 |
| NS * Experiencing difficulties | 0.20 (-0.10, 0.50) |  | 0.194 | -0.06 (-0.36, 0.23) |  | 0.683 | -0.11 (-0.41, 0.18) |  | 0.452 | 0.23 (-0.16, 0.61) |  | 0.256 |
| NS * Living comfortably | -0.04 (-0.21, 0.13) |  | 0.638 | -0.09 (-0.25, 0.07) |  | 0.271 | -0.04 (-0.21, 0.13) |  | 0.658 | 0.15 (-0.07, 0.37) |  | 0.185 |
| MCS * Experiencing difficulties | 0.01 (-0.27, 0.28) |  | 0.965 | -0.13 (-0.40, 0.14) |  | 0.342 | -0.25 (-0.52, 0.02) |  | 0.072 | 0.26 (-0.11, 0.63) |  | 0.163 |
| MCS * Living comfortably | -0.10 (-0.26, 0.07) |  | 0.240 | -0.11 (-0.26, 0.05) |  | 0.194 | -0.05 (-0.23, 0.12) |  | 0.544 | 0.18 (-0.04, 0.40) |  | 0.109 |
| Linear change * cohort (ref. NCDS) *  pre-pandemic financial situation (ref. Doing all right) |  | 10.1 | 0.259 |  | 12.6 | 0.125 |  | 6.9 | 0.55 |  | 10.2 | 0.248 |
| NSHD * Experiencing difficulties | -0.48 (-1.41, 0.45) |  | 0.314 | -0.26 (-1.35, 0.82) |  | 0.634 | 0.13 (-0.62, 0.88) |  | 0.741 | 1.41 (-0.56, 3.39) |  | 0.161 |
| NSHD * Living comfortably | 0.04 (-0.27, 0.34) |  | 0.822 | 0.25 (-0.04, 0.54) |  | 0.091 | 0.01 (-0.39, 0.41) |  | 0.952 | -0.08 (-0.63, 0.48) |  | 0.782 |
| BCS * Experiencing difficulties | 0.44 (-0.02, 0.89) |  | 0.060 | 0.55 (0.10, 0.99) |  | 0.017 | 0.13 (-0.31, 0.57) |  | 0.565 | -0.40 (-0.96, 0.16) |  | 0.160 |
| BCS * Living comfortably | 0.08 (-0.13, 0.29) |  | 0.481 | 0.27 (0.07, 0.46) |  | 0.008 | 0.01 (-0.21, 0.24) |  | 0.909 | -0.18 (-0.47, 0.12) |  | 0.243 |
| NS * Experiencing difficulties | -0.17 (-0.70, 0.37) |  | 0.540 | 0.36 (-0.16, 0.88) |  | 0.171 | 0.44 (-0.06, 0.95) |  | 0.083 | -0.43 (-1.09, 0.23) |  | 0.204 |
| NS * Living comfortably | 0.01 (-0.30, 0.32) |  | 0.948 | 0.14 (-0.15, 0.43) |  | 0.357 | -0.13 (-0.43, 0.17) |  | 0.383 | -0.29 (-0.67, 0.10) |  | 0.140 |
| MCS * Experiencing difficulties | -0.11 (-0.64, 0.42) |  | 0.686 | 0.32 (-0.21, 0.85) |  | 0.232 | 0.00 (-0.51, 0.51) |  | 1.000 | -0.31 (-0.98, 0.37) |  | 0.375 |
| MCS * Living comfortably | 0.22 (-0.12, 0.56) |  | 0.201 | 0.14 (-0.20, 0.47) |  | 0.414 | 0.08 (-0.27, 0.42) |  | 0.665 | -0.48 (-0.91, -0.06) |  | 0.026 |
| Quadratic change * cohort (ref. NCDS) *  pre-pandemic financial situation (ref. Doing all right) |  | 10.2 | 0.252 |  | 10 | 0.262 |  | 11.9 | 0.154 |  | 11.4 | 0.179 |
| NSHD * Experiencing difficulties | 0.18 (-0.21, 0.57) |  | 0.371 | 0.08 (-0.42, 0.57) |  | 0.765 | -0.11 (-0.51, 0.30) |  | 0.608 | -0.86 (-2.12, 0.39) |  | 0.178 |
| NSHD * Living comfortably | -0.02 (-0.18, 0.13) |  | 0.778 | -0.09 (-0.22, 0.05) |  | 0.200 | -0.04 (-0.23, 0.14) |  | 0.651 | 0.04 (-0.22, 0.30) |  | 0.764 |
| BCS * Experiencing difficulties | -0.17 (-0.38, 0.04) |  | 0.108 | -0.22 (-0.43, -0.01) |  | 0.040 | -0.03 (-0.24, 0.17) |  | 0.760 | 0.19 (-0.07, 0.45) |  | 0.157 |
| BCS * Living comfortably | -0.03 (-0.13, 0.06) |  | 0.502 | -0.11 (-0.20, -0.02) |  | 0.016 | 0.01 (-0.09, 0.12) |  | 0.834 | 0.10 (-0.04, 0.24) |  | 0.169 |
| NS * Experiencing difficulties | 0.08 (-0.16, 0.31) |  | 0.514 | -0.16 (-0.39, 0.07) |  | 0.176 | -0.20 (-0.42, 0.02) |  | 0.077 | 0.16 (-0.14, 0.45) |  | 0.304 |
| NS * Living comfortably | 0.00 (-0.14, 0.14) |  | 0.961 | -0.03 (-0.16, 0.10) |  | 0.675 | 0.11 (-0.03, 0.24) |  | 0.118 | 0.09 (-0.08, 0.26) |  | 0.318 |
| MCS * Experiencing difficulties | 0.05 (-0.19, 0.30) |  | 0.674 | -0.17 (-0.42, 0.08) |  | 0.189 | 0.03 (-0.22, 0.27) |  | 0.836 | 0.16 (-0.16, 0.47) |  | 0.331 |
| MCS * Living comfortably | -0.14 (-0.29, 0.02) |  | 0.085 | -0.08 (-0.24, 0.08) |  | 0.324 | -0.05 (-0.22, 0.11) |  | 0.511 | 0.27 (0.08, 0.46) |  | 0.006 |
| **Sensitivity models** |  | | |  | | |  | | |  | | |
| N participants | 21,312 |  |  | 21,309 |  |  | 21,343 |  |  | 21,381 |  |  |
| N observations | 46,337 |  |  | 46,326 |  |  | 46,395 |  |  | 46,585 |  |  |
|  | ***B* (95% CI)** | **χ2** | ***p*** | ***B* (95% CI)** | **χ2** | ***p*** | ***B* (95% CI)** | **χ2** | ***p*** | ***B* (95% CI)** | **χ2** | ***p*** |
| Time (linear) | 0.12 (0.01, 0.24) |  | 0.036 | 0.04 (-0.06, 0.15) |  | 0.426 | -0.27 (-0.40, -0.14) |  | <0.001 | -0.06 (-0.22, 0.10) |  | 0.451 |
| Time (quadratic) | -0.05 (-0.10, 0.01) |  | 0.079 | 0.02 (-0.03, 0.07) |  | 0.409 | 0.18 (0.13, 0.24) |  | <0.001 | -0.08 (-0.16, -0.01) |  | 0.028 |
| Cohort (ref. NCDS) |  | 346.3 | <0.001 |  | 437.7 | <0.001 |  | 269.4 | <0.001 |  | 232.6 | <0.001 |
| NSHD | -0.20 (-0.35, -0.06) |  | 0.006 | -0.08 (-0.24, 0.08) |  | 0.314 | -0.10 (-0.32, 0.11) |  | 0.345 | 0.06 (-0.23, 0.35) |  | 0.682 |
| BCS | 0.14 (0.04, 0.23) |  | 0.006 | 0.16 (0.07, 0.26) |  | <0.001 | -0.04 (-0.15, 0.07) |  | 0.490 | -0.15 (-0.29, -0.02) |  | 0.027 |
| NS | 0.66 (0.52, 0.79) |  | <0.001 | 0.69 (0.56, 0.82) |  | <0.001 | 0.41 (0.27, 0.55) |  | <0.001 | -0.56 (-0.73, -0.38) |  | <0.001 |
| MCS | 1.00 (0.88, 1.13) |  | <0.001 | 1.15 (1.03, 1.27) |  | <0.001 | 0.93 (0.80, 1.06) |  | <0.001 | -1.19 (-1.35, -1.02) |  | <0.001 |
| Linear change * cohort (ref. NCDS) |  | 6.7 | 0.151 |  | 26.2 | <0.001 |  | 10.3 | 0.036 |  | 26.1 | <0.001 |
| NSHD | 0.21 (-0.06, 0.47) |  | 0.133 | -0.02 (-0.28, 0.23) |  | 0.872 | -0.09 (-0.44, 0.26) |  | 0.623 | -0.11 (-0.60, 0.37) |  | 0.646 |
| BCS | 0.00 (-0.17, 0.17) |  | 0.963 | -0.26 (-0.42, -0.10) |  | 0.001 | 0.20 (0.01, 0.38) |  | 0.036 | 0.09 (-0.14, 0.33) |  | 0.441 |
| NS | 0.10 (-0.14, 0.34) |  | 0.402 | -0.37 (-0.59, -0.14) |  | 0.001 | 0.16 (-0.08, 0.40) |  | 0.188 | 0.47 (0.18, 0.77) |  | 0.001 |
| MCS | 0.25 (0.01, 0.50) |  | 0.041 | -0.49 (-0.73, -0.25) |  | <0.001 | 0.33 (0.09, 0.58) |  | 0.008 | 0.66 (0.35, 0.96) |  | <0.001 |
| Quadratic change * cohort (ref. NCDS) |  | 1.9 | 0.753 |  | 25.1 | <0.001 |  | 15.3 | 0.004 |  | 20.5 | <0.001 |
| NSHD | -0.07 (-0.21, 0.06) |  | 0.281 | -0.02 (-0.14, 0.10) |  | 0.730 | 0.06 (-0.11, 0.23) |  | 0.478 | 0.05 (-0.18, 0.27) |  | 0.696 |
| BCS | -0.01 (-0.08, 0.07) |  | 0.899 | 0.11 (0.03, 0.18) |  | 0.005 | -0.11 (-0.20, -0.03) |  | 0.011 | -0.03 (-0.14, 0.08) |  | 0.603 |
| NS | -0.03 (-0.14, 0.08) |  | 0.553 | 0.14 (0.04, 0.24) |  | 0.006 | -0.10 (-0.20, 0.01) |  | 0.075 | -0.16 (-0.29, -0.02) |  | 0.020 |
| MCS | -0.05 (-0.16, 0.06) |  | 0.385 | 0.24 (0.12, 0.35) |  | <0.001 | -0.18 (-0.29, -0.07) |  | 0.002 | -0.28 (-0.42, -0.14) |  | <0.001 |
| Pre-pandemic financial situation (ref. Doing all right) |  | 110.8 | <0.001 |  | 192.6 | <0.001 |  | 237.6 | <0.001 |  | 268.2 | <0.001 |
| Experiencing difficulties | 0.61 (0.43, 0.79) |  | <0.001 | 0.87 (0.68, 1.05) |  | <0.001 | 0.98 (0.78, 1.17) |  | <0.001 | -1.40 (-1.66, -1.14) |  | <0.001 |
| Living comfortably | -0.22 (-0.30, -0.14) |  | <0.001 | -0.27 (-0.34, -0.19) |  | <0.001 | -0.38 (-0.47, -0.28) |  | <0.001 | 0.51 (0.40, 0.63) |  | <0.001 |
| Linear change *  pre-pandemic financial situation (ref. Doing all right) |  | 4 | 0.139 |  | 8.5 | 0.014 |  | 0.2 | 0.89 |  | 10.6 | 0.005 |
| Experiencing difficulties | 0.14 (-0.16, 0.44) |  | 0.355 | -0.09 (-0.39, 0.21) |  | 0.563 | -0.07 (-0.36, 0.22) |  | 0.629 | 0.13 (-0.28, 0.54) |  | 0.531 |
| Living comfortably | -0.09 (-0.23, 0.04) |  | 0.179 | -0.18 (-0.30, -0.06) |  | 0.004 | -0.01 (-0.16, 0.13) |  | 0.849 | 0.32 (0.12, 0.51) |  | 0.001 |
| Quadratic change *  pre-pandemic financial situation (ref. Doing all right) |  | 5.3 | 0.07 |  | 5 | 0.023 |  | 0 | 0.994 |  | 18.7 | <0.001 |
| Experiencing difficulties | -0.09 (-0.23, 0.04) |  | 0.183 | 0.03 (-0.11, 0.17) |  | 0.699 | 0.01 (-0.13, 0.14) |  | 0.929 | -0.01 (-0.20, 0.18) |  | 0.952 |
| Living comfortably | 0.04 (-0.02, 0.10) |  | 0.184 | 0.08 (0.02, 0.14) |  | 0.007 | 0.00 (-0.07, 0.07) |  | 0.984 | -0.19 (-0.28, -0.10) |  | <0.001 |
| Cohort (ref. NCDS) *  pre-pandemic financial situation (ref. Doing all right) |  | 6 | 0.653 |  | 5 | 0.756 |  | 15.4 | 0.053 |  | 11.1 | 0.194 |
| NSHD * Experiencing difficulties | -0.03 (-0.70, 0.64) |  | 0.927 | -0.09 (-0.77, 0.59) |  | 0.795 | -0.49 (-1.14, 0.16) |  | 0.137 | 0.39 (-0.57, 1.34) |  | 0.428 |
| NSHD * Living comfortably | 0.06 (-0.11, 0.22) |  | 0.498 | -0.03 (-0.21, 0.14) |  | 0.708 | 0.14 (-0.10, 0.38) |  | 0.261 | -0.23 (-0.57, 0.10) |  | 0.167 |
| BCS * Experiencing difficulties | -0.08 (-0.33, 0.17) |  | 0.542 | -0.21 (-0.47, 0.04) |  | 0.100 | -0.12 (-0.39, 0.15) |  | 0.380 | 0.28 (-0.06, 0.62) |  | 0.103 |
| BCS * Living comfortably | 0.01 (-0.11, 0.13) |  | 0.864 | -0.01 (-0.12, 0.11) |  | 0.903 | 0.03 (-0.11, 0.17) |  | 0.662 | 0.00 (-0.18, 0.17) |  | 0.976 |
| NS * Experiencing difficulties | 0.14 (-0.17, 0.45) |  | 0.384 | -0.13 (-0.44, 0.18) |  | 0.410 | -0.22 (-0.53, 0.09) |  | 0.173 | 0.32 (-0.09, 0.72) |  | 0.128 |
| NS * Living comfortably | -0.07 (-0.24, 0.11) |  | 0.459 | -0.08 (-0.25, 0.09) |  | 0.341 | -0.02 (-0.20, 0.16) |  | 0.859 | 0.11 (-0.11, 0.34) |  | 0.322 |
| MCS * Experiencing difficulties | -0.10 (-0.39, 0.20) |  | 0.526 | -0.24 (-0.53, 0.04) |  | 0.097 | -0.46 (-0.76, -0.17) |  | 0.002 | 0.50 (0.12, 0.89) |  | 0.010 |
| MCS * Living comfortably | -0.10 (-0.27, 0.08) |  | 0.275 | -0.07 (-0.23, 0.10) |  | 0.436 | 0.02 (-0.16, 0.20) |  | 0.807 | 0.11 (-0.12, 0.34) |  | 0.344 |
| Linear change * cohort (ref. NCDS) *  pre-pandemic financial situation (ref. Doing all right) |  | 10.4 | 0.236 |  | 12.2 | 0.142 |  | 6.6 | 0.585 |  | 10.2 | 0.253 |
| NSHD * Experiencing difficulties | -0.48 (-1.44, 0.47) |  | 0.322 | -0.25 (-1.31, 0.81) |  | 0.643 | 0.14 (-0.60, 0.88) |  | 0.714 | 1.44 (-0.56, 3.45) |  | 0.158 |
| NSHD * Living comfortably | 0.03 (-0.29, 0.34) |  | 0.873 | 0.24 (-0.05, 0.53) |  | 0.109 | 0.01 (-0.39, 0.40) |  | 0.978 | -0.06 (-0.61, 0.50) |  | 0.845 |
| BCS * Experiencing difficulties | 0.42 (-0.04, 0.89) |  | 0.071 | 0.54 (0.08, 0.99) |  | 0.020 | 0.10 (-0.35, 0.55) |  | 0.659 | -0.36 (-0.93, 0.20) |  | 0.206 |
| BCS * Living comfortably | 0.07 (-0.14, 0.28) |  | 0.510 | 0.27 (0.07, 0.47) |  | 0.007 | 0.01 (-0.22, 0.23) |  | 0.954 | -0.18 (-0.47, 0.12) |  | 0.244 |
| NS * Experiencing difficulties | -0.19 (-0.72, 0.35) |  | 0.488 | 0.35 (-0.17, 0.88) |  | 0.190 | 0.43 (-0.07, 0.94) |  | 0.094 | -0.39 (-1.06, 0.28) |  | 0.254 |
| NS * Living comfortably | 0.03 (-0.28, 0.35) |  | 0.844 | 0.15 (-0.14, 0.44) |  | 0.318 | -0.12 (-0.42, 0.18) |  | 0.421 | -0.29 (-0.68, 0.10) |  | 0.141 |
| MCS * Experiencing difficulties | -0.11 (-0.65, 0.43) |  | 0.695 | 0.30 (-0.24, 0.85) |  | 0.272 | -0.02 (-0.54, 0.50) |  | 0.951 | -0.27 (-0.95, 0.42) |  | 0.444 |
| MCS * Living comfortably | 0.26 (-0.09, 0.60) |  | 0.142 | 0.16 (-0.18, 0.50) |  | 0.352 | 0.09 (-0.25, 0.44) |  | 0.596 | -0.50 (-0.93, -0.07) |  | 0.022 |
| Quadratic change * cohort (ref. NCDS) *  pre-pandemic financial situation (ref. Doing all right) |  | 10.6 | 0.226 |  | 9.8 | 0.277 |  | 11.6 | 0.168 |  | 11.5 | 0.176 |
| NSHD * Experiencing difficulties | 0.18 (-0.22, 0.58) |  | 0.374 | 0.07 (-0.41, 0.56) |  | 0.774 | -0.11 (-0.51, 0.29) |  | 0.587 | -0.88 (-2.15, 0.39) |  | 0.176 |
| NSHD * Living comfortably | -0.02 (-0.18, 0.14) |  | 0.796 | -0.09 (-0.22, 0.05) |  | 0.216 | -0.04 (-0.23, 0.15) |  | 0.652 | 0.03 (-0.23, 0.30) |  | 0.807 |
| BCS * Experiencing difficulties | -0.17 (-0.38, 0.05) |  | 0.122 | -0.22 (-0.43, 0.00) |  | 0.046 | -0.02 (-0.23, 0.18) |  | 0.818 | 0.18 (-0.08, 0.44) |  | 0.183 |
| BCS * Living comfortably | -0.03 (-0.13, 0.07) |  | 0.533 | -0.11 (-0.21, -0.02) |  | 0.014 | 0.01 (-0.09, 0.12) |  | 0.803 | 0.10 (-0.04, 0.24) |  | 0.169 |
| NS * Experiencing difficulties | 0.08 (-0.15, 0.32) |  | 0.483 | -0.16 (-0.39, 0.07) |  | 0.180 | -0.20 (-0.43, 0.02) |  | 0.077 | 0.15 (-0.15, 0.45) |  | 0.329 |
| NS * Living comfortably | -0.01 (-0.15, 0.14) |  | 0.930 | -0.04 (-0.17, 0.10) |  | 0.597 | 0.10 (-0.03, 0.23) |  | 0.147 | 0.09 (-0.08, 0.27) |  | 0.299 |
| MCS * Experiencing difficulties | 0.05 (-0.19, 0.30) |  | 0.680 | -0.16 (-0.41, 0.10) |  | 0.221 | 0.03 (-0.21, 0.28) |  | 0.790 | 0.13 (-0.18, 0.45) |  | 0.411 |
| MCS * Living comfortably | -0.15 (-0.31, 0.01) |  | 0.062 | -0.09 (-0.24, 0.07) |  | 0.289 | -0.06 (-0.22, 0.10) |  | 0.468 | 0.27 (0.08, 0.47) |  | 0.006 |

*Note.* Adjusted models included birth sex, highest qualification achieved, pre-pandemic self-reported health, pre-pandemic psychological distress, and household composition as covariates. Sensitivity models correspond to the unadjusted models after restricting the analytical sample to that of the adjusted models. BCS: British Cohort Study, 1970 birth cohort; GAD-2: 2-item General Anxiety Disorder questionnaire; MCS: Millennium Cohort Study, 2000 birth cohort; NCDS: National Child and Development Study, 1958 birth cohort; NS: Next Steps, 1990 cohort; NSHD: National Survey of Health and Development, 1946 birth cohort; ONS: UK Office for National Statistics; PHQ-2: 2-item Patient Health Questionnaire; UCLA-3: 3-item UCLA loneliness scale. χ2: Wald test performed to assess the overall statistical significance of the interaction terms; χ2 statistics have 4 degrees of freedom for time*cohort interaction terms, 2 degrees of freedom for time*pre-pandemic financial situation interaction terms; and 8 degrees of freedom for the rest of interaction terms.

#### Table S7.2. Unadjusted and adjusted marginal mean estimates and 95% confidence intervals by pre-pandemic financial situation.

|  |  |  | **Anxiety symptomatology (GAD-2)** | **Depressive symptomatology (PHQ-2)** | **Feelings of loneliness (UCLA-3)** | **Life satisfaction (ONS single question)** |
| --- | --- | --- | --- | --- | --- | --- |
| **Cohort** | **Pre-pandemic financial situation** | **Survey wave** | **Unadjusted marginal mean (95% CI)** | **Unadjusted marginal mean (95% CI)** | **Unadjusted marginal mean (95% CI)** | **Unadjusted marginal mean (95% CI)** |
| NSHD | Experiencing difficulties | 1 | 1.10 (0.65, 1.55) | 1.33 (0.78, 1.88) | 4.62 (4.12, 5.11) | 6.25 (5.50, 7.01) |
| NSHD | Experiencing difficulties | 2 | 1.18 (0.73, 1.62) | 1.19 (0.76, 1.62) | 4.61 (4.09, 5.13) | 6.53 (5.80, 7.26) |
| NSHD | Experiencing difficulties | 3 | 1.09 (0.57, 1.61) | 1.13 (0.70, 1.56) | 4.73 (4.19, 5.28) | 5.57 (4.66, 6.47) |
| NSHD | Doing all right | 1 | 0.62 (0.49, 0.74) | 0.66 (0.52, 0.80) | 4.17 (3.99, 4.35) | 7.34 (7.08, 7.59) |
| NSHD | Doing all right | 2 | 0.80 (0.65, 0.95) | 0.66 (0.53, 0.80) | 4.05 (3.87, 4.22) | 7.16 (6.93, 7.40) |
| NSHD | Doing all right | 3 | 0.79 (0.63, 0.95) | 0.73 (0.59, 0.88) | 4.45 (4.25, 4.64) | 6.77 (6.52, 7.03) |
| NSHD | Living comfortably | 1 | 0.42 (0.36, 0.49) | 0.36 (0.30, 0.41) | 3.86 (3.77, 3.95) | 7.70 (7.57, 7.83) |
| NSHD | Living comfortably | 2 | 0.56 (0.50, 0.63) | 0.42 (0.36, 0.48) | 3.70 (3.62, 3.79) | 7.59 (7.47, 7.71) |
| NSHD | Living comfortably | 3 | 0.53 (0.46, 0.60) | 0.49 (0.42, 0.55) | 3.99 (3.89, 4.08) | 7.12 (6.99, 7.25) |
| NCDS | Experiencing difficulties | 1 | 1.43 (1.27, 1.58) | 1.59 (1.43, 1.75) | 5.20 (5.03, 5.37) | 5.90 (5.68, 6.13) |
| NCDS | Experiencing difficulties | 2 | 1.55 (1.41, 1.69) | 1.62 (1.47, 1.77) | 5.04 (4.89, 5.19) | 5.92 (5.73, 6.10) |
| NCDS | Experiencing difficulties | 3 | 1.38 (1.25, 1.52) | 1.67 (1.53, 1.81) | 5.30 (5.14, 5.45) | 5.74 (5.56, 5.93) |
| NCDS | Doing all right | 1 | 0.81 (0.74, 0.88) | 0.72 (0.66, 0.78) | 4.25 (4.17, 4.33) | 7.31 (7.21, 7.40) |
| NCDS | Doing all right | 2 | 0.87 (0.80, 0.93) | 0.79 (0.73, 0.85) | 4.17 (4.10, 4.23) | 7.19 (7.10, 7.27) |
| NCDS | Doing all right | 3 | 0.85 (0.79, 0.90) | 0.90 (0.84, 0.95) | 4.45 (4.38, 4.52) | 6.86 (6.77, 6.94) |
| NCDS | Living comfortably | 1 | 0.57 (0.53, 0.61) | 0.46 (0.42, 0.50) | 3.88 (3.83, 3.93) | 7.85 (7.78, 7.91) |
| NCDS | Living comfortably | 2 | 0.59 (0.56, 0.63) | 0.43 (0.40, 0.46) | 3.77 (3.73, 3.81) | 7.83 (7.77, 7.88) |
| NCDS | Living comfortably | 3 | 0.60 (0.56, 0.64) | 0.57 (0.54, 0.60) | 4.04 (3.99, 4.08) | 7.27 (7.21, 7.33) |
| BCS | Experiencing difficulties | 1 | 1.60 (1.45, 1.75) | 1.66 (1.51, 1.80) | 5.15 (5.00, 5.30) | 5.98 (5.79, 6.16) |
| BCS | Experiencing difficulties | 2 | 1.86 (1.73, 1.99) | 1.75 (1.61, 1.88) | 5.13 (4.99, 5.27) | 5.83 (5.67, 5.98) |
| BCS | Experiencing difficulties | 3 | 1.70 (1.58, 1.82) | 1.88 (1.75, 2.00) | 5.24 (5.11, 5.36) | 5.78 (5.63, 5.93) |
| BCS | Doing all right | 1 | 0.92 (0.85, 0.98) | 0.89 (0.83, 0.96) | 4.21 (4.14, 4.28) | 7.19 (7.10, 7.28) |
| BCS | Doing all right | 2 | 0.99 (0.94, 1.05) | 0.84 (0.78, 0.89) | 4.25 (4.18, 4.32) | 7.07 (7.00, 7.15) |
| BCS | Doing all right | 3 | 0.98 (0.92, 1.03) | 0.98 (0.92, 1.03) | 4.41 (4.35, 4.48) | 6.75 (6.67, 6.83) |
| BCS | Living comfortably | 1 | 0.69 (0.64, 0.74) | 0.59 (0.55, 0.64) | 3.87 (3.81, 3.93) | 7.71 (7.63, 7.78) |
| BCS | Living comfortably | 2 | 0.76 (0.71, 0.81) | 0.56 (0.52, 0.61) | 3.89 (3.84, 3.95) | 7.65 (7.58, 7.73) |
| BCS | Living comfortably | 3 | 0.75 (0.70, 0.80) | 0.73 (0.68, 0.78) | 4.09 (4.03, 4.15) | 7.20 (7.13, 7.28) |
| NS | Experiencing difficulties | 1 | 2.16 (1.95, 2.37) | 2.12 (1.92, 2.33) | 5.41 (5.21, 5.61) | 5.79 (5.53, 6.05) |
| NS | Experiencing difficulties | 2 | 2.27 (2.12, 2.42) | 2.14 (1.99, 2.29) | 5.51 (5.37, 5.66) | 5.76 (5.58, 5.94) |
| NS | Experiencing difficulties | 3 | 2.22 (2.08, 2.36) | 2.17 (2.04, 2.31) | 5.51 (5.39, 5.64) | 5.61 (5.45, 5.78) |
| NS | Doing all right | 1 | 1.46 (1.35, 1.57) | 1.41 (1.30, 1.52) | 4.65 (4.54, 4.76) | 6.76 (6.62, 6.89) |
| NS | Doing all right | 2 | 1.58 (1.49, 1.67) | 1.24 (1.16, 1.32) | 4.64 (4.55, 4.72) | 6.93 (6.83, 7.03) |
| NS | Doing all right | 3 | 1.60 (1.51, 1.68) | 1.43 (1.36, 1.51) | 4.80 (4.72, 4.89) | 6.61 (6.51, 6.70) |
| NS | Living comfortably | 1 | 1.17 (1.07, 1.27) | 1.10 (1.00, 1.19) | 4.27 (4.17, 4.36) | 7.37 (7.24, 7.49) |
| NS | Living comfortably | 2 | 1.29 (1.20, 1.37) | 0.92 (0.84, 0.99) | 4.22 (4.15, 4.30) | 7.49 (7.39, 7.59) |
| NS | Living comfortably | 3 | 1.32 (1.24, 1.40) | 1.19 (1.11, 1.26) | 4.51 (4.44, 4.59) | 6.92 (6.82, 7.02) |
| MCS | Experiencing difficulties | 1 | 2.31 (2.12, 2.50) | 2.51 (2.33, 2.69) | 5.66 (5.49, 5.84) | 5.23 (5.00, 5.46) |
| MCS | Experiencing difficulties | 2 | 2.56 (2.39, 2.73) | 2.40 (2.23, 2.56) | 5.68 (5.52, 5.85) | 5.44 (5.25, 5.64) |
| MCS | Experiencing difficulties | 3 | 2.58 (2.44, 2.73) | 2.52 (2.39, 2.66) | 5.80 (5.66, 5.94) | 5.27 (5.10, 5.45) |
| MCS | Doing all right | 1 | 1.79 (1.69, 1.89) | 1.93 (1.83, 2.02) | 5.21 (5.11, 5.31) | 6.10 (5.97, 6.23) |
| MCS | Doing all right | 2 | 2.06 (1.96, 2.15) | 1.67 (1.58, 1.76) | 5.21 (5.12, 5.31) | 6.36 (6.25, 6.46) |
| MCS | Doing all right | 3 | 2.15 (2.06, 2.23) | 2.01 (1.93, 2.09) | 5.30 (5.22, 5.38) | 5.86 (5.76, 5.96) |
| MCS | Living comfortably | 1 | 1.46 (1.35, 1.57) | 1.54 (1.44, 1.65) | 4.83 (4.72, 4.94) | 6.73 (6.59, 6.87) |
| MCS | Living comfortably | 2 | 1.78 (1.68, 1.89) | 1.33 (1.23, 1.42) | 4.89 (4.78, 5.00) | 6.87 (6.75, 6.99) |
| MCS | Living comfortably | 3 | 1.74 (1.65, 1.83) | 1.64 (1.55, 1.73) | 4.90 (4.81, 4.99) | 6.44 (6.33, 6.55) |
| **Cohort** | **Pre-pandemic financial situation** | **Survey wave** | **Adjusted marginal mean (95% CI)** | **Adjusted marginal mean (95% CI)** | **Adjusted marginal mean (95% CI)** | **Adjusted marginal mean (95% CI)** |
| NSHD | Experiencing difficulties | 1 | 1.17 (0.61, 1.72) | 1.38 (0.75, 2.00) | 4.60 (4.06, 5.15) | 6.33 (5.50, 7.16) |
| NSHD | Experiencing difficulties | 2 | 1.11 (0.66, 1.55) | 1.13 (0.70, 1.56) | 4.42 (3.91, 4.94) | 6.82 (5.84, 7.80) |
| NSHD | Experiencing difficulties | 3 | 0.99 (0.40, 1.57) | 1.11 (0.66, 1.57) | 4.56 (4.01, 5.11) | 5.47 (4.15, 6.78) |
| NSHD | Doing all right | 1 | 0.57 (0.44, 0.69) | 0.59 (0.45, 0.73) | 4.18 (3.98, 4.38) | 7.31 (7.03, 7.58) |
| NSHD | Doing all right | 2 | 0.77 (0.62, 0.92) | 0.61 (0.47, 0.74) | 4.06 (3.88, 4.25) | 7.10 (6.85, 7.35) |
| NSHD | Doing all right | 3 | 0.73 (0.56, 0.91) | 0.63 (0.49, 0.77) | 4.44 (4.23, 4.66) | 6.81 (6.54, 7.08) |
| NSHD | Living comfortably | 1 | 0.54 (0.46, 0.61) | 0.48 (0.42, 0.54) | 4.14 (4.04, 4.24) | 7.38 (7.24, 7.52) |
| NSHD | Living comfortably | 2 | 0.70 (0.62, 0.78) | 0.55 (0.48, 0.62) | 3.97 (3.88, 4.07) | 7.27 (7.14, 7.40) |
| NSHD | Living comfortably | 3 | 0.66 (0.58, 0.74) | 0.61 (0.54, 0.68) | 4.21 (4.11, 4.32) | 6.77 (6.62, 6.91) |
| NCDS | Experiencing difficulties | 1 | 1.15 (1.00, 1.31) | 1.31 (1.15, 1.48) | 4.93 (4.76, 5.10) | 6.25 (6.02, 6.48) |
| NCDS | Experiencing difficulties | 2 | 1.26 (1.13, 1.40) | 1.30 (1.16, 1.44) | 4.76 (4.61, 4.91) | 6.25 (6.07, 6.44) |
| NCDS | Experiencing difficulties | 3 | 1.11 (0.98, 1.24) | 1.40 (1.26, 1.54) | 4.99 (4.84, 5.14) | 6.06 (5.87, 6.25) |
| NCDS | Doing all right | 1 | 0.78 (0.72, 0.85) | 0.72 (0.66, 0.78) | 4.28 (4.21, 4.36) | 7.29 (7.20, 7.38) |
| NCDS | Doing all right | 2 | 0.86 (0.80, 0.92) | 0.78 (0.73, 0.84) | 4.20 (4.13, 4.27) | 7.14 (7.06, 7.23) |
| NCDS | Doing all right | 3 | 0.84 (0.78, 0.89) | 0.89 (0.83, 0.95) | 4.48 (4.41, 4.55) | 6.84 (6.75, 6.93) |
| NCDS | Living comfortably | 1 | 0.67 (0.63, 0.71) | 0.61 (0.57, 0.65) | 4.07 (4.02, 4.13) | 7.62 (7.55, 7.69) |
| NCDS | Living comfortably | 2 | 0.69 (0.65, 0.74) | 0.57 (0.53, 0.60) | 3.97 (3.92, 4.02) | 7.61 (7.55, 7.67) |
| NCDS | Living comfortably | 3 | 0.71 (0.66, 0.75) | 0.73 (0.69, 0.76) | 4.24 (4.19, 4.29) | 7.05 (6.99, 7.12) |
| BCS | Experiencing difficulties | 1 | 1.12 (0.97, 1.27) | 1.22 (1.07, 1.37) | 4.77 (4.61, 4.93) | 6.39 (6.20, 6.59) |
| BCS | Experiencing difficulties | 2 | 1.49 (1.34, 1.63) | 1.38 (1.24, 1.53) | 4.79 (4.64, 4.94) | 6.24 (6.06, 6.41) |
| BCS | Experiencing difficulties | 3 | 1.23 (1.10, 1.36) | 1.42 (1.29, 1.55) | 4.92 (4.77, 5.06) | 6.21 (6.05, 6.37) |
| BCS | Doing all right | 1 | 0.91 (0.84, 0.98) | 0.92 (0.85, 0.98) | 4.31 (4.23, 4.39) | 7.09 (6.99, 7.19) |
| BCS | Doing all right | 2 | 0.98 (0.91, 1.04) | 0.83 (0.76, 0.89) | 4.31 (4.24, 4.39) | 7.00 (6.92, 7.09) |
| BCS | Doing all right | 3 | 0.94 (0.87, 1.00) | 0.98 (0.92, 1.04) | 4.46 (4.39, 4.53) | 6.70 (6.61, 6.79) |
| BCS | Living comfortably | 1 | 0.83 (0.77, 0.89) | 0.80 (0.74, 0.85) | 4.12 (4.05, 4.19) | 7.43 (7.34, 7.52) |
| BCS | Living comfortably | 2 | 0.89 (0.83, 0.94) | 0.76 (0.71, 0.81) | 4.13 (4.06, 4.19) | 7.40 (7.32, 7.48) |
| BCS | Living comfortably | 3 | 0.85 (0.80, 0.91) | 0.89 (0.84, 0.94) | 4.30 (4.24, 4.37) | 6.96 (6.88, 7.05) |
| NS | Experiencing difficulties | 1 | 1.87 (1.64, 2.09) | 1.86 (1.65, 2.08) | 5.12 (4.92, 5.33) | 6.05 (5.78, 6.32) |
| NS | Experiencing difficulties | 2 | 1.98 (1.82, 2.13) | 1.83 (1.68, 1.99) | 5.26 (5.11, 5.41) | 6.09 (5.90, 6.28) |
| NS | Experiencing difficulties | 3 | 1.89 (1.75, 2.03) | 1.86 (1.72, 2.00) | 5.20 (5.07, 5.33) | 5.95 (5.77, 6.12) |
| NS | Doing all right | 1 | 1.30 (1.18, 1.41) | 1.33 (1.22, 1.44) | 4.59 (4.48, 4.70) | 6.86 (6.72, 7.00) |
| NS | Doing all right | 2 | 1.46 (1.36, 1.55) | 1.18 (1.09, 1.26) | 4.57 (4.48, 4.65) | 7.03 (6.92, 7.14) |
| NS | Doing all right | 3 | 1.44 (1.35, 1.52) | 1.32 (1.24, 1.40) | 4.72 (4.63, 4.80) | 6.74 (6.64, 6.84) |
| NS | Living comfortably | 1 | 1.15 (1.04, 1.25) | 1.13 (1.03, 1.23) | 4.34 (4.24, 4.44) | 7.34 (7.21, 7.47) |
| NS | Living comfortably | 2 | 1.27 (1.18, 1.36) | 0.98 (0.90, 1.05) | 4.28 (4.19, 4.36) | 7.45 (7.34, 7.55) |
| NS | Living comfortably | 3 | 1.30 (1.21, 1.39) | 1.23 (1.15, 1.31) | 4.60 (4.51, 4.68) | 6.88 (6.77, 6.99) |
| MCS | Experiencing difficulties | 1 | 2.09 (1.89, 2.29) | 2.28 (2.09, 2.47) | 5.26 (5.08, 5.44) | 5.70 (5.46, 5.95) |
| MCS | Experiencing difficulties | 2 | 2.37 (2.19, 2.56) | 2.19 (2.01, 2.36) | 5.28 (5.11, 5.46) | 5.92 (5.70, 6.13) |
| MCS | Experiencing difficulties | 3 | 2.39 (2.23, 2.54) | 2.34 (2.20, 2.49) | 5.40 (5.24, 5.55) | 5.67 (5.48, 5.86) |
| MCS | Doing all right | 1 | 1.71 (1.60, 1.82) | 1.82 (1.71, 1.92) | 4.87 (4.76, 4.98) | 6.48 (6.34, 6.62) |
| MCS | Doing all right | 2 | 2.02 (1.91, 2.12) | 1.64 (1.54, 1.75) | 4.95 (4.84, 5.05) | 6.70 (6.57, 6.82) |
| MCS | Doing all right | 3 | 2.12 (2.02, 2.21) | 1.98 (1.89, 2.07) | 5.03 (4.94, 5.13) | 6.18 (6.06, 6.29) |
| MCS | Living comfortably | 1 | 1.50 (1.39, 1.62) | 1.60 (1.49, 1.71) | 4.60 (4.48, 4.73) | 6.99 (6.84, 7.14) |
| MCS | Living comfortably | 2 | 1.84 (1.72, 1.95) | 1.38 (1.28, 1.49) | 4.69 (4.57, 4.81) | 7.13 (7.00, 7.27) |
| MCS | Living comfortably | 3 | 1.78 (1.67, 1.88) | 1.68 (1.58, 1.78) | 4.67 (4.57, 4.78) | 6.69 (6.56, 6.81) |

*Note.* Adjusted models included birth sex, highest qualification achieved, pre-pandemic self-reported health, pre-pandemic psychological distress, and household composition as covariates. BCS: British Cohort Study, 1970 birth cohort; GAD-2: 2-item General Anxiety Disorder questionnaire; MCS: Millennium Cohort Study, 2000 birth cohort; NCDS: National Child and Development Study, 1958 birth cohort; NS: Next Steps, 1990 cohort; NSHD: National Survey of Health and Development, 1946 birth cohort; ONS: UK Office for National Statistics; PHQ-2: 2-item Patient Health Questionnaire; UCLA-3: 3-item UCLA loneliness scale. Survey wave 1: May 2020; survey wave 2: September/October 2020; survey wave 3: February/March 2021.

#### Table S7.3. Difference-in-differences (change in difference between people in worse vs best pre-pandemic financial situation between first vs last time point) estimates and 95% confidence intervals.

|  | *DID* (95% CI) | *p* |
| --- | --- | --- |
| GAD-2 | 0.08 (-0.10, 0.27) | 0.387 |
| PHQ-2 | 0.09 (-0.05, 0.22) | 0.197 |
| UCLA-3 | 0.07 (-0.07, 0.21) | 0.320 |
| Life satisfaction | -0.21 (-0.54, 0.12) | 0.212 |

*Note*. DID: difference-in-differences; GAD-2: 2-item General Anxiety Disorder questionnaire; PHQ-2: 2-item Patient Health Questionnaire; UCLA-3: 3-item UCLA loneliness scale.

#### Figure S7.1. Unadjusted and adjusted (by birth sex, highest qualification achieved, pre-pandemic self-reported health, pre-pandemic psychological distress, and household composition) anxiety symptomatology (GAD-2) marginal mean estimates and 95% confidence intervals by pre-pandemic financial situation.

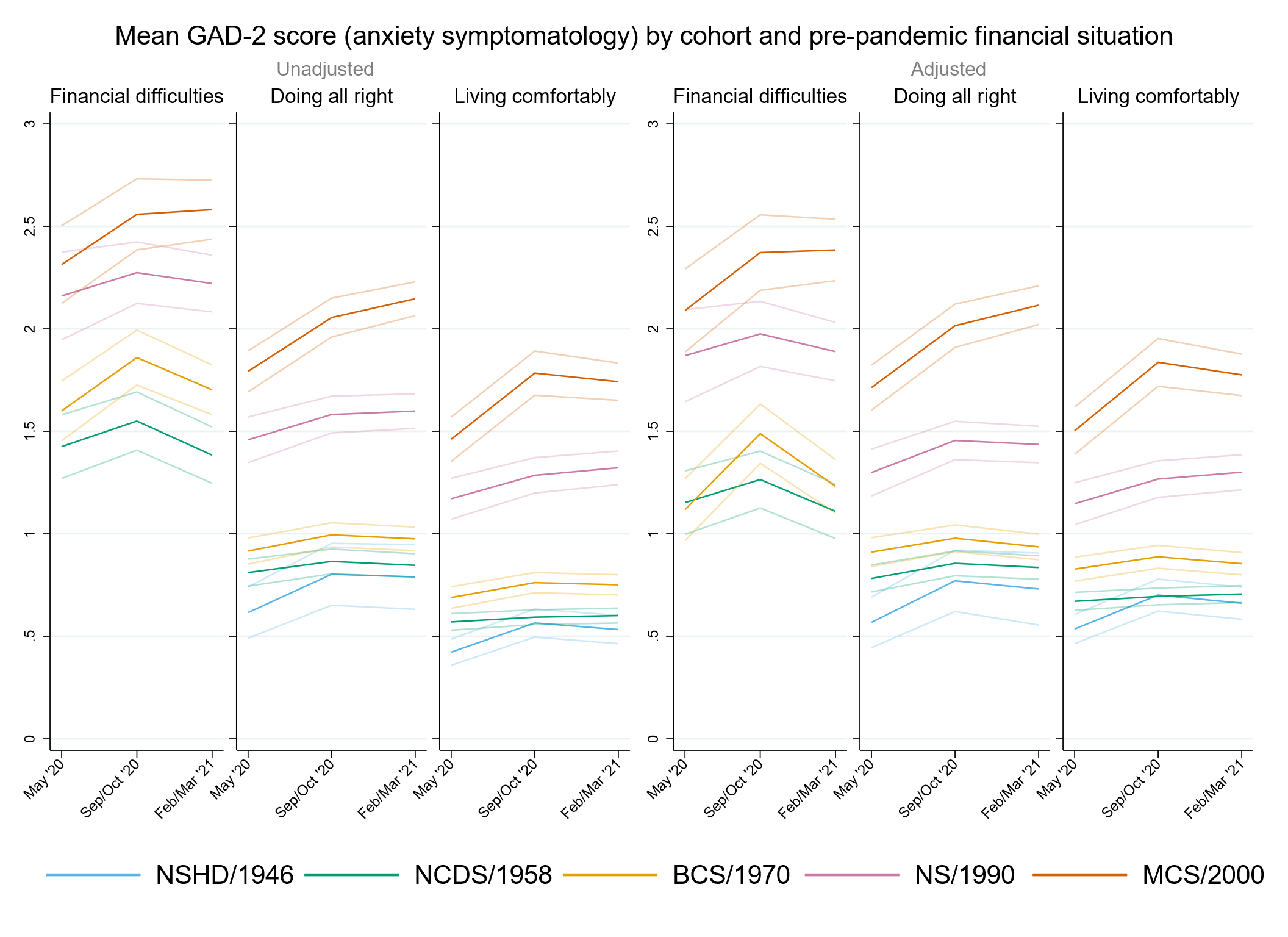

#### Figure S7.2. Unadjusted and adjusted (by birth sex, highest qualification achieved, pre-pandemic self-reported health, pre-pandemic psychological distress, and household composition) depressive symptomatology (PHQ-2) marginal mean estimates and 95% confidence intervals by pre-pandemic financial situation.

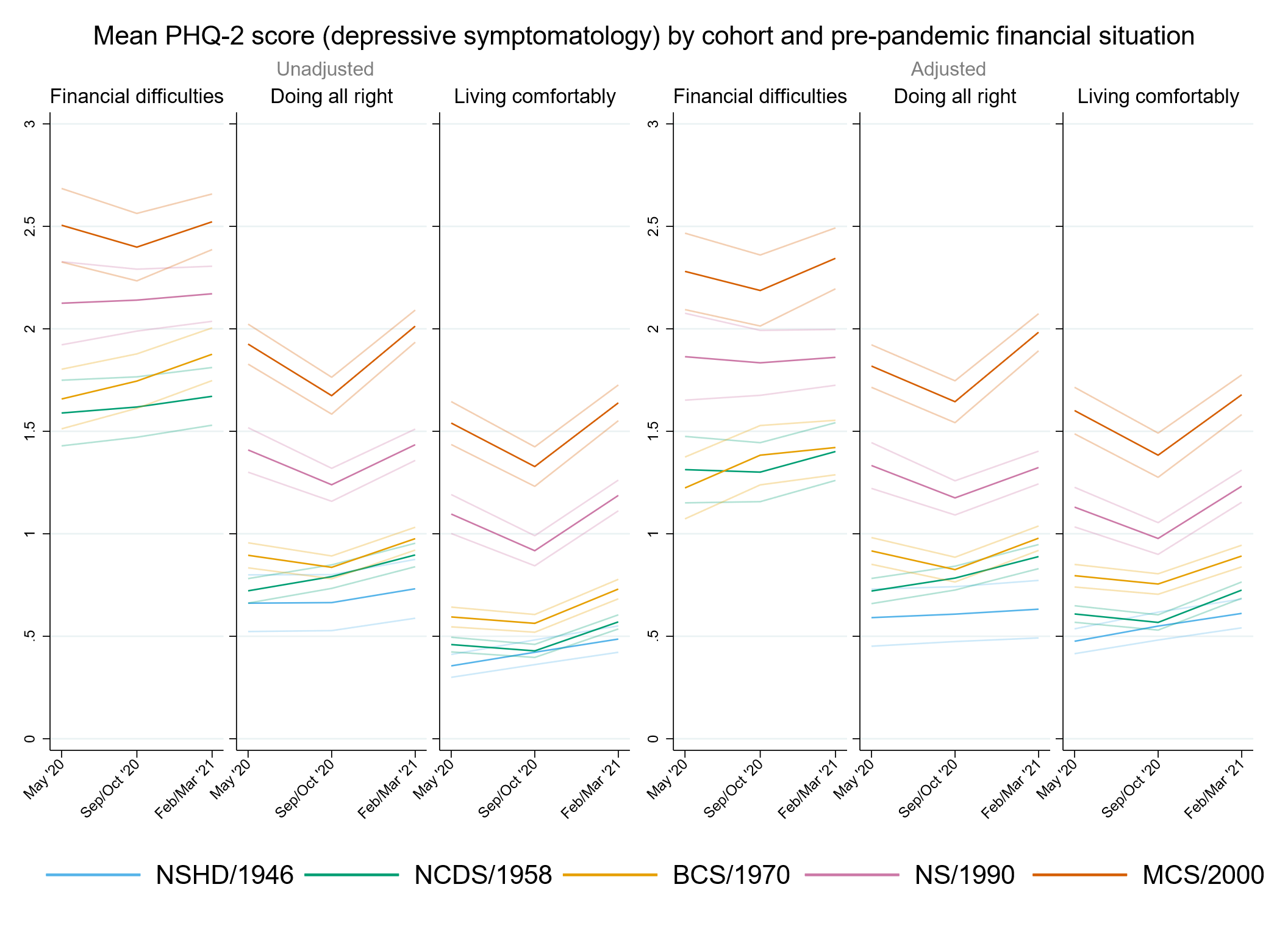

#### Figure S7.3. Unadjusted and adjusted (by birth sex, highest qualification achieved, pre-pandemic self-reported health, pre-pandemic psychological distress, and household composition) loneliness (UCLA-3) marginal mean estimates and 95% confidence intervals by pre-pandemic financial situation.

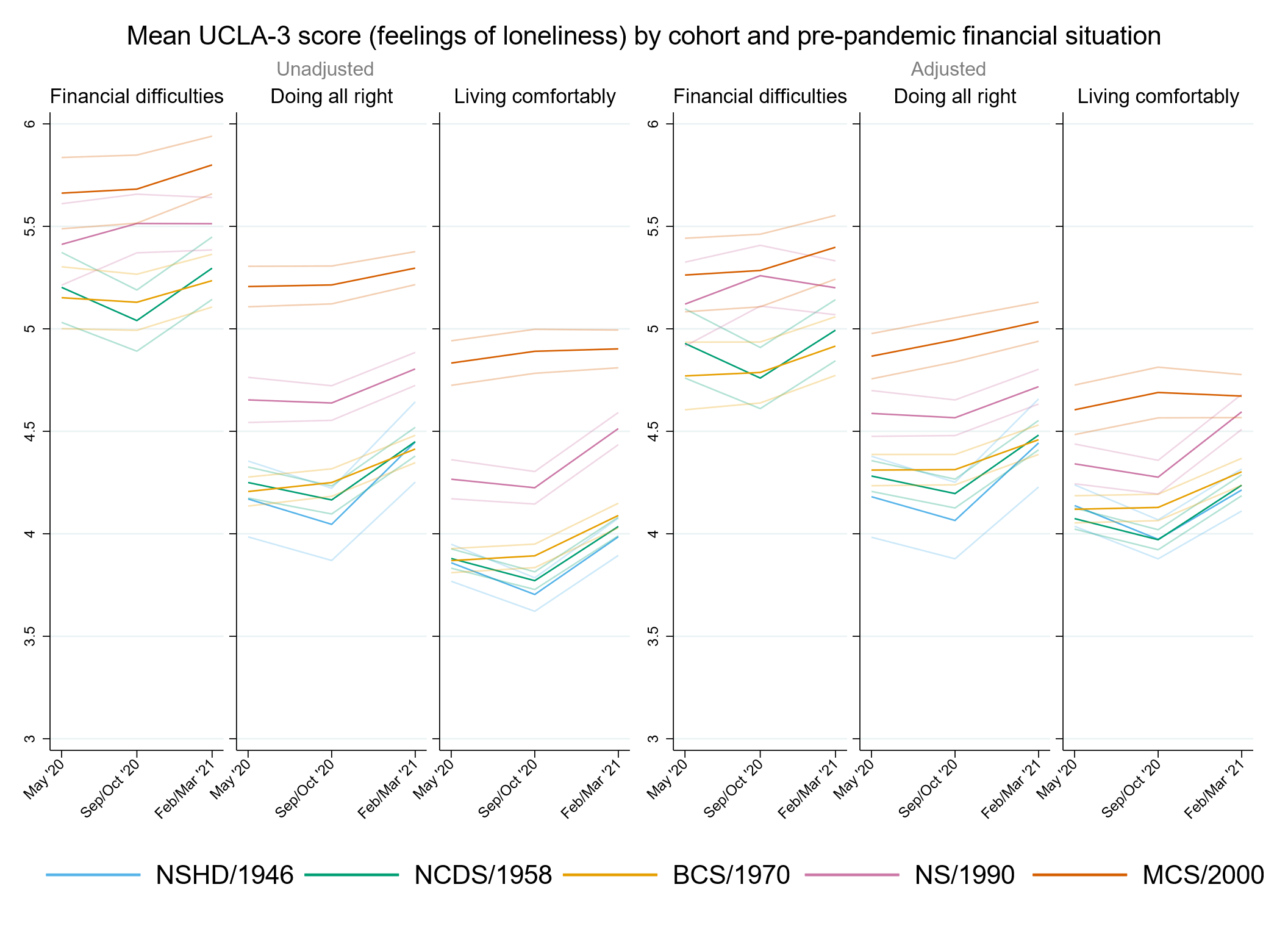

#### Figure S7.4. Unadjusted and adjusted (by birth sex, highest qualification achieved, pre-pandemic self-reported health, pre-pandemic psychological distress, and household composition) life satisfaction marginal mean estimates and 95% confidence intervals by pre-pandemic financial situation.

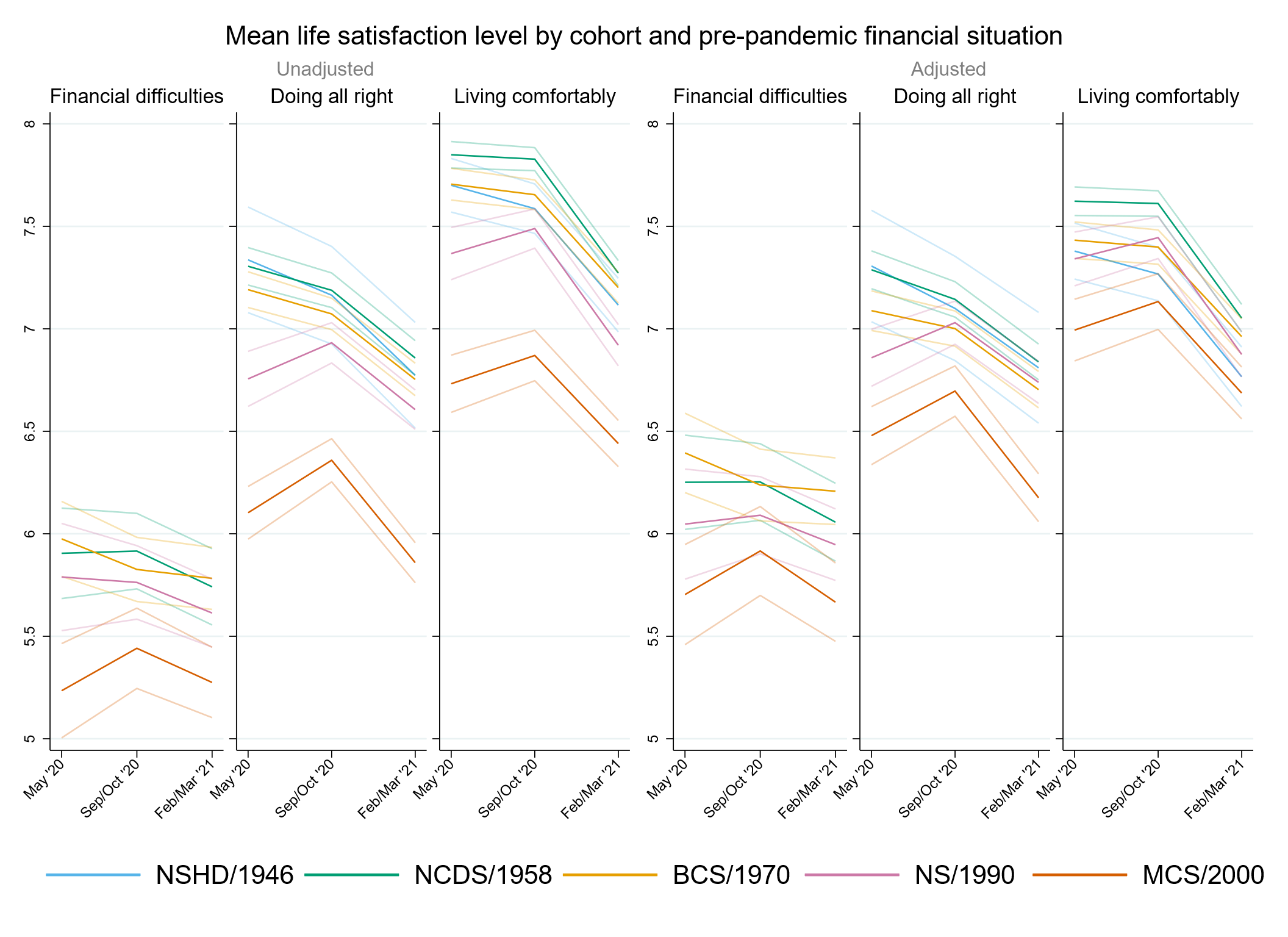

### Appendix S8. Results by relationship status.

#### Table S8.1. Results of multilevel growth curve models by relationship status.

|  | **Anxiety symptomatology (GAD-2)** | | | **Depressive symptomatology (PHQ-2)** | | | **Feelings of loneliness (UCLA-3)** | | | **Life satisfaction (ONS single question)** | | |
| --- | --- | --- | --- | --- | --- | --- | --- | --- | --- | --- | --- | --- |
| **Unadjusted models** |  | | |  | | |  | | |  | | |
| N participants | 24,547 |  |  | 24,545 |  |  | 24,574 |  |  | 24,628 |  |  |
| N observations | 52,346 |  |  | 59,329 |  |  | 52,411 |  |  | 52,636 |  |  |
|  | ***B* (95% CI)** | **χ2** | ***p*** | ***B* (95% CI)** | **χ2** | ***p*** | ***B* (95% CI)** | **χ2** | ***p*** | ***B* (95% CI)** | **χ2** | ***p*** |
| Time (linear) | 0.03 (-0.14, 0.20) |  | 0.753 | -0.01 (-0.18, 0.16) |  | 0.915 | -0.38 (-0.60, -0.17) |  | <0.001 | 0.01 (-0.23, 0.24) |  | 0.955 |
| Time (quadratic) | -0.02 (-0.10, 0.06) |  | 0.647 | 0.03 (-0.05, 0.11) |  | 0.477 | 0.21 (0.12, 0.31) |  | <0.001 | -0.10 (-0.21, 0.01) |  | 0.087 |
| Cohort (ref. NCDS) |  | 124 | <0.001 |  | 188.7 | <0.001 |  | 33.7 | <0.001 |  | 46.1 | <0.001 |
| NSHD | -0.39 (-0.76, -0.03) |  | 0.036 | -0.66 (-1.07, -0.25) |  | 0.002 | -0.59 (-1.27, 0.09) |  | 0.092 | 1.13 (0.18, 2.09) |  | 0.019 |
| BCS | 0.28 (0.11, 0.45) |  | 0.001 | 0.34 (0.16, 0.51) |  | <0.001 | 0.23 (0.02, 0.43) |  | 0.028 | -0.27 (-0.50, -0.04) |  | 0.024 |
| NS | 0.82 (0.62, 1.02) |  | <0.001 | 0.89 (0.69, 1.09) |  | <0.001 | 0.56 (0.35, 0.76) |  | <0.001 | -0.66 (-0.93, -0.40) |  | <0.001 |
| MCS | 0.62 (0.48, 0.75) |  | <0.001 | 0.81 (0.67, 0.95) |  | <0.001 | 0.24 (0.07, 0.40) |  | 0.005 | -0.52 (-0.72, -0.33) |  | <0.001 |
| Linear change * cohort (ref. NCDS) |  | 14.9 | 0.005 |  | 10.1 | 0.038 |  | 10.9 | 0.028 |  | 22.2 | <0.001 |
| NSHD | 0.13 (-0.37, 0.63) |  | 0.610 | 0.35 (-0.52, 1.23) |  | 0.430 | -0.52 (-1.65, 0.60) |  | 0.360 | -1.43 (-3.59, 0.74) |  | 0.196 |
| BCS | 0.20 (-0.10, 0.49) |  | 0.190 | -0.11 (-0.39, 0.17) |  | 0.435 | 0.27 (-0.07, 0.60) |  | 0.118 | -0.35 (-0.71, 0.01) |  | 0.059 |
| NS | 0.07 (-0.27, 0.40) |  | 0.691 | -0.24 (-0.58, 0.10) |  | 0.163 | 0.32 (-0.01, 0.65) |  | 0.057 | 0.26 (-0.15, 0.68) |  | 0.217 |
| MCS | 0.46 (0.22, 0.70) |  | <0.001 | -0.36 (-0.60, -0.11) |  | 0.004 | 0.41 (0.14, 0.68) |  | 0.003 | 0.42 (0.10, 0.74) |  | 0.010 |
| Quadratic change * cohort (ref. NCDS) |  | 5.9 | 0.207 |  | 9.4 | 0.052 |  | 18.1 | 0.001 |  | 19.7 | <0.001 |
| NSHD | -0.05 (-0.31, 0.21) |  | 0.730 | -0.05 (-0.48, 0.37) |  | 0.800 | 0.38 (-0.12, 0.88) |  | 0.139 | 0.46 (-0.54, 1.45) |  | 0.368 |
| BCS | -0.08 (-0.22, 0.06) |  | 0.258 | 0.06 (-0.06, 0.19) |  | 0.328 | -0.14 (-0.29, 0.02) |  | 0.084 | 0.20 (0.02, 0.37) |  | 0.026 |
| NS | -0.04 (-0.19, 0.11) |  | 0.603 | 0.06 (-0.09, 0.22) |  | 0.407 | -0.21 (-0.36, -0.06) |  | 0.007 | -0.05 (-0.24, 0.14) |  | 0.595 |
| MCS | -0.14 (-0.25, -0.02) |  | 0.017 | 0.17 (0.06, 0.29) |  | 0.003 | -0.23 (-0.35, -0.10) |  | <0.001 | -0.16 (-0.31, -0.01) |  | 0.037 |
| Relationship status (ref. Not in a relationship) | -0.30 (-0.41, -0.19) |  | <0.001 | -0.42 (-0.54, -0.31) |  | <0.001 | -1.12 (-1.26, -0.98) |  | <0.001 | 1.06 (0.89, 1.23) |  | <0.001 |
| Linear change *  relationship status (ref. Not in a relationship) | 0.05 (-0.13, 0.24) |  | 0.569 | -0.04 (-0.22, 0.14) |  | 0.649 | 0.11 (-0.11, 0.33) |  | 0.320 | 0.15 (-0.10, 0.41) |  | 0.231 |
| Quadratic change *  relationship status (ref. Not in a relationship) | -0.01 (-0.10, 0.07) |  | 0.733 | 0.03 (-0.06, 0.11) |  | 0.519 | -0.04 (-0.14, 0.06) |  | 0.461 | -0.11 (-0.23, 0.01) |  | 0.077 |
| Cohort (ref. NCDS) *  relationship status (ref. Not in a relationship) |  | 88.2 | <0.001 |  | 78 | <0.001 |  | 112.4 | <0.001 |  | 61.1 | <0.001 |
| NSHD | 0.18 (-0.19, 0.55) |  | 0.343 | 0.46 (0.05, 0.88) |  | 0.029 | 0.46 (-0.23, 1.15) |  | 0.189 | -1.08 (-2.04, -0.12) |  | 0.028 |
| BCS | -0.09 (-0.27, 0.09) |  | 0.317 | -0.11 (-0.29, 0.07) |  | 0.224 | -0.15 (-0.37, 0.06) |  | 0.157 | 0.00 (-0.25, 0.24) |  | 0.970 |
| NS | -0.13 (-0.35, 0.09) |  | 0.241 | -0.21 (-0.42, 0.01) |  | 0.061 | -0.16 (-0.39, 0.06) |  | 0.155 | 0.11 (-0.17, 0.40) |  | 0.448 |
| MCS | 0.72 (0.54, 0.90) |  | <0.001 | 0.60 (0.43, 0.78) |  | <0.001 | 0.77 (0.58, 0.97) |  | <0.001 | -0.79 (-1.04, -0.54) |  | <0.001 |
| Linear change * cohort (ref. NCDS) *  relationship status (ref. Not in a relationship) |  | 1.9 | 0.751 |  | 1.8 | 0.765 |  | 1.7 | 0.783 |  | 4.1 | 0.397 |
| NSHD | -0.03 (-0.55, 0.48) |  | 0.900 | -0.30 (-1.18, 0.58) |  | 0.508 | 0.44 (-0.69, 1.57) |  | 0.444 | 1.43 (-0.75, 3.61) |  | 0.197 |
| BCS | -0.10 (-0.42, 0.21) |  | 0.514 | 0.05 (-0.24, 0.35) |  | 0.714 | -0.02 (-0.37, 0.33) |  | 0.913 | 0.26 (-0.12, 0.65) |  | 0.181 |
| NS | 0.07 (-0.30, 0.43) |  | 0.721 | -0.03 (-0.40, 0.33) |  | 0.867 | -0.14 (-0.50, 0.22) |  | 0.456 | -0.06 (-0.52, 0.39) |  | 0.794 |
| MCS | -0.19 (-0.51, 0.14) |  | 0.271 | -0.16 (-0.48, 0.17) |  | 0.339 | -0.14 (-0.48, 0.21) |  | 0.443 | 0.01 (-0.41, 0.44) |  | 0.945 |
| Quadratic change * cohort (ref. NCDS) *  relationship status (ref. Not in a relationship) |  | 0.8 | 0.937 |  | 2.6 | 0.633 |  | 6 | 0.198 |  | 3.5 | 0.481 |
| NSHD | 0.02 (-0.25, 0.28) |  | 0.908 | 0.02 (-0.40, 0.45) |  | 0.921 | -0.34 (-0.84, 0.16) |  | 0.187 | -0.48 (-1.48, 0.52) |  | 0.345 |
| BCS | 0.04 (-0.10, 0.19) |  | 0.567 | -0.04 (-0.17, 0.10) |  | 0.601 | 0.02 (-0.14, 0.19) |  | 0.773 | -0.14 (-0.32, 0.05) |  | 0.145 |
| NS | 0.02 (-0.15, 0.18) |  | 0.860 | 0.07 (-0.09, 0.24) |  | 0.400 | 0.14 (-0.03, 0.30) |  | 0.098 | -0.01 (-0.21, 0.20) |  | 0.947 |
| MCS | 0.06 (-0.09, 0.22) |  | 0.409 | 0.08 (-0.07, 0.23) |  | 0.312 | 0.10 (-0.06, 0.26) |  | 0.205 | 0.00 (-0.20, 0.20) |  | 1.000 |
| **Adjusted models** |  | | |  | | |  | | |  | | |
| N participants | 20,955 |  |  | 20,956 |  |  | 20,981 |  |  | 21,023 |  |  |
| N observations | 45,644 |  |  | 45,635 |  |  | 45,696 |  |  | 45,888 |  |  |
|  | ***B* (95% CI)** | **χ2** | ***p*** | ***B* (95% CI)** | **χ2** | ***p*** | ***B* (95% CI)** | **χ2** | ***p*** | ***B* (95% CI)** | **χ2** | ***p*** |
| Time (linear) | 0.11 (-0.07, 0.30) |  | 0.234 | -0.06 (-0.24, 0.13) |  | 0.556 | -0.31 (-0.54, -0.09) |  | 0.007 | 0.05 (-0.21, 0.31) |  | 0.682 |
| Time (quadratic) | -0.06 (-0.15, 0.03) |  | 0.166 | 0.05 (-0.04, 0.14) |  | 0.265 | 0.19 (0.08, 0.29) |  | <0.001 | -0.13 (-0.26, -0.01) |  | 0.037 |
| Cohort (ref. NCDS) |  | 140.5 | <0.001 |  | 219 | <0.001 |  | 47.5 | <0.001 |  | 53.3 | <0.001 |
| NSHD | -0.44 (-0.89, 0.01) |  | 0.056 | -0.69 (-1.12, -0.25) |  | 0.002 | -0.15 (-0.98, 0.68) |  | 0.723 | 0.73 (-0.07, 1.52) |  | 0.073 |
| BCS | 0.17 (0.00, 0.35) |  | 0.056 | 0.13 (-0.04, 0.31) |  | 0.141 | 0.03 (-0.18, 0.24) |  | 0.797 | 0.00 (-0.24, 0.23) |  | 0.977 |
| NS | 0.74 (0.53, 0.95) |  | <0.001 | 0.82 (0.61, 1.03) |  | <0.001 | 0.58 (0.37, 0.80) |  | <0.001 | -0.56 (-0.84, -0.28) |  | <0.001 |
| MCS | 0.77 (0.62, 0.92) |  | <0.001 | 0.92 (0.77, 1.08) |  | <0.001 | 0.43 (0.26, 0.61) |  | <0.001 | -0.58 (-0.79, -0.37) |  | <0.001 |
| Linear change * cohort (ref. NCDS) |  | 12.6 | 0.013 |  | 3.6 | 0.469 |  | 10.6 | 0.032 |  | 16.1 | 0.003 |
| NSHD | 0.16 (-0.60, 0.93) |  | 0.674 | 0.39 (-0.66, 1.44) |  | 0.466 | -0.35 (-1.71, 1.01) |  | 0.616 | -2.31 (-4.46, -0.15) |  | 0.036 |
| BCS | 0.04 (-0.31, 0.40) |  | 0.806 | -0.04 (-0.35, 0.26) |  | 0.777 | 0.25 (-0.10, 0.61) |  | 0.162 | -0.35 (-0.74, 0.05) |  | 0.089 |
| NS | 0.09 (-0.28, 0.47) |  | 0.623 | -0.15 (-0.52, 0.22) |  | 0.427 | 0.31 (-0.05, 0.67) |  | 0.094 | 0.11 (-0.36, 0.57) |  | 0.653 |
| MCS | 0.45 (0.18, 0.71) |  | 0.001 | -0.21 (-0.48, 0.06) |  | 0.120 | 0.47 (0.17, 0.77) |  | 0.002 | 0.30 (-0.05, 0.65) |  | 0.089 |
| Quadratic change * cohort (ref. NCDS) |  | 4.1 | 0.393 |  | 3.9 | 0.422 |  | 14.9 | 0.005 |  | 15 | 0.005 |
| NSHD | -0.05 (-0.50, 0.41) |  | 0.833 | -0.09 (-0.61, 0.42) |  | 0.721 | 0.13 (-0.45, 0.70) |  | 0.664 | 1.03 (0.05, 2.01) |  | 0.040 |
| BCS | -0.02 (-0.19, 0.14) |  | 0.774 | 0.04 (-0.10, 0.18) |  | 0.589 | -0.13 (-0.30, 0.03) |  | 0.112 | 0.19 (0.00, 0.38) |  | 0.054 |
| NS | -0.05 (-0.22, 0.12) |  | 0.548 | 0.01 (-0.16, 0.17) |  | 0.928 | -0.21 (-0.38, -0.04) |  | 0.013 | 0.05 (-0.16, 0.26) |  | 0.651 |
| MCS | -0.12 (-0.24, 0.00) |  | 0.049 | 0.11 (-0.01, 0.24) |  | 0.073 | -0.25 (-0.39, -0.12) |  | <0.001 | -0.11 (-0.27, 0.06) |  | 0.205 |
| Relationship status (ref. Not in a relationship) | -0.02 (-0.14, 0.10) |  | 0.739 | -0.21 (-0.33, -0.08) |  | 0.001 | -0.77 (-0.92, -0.61) |  | <0.001 | 0.65 (0.47, 0.83) |  | <0.001 |
| Linear change *  relationship status (ref. Not in a relationship) | -0.03 (-0.23, 0.17) |  | 0.758 | -0.01 (-0.20, 0.19) |  | 0.958 | 0.05 (-0.18, 0.29) |  | 0.669 | 0.08 (-0.19, 0.36) |  | 0.554 |
| Quadratic change *  relationship status (ref. Not in a relationship) | 0.03 (-0.06, 0.12) |  | 0.535 | 0.01 (-0.08, 0.10) |  | 0.784 | -0.02 (-0.13, 0.09) |  | 0.767 | -0.06 (-0.19, 0.07) |  | 0.374 |
| Cohort (ref. NCDS) *  relationship status (ref. Not in a relationship) |  | 22.4 | <0.001 |  | 27.4 | <0.001 |  | 32.3 | <0.001 |  | 16.3 | 0.003 |
| NSHD | 0.30 (-0.16, 0.76) |  | 0.200 | 0.53 (0.09, 0.98) |  | 0.018 | 0.10 (-0.73, 0.93) |  | 0.819 | -0.79 (-1.60, 0.01) |  | 0.054 |
| BCS | -0.04 (-0.22, 0.15) |  | 0.706 | 0.06 (-0.12, 0.24) |  | 0.519 | 0.03 (-0.19, 0.25) |  | 0.820 | -0.24 (-0.50, 0.01) |  | 0.062 |
| NS | -0.22 (-0.45, 0.01) |  | 0.056 | -0.26 (-0.49, -0.04) |  | 0.022 | -0.31 (-0.54, -0.09) |  | 0.007 | 0.16 (-0.14, 0.46) |  | 0.285 |
| MCS | 0.31 (0.13, 0.50) |  | 0.001 | 0.30 (0.11, 0.48) |  | 0.002 | 0.35 (0.14, 0.55) |  | 0.001 | -0.33 (-0.59, -0.08) |  | 0.011 |
| Linear change * cohort (ref. NCDS) *  relationship status (ref. Not in a relationship) |  | 1.7 | 0.794 |  | 3.3 | 0.516 |  | 1.7 | 0.787 |  | 6.2 | 0.183 |
| NSHD | -0.04 (-0.82, 0.73) |  | 0.914 | -0.31 (-1.37, 0.75) |  | 0.568 | 0.20 (-1.17, 1.57) |  | 0.770 | 2.35 (0.17, 4.52) |  | 0.034 |
| BCS | 0.08 (-0.29, 0.45) |  | 0.679 | -0.01 (-0.33, 0.31) |  | 0.964 | -0.05 (-0.42, 0.32) |  | 0.809 | 0.32 (-0.11, 0.74) |  | 0.147 |
| NS | 0.05 (-0.35, 0.46) |  | 0.791 | -0.08 (-0.48, 0.32) |  | 0.703 | -0.14 (-0.53, 0.25) |  | 0.480 | 0.13 (-0.38, 0.63) |  | 0.628 |
| MCS | -0.18 (-0.53, 0.18) |  | 0.329 | -0.30 (-0.65, 0.05) |  | 0.097 | -0.22 (-0.60, 0.15) |  | 0.246 | 0.22 (-0.24, 0.67) |  | 0.359 |
| Quadratic change * cohort (ref. NCDS) *  relationship status (ref. Not in a relationship) |  | 0.8 | 0.94 |  | 4.6 | 0.327 |  | 4.9 | 0.298 |  | 6.3 | 0.175 |
| NSHD | 0.01 (-0.45, 0.48) |  | 0.950 | 0.05 (-0.47, 0.57) |  | 0.849 | -0.06 (-0.64, 0.52) |  | 0.833 | -1.08 (-2.07, -0.09) |  | 0.033 |
| BCS | -0.03 (-0.21, 0.14) |  | 0.705 | -0.02 (-0.17, 0.13) |  | 0.753 | 0.03 (-0.14, 0.20) |  | 0.721 | -0.14 (-0.35, 0.06) |  | 0.166 |
| NS | 0.02 (-0.17, 0.20) |  | 0.873 | 0.10 (-0.08, 0.28) |  | 0.263 | 0.15 (-0.03, 0.33) |  | 0.094 | -0.13 (-0.36, 0.10) |  | 0.272 |
| MCS | 0.05 (-0.11, 0.22) |  | 0.513 | 0.14 (-0.02, 0.31) |  | 0.089 | 0.16 (-0.02, 0.33) |  | 0.084 | -0.11 (-0.32, 0.10) |  | 0.318 |
| **Sensitivity models** |  | | |  | | |  | | |  | | |
| N participants | 20,955 |  |  | 20,956 |  |  | 20,981 |  |  | 21,023 |  |  |
| N observations | 45,644 |  |  | 45,635 |  |  | 45,696 |  |  | 45,888 |  |  |
|  | ***B* (95% CI)** | **χ2** | ***p*** | ***B* (95% CI)** | **χ2** | ***p*** | ***B* (95% CI)** | **χ2** | ***p*** | ***B* (95% CI)** | **χ2** | ***p*** |
| Time (linear) | 0.12 (-0.07, 0.31) |  | 0.225 | -0.06 (-0.25, 0.13) |  | 0.558 | -0.32 (-0.55, -0.09) |  | 0.006 | 0.04 (-0.22, 0.31) |  | 0.743 |
| Time (quadratic) | -0.06 (-0.15, 0.03) |  | 0.175 | 0.05 (-0.04, 0.14) |  | 0.259 | 0.19 (0.08, 0.29) |  | <0.001 | -0.13 (-0.26, -0.01) |  | 0.041 |
| Cohort (ref. NCDS) |  | 106.9 | <0.001 |  | 154.7 | <0.001 |  | 32 | <0.001 |  | 41.3 | <0.001 |
| NSHD | -0.24 (-0.69, 0.22) |  | 0.308 | -0.51 (-0.93, -0.08) |  | 0.020 | -0.05 (-0.88, 0.77) |  | 0.900 | 0.52 (-0.34, 1.39) |  | 0.232 |
| BCS | 0.23 (0.04, 0.41) |  | 0.019 | 0.17 (-0.02, 0.36) |  | 0.084 | 0.04 (-0.18, 0.26) |  | 0.728 | -0.07 (-0.32, 0.18) |  | 0.583 |
| NS | 0.83 (0.61, 1.05) |  | <0.001 | 0.88 (0.66, 1.10) |  | <0.001 | 0.59 (0.37, 0.81) |  | <0.001 | -0.68 (-0.96, -0.40) |  | <0.001 |
| MCS | 0.64 (0.49, 0.78) |  | <0.001 | 0.76 (0.61, 0.91) |  | <0.001 | 0.22 (0.05, 0.40) |  | 0.013 | -0.49 (-0.70, -0.28) |  | <0.001 |
| Linear change * cohort (ref. NCDS) |  | 11 | 0.027 |  | 3.6 | 0.465 |  | 10.2 | 0.037 |  | 16.4 | 0.003 |
| NSHD | 0.16 (-0.63, 0.95) |  | 0.695 | 0.35 (-0.73, 1.42) |  | 0.527 | -0.36 (-1.72, 1.00) |  | 0.602 | -2.26 (-4.42, -0.09) |  | 0.041 |
| BCS | 0.07 (-0.29, 0.43) |  | 0.690 | -0.03 (-0.34, 0.28) |  | 0.852 | 0.26 (-0.09, 0.62) |  | 0.150 | -0.35 (-0.75, 0.05) |  | 0.087 |
| NS | 0.10 (-0.28, 0.48) |  | 0.602 | -0.15 (-0.52, 0.23) |  | 0.440 | 0.30 (-0.06, 0.67) |  | 0.103 | 0.12 (-0.34, 0.59) |  | 0.608 |
| MCS | 0.43 (0.16, 0.70) |  | 0.002 | -0.22 (-0.49, 0.05) |  | 0.113 | 0.47 (0.17, 0.77) |  | 0.002 | 0.32 (-0.03, 0.67) |  | 0.073 |
| Quadratic change * cohort (ref. NCDS) |  | 4.1 | 0.396 |  | 3.4 | 0.489 |  | 15 | 0.005 |  | 14.6 | 0.006 |
| NSHD | -0.05 (-0.52, 0.42) |  | 0.822 | -0.08 (-0.61, 0.45) |  | 0.769 | 0.13 (-0.45, 0.70) |  | 0.667 | 1.02 (0.03, 2.00) |  | 0.042 |
| BCS | -0.04 (-0.21, 0.13) |  | 0.658 | 0.03 (-0.11, 0.18) |  | 0.661 | -0.14 (-0.30, 0.03) |  | 0.101 | 0.19 (0.00, 0.39) |  | 0.050 |
| NS | -0.05 (-0.22, 0.12) |  | 0.547 | 0.01 (-0.16, 0.17) |  | 0.943 | -0.21 (-0.38, -0.04) |  | 0.014 | 0.04 (-0.17, 0.25) |  | 0.693 |
| MCS | -0.13 (-0.25, 0.00) |  | 0.046 | 0.11 (-0.02, 0.24) |  | 0.092 | -0.26 (-0.40, -0.12) |  | <0.001 | -0.10 (-0.27, 0.06) |  | 0.223 |
| Relationship status (ref. Not in a relationship) | -0.29 (-0.41, -0.17) |  | <0.001 | -0.45 (-0.58, -0.33) |  | <0.001 | -1.12 (-1.27, -0.97) |  | <0.001 | 1.08 (0.90, 1.26) |  | <0.001 |
| Linear change *  relationship status (ref. Not in a relationship) | -0.03 (-0.23, 0.17) |  | 0.777 | 0.00 (-0.20, 0.20) |  | 1.000 | 0.06 (-0.18, 0.30) |  | 0.618 | 0.09 (-0.19, 0.36) |  | 0.550 |
| Quadratic change *  relationship status (ref. Not in a relationship) | 0.03 (-0.07, 0.12) |  | 0.571 | 0.01 (-0.08, 0.10) |  | 0.832 | -0.02 (-0.13, 0.09) |  | 0.729 | -0.06 (-0.19, 0.07) |  | 0.376 |
| Cohort (ref. NCDS) *  relationship status (ref. Not in a relationship) |  | 67.7 | <0.001 |  | 57.9 | <0.001 |  | 83.9 | <0.001 |  | 44.3 | <0.001 |
| NSHD | 0.03 (-0.43, 0.49) |  | 0.887 | 0.32 (-0.11, 0.75) |  | 0.148 | -0.07 (-0.90, 0.76) |  | 0.861 | -0.48 (-1.35, 0.40) |  | 0.283 |
| BCS | -0.03 (-0.23, 0.17) |  | 0.745 | 0.06 (-0.14, 0.26) |  | 0.539 | 0.04 (-0.19, 0.27) |  | 0.754 | -0.22 (-0.48, 0.05) |  | 0.112 |
| NS | -0.15 (-0.38, 0.09) |  | 0.226 | -0.21 (-0.45, 0.02) |  | 0.074 | -0.21 (-0.45, 0.03) |  | 0.082 | 0.15 (-0.16, 0.45) |  | 0.351 |
| MCS | 0.70 (0.50, 0.89) |  | <0.001 | 0.61 (0.42, 0.80) |  | <0.001 | 0.75 (0.54, 0.96) |  | <0.001 | -0.76 (-1.02, -0.49) |  | <0.001 |
| Linear change * cohort (ref. NCDS) *  relationship status (ref. Not in a relationship) |  | 1 | 0.908 |  | 2.6 | 0.632 |  | 1.4 | 0.844 |  | 6.3 | 0.178 |
| NSHD | -0.05 (-0.86, 0.76) |  | 0.901 | -0.27 (-1.35, 0.81) |  | 0.626 | 0.21 (-1.16, 1.58) |  | 0.764 | 2.31 (0.13, 4.49) |  | 0.038 |
| BCS | 0.03 (-0.34, 0.41) |  | 0.862 | -0.04 (-0.36, 0.29) |  | 0.826 | -0.07 (-0.44, 0.30) |  | 0.715 | 0.34 (-0.09, 0.77) |  | 0.119 |
| NS | 0.04 (-0.37, 0.45) |  | 0.850 | -0.08 (-0.49, 0.32) |  | 0.692 | -0.13 (-0.53, 0.26) |  | 0.505 | 0.12 (-0.39, 0.62) |  | 0.653 |
| MCS | -0.15 (-0.51, 0.21) |  | 0.407 | -0.28 (-0.65, 0.08) |  | 0.126 | -0.20 (-0.59, 0.18) |  | 0.292 | 0.17 (-0.29, 0.64) |  | 0.471 |
| Quadratic change * cohort (ref. NCDS) *  relationship status (ref. Not in a relationship) |  | 0.4 | 0.981 |  | 3.9 | 0.415 |  | 4.3 | 0.369 |  | 6.4 | 0.171 |
| NSHD | 0.02 (-0.45, 0.50) |  | 0.921 | 0.04 (-0.50, 0.57) |  | 0.894 | -0.06 (-0.64, 0.52) |  | 0.840 | -1.07 (-2.06, -0.08) |  | 0.035 |
| BCS | -0.01 (-0.19, 0.17) |  | 0.894 | -0.01 (-0.16, 0.14) |  | 0.898 | 0.04 (-0.13, 0.22) |  | 0.630 | -0.16 (-0.36, 0.05) |  | 0.130 |
| NS | 0.02 (-0.17, 0.20) |  | 0.859 | 0.10 (-0.08, 0.28) |  | 0.269 | 0.15 (-0.03, 0.33) |  | 0.106 | -0.12 (-0.35, 0.11) |  | 0.300 |
| MCS | 0.05 (-0.12, 0.21) |  | 0.576 | 0.14 (-0.03, 0.31) |  | 0.104 | 0.15 (-0.03, 0.33) |  | 0.103 | -0.09 (-0.31, 0.12) |  | 0.395 |

*Note.* Adjusted models included birth sex, highest qualification achieved, pre-pandemic self-reported health, pre-pandemic psychological distress, and household composition as covariates. Sensitivity models correspond to the unadjusted models after restricting the analytical sample to that of the adjusted models. BCS: British Cohort Study, 1970 birth cohort; GAD-2: 2-item General Anxiety Disorder questionnaire; MCS: Millennium Cohort Study, 2000 birth cohort; NCDS: National Child and Development Study, 1958 birth cohort; NS: Next Steps, 1990 cohort; NSHD: National Survey of Health and Development, 1946 birth cohort; ONS: UK Office for National Statistics; PHQ-2: 2-item Patient Health Questionnaire; UCLA-3: 3-item UCLA loneliness scale. χ2: Wald test performed to assess the overall statistical significance of the interaction terms; all χ2 statistics in this table have 4 degrees of freedom.

#### Table S8.2. Unadjusted and adjusted marginal mean estimates and 95% confidence intervals by relationship status.

|  |  |  | **Anxiety symptomatology (GAD-2)** | **Depressive symptomatology (PHQ-2)** | **Feelings of loneliness (UCLA-3)** | **Life satisfaction (ONS single question)** |
| --- | --- | --- | --- | --- | --- | --- |
| **Cohort** | **Relationship status** | **Survey wave** | **Unadjusted marginal mean (95% CI)** | **Unadjusted marginal mean (95% CI)** | **Unadjusted marginal mean (95% CI)** | **Unadjusted marginal mean (95% CI)** |
| NSHD | Not in a relationship | 1 | 0.59 (0.24, 0.94) | 0.35 (-0.05, 0.74) | 4.47 (3.80, 5.13) | 7.73 (6.79, 8.67) |
| NSHD | Not in a relationship | 2 | 0.69 (0.34, 1.03) | 0.66 (0.30, 1.02) | 4.15 (3.60, 4.70) | 6.67 (5.87, 7.46) |
| NSHD | Not in a relationship | 3 | 0.65 (0.30, 1.01) | 0.93 (0.51, 1.36) | 5.02 (4.46, 5.57) | 6.32 (5.58, 7.05) |
| NSHD | In a relationship | 1 | 0.47 (0.41, 0.54) | 0.39 (0.33, 0.44) | 3.81 (3.73, 3.89) | 7.72 (7.59, 7.84) |
| NSHD | In a relationship | 2 | 0.59 (0.53, 0.65) | 0.41 (0.36, 0.46) | 3.67 (3.60, 3.73) | 7.65 (7.54, 7.75) |
| NSHD | In a relationship | 3 | 0.58 (0.51, 0.64) | 0.49 (0.43, 0.55) | 3.96 (3.88, 4.04) | 7.11 (6.99, 7.23) |
| NCDS | Not in a relationship | 1 | 0.98 (0.87, 1.09) | 1.01 (0.90, 1.11) | 5.05 (4.92, 5.19) | 6.60 (6.43, 6.76) |
| NCDS | Not in a relationship | 2 | 0.99 (0.89, 1.10) | 1.02 (0.92, 1.13) | 4.89 (4.76, 5.01) | 6.50 (6.35, 6.65) |
| NCDS | Not in a relationship | 3 | 0.96 (0.86, 1.07) | 1.10 (1.00, 1.21) | 5.15 (5.01, 5.28) | 6.21 (6.06, 6.37) |
| NCDS | In a relationship | 1 | 0.68 (0.65, 0.72) | 0.58 (0.55, 0.61) | 3.94 (3.90, 3.98) | 7.66 (7.60, 7.71) |
| NCDS | In a relationship | 2 | 0.73 (0.70, 0.76) | 0.59 (0.55, 0.62) | 3.84 (3.80, 3.88) | 7.61 (7.56, 7.66) |
| NCDS | In a relationship | 3 | 0.71 (0.68, 0.75) | 0.71 (0.67, 0.74) | 4.10 (4.06, 4.14) | 7.15 (7.10, 7.20) |
| BCS | Not in a relationship | 1 | 1.26 (1.13, 1.40) | 1.34 (1.21, 1.47) | 5.28 (5.13, 5.43) | 6.33 (6.16, 6.49) |
| BCS | Not in a relationship | 2 | 1.39 (1.25, 1.53) | 1.31 (1.18, 1.44) | 5.24 (5.09, 5.40) | 6.08 (5.92, 6.24) |
| BCS | Not in a relationship | 3 | 1.33 (1.19, 1.46) | 1.47 (1.34, 1.61) | 5.36 (5.21, 5.50) | 6.03 (5.86, 6.19) |
| BCS | In a relationship | 1 | 0.87 (0.83, 0.92) | 0.80 (0.76, 0.85) | 4.01 (3.96, 4.06) | 7.39 (7.32, 7.45) |
| BCS | In a relationship | 2 | 0.98 (0.94, 1.02) | 0.78 (0.74, 0.82) | 4.05 (4.01, 4.09) | 7.31 (7.26, 7.37) |
| BCS | In a relationship | 3 | 0.94 (0.90, 0.98) | 0.93 (0.89, 0.97) | 4.21 (4.17, 4.26) | 6.94 (6.89, 7.00) |
| NS | Not in a relationship | 1 | 1.80 (1.63, 1.97) | 1.90 (1.73, 2.06) | 5.61 (5.46, 5.77) | 5.93 (5.72, 6.14) |
| NS | Not in a relationship | 2 | 1.84 (1.70, 1.98) | 1.74 (1.60, 1.88) | 5.56 (5.42, 5.69) | 6.05 (5.89, 6.21) |
| NS | Not in a relationship | 3 | 1.76 (1.63, 1.88) | 1.77 (1.65, 1.89) | 5.52 (5.39, 5.64) | 5.87 (5.72, 6.01) |
| NS | In a relationship | 1 | 1.37 (1.29, 1.45) | 1.26 (1.19, 1.34) | 4.33 (4.26, 4.40) | 7.10 (7.01, 7.20) |
| NS | In a relationship | 2 | 1.53 (1.46, 1.59) | 1.14 (1.08, 1.19) | 4.35 (4.29, 4.41) | 7.20 (7.13, 7.27) |
| NS | In a relationship | 3 | 1.57 (1.51, 1.63) | 1.39 (1.33, 1.45) | 4.58 (4.53, 4.64) | 6.76 (6.69, 6.84) |
| MCS | Not in a relationship | 1 | 1.60 (1.51, 1.68) | 1.81 (1.73, 1.90) | 5.29 (5.20, 5.38) | 6.07 (5.95, 6.19) |
| MCS | Not in a relationship | 2 | 1.93 (1.84, 2.01) | 1.65 (1.57, 1.73) | 5.31 (5.22, 5.39) | 6.24 (6.14, 6.34) |
| MCS | Not in a relationship | 3 | 1.95 (1.88, 2.02) | 1.89 (1.82, 1.96) | 5.30 (5.22, 5.37) | 5.89 (5.80, 5.98) |
| MCS | In a relationship | 1 | 2.02 (1.91, 2.13) | 1.99 (1.89, 2.10) | 4.94 (4.84, 5.05) | 6.35 (6.21, 6.48) |
| MCS | In a relationship | 2 | 2.27 (2.16, 2.37) | 1.74 (1.64, 1.83) | 5.00 (4.90, 5.10) | 6.57 (6.46, 6.69) |
| MCS | In a relationship | 3 | 2.31 (2.22, 2.40) | 2.09 (2.01, 2.18) | 5.17 (5.08, 5.25) | 6.07 (5.96, 6.17) |
| **Cohort** | **Relationship status** | **Survey wave** | **Adjusted marginal mean (95% CI)** | **Adjusted marginal mean (95% CI)** | **Adjusted marginal mean (95% CI)** | **Adjusted marginal mean (95% CI)** |
| NSHD | Not in a relationship | 1 | 0.33 (-0.11, 0.77) | 0.18 (-0.24, 0.60) | 4.63 (3.82, 5.45) | 7.60 (6.83, 8.38) |
| NSHD | Not in a relationship | 2 | 0.50 (0.04, 0.96) | 0.47 (0.01, 0.93) | 4.29 (3.59, 4.98) | 6.25 (5.41, 7.08) |
| NSHD | Not in a relationship | 3 | 0.45 (-0.14, 1.04) | 0.67 (0.20, 1.14) | 4.57 (3.82, 5.31) | 6.69 (6.01, 7.38) |
| NSHD | In a relationship | 1 | 0.61 (0.54, 0.69) | 0.50 (0.44, 0.57) | 3.96 (3.87, 4.06) | 7.46 (7.32, 7.60) |
| NSHD | In a relationship | 2 | 0.75 (0.67, 0.83) | 0.54 (0.48, 0.61) | 3.79 (3.71, 3.88) | 7.39 (7.26, 7.53) |
| NSHD | In a relationship | 3 | 0.75 (0.67, 0.84) | 0.62 (0.55, 0.69) | 4.09 (3.99, 4.19) | 6.84 (6.69, 7.00) |
| NCDS | Not in a relationship | 1 | 0.77 (0.66, 0.88) | 0.87 (0.75, 0.99) | 4.78 (4.64, 4.93) | 6.88 (6.71, 7.05) |
| NCDS | Not in a relationship | 2 | 0.83 (0.72, 0.94) | 0.86 (0.75, 0.97) | 4.66 (4.52, 4.80) | 6.80 (6.64, 6.96) |
| NCDS | Not in a relationship | 3 | 0.76 (0.65, 0.86) | 0.95 (0.84, 1.06) | 4.90 (4.76, 5.05) | 6.45 (6.28, 6.62) |
| NCDS | In a relationship | 1 | 0.75 (0.71, 0.80) | 0.66 (0.62, 0.70) | 4.02 (3.97, 4.06) | 7.52 (7.46, 7.59) |
| NCDS | In a relationship | 2 | 0.80 (0.76, 0.85) | 0.66 (0.62, 0.70) | 3.92 (3.88, 3.97) | 7.47 (7.41, 7.53) |
| NCDS | In a relationship | 3 | 0.79 (0.75, 0.83) | 0.79 (0.75, 0.82) | 4.17 (4.13, 4.22) | 7.03 (6.96, 7.09) |
| BCS | Not in a relationship | 1 | 0.95 (0.80, 1.09) | 1.00 (0.86, 1.13) | 4.81 (4.65, 4.97) | 6.87 (6.69, 7.05) |
| BCS | Not in a relationship | 2 | 1.02 (0.87, 1.17) | 0.99 (0.86, 1.11) | 4.80 (4.64, 4.97) | 6.64 (6.46, 6.81) |
| BCS | Not in a relationship | 3 | 0.92 (0.78, 1.06) | 1.15 (1.01, 1.29) | 4.90 (4.74, 5.06) | 6.51 (6.32, 6.69) |
| BCS | In a relationship | 1 | 0.89 (0.84, 0.94) | 0.85 (0.80, 0.90) | 4.07 (4.01, 4.13) | 7.28 (7.20, 7.35) |
| BCS | In a relationship | 2 | 1.00 (0.95, 1.05) | 0.81 (0.77, 0.86) | 4.08 (4.03, 4.14) | 7.24 (7.17, 7.31) |
| BCS | In a relationship | 3 | 0.94 (0.89, 0.98) | 0.93 (0.89, 0.98) | 4.23 (4.18, 4.29) | 6.89 (6.83, 6.96) |
| NS | Not in a relationship | 1 | 1.51 (1.33, 1.70) | 1.68 (1.51, 1.86) | 5.37 (5.20, 5.53) | 6.32 (6.09, 6.55) |
| NS | Not in a relationship | 2 | 1.61 (1.46, 1.76) | 1.54 (1.39, 1.68) | 5.34 (5.19, 5.49) | 6.39 (6.21, 6.58) |
| NS | Not in a relationship | 3 | 1.48 (1.34, 1.62) | 1.50 (1.37, 1.63) | 5.26 (5.12, 5.41) | 6.30 (6.13, 6.47) |
| NS | In a relationship | 1 | 1.27 (1.19, 1.35) | 1.21 (1.14, 1.29) | 4.29 (4.21, 4.37) | 7.13 (7.02, 7.23) |
| NS | In a relationship | 2 | 1.43 (1.36, 1.50) | 1.10 (1.04, 1.16) | 4.31 (4.24, 4.37) | 7.22 (7.14, 7.31) |
| NS | In a relationship | 3 | 1.46 (1.39, 1.52) | 1.33 (1.27, 1.39) | 4.55 (4.49, 4.61) | 6.77 (6.69, 6.85) |
| MCS | Not in a relationship | 1 | 1.54 (1.44, 1.65) | 1.79 (1.69, 1.89) | 5.21 (5.10, 5.33) | 6.30 (6.16, 6.43) |
| MCS | Not in a relationship | 2 | 1.92 (1.81, 2.02) | 1.69 (1.59, 1.79) | 5.30 (5.20, 5.41) | 6.41 (6.29, 6.54) |
| MCS | Not in a relationship | 3 | 1.93 (1.83, 2.02) | 1.91 (1.82, 2.00) | 5.26 (5.17, 5.36) | 6.05 (5.93, 6.17) |
| MCS | In a relationship | 1 | 1.84 (1.71, 1.96) | 1.88 (1.76, 2.00) | 4.79 (4.68, 4.91) | 6.61 (6.46, 6.77) |
| MCS | In a relationship | 2 | 2.09 (1.97, 2.20) | 1.63 (1.52, 1.74) | 4.85 (4.73, 4.97) | 6.86 (6.72, 7.00) |
| MCS | In a relationship | 3 | 2.14 (2.04, 2.24) | 2.02 (1.92, 2.11) | 5.05 (4.95, 5.16) | 6.29 (6.16, 6.42) |

*Note.* Adjusted models included birth sex, highest qualification achieved, pre-pandemic self-reported health, pre-pandemic psychological distress, and household composition as covariates. BCS: British Cohort Study, 1970 birth cohort; GAD-2: 2-item General Anxiety Disorder questionnaire; MCS: Millennium Cohort Study, 2000 birth cohort; NCDS: National Child and Development Study, 1958 birth cohort; NS: Next Steps, 1990 cohort; NSHD: National Survey of Health and Development, 1946 birth cohort; ONS: UK Office for National Statistics; PHQ-2: 2-item Patient Health Questionnaire; UCLA-3: 3-item UCLA loneliness scale. Survey wave 1: May 2020; survey wave 2: September/October 2020; survey wave 3: February/March 2021.

#### Figure S8.1. Unadjusted and adjusted (by birth sex, highest qualification achieved, pre-pandemic self-reported health, pre-pandemic psychological distress, and household composition) anxiety symptomatology (GAD-2) marginal mean estimates and 95% confidence intervals by relationship status.

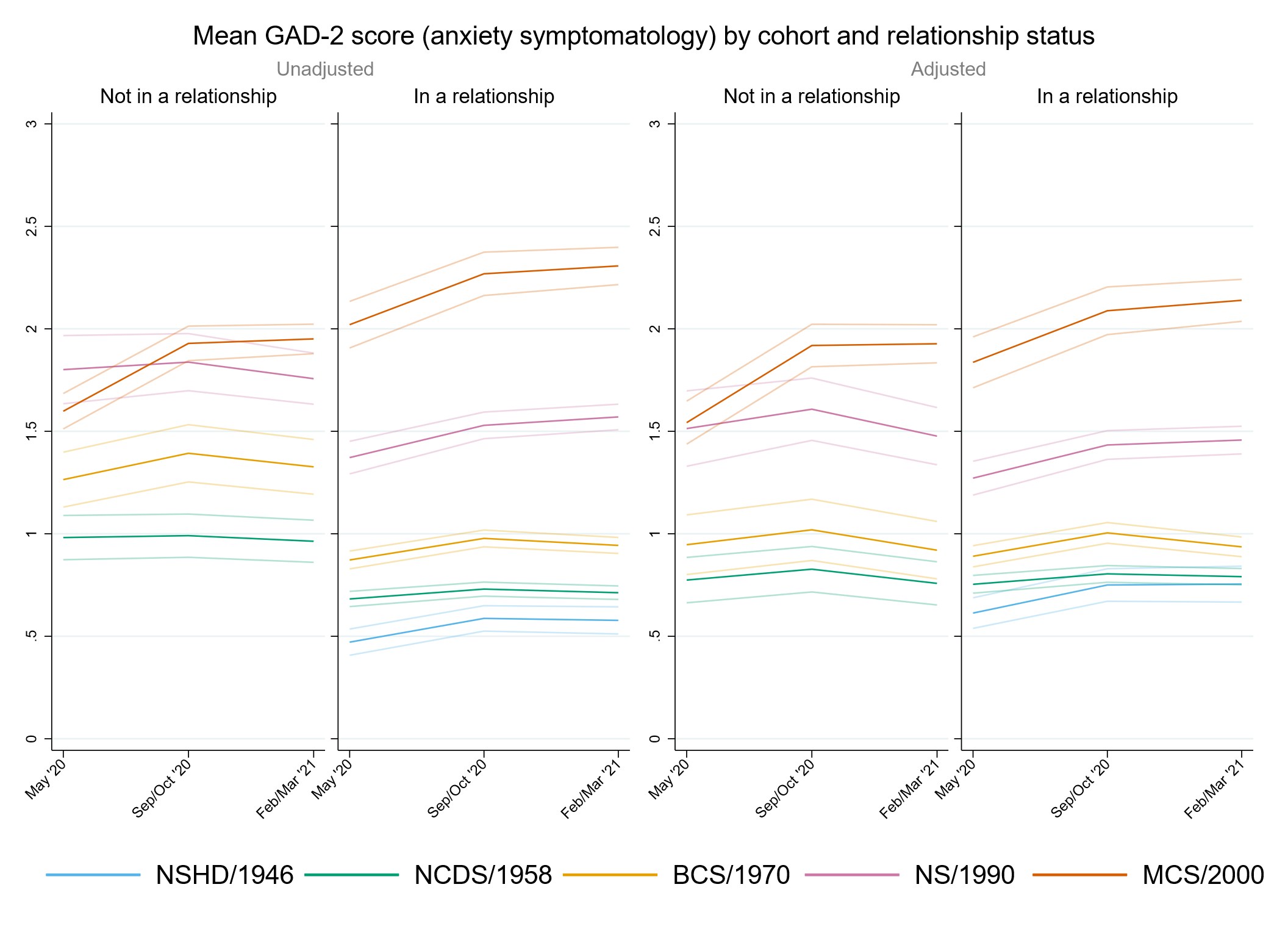

#### Figure S8.2. Unadjusted and adjusted (by birth sex, highest qualification achieved, pre-pandemic self-reported health, pre-pandemic psychological distress, and household composition) depressive symptomatology (PHQ-2) marginal mean estimates and 95% confidence intervals by relationship status.

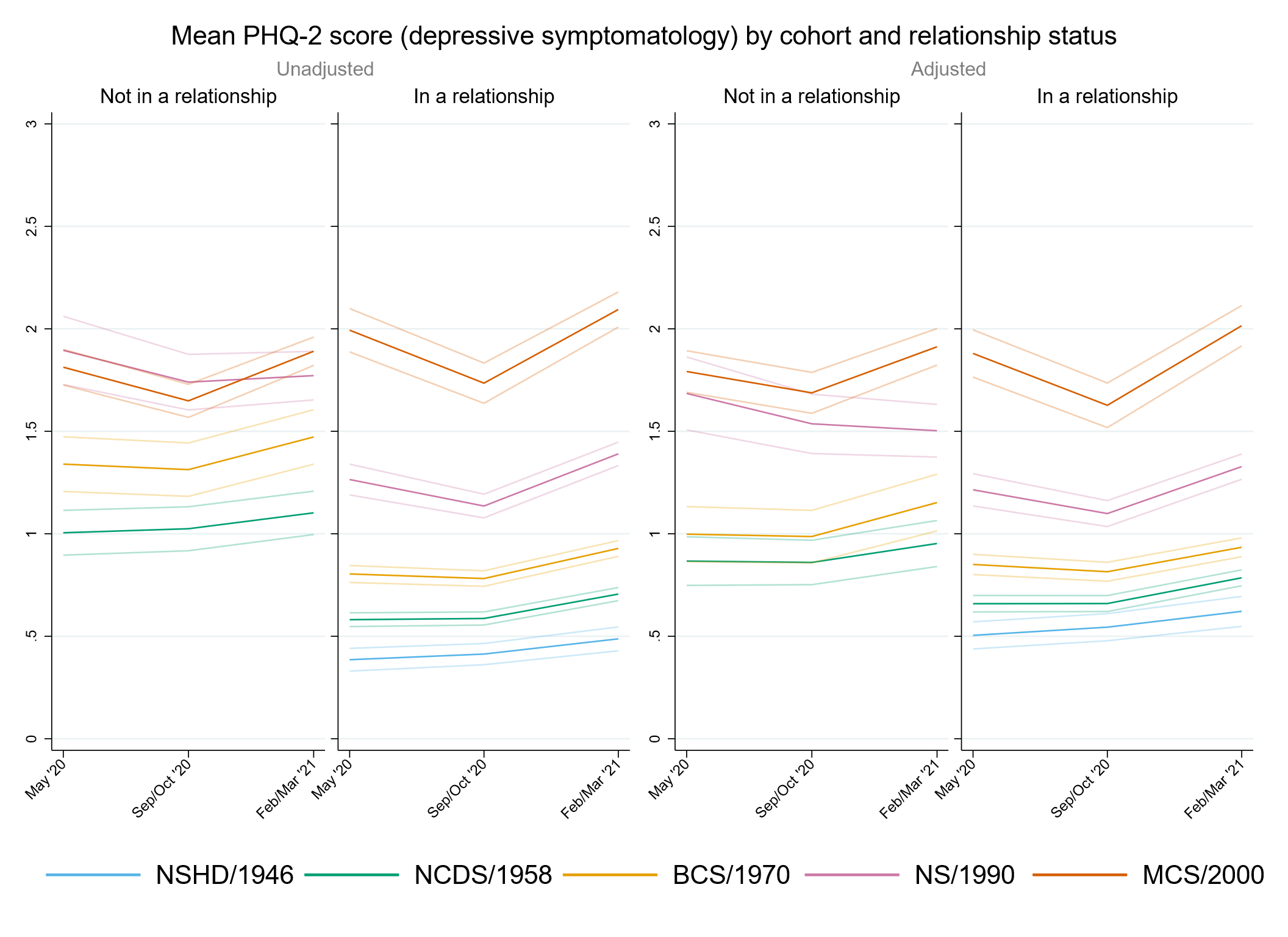

#### Figure S8.3. Unadjusted and adjusted (by birth sex, highest qualification achieved, pre-pandemic self-reported health, pre-pandemic psychological distress, and household composition) loneliness (UCLA-3) marginal mean estimates and 95% confidence intervals by relationship status.

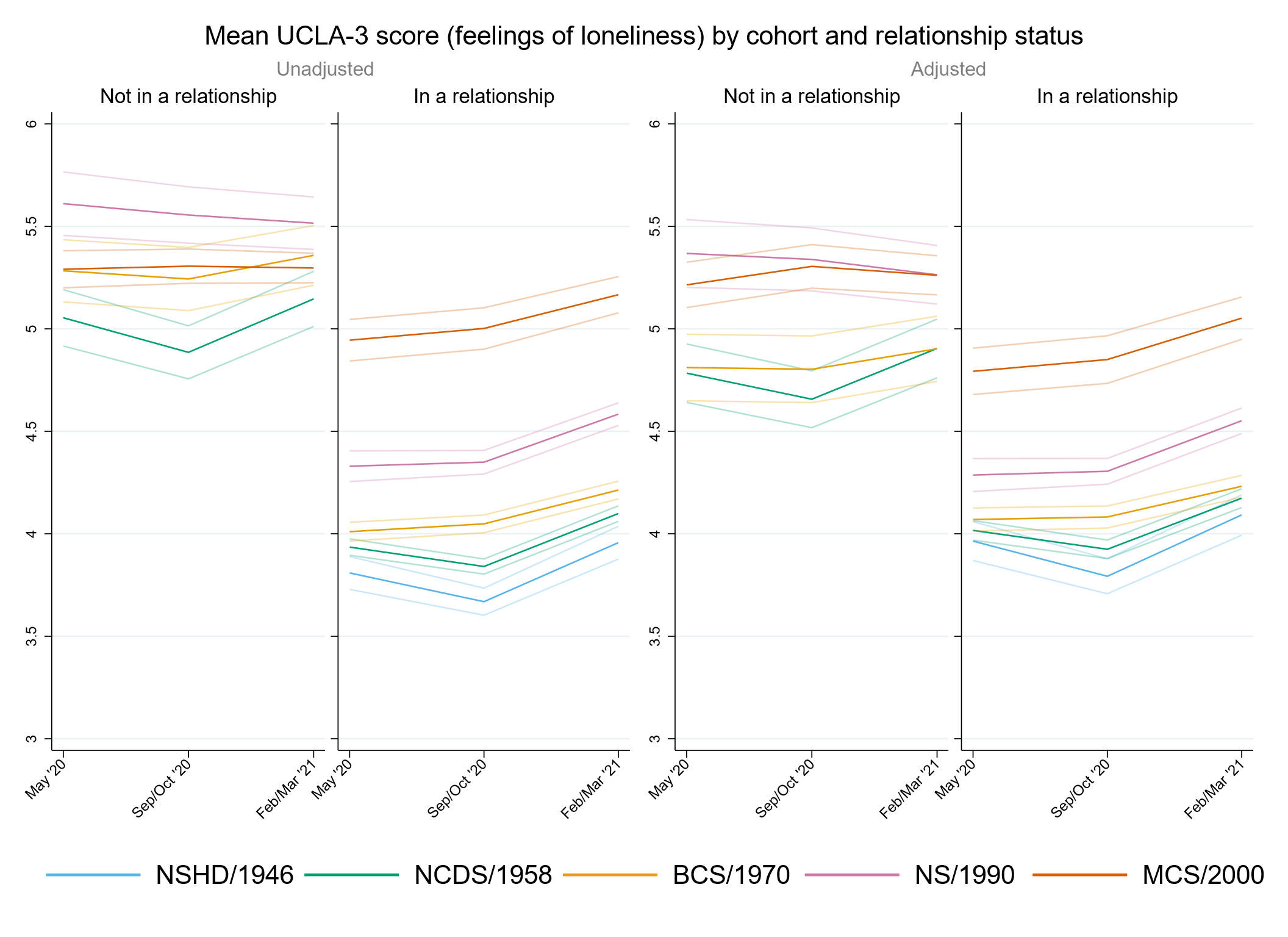

#### Figure S8.4. Unadjusted and adjusted (by birth sex, highest qualification achieved, pre-pandemic self-reported health, pre-pandemic psychological distress, and household composition) life satisfaction marginal mean estimates and 95% confidence intervals by relationship status.

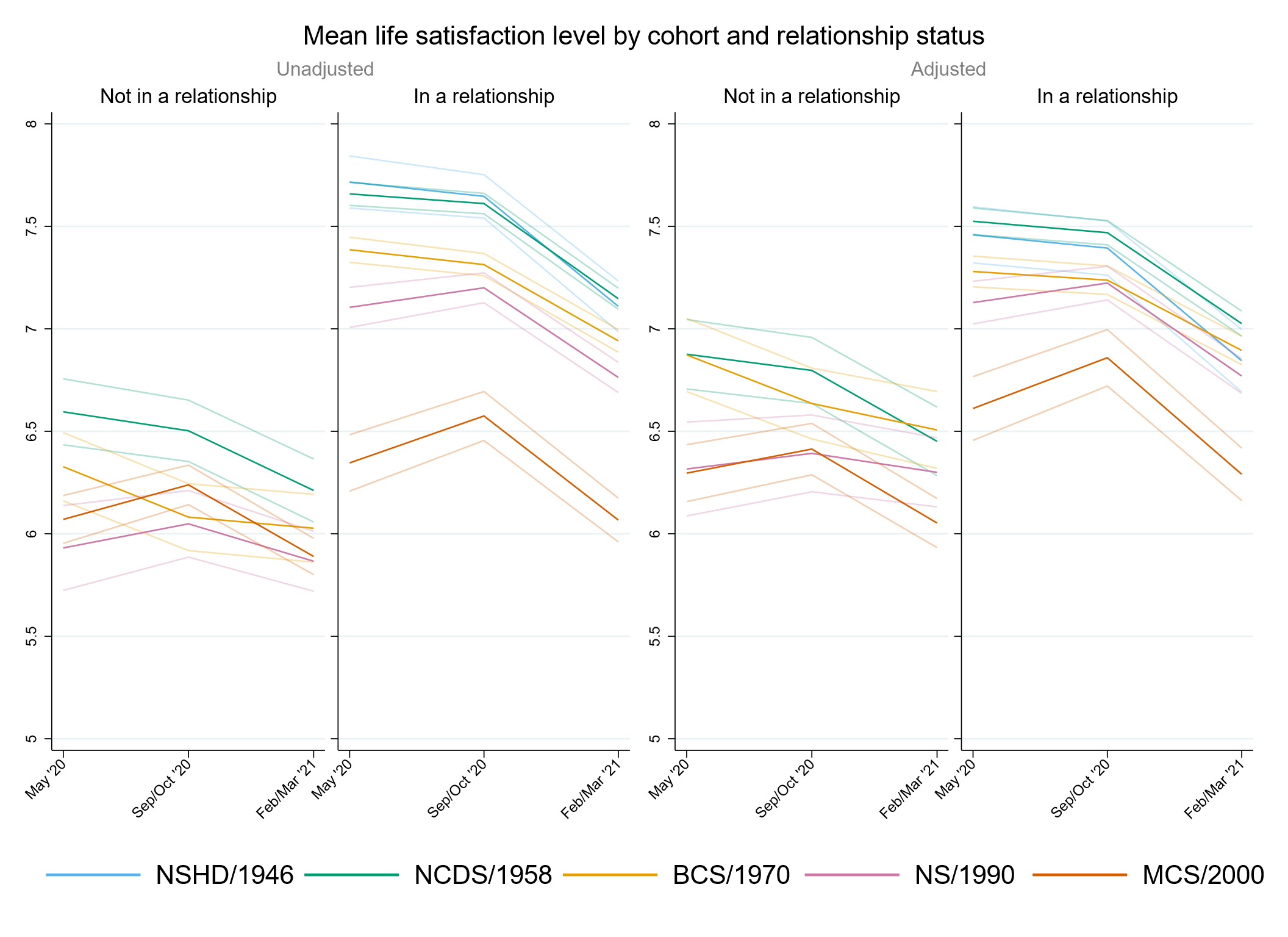

### Appendix S9. Results by housing tenure.

#### Table S9.1. Results of multilevel growth curve models by housing tenure.

|  | **Anxiety symptomatology (GAD-2)** | | | **Depressive symptomatology (PHQ-2)** | | | **Feelings of loneliness (UCLA-3)** | | | **Life satisfaction (ONS single question)** | | |
| --- | --- | --- | --- | --- | --- | --- | --- | --- | --- | --- | --- | --- |
| **Unadjusted models** |  | | |  | | |  | | |  | | |
| N participants | 23,117 |  |  | 23,111 |  |  | 23,141 |  |  | 23,174 |  |  |
| N observations | 50,511 |  |  | 50,487 |  |  | 50,571 |  |  | 50,760 |  |  |
|  | ***B* (95% CI)** | **χ2** | ***p*** | ***B* (95% CI)** | **χ2** | ***p*** | ***B* (95% CI)** | **χ2** | ***p*** | ***B* (95% CI)** | **χ2** | ***p*** |
| Time (linear) | 0.21 (0.00, 0.42) |  | 0.050 | 0.16 (-0.03, 0.36) |  | 0.100 | -0.38 (-0.61, -0.16) |  | 0.001 | -0.04 (-0.35, 0.27) |  | 0.807 |
| Time (quadratic) | -0.10 (-0.20, -0.01) |  | 0.038 | -0.03 (-0.12, 0.06) |  | 0.485 | 0.25 (0.15, 0.35) |  | <0.001 | -0.07 (-0.21, 0.07) |  | 0.346 |
| Cohort (ref. NCDS) |  | 137 | <0.001 |  | 208.3 | <0.001 |  | 50.4 | <0.001 |  | 50.5 | <0.001 |
| NSHD | -0.27 (-0.58, 0.03) |  | 0.082 | -0.30 (-0.56, -0.05) |  | 0.021 | -0.28 (-0.71, 0.16) |  | 0.215 | 0.25 (-0.40, 0.91) |  | 0.446 |
| BCS | 0.39 (0.18, 0.59) |  | <0.001 | 0.45 (0.26, 0.65) |  | <0.001 | 0.27 (0.05, 0.50) |  | 0.015 | -0.59 (-0.87, -0.30) |  | <0.001 |
| NS | 0.64 (0.46, 0.82) |  | <0.001 | 0.68 (0.51, 0.86) |  | <0.001 | 0.29 (0.10, 0.49) |  | 0.004 | -0.54 (-0.79, -0.28) |  | <0.001 |
| MCS | 0.86 (0.69, 1.03) |  | <0.001 | 1.00 (0.84, 1.17) |  | <0.001 | 0.59 (0.40, 0.77) |  | <0.001 | -0.89 (-1.13, -0.64) |  | <0.001 |
| Linear change * cohort (ref. NCDS) |  | 1.8 | 0.782 |  | 29.3 | <0.001 |  | 14.8 | 0.005 |  | 10 | 0.041 |
| NSHD | -0.03 (-0.52, 0.47) |  | 0.913 | -0.30 (-0.71, 0.11) |  | 0.153 | -0.47 (-1.02, 0.09) |  | 0.098 | 0.17 (-0.80, 1.14) |  | 0.727 |
| BCS | -0.02 (-0.36, 0.32) |  | 0.906 | -0.36 (-0.67, -0.05) |  | 0.023 | 0.28 (-0.06, 0.61) |  | 0.108 | 0.04 (-0.40, 0.47) |  | 0.866 |
| NS | 0.06 (-0.24, 0.35) |  | 0.705 | -0.47 (-0.75, -0.19) |  | 0.001 | 0.37 (0.07, 0.67) |  | 0.014 | 0.22 (-0.18, 0.62) |  | 0.289 |
| MCS | 0.15 (-0.13, 0.43) |  | 0.296 | -0.73 (-1.00, -0.46) |  | <0.001 | 0.34 (0.05, 0.63) |  | 0.021 | 0.53 (0.14, 0.92) |  | 0.008 |
| Quadratic change * cohort (ref. NCDS) |  | 0.5 | 0.974 |  | 27.6 | <0.001 |  | 20.9 | <0.001 |  | 10.6 | 0.031 |
| NSHD | -0.06 (-0.29, 0.16) |  | 0.582 | 0.06 (-0.15, 0.26) |  | 0.589 | 0.20 (-0.05, 0.45) |  | 0.112 | -0.11 (-0.59, 0.37) |  | 0.644 |
| BCS | 0.02 (-0.13, 0.17) |  | 0.792 | 0.18 (0.04, 0.32) |  | 0.014 | -0.15 (-0.30, 0.00) |  | 0.056 | 0.00 (-0.19, 0.20) |  | 0.966 |
| NS | 0.00 (-0.13, 0.14) |  | 0.949 | 0.19 (0.07, 0.32) |  | 0.003 | -0.20 (-0.33, -0.07) |  | 0.003 | -0.07 (-0.25, 0.11) |  | 0.442 |
| MCS | 0.00 (-0.13, 0.12) |  | 0.979 | 0.32 (0.20, 0.45) |  | <0.001 | -0.21 (-0.34, -0.08) |  | 0.001 | -0.23 (-0.40, -0.05) |  | 0.011 |
| Housing tenure (ref. Rented/rent-free/other arrangement) | -0.32 (-0.46, -0.18) |  | <0.001 | -0.42 (-0.55, -0.29) |  | <0.001 | -0.62 (-0.79, -0.46) |  | <0.001 | 0.57 (0.36, 0.79) |  | <0.001 |
| Linear change * housing tenure (ref. Rented/rent-free/other arrangement) | -0.15 (-0.37, 0.07) |  | 0.195 | -0.23 (-0.43, -0.03) |  | 0.024 | 0.12 (-0.11, 0.35) |  | 0.322 | 0.19 (-0.14, 0.51) |  | 0.257 |
| Quadratic change * housing tenure (ref. Rented/rent-free/other arrangement) | 0.07 (-0.02, 0.17) |  | 0.141 | 0.10 (0.00, 0.19) |  | 0.046 | -0.07 (-0.18, 0.03) |  | 0.184 | -0.14 (-0.29, 0.01) |  | 0.065 |
| Cohort (ref. NCDS) * housing tenure (ref. Rented/rent-free/other arrangement) |  | 10.7 | 0.03 |  | 23.9 | <0.001 |  | 41.5 | <0.001 |  | 37.3 | <0.001 |
| NSHD | 0.03 (-0.29, 0.34) |  | 0.872 | 0.10 (-0.16, 0.37) |  | 0.452 | 0.16 (-0.29, 0.61) |  | 0.481 | -0.19 (-0.86, 0.47) |  | 0.572 |
| BCS | -0.26 (-0.48, -0.05) |  | 0.015 | -0.31 (-0.52, -0.11) |  | 0.002 | -0.28 (-0.51, -0.05) |  | 0.019 | 0.44 (0.14, 0.74) |  | 0.004 |
| NS | -0.03 (-0.24, 0.18) |  | 0.746 | -0.12 (-0.32, 0.08) |  | 0.245 | 0.00 (-0.22, 0.22) |  | 0.977 | 0.09 (-0.20, 0.38) |  | 0.524 |
| MCS | 0.10 (-0.12, 0.31) |  | 0.377 | 0.20 (-0.01, 0.40) |  | 0.058 | 0.45 (0.23, 0.68) |  | <0.001 | -0.46 (-0.75, -0.16) |  | 0.002 |
| Linear change * cohort (ref. NCDS) * housing tenure (ref. Rented/rent-free/other arrangement) |  | 3.4 | 0.501 |  | 8.8 | 0.065 |  | 5.3 | 0.259 |  | 2.3 | 0.673 |
| NSHD | 0.21 (-0.30, 0.72) |  | 0.420 | 0.43 (0.01, 0.86) |  | 0.047 | 0.41 (-0.16, 0.99) |  | 0.157 | -0.28 (-1.28, 0.71) |  | 0.580 |
| BCS | 0.14 (-0.21, 0.48) |  | 0.445 | 0.36 (0.04, 0.68) |  | 0.029 | -0.04 (-0.39, 0.31) |  | 0.834 | -0.18 (-0.63, 0.28) |  | 0.443 |
| NS | 0.00 (-0.34, 0.35) |  | 0.978 | 0.27 (-0.05, 0.59) |  | 0.098 | -0.23 (-0.57, 0.11) |  | 0.180 | 0.14 (-0.31, 0.59) |  | 0.545 |
| MCS | 0.29 (-0.07, 0.65) |  | 0.111 | 0.41 (0.07, 0.75) |  | 0.019 | -0.04 (-0.39, 0.31) |  | 0.830 | -0.08 (-0.56, 0.40) |  | 0.740 |
| Quadratic change * cohort (ref. NCDS) * housing tenure (ref. Rented/rent-free/other arrangement) |  | 3.7 | 0.443 |  | 7.4 | 0.118 |  | 7.3 | 0.122 |  | 2.8 | 0.601 |
| NSHD | 0.00 (-0.23, 0.23) |  | 0.996 | -0.12 (-0.34, 0.09) |  | 0.268 | -0.18 (-0.44, 0.08) |  | 0.185 | 0.15 (-0.35, 0.64) |  | 0.560 |
| BCS | -0.07 (-0.22, 0.09) |  | 0.400 | -0.17 (-0.32, -0.02) |  | 0.023 | 0.04 (-0.12, 0.20) |  | 0.636 | 0.08 (-0.13, 0.28) |  | 0.463 |
| NS | -0.01 (-0.16, 0.14) |  | 0.868 | -0.10 (-0.25, 0.04) |  | 0.168 | 0.14 (-0.01, 0.29) |  | 0.062 | -0.05 (-0.25, 0.15) |  | 0.604 |
| MCS | -0.14 (-0.31, 0.02) |  | 0.079 | -0.18 (-0.34, -0.02) |  | 0.027 | 0.05 (-0.11, 0.21) |  | 0.526 | 0.08 (-0.13, 0.30) |  | 0.438 |
| **Adjusted models** |  | | |  | | |  | | |  | | |
| N participants | 19,723 |  |  | 19,718 |  |  | 19,743 |  |  | 19,769 |  |  |
| N observations | 44,028 |  |  | 44,012 |  |  | 44,077 |  |  | 44,244 |  |  |
|  | ***B* (95% CI)** | **χ2** | ***p*** | ***B* (95% CI)** | **χ2** | ***p*** | ***B* (95% CI)** | **χ2** | ***p*** | ***B* (95% CI)** | **χ2** | ***p*** |
| Time (linear) | 0.21 (-0.02, 0.43) |  | 0.071 | 0.05 (-0.17, 0.27) |  | 0.680 | -0.42 (-0.65, -0.19) |  | <0.001 | -0.05 (-0.38, 0.29) |  | 0.783 |
| Time (quadratic) | -0.09 (-0.19, 0.01) |  | 0.090 | 0.02 (-0.09, 0.12) |  | 0.774 | 0.27 (0.16, 0.37) |  | <0.001 | -0.06 (-0.22, 0.09) |  | 0.436 |
| Cohort (ref. NCDS) |  | 118.7 | <0.001 |  | 160.1 | <0.001 |  | 23.4 | <0.001 |  | 27.4 | <0.001 |
| NSHD | 0.10 (-0.27, 0.47) |  | 0.600 | -0.08 (-0.36, 0.20) |  | 0.593 | -0.09 (-0.62, 0.44) |  | 0.741 | -0.17 (-0.91, 0.57) |  | 0.659 |
| BCS | 0.29 (0.08, 0.51) |  | 0.007 | 0.33 (0.13, 0.54) |  | 0.002 | 0.13 (-0.10, 0.36) |  | 0.275 | -0.57 (-0.87, -0.27) |  | <0.001 |
| NS | 0.61 (0.42, 0.79) |  | <0.001 | 0.64 (0.46, 0.82) |  | <0.001 | 0.31 (0.11, 0.51) |  | 0.002 | -0.53 (-0.80, -0.27) |  | <0.001 |
| MCS | 0.94 (0.76, 1.11) |  | <0.001 | 1.01 (0.84, 1.18) |  | <0.001 | 0.43 (0.24, 0.62) |  | <0.001 | -0.65 (-0.90, -0.39) |  | <0.001 |
| Linear change * cohort (ref. NCDS) |  | 2 | 0.736 |  | 15.2 | 0.004 |  | 16.1 | 0.003 |  | 6.1 | 0.193 |
| NSHD | 0.00 (-0.58, 0.57) |  | 0.993 | -0.09 (-0.52, 0.34) |  | 0.691 | -0.42 (-1.02, 0.18) |  | 0.173 | 0.71 (-0.78, 2.19) |  | 0.353 |
| BCS | 0.15 (-0.24, 0.54) |  | 0.443 | -0.23 (-0.59, 0.13) |  | 0.210 | 0.31 (-0.06, 0.68) |  | 0.105 | 0.19 (-0.28, 0.67) |  | 0.429 |
| NS | 0.15 (-0.17, 0.47) |  | 0.367 | -0.31 (-0.62, 0.00) |  | 0.047 | 0.42 (0.10, 0.73) |  | 0.010 | 0.23 (-0.21, 0.66) |  | 0.312 |
| MCS | 0.20 (-0.10, 0.50) |  | 0.194 | -0.57 (-0.87, -0.27) |  | <0.001 | 0.45 (0.15, 0.76) |  | 0.004 | 0.49 (0.07, 0.91) |  | 0.022 |
| Quadratic change * cohort (ref. NCDS) |  | 1.6 | 0.815 |  | 18.5 | 0.001 |  | 21.1 | <0.001 |  | 7.6 | 0.107 |
| NSHD | -0.10 (-0.36, 0.16) |  | 0.447 | -0.08 (-0.30, 0.14) |  | 0.463 | 0.11 (-0.14, 0.36) |  | 0.378 | -0.43 (-1.30, 0.44) |  | 0.336 |
| BCS | -0.10 (-0.28, 0.08) |  | 0.271 | 0.09 (-0.08, 0.26) |  | 0.291 | -0.17 (-0.33, 0.00) |  | 0.050 | -0.04 (-0.26, 0.17) |  | 0.695 |
| NS | -0.05 (-0.19, 0.09) |  | 0.487 | 0.11 (-0.03, 0.25) |  | 0.114 | -0.23 (-0.37, -0.08) |  | 0.002 | -0.06 (-0.26, 0.13) |  | 0.528 |
| MCS | -0.03 (-0.17, 0.11) |  | 0.658 | 0.26 (0.12, 0.40) |  | <0.001 | -0.26 (-0.40, -0.12) |  | <0.001 | -0.22 (-0.41, -0.03) |  | 0.023 |
| Housing tenure (ref. Rented/rent-free/other arrangement) | -0.05 (-0.18, 0.09) |  | 0.487 | -0.18 (-0.32, -0.05) |  | 0.007 | -0.26 (-0.42, -0.11) |  | 0.001 | 0.19 (-0.02, 0.40) |  | 0.081 |
| Linear change * housing tenure (ref. Rented/rent-free/other arrangement) | -0.14 (-0.38, 0.09) |  | 0.239 | -0.13 (-0.35, 0.10) |  | 0.270 | 0.15 (-0.09, 0.39) |  | 0.214 | 0.19 (-0.16, 0.54) |  | 0.286 |
| Quadratic change * housing tenure (ref. Rented/rent-free/other arrangement) | 0.06 (-0.04, 0.17) |  | 0.246 | 0.06 (-0.05, 0.17) |  | 0.294 | -0.09 (-0.20, 0.02) |  | 0.108 | -0.14 (-0.30, 0.02) |  | 0.083 |
| Cohort (ref. NCDS) * housing tenure (ref. Rented/rent-free/other arrangement) |  | 4.4 | 0.351 |  | 6.9 | 0.139 |  | 7.1 | 0.29 |  | 14.7 | 0.005 |
| NSHD | -0.30 (-0.68, 0.08) |  | 0.122 | -0.09 (-0.38, 0.19) |  | 0.525 | 0.06 (-0.48, 0.60) |  | 0.817 | 0.08 (-0.68, 0.83) |  | 0.845 |
| BCS | -0.19 (-0.41, 0.03) |  | 0.089 | -0.20 (-0.41, 0.01) |  | 0.066 | -0.11 (-0.34, 0.13) |  | 0.380 | 0.40 (0.09, 0.71) |  | 0.011 |
| NS | -0.11 (-0.32, 0.10) |  | 0.315 | -0.15 (-0.36, 0.05) |  | 0.145 | -0.10 (-0.32, 0.12) |  | 0.352 | 0.20 (-0.09, 0.49) |  | 0.172 |
| MCS | -0.07 (-0.29, 0.14) |  | 0.491 | 0.05 (-0.15, 0.26) |  | 0.608 | 0.16 (-0.06, 0.39) |  | 0.157 | -0.16 (-0.46, 0.13) |  | 0.286 |
| Linear change * cohort (ref. NCDS) * housing tenure (ref. Rented/rent-free/other arrangement) |  | 3.3 | 0.508 |  | 3.2 | 0.525 |  | 5 | 0.29 |  | 4.4 | 0.357 |
| NSHD | 0.20 (-0.39, 0.80) |  | 0.508 | 0.22 (-0.23, 0.67) |  | 0.344 | 0.35 (-0.27, 0.98) |  | 0.267 | -0.82 (-2.33, 0.69) |  | 0.288 |
| BCS | -0.05 (-0.45, 0.36) |  | 0.822 | 0.24 (-0.14, 0.61) |  | 0.215 | -0.09 (-0.48, 0.30) |  | 0.650 | -0.30 (-0.80, 0.20) |  | 0.237 |
| NS | -0.09 (-0.46, 0.28) |  | 0.637 | 0.15 (-0.20, 0.50) |  | 0.404 | -0.29 (-0.64, 0.07) |  | 0.115 | 0.15 (-0.34, 0.64) |  | 0.546 |
| MCS | 0.25 (-0.14, 0.63) |  | 0.206 | 0.31 (-0.07, 0.69) |  | 0.107 | -0.11 (-0.49, 0.27) |  | 0.570 | -0.04 (-0.55, 0.47) |  | 0.880 |
| Quadratic change * cohort (ref. NCDS) * housing tenure (ref. Rented/rent-free/other arrangement) |  | 3.8 | 0.437 |  | 3.4 | 0.496 |  | 6.4 | 0.171 |  | 4.2 | 0.384 |
| NSHD | 0.04 (-0.24, 0.31) |  | 0.798 | 0.02 (-0.21, 0.24) |  | 0.883 | -0.09 (-0.35, 0.18) |  | 0.518 | 0.47 (-0.41, 1.35) |  | 0.295 |
| BCS | 0.05 (-0.13, 0.24) |  | 0.570 | -0.09 (-0.27, 0.08) |  | 0.294 | 0.06 (-0.11, 0.24) |  | 0.493 | 0.12 (-0.10, 0.35) |  | 0.294 |
| NS | 0.04 (-0.13, 0.20) |  | 0.640 | -0.05 (-0.21, 0.11) |  | 0.576 | 0.17 (0.01, 0.33) |  | 0.033 | -0.07 (-0.29, 0.15) |  | 0.543 |
| MCS | -0.12 (-0.29, 0.06) |  | 0.187 | -0.14 (-0.32, 0.03) |  | 0.109 | 0.09 (-0.09, 0.27) |  | 0.313 | 0.06 (-0.17, 0.29) |  | 0.624 |
| **Sensitivity models** |  | | |  | | |  | | |  | | |
| N participants | 19,723 |  |  | 19,718 |  |  | 19,743 |  |  | 19,769 |  |  |
| N observations | 44,028 |  |  | 44,012 |  |  | 44,077 |  |  | 44,244 |  |  |
|  | ***B* (95% CI)** | **χ2** | ***p*** | ***B* (95% CI)** | **χ2** | ***p*** | ***B* (95% CI)** | **χ2** | ***p*** | ***B* (95% CI)** | **χ2** | ***p*** |
| Time (linear) | 0.23 (0.00, 0.45) |  | 0.050 | 0.07 (-0.15, 0.29) |  | 0.545 | -0.39 (-0.62, -0.16) |  | 0.001 | -0.09 (-0.42, 0.25) |  | 0.620 |
| Time (quadratic) | -0.10 (-0.20, 0.01) |  | 0.069 | 0.01 (-0.10, 0.11) |  | 0.891 | 0.26 (0.15, 0.36) |  | <0.001 | -0.05 (-0.21, 0.10) |  | 0.512 |
| Cohort (ref. NCDS) |  | 107.4 | <0.001 |  | 129.4 | <0.001 |  | 35.7 | <0.001 |  | 41.4 | <0.001 |
| NSHD | -0.09 (-0.48, 0.30) |  | 0.644 | -0.24 (-0.54, 0.05) |  | 0.108 | -0.29 (-0.82, 0.24) |  | 0.283 | 0.11 (-0.66, 0.88) |  | 0.780 |
| BCS | 0.37 (0.14, 0.59) |  | 0.001 | 0.37 (0.15, 0.59) |  | 0.001 | 0.14 (-0.10, 0.39) |  | 0.248 | -0.61 (-0.92, -0.30) |  | <0.001 |
| NS | 0.64 (0.45, 0.83) |  | <0.001 | 0.59 (0.40, 0.78) |  | <0.001 | 0.26 (0.05, 0.47) |  | 0.016 | -0.53 (-0.81, -0.26) |  | <0.001 |
| MCS | 0.87 (0.69, 1.05) |  | <0.001 | 0.88 (0.70, 1.06) |  | <0.001 | 0.52 (0.32, 0.72) |  | <0.001 | -0.81 (-1.07, -0.55) |  | <0.001 |
| Linear change * cohort (ref. NCDS) |  | 1.6 | 0.819 |  | 15.5 | 0.004 |  | 14.8 | 0.005 |  | 6.7 | 0.152 |
| NSHD | -0.03 (-0.61, 0.55) |  | 0.917 | -0.10 (-0.53, 0.33) |  | 0.638 | -0.41 (-1.01, 0.19) |  | 0.177 | 0.75 (-0.77, 2.26) |  | 0.335 |
| BCS | 0.13 (-0.26, 0.53) |  | 0.501 | -0.26 (-0.62, 0.11) |  | 0.168 | 0.29 (-0.08, 0.66) |  | 0.123 | 0.22 (-0.25, 0.70) |  | 0.355 |
| NS | 0.12 (-0.20, 0.44) |  | 0.472 | -0.33 (-0.64, -0.02) |  | 0.038 | 0.41 (0.09, 0.73) |  | 0.013 | 0.24 (-0.19, 0.68) |  | 0.276 |
| MCS | 0.17 (-0.13, 0.48) |  | 0.265 | -0.59 (-0.89, -0.28) |  | <0.001 | 0.43 (0.12, 0.74) |  | 0.007 | 0.52 (0.10, 0.95) |  | 0.015 |
| Quadratic change * cohort (ref. NCDS) |  | 1.3 | 0.868 |  | 17.8 | 0.001 |  | 20.6 | <0.001 |  | 7.2 | 0.124 |
| NSHD | -0.09 (-0.36, 0.17) |  | 0.493 | -0.08 (-0.30, 0.14) |  | 0.486 | 0.11 (-0.14, 0.36) |  | 0.399 | -0.44 (-1.33, 0.45) |  | 0.329 |
| BCS | -0.09 (-0.27, 0.09) |  | 0.316 | 0.10 (-0.07, 0.27) |  | 0.233 | -0.16 (-0.33, 0.01) |  | 0.060 | -0.05 (-0.27, 0.16) |  | 0.619 |
| NS | -0.04 (-0.19, 0.10) |  | 0.576 | 0.12 (-0.02, 0.26) |  | 0.100 | -0.23 (-0.37, -0.08) |  | 0.002 | -0.06 (-0.26, 0.13) |  | 0.522 |
| MCS | -0.03 (-0.17, 0.11) |  | 0.685 | 0.26 (0.12, 0.41) |  | <0.001 | -0.26 (-0.40, -0.12) |  | <0.001 | -0.22 (-0.41, -0.03) |  | 0.025 |
| Housing tenure (ref. Rented/rent-free/other arrangement) | -0.31 (-0.45, -0.17) |  | <0.001 | -0.51 (-0.66, -0.37) |  | <0.001 | -0.67 (-0.84, -0.50) |  | <0.001 | 0.60 (0.38, 0.82) |  | <0.001 |
| Linear change * housing tenure (ref. Rented/rent-free/other arrangement) | -0.16 (-0.39, 0.08) |  | 0.191 | -0.14 (-0.37, 0.09) |  | 0.219 | 0.13 (-0.11, 0.37) |  | 0.294 | 0.22 (-0.13, 0.57) |  | 0.223 |
| Quadratic change * housing tenure (ref. Rented/rent-free/other arrangement) | 0.07 (-0.04, 0.18) |  | 0.204 | 0.06 (-0.04, 0.17) |  | 0.250 | -0.08 (-0.19, 0.03) |  | 0.140 | -0.15 (-0.31, 0.01) |  | 0.070 |
| Cohort (ref. NCDS) * housing tenure (ref. Rented/rent-free/other arrangement) |  | 7.5 | 0.111 |  | 19.8 | <0.001 |  | 33 | <0.001 |  | 35 | <0.001 |
| NSHD | -0.14 (-0.54, 0.26) |  | 0.483 | 0.06 (-0.24, 0.37) |  | 0.689 | 0.21 (-0.33, 0.75) |  | 0.444 | -0.10 (-0.88, 0.69) |  | 0.810 |
| BCS | -0.23 (-0.47, 0.00) |  | 0.051 | -0.23 (-0.46, 0.00) |  | 0.051 | -0.14 (-0.40, 0.11) |  | 0.262 | 0.44 (0.12, 0.77) |  | 0.008 |
| NS | -0.03 (-0.25, 0.19) |  | 0.769 | -0.04 (-0.25, 0.18) |  | 0.744 | 0.02 (-0.22, 0.25) |  | 0.885 | 0.10 (-0.20, 0.41) |  | 0.507 |
| MCS | 0.11 (-0.12, 0.33) |  | 0.354 | 0.30 (0.09, 0.52) |  | 0.006 | 0.51 (0.27, 0.75) |  | <0.001 | -0.50 (-0.81, -0.19) |  | 0.002 |
| Linear change * cohort (ref. NCDS) * housing tenure (ref. Rented/rent-free/other arrangement) |  | 3.5 | 0.475 |  | 3.5 | 0.478 |  | 4.7 | 0.321 |  | 4.7 | 0.322 |
| NSHD | 0.23 (-0.37, 0.82) |  | 0.460 | 0.23 (-0.22, 0.68) |  | 0.315 | 0.34 (-0.28, 0.96) |  | 0.279 | -0.85 (-2.39, 0.68) |  | 0.276 |
| BCS | -0.04 (-0.44, 0.37) |  | 0.857 | 0.25 (-0.12, 0.63) |  | 0.185 | -0.08 (-0.47, 0.30) |  | 0.671 | -0.32 (-0.82, 0.18) |  | 0.204 |
| NS | -0.07 (-0.43, 0.30) |  | 0.729 | 0.16 (-0.19, 0.52) |  | 0.373 | -0.28 (-0.64, 0.08) |  | 0.126 | 0.14 (-0.35, 0.63) |  | 0.564 |
| MCS | 0.27 (-0.11, 0.66) |  | 0.165 | 0.33 (-0.06, 0.71) |  | 0.096 | -0.09 (-0.47, 0.30) |  | 0.654 | -0.08 (-0.59, 0.44) |  | 0.772 |
| Quadratic change * cohort (ref. NCDS) * housing tenure (ref. Rented/rent-free/other arrangement) |  | 3.9 | 0.419 |  | 3.6 | 0.466 |  | 6.3 | 0.178 |  | 4.6 | 0.329 |
| NSHD | 0.03 (-0.25, 0.30) |  | 0.847 | 0.01 (-0.21, 0.24) |  | 0.912 | -0.08 (-0.34, 0.18) |  | 0.545 | 0.48 (-0.41, 1.38) |  | 0.288 |
| BCS | 0.05 (-0.14, 0.23) |  | 0.602 | -0.10 (-0.28, 0.07) |  | 0.254 | 0.06 (-0.12, 0.23) |  | 0.514 | 0.13 (-0.10, 0.35) |  | 0.268 |
| NS | 0.03 (-0.14, 0.20) |  | 0.718 | -0.05 (-0.21, 0.11) |  | 0.534 | 0.17 (0.01, 0.33) |  | 0.032 | -0.07 (-0.29, 0.15) |  | 0.523 |
| MCS | -0.13 (-0.30, 0.05) |  | 0.154 | -0.15 (-0.33, 0.03) |  | 0.102 | 0.08 (-0.09, 0.26) |  | 0.361 | 0.07 (-0.17, 0.30) |  | 0.565 |

*Note.* Adjusted models included birth sex, highest qualification achieved, pre-pandemic self-reported health, pre-pandemic psychological distress, and household composition as covariates. Sensitivity models correspond to the unadjusted models after restricting the analytical sample to that of the adjusted models. BCS: British Cohort Study, 1970 birth cohort; GAD-2: 2-item General Anxiety Disorder questionnaire; MCS: Millennium Cohort Study, 2000 birth cohort; NCDS: National Child and Development Study, 1958 birth cohort; NS: Next Steps, 1990 cohort; NSHD: National Survey of Health and Development, 1946 birth cohort; ONS: UK Office for National Statistics; PHQ-2: 2-item Patient Health Questionnaire; UCLA-3: 3-item UCLA loneliness scale. χ2: Wald test performed to assess the overall statistical significance of the interaction terms; all χ2 statistics in this table have 4 degrees of freedom.

#### Table S9.2. Unadjusted and adjusted marginal mean estimates and 95% confidence intervals by housing tenure.

|  |  |  | **Anxiety symptomatology (GAD-2)** | **Depressive symptomatology (PHQ-2)** | **Feelings of loneliness (UCLA-3)** | **Life satisfaction (ONS single question)** |
| --- | --- | --- | --- | --- | --- | --- |
| **Cohort** | **Housing tenure** | **Survey wave** | **Unadjusted marginal mean (95% CI)** | **Unadjusted marginal mean (95% CI)** | **Unadjusted marginal mean (95% CI)** | **Unadjusted marginal mean (95% CI)** |
| NSHD | No (rented/rent-free/other arrangement) | 1 | 0.75 (0.48, 1.03) | 0.73 (0.50, 0.95) | 4.41 (4.00, 4.82) | 7.22 (6.60, 7.84) |
| NSHD | No (rented/rent-free/other arrangement) | 2 | 0.78 (0.52, 1.03) | 0.62 (0.40, 0.83) | 4.01 (3.71, 4.30) | 7.18 (6.70, 7.65) |
| NSHD | No (rented/rent-free/other arrangement) | 3 | 0.47 (0.28, 0.66) | 0.55 (0.34, 0.77) | 4.51 (4.12, 4.89) | 6.76 (6.14, 7.39) |
| NSHD | Yes (house owned/partly owned) | 1 | 0.46 (0.40, 0.52) | 0.41 (0.35, 0.47) | 3.95 (3.86, 4.03) | 7.61 (7.48, 7.73) |
| NSHD | Yes (house owned/partly owned) | 2 | 0.62 (0.56, 0.68) | 0.48 (0.42, 0.53) | 3.83 (3.75, 3.90) | 7.47 (7.37, 7.58) |
| NSHD | Yes (house owned/partly owned) | 3 | 0.60 (0.54, 0.67) | 0.54 (0.48, 0.60) | 4.12 (4.03, 4.20) | 6.99 (6.87, 7.10) |
| NCDS | No (rented/rent-free/other arrangement) | 1 | 1.03 (0.89, 1.16) | 1.03 (0.90, 1.16) | 4.69 (4.53, 4.84) | 6.97 (6.77, 7.17) |
| NCDS | No (rented/rent-free/other arrangement) | 2 | 1.14 (1.02, 1.25) | 1.16 (1.04, 1.28) | 4.55 (4.42, 4.68) | 6.86 (6.70, 7.03) |
| NCDS | No (rented/rent-free/other arrangement) | 3 | 1.05 (0.94, 1.16) | 1.23 (1.11, 1.34) | 4.90 (4.77, 5.03) | 6.62 (6.45, 6.79) |
| NCDS | Yes (house owned/partly owned) | 1 | 0.71 (0.67, 0.74) | 0.61 (0.58, 0.65) | 4.06 (4.02, 4.11) | 7.54 (7.49, 7.60) |
| NCDS | Yes (house owned/partly owned) | 2 | 0.75 (0.71, 0.78) | 0.61 (0.58, 0.64) | 3.97 (3.93, 4.01) | 7.49 (7.44, 7.54) |
| NCDS | Yes (house owned/partly owned) | 3 | 0.73 (0.70, 0.77) | 0.73 (0.70, 0.77) | 4.23 (4.19, 4.27) | 7.01 (6.96, 7.07) |
| BCS | No (rented/rent-free/other arrangement) | 1 | 1.41 (1.26, 1.56) | 1.48 (1.33, 1.63) | 4.96 (4.80, 5.12) | 6.39 (6.19, 6.58) |
| BCS | No (rented/rent-free/other arrangement) | 2 | 1.52 (1.40, 1.65) | 1.43 (1.31, 1.55) | 4.95 (4.82, 5.09) | 6.32 (6.16, 6.48) |
| BCS | No (rented/rent-free/other arrangement) | 3 | 1.47 (1.35, 1.59) | 1.66 (1.54, 1.79) | 5.14 (5.01, 5.27) | 6.13 (5.97, 6.28) |
| BCS | Yes (house owned/partly owned) | 1 | 0.83 (0.78, 0.87) | 0.75 (0.71, 0.79) | 4.06 (4.01, 4.11) | 7.40 (7.33, 7.46) |
| BCS | Yes (house owned/partly owned) | 2 | 0.94 (0.90, 0.98) | 0.75 (0.71, 0.79) | 4.10 (4.05, 4.14) | 7.28 (7.23, 7.34) |
| BCS | Yes (house owned/partly owned) | 3 | 0.90 (0.86, 0.94) | 0.89 (0.85, 0.93) | 4.27 (4.22, 4.31) | 6.91 (6.85, 6.97) |
| NS | No (rented/rent-free/other arrangement) | 1 | 1.66 (1.54, 1.79) | 1.72 (1.59, 1.84) | 4.98 (4.86, 5.10) | 6.44 (6.28, 6.59) |
| NS | No (rented/rent-free/other arrangement) | 2 | 1.84 (1.74, 1.93) | 1.57 (1.48, 1.66) | 5.01 (4.92, 5.11) | 6.47 (6.36, 6.59) |
| NS | No (rented/rent-free/other arrangement) | 3 | 1.82 (1.73, 1.91) | 1.74 (1.65, 1.82) | 5.14 (5.05, 5.22) | 6.24 (6.13, 6.34) |
| NS | Yes (house owned/partly owned) | 1 | 1.31 (1.21, 1.41) | 1.18 (1.09, 1.27) | 4.36 (4.27, 4.45) | 7.10 (6.98, 7.22) |
| NS | Yes (house owned/partly owned) | 2 | 1.40 (1.33, 1.48) | 1.07 (1.00, 1.13) | 4.35 (4.28, 4.42) | 7.28 (7.20, 7.36) |
| NS | Yes (house owned/partly owned) | 3 | 1.43 (1.36, 1.50) | 1.26 (1.20, 1.32) | 4.57 (4.50, 4.64) | 6.80 (6.71, 6.88) |
| MCS | No (rented/rent-free/other arrangement) | 1 | 1.89 (1.78, 1.99) | 2.03 (1.93, 2.14) | 5.27 (5.17, 5.38) | 6.08 (5.95, 6.22) |
| MCS | No (rented/rent-free/other arrangement) | 2 | 2.14 (2.05, 2.24) | 1.76 (1.67, 1.84) | 5.26 (5.17, 5.35) | 6.28 (6.18, 6.38) |
| MCS | No (rented/rent-free/other arrangement) | 3 | 2.20 (2.12, 2.28) | 2.06 (1.99, 2.14) | 5.32 (5.24, 5.39) | 5.88 (5.79, 5.97) |
| MCS | Yes (house owned/partly owned) | 1 | 1.66 (1.54, 1.79) | 1.82 (1.70, 1.93) | 5.10 (4.99, 5.22) | 6.20 (6.04, 6.36) |
| MCS | Yes (house owned/partly owned) | 2 | 2.00 (1.90, 2.10) | 1.63 (1.54, 1.73) | 5.15 (5.05, 5.25) | 6.45 (6.33, 6.56) |
| MCS | Yes (house owned/partly owned) | 3 | 1.99 (1.90, 2.08) | 1.87 (1.78, 1.96) | 5.23 (5.13, 5.32) | 5.99 (5.88, 6.11) |
| **Cohort** | **Housing tenure** | **Survey wave** | **Adjusted marginal mean (95% CI)** | **Adjusted marginal mean (95% CI)** | **Adjusted marginal mean (95% CI)** | **Adjusted marginal mean (95% CI)** |
| NSHD | No (rented/rent-free/other arrangement) | 1 | 0.92 (0.57, 1.27) | 0.82 (0.57, 1.07) | 4.39 (3.88, 4.90) | 7.02 (6.31, 7.73) |
| NSHD | No (rented/rent-free/other arrangement) | 2 | 0.94 (0.64, 1.23) | 0.71 (0.46, 0.96) | 3.93 (3.61, 4.25) | 7.19 (6.55, 7.83) |
| NSHD | No (rented/rent-free/other arrangement) | 3 | 0.57 (0.32, 0.81) | 0.47 (0.27, 0.67) | 4.23 (3.81, 4.64) | 6.38 (5.40, 7.37) |
| NSHD | Yes (house owned/partly owned) | 1 | 0.57 (0.51, 0.64) | 0.54 (0.48, 0.61) | 4.19 (4.10, 4.29) | 7.28 (7.15, 7.41) |
| NSHD | Yes (house owned/partly owned) | 2 | 0.75 (0.67, 0.82) | 0.60 (0.53, 0.67) | 4.06 (3.97, 4.15) | 7.15 (7.03, 7.28) |
| NSHD | Yes (house owned/partly owned) | 3 | 0.73 (0.65, 0.82) | 0.67 (0.60, 0.74) | 4.33 (4.23, 4.43) | 6.70 (6.56, 6.83) |
| NCDS | No (rented/rent-free/other arrangement) | 1 | 0.82 (0.69, 0.95) | 0.90 (0.77, 1.02) | 4.48 (4.33, 4.63) | 7.19 (6.98, 7.39) |
| NCDS | No (rented/rent-free/other arrangement) | 2 | 0.94 (0.82, 1.06) | 0.96 (0.84, 1.08) | 4.32 (4.20, 4.45) | 7.08 (6.91, 7.25) |
| NCDS | No (rented/rent-free/other arrangement) | 3 | 0.88 (0.76, 0.99) | 1.05 (0.93, 1.17) | 4.70 (4.57, 4.83) | 6.85 (6.67, 7.02) |
| NCDS | Yes (house owned/partly owned) | 1 | 0.77 (0.73, 0.82) | 0.71 (0.67, 0.75) | 4.22 (4.17, 4.27) | 7.38 (7.31, 7.44) |
| NCDS | Yes (house owned/partly owned) | 2 | 0.82 (0.78, 0.85) | 0.70 (0.67, 0.74) | 4.12 (4.08, 4.17) | 7.31 (7.26, 7.37) |
| NCDS | Yes (house owned/partly owned) | 3 | 0.80 (0.77, 0.84) | 0.84 (0.80, 0.88) | 4.38 (4.34, 4.43) | 6.84 (6.79, 6.90) |
| BCS | No (rented/rent-free/other arrangement) | 1 | 1.12 (0.95, 1.29) | 1.23 (1.07, 1.39) | 4.61 (4.44, 4.78) | 6.62 (6.40, 6.83) |
| BCS | No (rented/rent-free/other arrangement) | 2 | 1.29 (1.15, 1.42) | 1.15 (1.02, 1.28) | 4.59 (4.44, 4.74) | 6.66 (6.48, 6.83) |
| BCS | No (rented/rent-free/other arrangement) | 3 | 1.08 (0.95, 1.21) | 1.28 (1.15, 1.41) | 4.77 (4.63, 4.92) | 6.49 (6.32, 6.66) |
| BCS | Yes (house owned/partly owned) | 1 | 0.88 (0.83, 0.93) | 0.85 (0.80, 0.89) | 4.24 (4.18, 4.30) | 7.21 (7.13, 7.28) |
| BCS | Yes (house owned/partly owned) | 2 | 0.98 (0.93, 1.02) | 0.84 (0.80, 0.88) | 4.26 (4.20, 4.31) | 7.11 (7.05, 7.18) |
| BCS | Yes (house owned/partly owned) | 3 | 0.93 (0.89, 0.98) | 0.97 (0.93, 1.02) | 4.41 (4.36, 4.47) | 6.77 (6.70, 6.84) |
| NS | No (rented/rent-free/other arrangement) | 1 | 1.43 (1.30, 1.56) | 1.54 (1.41, 1.66) | 4.79 (4.66, 4.92) | 6.65 (6.49, 6.82) |
| NS | No (rented/rent-free/other arrangement) | 2 | 1.64 (1.54, 1.74) | 1.40 (1.30, 1.49) | 4.82 (4.72, 4.92) | 6.71 (6.59, 6.83) |
| NS | No (rented/rent-free/other arrangement) | 3 | 1.57 (1.48, 1.67) | 1.52 (1.43, 1.61) | 4.93 (4.84, 5.02) | 6.51 (6.40, 6.62) |
| NS | Yes (house owned/partly owned) | 1 | 1.27 (1.18, 1.37) | 1.20 (1.11, 1.30) | 4.42 (4.33, 4.51) | 7.05 (6.93, 7.16) |
| NS | Yes (house owned/partly owned) | 2 | 1.36 (1.28, 1.44) | 1.10 (1.03, 1.17) | 4.40 (4.33, 4.47) | 7.23 (7.14, 7.31) |
| NS | Yes (house owned/partly owned) | 3 | 1.37 (1.30, 1.45) | 1.28 (1.21, 1.34) | 4.63 (4.55, 4.70) | 6.74 (6.65, 6.83) |
| MCS | No (rented/rent-free/other arrangement) | 1 | 1.76 (1.64, 1.88) | 1.91 (1.79, 2.02) | 4.91 (4.79, 5.02) | 6.54 (6.39, 6.69) |
| MCS | No (rented/rent-free/other arrangement) | 2 | 2.04 (1.94, 2.15) | 1.66 (1.56, 1.76) | 4.94 (4.84, 5.05) | 6.70 (6.58, 6.82) |
| MCS | No (rented/rent-free/other arrangement) | 3 | 2.09 (2.00, 2.18) | 1.97 (1.88, 2.06) | 4.98 (4.89, 5.07) | 6.30 (6.19, 6.41) |
| MCS | Yes (house owned/partly owned) | 1 | 1.64 (1.51, 1.77) | 1.78 (1.65, 1.90) | 4.81 (4.68, 4.94) | 6.57 (6.40, 6.74) |
| MCS | Yes (house owned/partly owned) | 2 | 1.98 (1.86, 2.09) | 1.63 (1.52, 1.73) | 4.88 (4.77, 5.00) | 6.79 (6.66, 6.93) |
| MCS | Yes (house owned/partly owned) | 3 | 1.97 (1.86, 2.07) | 1.86 (1.76, 1.96) | 4.96 (4.85, 5.07) | 6.29 (6.15, 6.42) |

*Note.* Adjusted models included birth sex, highest qualification achieved, pre-pandemic self-reported health, pre-pandemic psychological distress, and household composition as covariates. BCS: British Cohort Study, 1970 birth cohort; GAD-2: 2-item General Anxiety Disorder questionnaire; MCS: Millennium Cohort Study, 2000 birth cohort; NCDS: National Child and Development Study, 1958 birth cohort; NS: Next Steps, 1990 cohort; NSHD: National Survey of Health and Development, 1946 birth cohort; ONS: UK Office for National Statistics; PHQ-2: 2-item Patient Health Questionnaire; UCLA-3: 3-item UCLA loneliness scale. Survey wave 1: May 2020; survey wave 2: September/October 2020; survey wave 3: February/March 2021.

#### Figure S9.1. Unadjusted and adjusted (by birth sex, highest qualification achieved, pre-pandemic self-reported health, pre-pandemic psychological distress, and household composition) anxiety symptomatology (GAD-2) marginal mean estimates and 95% confidence intervals by housing tenure.

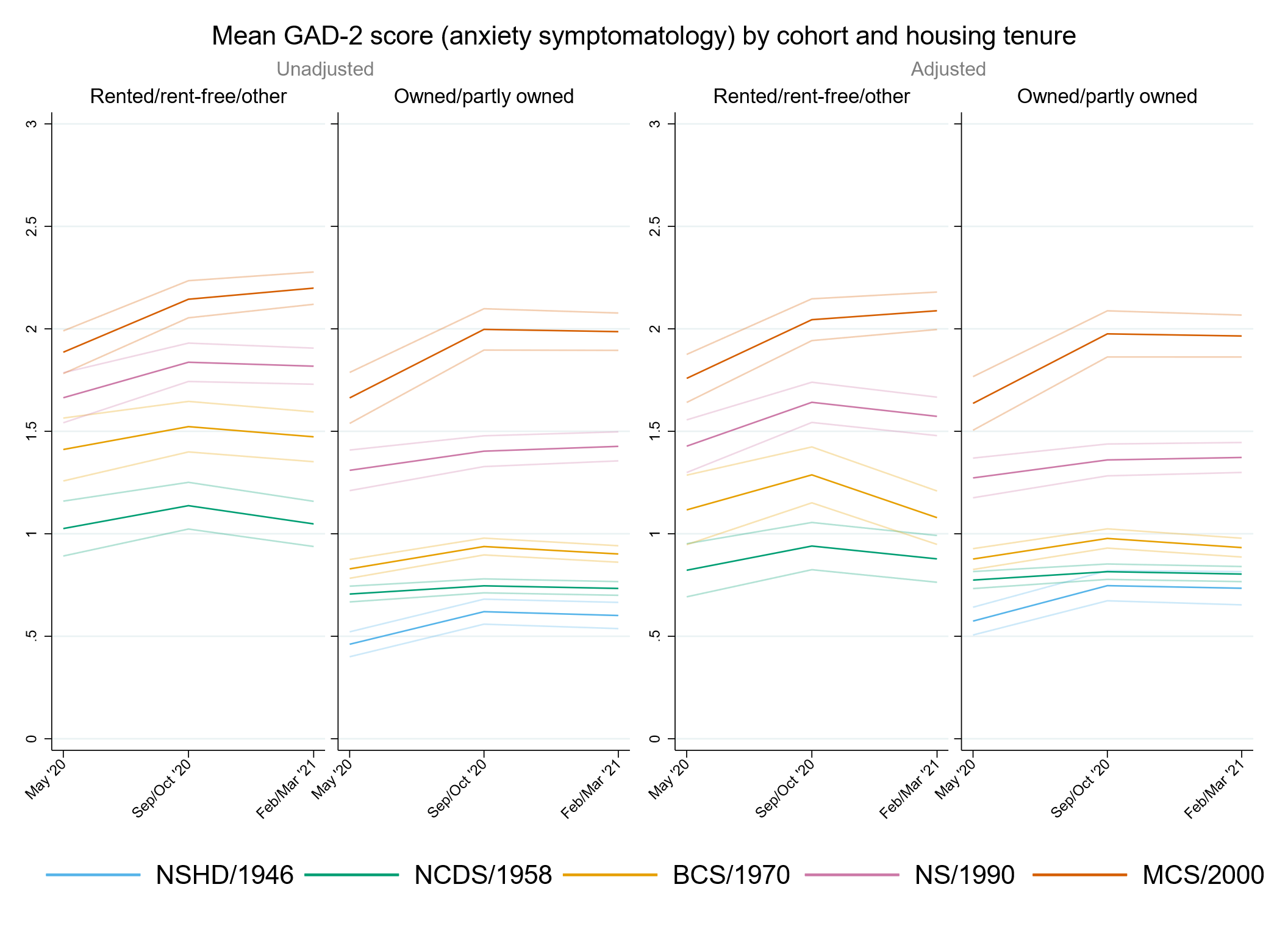

#### Figure S9.2. Unadjusted and adjusted (by birth sex, highest qualification achieved, pre-pandemic self-reported health, pre-pandemic psychological distress, and household composition) depressive symptomatology (PHQ-2) marginal mean estimates and 95% confidence intervals by housing tenure.

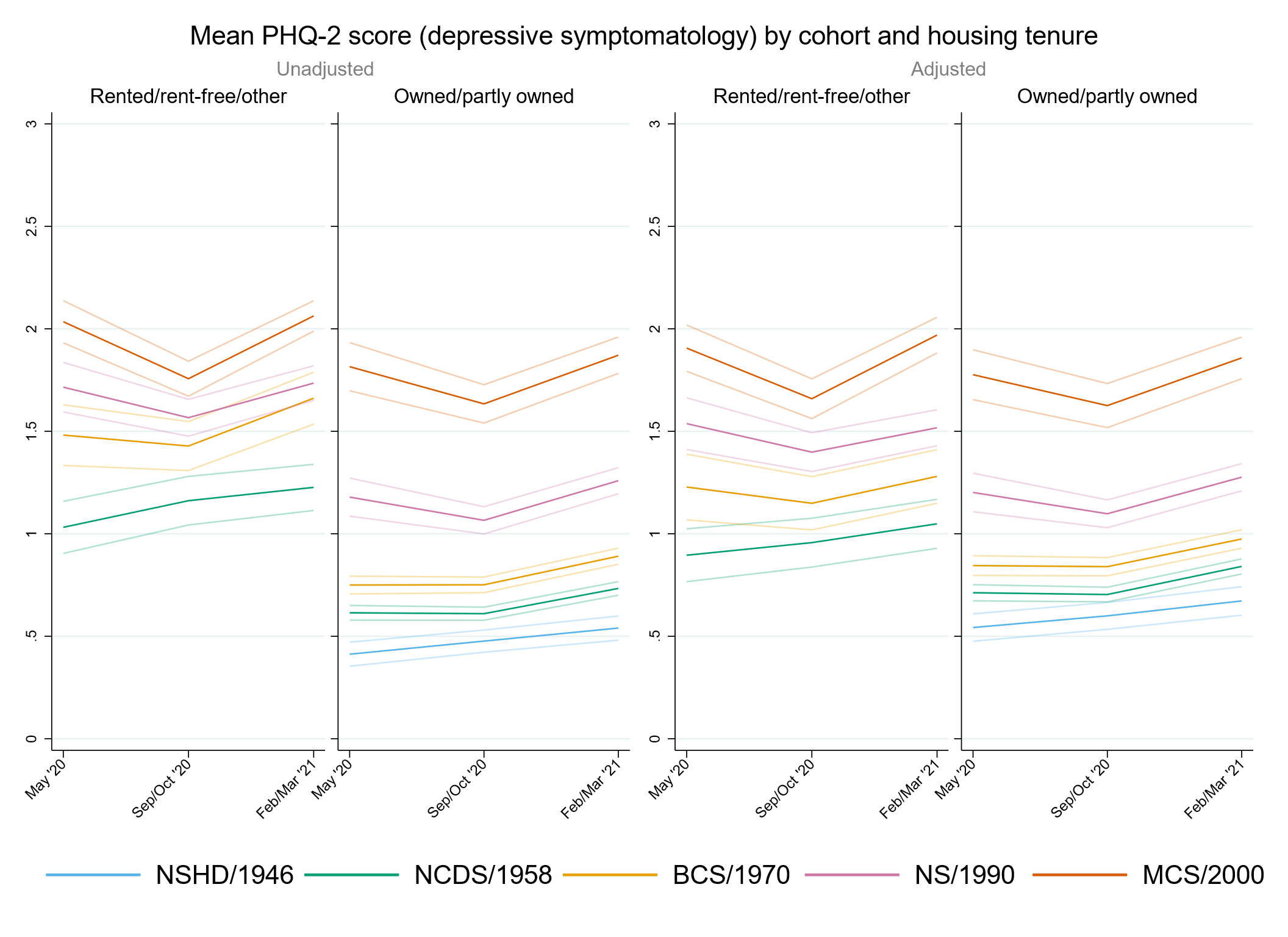

#### Figure S9.3. Unadjusted and adjusted (by birth sex, highest qualification achieved, pre-pandemic self-reported health, pre-pandemic psychological distress, and household composition) loneliness (UCLA-3) marginal mean estimates and 95% confidence intervals by housing tenure.

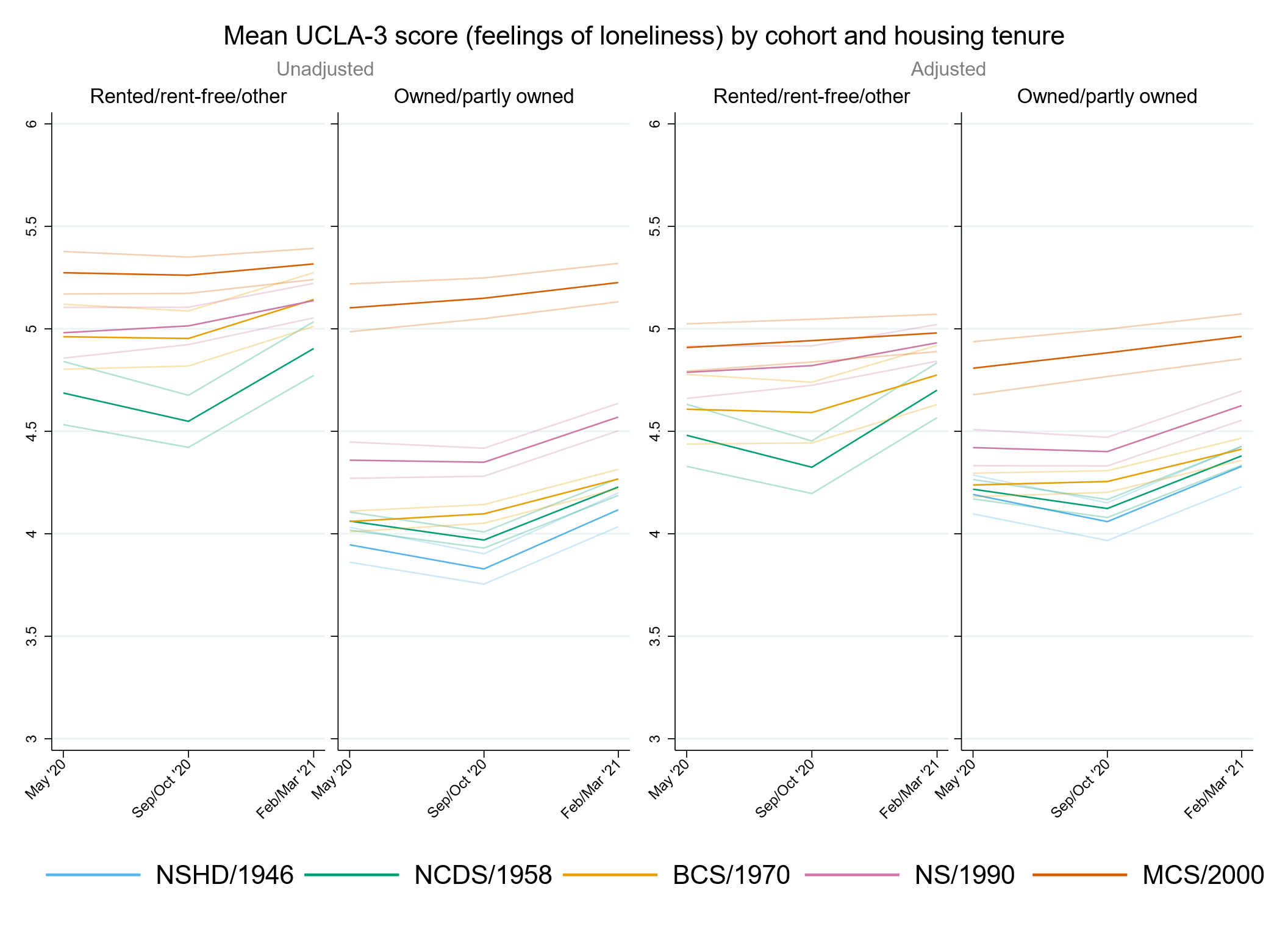

#### Figure S9.4. Unadjusted and adjusted (by birth sex, highest qualification achieved, pre-pandemic self-reported health, pre-pandemic psychological distress, and household composition) life satisfaction marginal mean estimates and 95% confidence intervals by housing tenure.

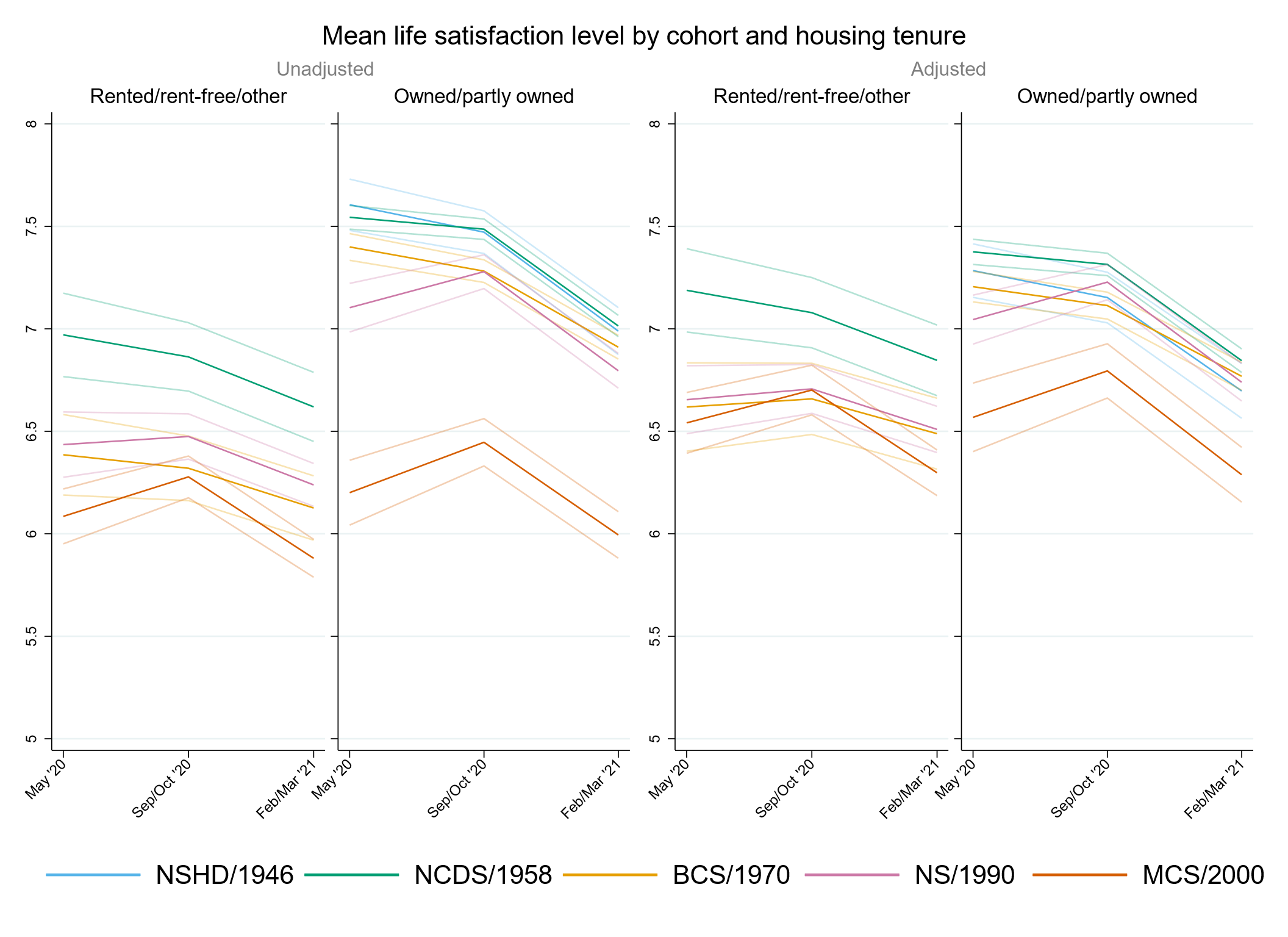

### Appendix S10. Results by urbanicity.

#### Table S10.1. Results of multilevel growth curve models by urbanicity.

|  | **Anxiety symptomatology (GAD-2)** | | | **Depressive symptomatology (PHQ-2)** | | | **Feelings of loneliness (UCLA-3)** | | | **Life satisfaction (ONS single question)** | | |
| --- | --- | --- | --- | --- | --- | --- | --- | --- | --- | --- | --- | --- |
| **Unadjusted models** |  |  |  |  |  |  |  |  |  |  |  |  |
| N participants | 24,458 |  |  | 24,456 |  |  | 24,481 |  |  | 24,533 |  |  |
| N observations | 52,062 |  |  | 52,045 |  |  | 52,123 |  |  | 52,344 |  |  |
|  | ***B* (95% CI)** | **χ2** | ***p*** | ***B* (95% CI)** | **χ2** | ***p*** | ***B* (95% CI)** | **χ2** | ***p*** | ***B* (95% CI)** | **χ2** | ***p*** |
| Time (linear) | 0.08 (0.01, 0.15) |  | 0.028 | -0.05 (-0.12, 0.01) |  | 0.117 | -0.27 (-0.35, -0.20) |  | <0.001 | 0.16 (0.06, 0.26) |  | 0.002 |
| Time (quad) | -0.03 (-0.06, 0.00) |  | 0.063 | 0.06 (0.03, 0.09) |  | <0.001 | 0.18 (0.14, 0.21) |  | <0.001 | -0.20 (-0.25, -0.15) |  | <0.001 |
| Cohort (ref. NCDS) |  | 782 | <0.001 |  | 1043.6 | <0.001 |  | 571.2 | <0.001 |  | 516.9 | <0.001 |
| NSHD | -0.24 (-0.33, -0.16) |  | <0.001 | -0.21 (-0.29, -0.13) |  | <0.001 | -0.14 (-0.25, -0.02) |  | 0.020 | 0.13 (-0.04, 0.29) |  | 0.131 |
| BCS | 0.19 (0.13, 0.26) |  | <0.001 | 0.23 (0.16, 0.29) |  | <0.001 | 0.09 (0.01, 0.16) |  | 0.022 | -0.27 (-0.36, -0.17) |  | <0.001 |
| NS | 0.74 (0.64, 0.83) |  | <0.001 | 0.77 (0.68, 0.85) |  | <0.001 | 0.48 (0.39, 0.58) |  | <0.001 | -0.65 (-0.78, -0.53) |  | <0.001 |
| MCS | 1.03 (0.94, 1.13) |  | <0.001 | 1.24 (1.15, 1.33) |  | <0.001 | 1.03 (0.94, 1.13) |  | <0.001 | -1.36 (-1.49, -1.23) |  | <0.001 |
| Linear change * cohort (ref. NCDS) |  | 17.8 | 0.001 |  | 45.4 | <0.001 |  | 33.2 | <0.001 |  | 33.6 | <0.001 |
| NSHD | 0.12 (-0.02, 0.26) |  | 0.091 | 0.07 (-0.07, 0.21) |  | 0.301 | -0.12 (-0.29, 0.06) |  | 0.194 | -0.20 (-0.47, 0.07) |  | 0.149 |
| BCS | 0.13 (0.01, 0.24) |  | 0.031 | -0.01 (-0.12, 0.09) |  | 0.803 | 0.26 (0.14, 0.38) |  | <0.001 | -0.16 (-0.31, 0.00) |  | 0.044 |
| NS | 0.11 (-0.05, 0.26) |  | 0.166 | -0.27 (-0.41, -0.12) |  | <0.001 | 0.24 (0.09, 0.39) |  | 0.002 | 0.20 (0.00, 0.39) |  | 0.046 |
| MCS | 0.35 (0.18, 0.53) |  | <0.001 | -0.47 (-0.63, -0.30) |  | <0.001 | 0.26 (0.09, 0.43) |  | 0.003 | 0.43 (0.21, 0.66) |  | <0.001 |
| Quad. change * cohort (ref. NCDS) |  | 8.7 | 0.071 |  | 47.5 | <0.001 |  | 38.2 | <0.001 |  | 23.3 | <0.001 |
| NSHD | -0.04 (-0.11, 0.03) |  | 0.245 | -0.04 (-0.11, 0.02) |  | 0.191 | 0.05 (-0.03, 0.13) |  | 0.201 | 0.05 (-0.07, 0.18) |  | 0.427 |
| BCS | -0.05 (-0.10, 0.00) |  | 0.060 | 0.01 (-0.04, 0.06) |  | 0.624 | -0.12 (-0.17, -0.06) |  | <0.001 | 0.09 (0.02, 0.16) |  | 0.014 |
| NS | -0.03 (-0.10, 0.04) |  | 0.450 | 0.11 (0.05, 0.18) |  | 0.001 | -0.11 (-0.18, -0.04) |  | 0.001 | -0.05 (-0.13, 0.04) |  | 0.271 |
| MCS | -0.11 (-0.19, -0.03) |  | 0.008 | 0.23 (0.15, 0.31) |  | <0.001 | -0.16 (-0.23, -0.08) |  | <0.001 | -0.14 (-0.24, -0.04) |  | 0.005 |
| Urbanicity: rural (ref. Urban) | -0.03 (-0.11, 0.05) |  | 0.464 | -0.08 (-0.16, -0.01) |  | 0.031 | -0.04 (-0.13, 0.05) |  | 0.366 | 0.15 (0.03, 0.27) |  | 0.013 |
| Linear change * rural (ref. Urban) | 0.02 (-0.10, 0.15) |  | 0.704 | 0.04 (-0.08, 0.15) |  | 0.531 | -0.03 (-0.16, 0.11) |  | 0.690 | -0.15 (-0.35, 0.04) |  | 0.114 |
| Quad. change * rural (ref. Urban) | -0.01 (-0.07, 0.05) |  | 0.674 | -0.01 (-0.06, 0.04) |  | 0.693 | 0.02 (-0.04, 0.08) |  | 0.511 | 0.05 (-0.04, 0.13) |  | 0.312 |
| Cohort (ref. NCDS) * rural (ref. Urban) |  | 1.5 | 0.825 |  | 0.9 | 0.926 |  | 1.0 | 0.910 |  | 4.4 | 0.353 |
| NSHD | -0.03 (-0.18, 0.12) |  | 0.725 | 0.01 (-0.13, 0.15) |  | 0.892 | -0.07 (-0.26, 0.13) |  | 0.491 | -0.10 (-0.38, 0.18) |  | 0.486 |
| BCS | 0.02 (-0.11, 0.15) |  | 0.755 | 0.00 (-0.12, 0.12) |  | 0.982 | -0.06 (-0.20, 0.08) |  | 0.375 | 0.05 (-0.13, 0.23) |  | 0.599 |
| NS | -0.08 (-0.29, 0.13) |  | 0.465 | -0.09 (-0.30, 0.12) |  | 0.383 | -0.03 (-0.24, 0.19) |  | 0.795 | 0.18 (-0.09, 0.44) |  | 0.202 |
| MCS | -0.07 (-0.26, 0.11) |  | 0.440 | -0.02 (-0.20, 0.16) |  | 0.865 | -0.02 (-0.21, 0.17) |  | 0.830 | 0.18 (-0.07, 0.42) |  | 0.154 |
| Linear change * cohort (ref. NCDS) * rural (ref. Urban) |  | 2.4 | 0.666 |  | 6.4 | 0.173 |  | 4.1 | 0.394 |  | 7.0 | 0.138 |
| NSHD | 0.11 (-0.16, 0.37) |  | 0.429 | 0.01 (-0.22, 0.24) |  | 0.929 | 0.13 (-0.17, 0.43) |  | 0.386 | 0.45 (-0.02, 0.91) |  | 0.059 |
| BCS | -0.10 (-0.32, 0.11) |  | 0.355 | -0.18 (-0.38, 0.02) |  | 0.081 | -0.14 (-0.36, 0.08) |  | 0.223 | 0.22 (-0.07, 0.52) |  | 0.143 |
| NS | 0.05 (-0.32, 0.41) |  | 0.805 | 0.14 (-0.22, 0.50) |  | 0.444 | -0.08 (-0.42, 0.26) |  | 0.659 | 0.01 (-0.42, 0.45) |  | 0.951 |
| MCS | -0.08 (-0.42, 0.26) |  | 0.635 | 0.21 (-0.15, 0.57) |  | 0.258 | 0.14 (-0.21, 0.49) |  | 0.432 | -0.20 (-0.66, 0.25) |  | 0.385 |
| Quad. change * cohort (ref. NCDS) * rural (ref. Urban) |  | 2.9 | 0.572 |  | 6.7 | 0.154 |  | 3.6 | 0.457 |  | 4.5 | 0.340 |
| NSHD | -0.06 (-0.18, 0.06) |  | 0.332 | -0.01 (-0.12, 0.09) |  | 0.793 | -0.06 (-0.20, 0.08) |  | 0.386 | -0.14 (-0.36, 0.08) |  | 0.206 |
| BCS | 0.03 (-0.06, 0.13) |  | 0.500 | 0.07 (-0.02, 0.16) |  | 0.142 | 0.06 (-0.04, 0.16) |  | 0.244 | -0.08 (-0.22, 0.05) |  | 0.234 |
| NS | -0.02 (-0.19, 0.14) |  | 0.804 | -0.04 (-0.20, 0.12) |  | 0.610 | 0.03 (-0.11, 0.18) |  | 0.657 | 0.00 (-0.20, 0.19) |  | 0.978 |
| MCS | 0.07 (-0.08, 0.23) |  | 0.364 | -0.14 (-0.31, 0.03) |  | 0.097 | -0.05 (-0.21, 0.11) |  | 0.533 | 0.10 (-0.11, 0.31) |  | 0.347 |
| **Adjusted models** |  |  |  |  |  |  |  |  |  |  |  |  |
| N participants | 21,008 |  |  | 21,006 |  |  | 21,027 |  |  | 21,067 |  |  |
| N observations | 45,754 |  |  | 45,742 |  |  | 45,799 |  |  | 45,989 |  |  |
|  | ***B* (95% CI)** | **χ2** | ***p*** | ***B* (95% CI)** | **χ2** | ***p*** | ***B* (95% CI)** | **χ2** | ***p*** | ***B* (95% CI)** | **χ2** | ***p*** |
| Time (linear) | 0.08 (0.01, 0.16) |  | 0.024 | -0.08 (-0.15, -0.01) |  | 0.020 | -0.28 (-0.36, -0.20) |  | <0.001 | 0.18 (0.07, 0.29) |  | 0.001 |
| Time (quad) | -0.03 (-0.07, 0.00) |  | 0.057 | 0.07 (0.04, 0.10) |  | <0.001 | 0.18 (0.14, 0.22) |  | <0.001 | -0.21 (-0.26, -0.16) |  | <0.001 |
| Cohort (ref. NCDS) |  | 425.7 | <0.001 |  | 572.0 | <0.001 |  | 131.2 | <0.001 |  | 140.9 | <0.001 |
| NSHD | -0.21 (-0.29, -0.12) |  | <0.001 | -0.18 (-0.27, -0.10) |  | <0.001 | -0.03 (-0.14, 0.09) |  | 0.670 | -0.01 (-0.17, 0.15) |  | 0.865 |
| BCS | 0.13 (0.06, 0.20) |  | <0.001 | 0.17 (0.10, 0.23) |  | <0.001 | 0.05 (-0.02, 0.13) |  | 0.171 | -0.22 (-0.32, -0.12) |  | <0.001 |
| NS | 0.58 (0.49, 0.67) |  | <0.001 | 0.63 (0.54, 0.72) |  | <0.001 | 0.32 (0.23, 0.41) |  | <0.001 | -0.44 (-0.57, -0.32) |  | <0.001 |
| MCS | 0.91 (0.80, 1.02) |  | <0.001 | 1.07 (0.96, 1.17) |  | <0.001 | 0.58 (0.46, 0.69) |  | <0.001 | -0.82 (-0.97, -0.67) |  | <0.001 |
| Linear change * cohort (ref. NCDS) |  | 20.1 | <0.001 |  | 28.9 | <0.001 |  | 33.3 | <0.001 |  | 27.9 | <0.001 |
| NSHD | 0.15 (0.00, 0.30) |  | 0.053 | 0.12 (-0.04, 0.27) |  | 0.139 | -0.13 (-0.32, 0.06) |  | 0.193 | -0.27 (-0.56, 0.02) |  | 0.072 |
| BCS | 0.15 (0.03, 0.27) |  | 0.015 | 0.03 (-0.08, 0.15) |  | 0.590 | 0.26 (0.14, 0.39) |  | <0.001 | -0.16 (-0.33, 0.01) |  | 0.060 |
| NS | 0.15 (-0.02, 0.32) |  | 0.078 | -0.19 (-0.35, -0.03) |  | 0.018 | 0.24 (0.08, 0.40) |  | 0.003 | 0.18 (-0.03, 0.38) |  | 0.097 |
| MCS | 0.38 (0.20, 0.57) |  | <0.001 | -0.39 (-0.57, -0.20) |  | <0.001 | 0.32 (0.14, 0.51) |  | 0.001 | 0.41 (0.17, 0.64) |  | 0.001 |
| Quad. change * cohort (ref. NCDS) |  | 11.2 | 0.025 |  | 32.0 | <0.001 |  | 37.3 | <0.001 |  | 20.9 | <0.001 |
| NSHD | -0.04 (-0.12, 0.03) |  | 0.274 | -0.07 (-0.14, 0.01) |  | 0.080 | 0.05 (-0.04, 0.14) |  | 0.290 | 0.09 (-0.05, 0.23) |  | 0.206 |
| BCS | -0.07 (-0.13, -0.01) |  | 0.014 | -0.02 (-0.07, 0.04) |  | 0.540 | -0.13 (-0.19, -0.07) |  | <0.001 | 0.10 (0.02, 0.18) |  | 0.014 |
| NS | -0.05 (-0.13, 0.02) |  | 0.175 | 0.07 (0.00, 0.14) |  | 0.050 | -0.11 (-0.18, -0.04) |  | 0.002 | -0.04 (-0.13, 0.06) |  | 0.453 |
| MCS | -0.12 (-0.20, -0.03) |  | 0.006 | 0.20 (0.11, 0.28) |  | <0.001 | -0.18 (-0.27, -0.09) |  | <0.001 | -0.14 (-0.25, -0.03) |  | 0.010 |
| Urbanicity: rural (ref. Urban) | -0.02 (-0.09, 0.06) |  | 0.631 | -0.05 (-0.12, 0.02) |  | 0.154 | 0.01 (-0.08, 0.09) |  | 0.862 | 0.09 (-0.02, 0.21) |  | 0.106 |
| Linear change * rural (ref. Urban) | 0.02 (-0.11, 0.15) |  | 0.770 | 0.07 (-0.06, 0.19) |  | 0.306 | -0.03 (-0.17, 0.11) |  | 0.675 | -0.18 (-0.38, 0.01) |  | 0.066 |
| Quad. change * rural (ref. Urban) | -0.01 (-0.07, 0.05) |  | 0.804 | -0.02 (-0.08, 0.04) |  | 0.432 | 0.02 (-0.05, 0.09) |  | 0.575 | 0.07 (-0.02, 0.16) |  | 0.152 |
| Cohort (ref. NCDS) * rural (ref. Urban) |  | 3.9 | 0.415 |  | 4.0 | 0.406 |  | 1.9 | 0.751 |  | 7.2 | 0.126 |
| NSHD | 0.07 (-0.09, 0.22) |  | 0.401 | 0.04 (-0.11, 0.18) |  | 0.620 | -0.08 (-0.29, 0.12) |  | 0.430 | -0.14 (-0.43, 0.15) |  | 0.336 |
| BCS | 0.04 (-0.08, 0.17) |  | 0.517 | 0.07 (-0.05, 0.19) |  | 0.238 | 0.00 (-0.14, 0.14) |  | 0.968 | 0.01 (-0.17, 0.19) |  | 0.898 |
| NS | -0.10 (-0.29, 0.10) |  | 0.347 | -0.09 (-0.28, 0.11) |  | 0.387 | 0.04 (-0.16, 0.24) |  | 0.694 | 0.13 (-0.13, 0.39) |  | 0.340 |
| MCS | -0.10 (-0.29, 0.09) |  | 0.288 | -0.07 (-0.25, 0.10) |  | 0.405 | -0.10 (-0.29, 0.09) |  | 0.321 | 0.26 (0.02, 0.50) |  | 0.033 |
| Linear change * cohort (ref. NCDS) * rural (ref. Urban) |  | 1.4 | 0.849 |  | 8.9 | 0.065 |  | 6.2 | 0.184 |  | 10.0 | 0.041 |
| NSHD | 0.10 (-0.21, 0.41) |  | 0.534 | -0.02 (-0.28, 0.25) |  | 0.904 | 0.16 (-0.20, 0.52) |  | 0.385 | 0.64 (0.10, 1.17) |  | 0.019 |
| BCS | -0.10 (-0.34, 0.14) |  | 0.430 | -0.26 (-0.49, -0.04) |  | 0.021 | -0.17 (-0.41, 0.07) |  | 0.161 | 0.27 (-0.04, 0.58) |  | 0.090 |
| NS | -0.02 (-0.39, 0.35) |  | 0.924 | 0.04 (-0.32, 0.40) |  | 0.832 | -0.16 (-0.50, 0.18) |  | 0.349 | 0.05 (-0.40, 0.51) |  | 0.824 |
| MCS | -0.06 (-0.42, 0.31) |  | 0.765 | 0.25 (-0.12, 0.63) |  | 0.185 | 0.23 (-0.15, 0.60) |  | 0.243 | -0.25 (-0.73, 0.22) |  | 0.299 |
| Quad. change * cohort (ref. NCDS) * rural (ref. Urban) |  | 2.3 | 0.678 |  | 9.0 | 0.062 |  | 5.1 | 0.277 |  | 7.2 | 0.125 |
| NSHD | -0.08 (-0.23, 0.07) |  | 0.302 | 0.00 (-0.12, 0.13) |  | 0.938 | -0.07 (-0.24, 0.11) |  | 0.448 | -0.26 (-0.55, 0.02) |  | 0.065 |
| BCS | 0.03 (-0.08, 0.14) |  | 0.558 | 0.11 (0.00, 0.21) |  | 0.045 | 0.08 (-0.03, 0.19) |  | 0.168 | -0.11 (-0.25, 0.04) |  | 0.142 |
| NS | 0.01 (-0.16, 0.18) |  | 0.880 | 0.00 (-0.16, 0.16) |  | 0.995 | 0.07 (-0.08, 0.22) |  | 0.365 | -0.02 (-0.23, 0.18) |  | 0.816 |
| MCS | 0.06 (-0.11, 0.23) |  | 0.485 | -0.16 (-0.33, 0.02) |  | 0.075 | -0.08 (-0.26, 0.10) |  | 0.365 | 0.11 (-0.11, 0.33) |  | 0.313 |
| **Sensitivity models** |  |  |  |  |  |  |  |  |  |  |  |  |
| N participants | 21,008 |  |  | 21,006 |  |  | 21,027 |  |  | 21,067 |  |  |
| N observations | 45,754 |  |  | 45,742 |  |  | 45,799 |  |  | 45,989 |  |  |
|  | ***B* (95% CI)** | **χ2** | ***p*** | ***B* (95% CI)** | **χ2** | ***p*** | ***B* (95% CI)** | **χ2** | ***p*** | ***B* (95% CI)** | **χ2** | ***p*** |
| Time (linear) | 0.09 (0.02, 0.16) |  | 0.014 | -0.07 (-0.14, 0.00) |  | 0.036 | -0.26 (-0.34, -0.18) |  | <0.001 | 0.17 (0.06, 0.27) |  | 0.002 |
| Time (quad) | -0.03 (-0.07, 0.00) |  | 0.043 | 0.07 (0.04, 0.10) |  | <0.001 | 0.17 (0.14, 0.21) |  | <0.001 | -0.20 (-0.26, -0.15) |  | <0.001 |
| Cohort (ref. NCDS) |  | 752.1 | <0.001 |  | 930.7 | <0.001 |  | 508.6 | <0.001 |  | 460.1 | <0.001 |
| NSHD | -0.27 (-0.35, -0.18) |  | <0.001 | -0.23 (-0.32, -0.15) |  | <0.001 | -0.13 (-0.25, 0.00) |  | 0.045 | 0.13 (-0.03, 0.30) |  | 0.121 |
| BCS | 0.18 (0.11, 0.25) |  | <0.001 | 0.20 (0.13, 0.26) |  | <0.001 | 0.06 (-0.02, 0.14) |  | 0.156 | -0.25 (-0.35, -0.15) |  | <0.001 |
| NS | 0.75 (0.65, 0.84) |  | <0.001 | 0.75 (0.66, 0.85) |  | <0.001 | 0.48 (0.39, 0.58) |  | <0.001 | -0.65 (-0.78, -0.52) |  | <0.001 |
| MCS | 1.05 (0.96, 1.15) |  | <0.001 | 1.22 (1.13, 1.32) |  | <0.001 | 1.03 (0.93, 1.13) |  | <0.001 | -1.33 (-1.46, -1.20) |  | <0.001 |
| Linear change * cohort (ref. NCDS) |  | 16.9 | 0.002 |  | 30.4 | <0.001 |  | 30.6 | <0.001 |  | 27.2 | <0.001 |
| NSHD | 0.14 (-0.01, 0.30) |  | 0.066 | 0.11 (-0.04, 0.27) |  | 0.148 | -0.13 (-0.32, 0.06) |  | 0.184 | -0.26 (-0.55, 0.04) |  | 0.085 |
| BCS | 0.14 (0.02, 0.26) |  | 0.026 | 0.02 (-0.09, 0.14) |  | 0.680 | 0.25 (0.12, 0.38) |  | <0.001 | -0.14 (-0.31, 0.03) |  | 0.098 |
| NS | 0.13 (-0.03, 0.30) |  | 0.121 | -0.20 (-0.36, -0.04) |  | 0.012 | 0.24 (0.08, 0.41) |  | 0.003 | 0.19 (-0.02, 0.40) |  | 0.074 |
| MCS | 0.36 (0.17, 0.54) |  | <0.001 | -0.41 (-0.60, -0.22) |  | <0.001 | 0.30 (0.12, 0.49) |  | 0.002 | 0.42 (0.18, 0.66) |  | 0.001 |
| Quad. change * cohort (ref. NCDS) |  | 9.7 | 0.046 |  | 32.2 | <0.001 |  | 35.3 | <0.001 |  | 18.9 | <0.001 |
| NSHD | -0.04 (-0.12, 0.04) |  | 0.288 | -0.07 (-0.14, 0.01) |  | 0.073 | 0.05 (-0.04, 0.14) |  | 0.301 | 0.09 (-0.05, 0.24) |  | 0.206 |
| BCS | -0.06 (-0.12, -0.01) |  | 0.025 | -0.01 (-0.07, 0.04) |  | 0.648 | -0.12 (-0.18, -0.06) |  | <0.001 | 0.09 (0.01, 0.17) |  | 0.024 |
| NS | -0.05 (-0.12, 0.03) |  | 0.237 | 0.07 (0.00, 0.14) |  | 0.040 | -0.11 (-0.19, -0.04) |  | 0.002 | -0.04 (-0.13, 0.05) |  | 0.404 |
| MCS | -0.11 (-0.20, -0.03) |  | 0.010 | 0.20 (0.11, 0.29) |  | <0.001 | -0.18 (-0.26, -0.09) |  | <0.001 | -0.14 (-0.24, -0.03) |  | 0.014 |
| Urbanicity: rural (ref. Urban) | -0.04 (-0.12, 0.04) |  | 0.309 | -0.09 (-0.17, -0.02) |  | 0.015 | -0.04 (-0.14, 0.05) |  | 0.353 | 0.14 (0.02, 0.26) |  | 0.022 |
| Linear change * rural (ref. Urban) | 0.02 (-0.12, 0.15) |  | 0.826 | 0.06 (-0.06, 0.19) |  | 0.329 | -0.04 (-0.18, 0.11) |  | 0.615 | -0.18 (-0.38, 0.02) |  | 0.078 |
| Quad. change * rural (ref. Urban) | -0.01 (-0.07, 0.06) |  | 0.863 | -0.02 (-0.08, 0.04) |  | 0.475 | 0.02 (-0.04, 0.09) |  | 0.511 | 0.06 (-0.03, 0.16) |  | 0.177 |
| Cohort (ref. NCDS) * rural (ref. Urban) |  | 4.6 | 0.333 |  | 4.4 | 0.358 |  | 0.7 | 0.956 |  | 8.1 | 0.090 |
| NSHD | 0.09 (-0.07, 0.26) |  | 0.263 | 0.09 (-0.06, 0.24) |  | 0.253 | -0.03 (-0.24, 0.19) |  | 0.803 | -0.20 (-0.50, 0.11) |  | 0.205 |
| BCS | 0.01 (-0.12, 0.14) |  | 0.862 | 0.04 (-0.09, 0.17) |  | 0.525 | -0.05 (-0.19, 0.10) |  | 0.532 | 0.06 (-0.13, 0.25) |  | 0.531 |
| NS | -0.14 (-0.35, 0.07) |  | 0.201 | -0.12 (-0.33, 0.09) |  | 0.250 | -0.03 (-0.24, 0.19) |  | 0.798 | 0.22 (-0.05, 0.50) |  | 0.114 |
| MCS | -0.09 (-0.29, 0.10) |  | 0.357 | -0.07 (-0.25, 0.12) |  | 0.476 | -0.07 (-0.27, 0.13) |  | 0.497 | 0.23 (-0.02, 0.48) |  | 0.075 |
| Linear change * cohort (ref. NCDS) * rural (ref. Urban) |  | 1.2 | 0.880 |  | 10.0 | 0.040 |  | 6.2 | 0.185 |  | 10.5 | 0.032 |
| NSHD | 0.09 (-0.22, 0.41) |  | 0.558 | -0.03 (-0.29, 0.24) |  | 0.846 | 0.15 (-0.21, 0.51) |  | 0.418 | 0.64 (0.11, 1.18) |  | 0.019 |
| BCS | -0.09 (-0.33, 0.15) |  | 0.479 | -0.27 (-0.49, -0.04) |  | 0.021 | -0.16 (-0.41, 0.08) |  | 0.179 | 0.26 (-0.06, 0.57) |  | 0.106 |
| NS | 0.04 (-0.34, 0.41) |  | 0.844 | 0.09 (-0.28, 0.46) |  | 0.638 | -0.13 (-0.47, 0.22) |  | 0.470 | -0.01 (-0.47, 0.45) |  | 0.967 |
| MCS | 0.01 (-0.36, 0.38) |  | 0.957 | 0.30 (-0.08, 0.68) |  | 0.120 | 0.27 (-0.11, 0.65) |  | 0.170 | -0.29 (-0.77, 0.19) |  | 0.229 |
| Quad. change * cohort (ref. NCDS) * rural (ref. Urban) |  | 1.7 | 0.798 |  | 9.9 | 0.043 |  | 5.0 | 0.288 |  | 7.4 | 0.117 |
| NSHD | -0.08 (-0.23, 0.08) |  | 0.321 | 0.01 (-0.12, 0.14) |  | 0.879 | -0.06 (-0.24, 0.11) |  | 0.471 | -0.27 (-0.55, 0.02) |  | 0.065 |
| BCS | 0.03 (-0.09, 0.14) |  | 0.632 | 0.11 (0.00, 0.21) |  | 0.052 | 0.07 (-0.04, 0.18) |  | 0.201 | -0.10 (-0.24, 0.05) |  | 0.186 |
| NS | -0.01 (-0.18, 0.16) |  | 0.909 | -0.02 (-0.18, 0.14) |  | 0.810 | 0.06 (-0.10, 0.21) |  | 0.466 | 0.00 (-0.21, 0.21) |  | 0.996 |
| MCS | 0.03 (-0.14, 0.20) |  | 0.742 | -0.18 (-0.36, -0.01) |  | 0.043 | -0.10 (-0.28, 0.08) |  | 0.264 | 0.13 (-0.09, 0.35) |  | 0.248 |

*Note.* Adjusted models included birth sex, highest qualification achieved, pre-pandemic self-reported health, pre-pandemic psychological distress, and household composition as covariates. Sensitivity models correspond to the unadjusted models after restricting the analytical sample to that of the adjusted models. BCS: British Cohort Study, 1970 birth cohort; GAD-2: 2-item General Anxiety Disorder questionnaire; MCS: Millennium Cohort Study, 2000 birth cohort; NCDS: National Child and Development Study, 1958 birth cohort; NS: Next Steps, 1990 cohort; NSHD: National Survey of Health and Development, 1946 birth cohort; ONS: UK Office for National Statistics; PHQ-2: 2-item Patient Health Questionnaire; UCLA-3: 3-item UCLA loneliness scale. χ2: Wald test performed to assess the overall statistical significance of the interaction terms; all χ2 statistics in this table have 4 degrees of freedom.

#### Table S10.2. Unadjusted and adjusted marginal mean estimates and 95% confidence intervals by urbanicity.

|  |  |  | **Anxiety symptomatology (GAD-2)** | **Depressive symptomatology (PHQ-2)** | **Feelings of loneliness (UCLA-3)** | **Life satisfaction (ONS single question)** |
| --- | --- | --- | --- | --- | --- | --- |
| Cohort | Urbanicity | Survey wave | Unadjusted marginal mean (95% CI) | Unadjusted marginal mean (95% CI) | Unadjusted marginal mean (95% CI) | Unadjusted marginal mean (95% CI) |
| NSHD | Urban | 1 | 0.51 (0.44, 0.59) | 0.48 (0.41, 0.56) | 4.02 (3.92, 4.12) | 7.54 (7.39, 7.69) |
| NSHD | Urban | 2 | 0.64 (0.57, 0.72) | 0.52 (0.45, 0.59) | 3.86 (3.77, 3.95) | 7.36 (7.23, 7.49) |
| NSHD | Urban | 3 | 0.63 (0.55, 0.71) | 0.58 (0.51, 0.65) | 4.16 (4.07, 4.26) | 6.87 (6.73, 7.01) |
| NSHD | Rural | 1 | 0.46 (0.35, 0.56) | 0.41 (0.32, 0.50) | 3.91 (3.77, 4.05) | 7.60 (7.39, 7.80) |
| NSHD | Rural | 2 | 0.64 (0.54, 0.75) | 0.47 (0.38, 0.56) | 3.82 (3.69, 3.94) | 7.61 (7.44, 7.78) |
| NSHD | Rural | 3 | 0.55 (0.45, 0.64) | 0.50 (0.41, 0.59) | 4.11 (3.97, 4.24) | 7.13 (6.94, 7.32) |
| NCDS | Urban | 1 | 0.76 (0.71, 0.80) | 0.69 (0.65, 0.73) | 4.16 (4.11, 4.21) | 7.42 (7.35, 7.48) |
| NCDS | Urban | 2 | 0.80 (0.76, 0.84) | 0.70 (0.66, 0.74) | 4.06 (4.01, 4.11) | 7.38 (7.32, 7.44) |
| NCDS | Urban | 3 | 0.79 (0.75, 0.83) | 0.82 (0.78, 0.86) | 4.32 (4.27, 4.37) | 6.94 (6.88, 7.00) |
| NCDS | Rural | 1 | 0.73 (0.66, 0.79) | 0.61 (0.55, 0.67) | 4.11 (4.04, 4.19) | 7.57 (7.47, 7.67) |
| NCDS | Rural | 2 | 0.79 (0.72, 0.85) | 0.64 (0.59, 0.70) | 4.01 (3.94, 4.08) | 7.42 (7.33, 7.52) |
| NCDS | Rural | 3 | 0.76 (0.70, 0.82) | 0.77 (0.71, 0.83) | 4.31 (4.23, 4.38) | 6.97 (6.88, 7.06) |
| BCS70 | Urban | 1 | 0.95 (0.90, 1.00) | 0.92 (0.87, 0.97) | 4.24 (4.19, 4.30) | 7.15 (7.08, 7.22) |
| BCS70 | Urban | 2 | 1.07 (1.02, 1.12) | 0.93 (0.88, 0.97) | 4.28 (4.23, 4.34) | 7.04 (6.98, 7.11) |
| BCS70 | Urban | 3 | 1.03 (0.99, 1.08) | 1.07 (1.02, 1.12) | 4.44 (4.39, 4.49) | 6.72 (6.66, 6.78) |
| BCS70 | Rural | 1 | 0.94 (0.85, 1.02) | 0.84 (0.76, 0.92) | 4.14 (4.05, 4.23) | 7.35 (7.24, 7.47) |
| BCS70 | Rural | 2 | 1.01 (0.93, 1.09) | 0.76 (0.69, 0.84) | 4.09 (4.01, 4.18) | 7.28 (7.17, 7.38) |
| BCS70 | Rural | 3 | 0.96 (0.88, 1.03) | 0.94 (0.87, 1.01) | 4.33 (4.25, 4.42) | 6.91 (6.81, 7.01) |
| NS | Urban | 1 | 1.49 (1.41, 1.57) | 1.46 (1.38, 1.54) | 4.64 (4.56, 4.72) | 6.76 (6.66, 6.86) |
| NS | Urban | 2 | 1.62 (1.56, 1.69) | 1.31 (1.25, 1.37) | 4.67 (4.61, 4.73) | 6.87 (6.80, 6.95) |
| NS | Urban | 3 | 1.64 (1.58, 1.70) | 1.50 (1.44, 1.55) | 4.84 (4.79, 4.90) | 6.49 (6.42, 6.56) |
| NS | Rural | 1 | 1.38 (1.20, 1.56) | 1.29 (1.11, 1.46) | 4.57 (4.39, 4.75) | 7.09 (6.87, 7.31) |
| NS | Rural | 2 | 1.55 (1.40, 1.70) | 1.26 (1.13, 1.40) | 4.55 (4.41, 4.70) | 7.11 (6.93, 7.28) |
| NS | Rural | 3 | 1.54 (1.40, 1.68) | 1.47 (1.34, 1.61) | 4.78 (4.64, 4.92) | 6.71 (6.54, 6.88) |
| MCS | Urban | 1 | 1.79 (1.71, 1.87) | 1.93 (1.85, 2.01) | 5.19 (5.11, 5.27) | 6.06 (5.95, 6.17) |
| MCS | Urban | 2 | 2.08 (2.01, 2.16) | 1.70 (1.63, 1.77) | 5.20 (5.13, 5.27) | 6.32 (6.23, 6.40) |
| MCS | Urban | 3 | 2.10 (2.03, 2.16) | 2.04 (1.97, 2.10) | 5.25 (5.19, 5.32) | 5.89 (5.81, 5.97) |
| MCS | Rural | 1 | 1.69 (1.54, 1.83) | 1.84 (1.69, 1.98) | 5.13 (4.98, 5.27) | 6.39 (6.21, 6.57) |
| MCS | Rural | 2 | 1.98 (1.84, 2.13) | 1.70 (1.55, 1.85) | 5.22 (5.07, 5.37) | 6.43 (6.25, 6.61) |
| MCS | Rural | 3 | 2.12 (1.99, 2.25) | 1.83 (1.71, 1.94) | 5.29 (5.16, 5.42) | 6.09 (5.94, 6.24) |
| Cohort | Urbanicity | Survey wave | Adjusted marginal mean (95% CI) | Adjusted marginal mean (95% CI) | Adjusted marginal mean (95% CI) | Adjusted marginal mean (95% CI) |
| NSHD | Urban | 1 | 0.58 (0.50, 0.65) | 0.57 (0.50, 0.65) | 4.23 (4.12, 4.34) | 7.29 (7.14, 7.44) |
| NSHD | Urban | 2 | 0.74 (0.65, 0.82) | 0.61 (0.54, 0.69) | 4.05 (3.95, 4.15) | 7.08 (6.94, 7.23) |
| NSHD | Urban | 3 | 0.74 (0.65, 0.84) | 0.66 (0.59, 0.74) | 4.33 (4.22, 4.44) | 6.65 (6.49, 6.80) |
| NSHD | Rural | 1 | 0.63 (0.51, 0.75) | 0.56 (0.45, 0.66) | 4.15 (3.99, 4.31) | 7.24 (7.02, 7.47) |
| NSHD | Rural | 2 | 0.82 (0.69, 0.95) | 0.63 (0.52, 0.73) | 4.06 (3.90, 4.21) | 7.29 (7.07, 7.51) |
| NSHD | Rural | 3 | 0.68 (0.55, 0.81) | 0.67 (0.56, 0.78) | 4.32 (4.16, 4.48) | 6.71 (6.43, 6.99) |
| NCDS | Urban | 1 | 0.78 (0.74, 0.83) | 0.76 (0.71, 0.80) | 4.25 (4.20, 4.31) | 7.30 (7.24, 7.37) |
| NCDS | Urban | 2 | 0.84 (0.79, 0.88) | 0.74 (0.71, 0.78) | 4.16 (4.11, 4.20) | 7.28 (7.22, 7.34) |
| NCDS | Urban | 3 | 0.82 (0.78, 0.86) | 0.88 (0.84, 0.92) | 4.41 (4.37, 4.46) | 6.83 (6.77, 6.90) |
| NCDS | Rural | 1 | 0.77 (0.70, 0.83) | 0.70 (0.64, 0.77) | 4.26 (4.19, 4.34) | 7.40 (7.30, 7.50) |
| NCDS | Rural | 2 | 0.83 (0.77, 0.89) | 0.73 (0.67, 0.79) | 4.15 (4.08, 4.22) | 7.25 (7.16, 7.34) |
| NCDS | Rural | 3 | 0.81 (0.75, 0.87) | 0.86 (0.80, 0.92) | 4.44 (4.37, 4.51) | 6.83 (6.73, 6.92) |
| BCS70 | Urban | 1 | 0.91 (0.86, 0.97) | 0.92 (0.87, 0.97) | 4.31 (4.25, 4.37) | 7.08 (7.01, 7.16) |
| BCS70 | Urban | 2 | 1.05 (0.99, 1.10) | 0.93 (0.88, 0.98) | 4.34 (4.29, 4.40) | 6.99 (6.92, 7.06) |
| BCS70 | Urban | 3 | 0.97 (0.92, 1.02) | 1.04 (0.99, 1.09) | 4.48 (4.42, 4.54) | 6.68 (6.61, 6.75) |
| BCS70 | Rural | 1 | 0.94 (0.85, 1.03) | 0.94 (0.86, 1.03) | 4.31 (4.22, 4.41) | 7.19 (7.07, 7.31) |
| BCS70 | Rural | 2 | 1.02 (0.93, 1.10) | 0.83 (0.76, 0.91) | 4.25 (4.16, 4.33) | 7.14 (7.04, 7.25) |
| BCS70 | Rural | 3 | 0.94 (0.87, 1.02) | 1.00 (0.93, 1.07) | 4.47 (4.38, 4.56) | 6.80 (6.69, 6.90) |
| NS | Urban | 1 | 1.36 (1.28, 1.45) | 1.39 (1.31, 1.46) | 4.57 (4.50, 4.65) | 6.86 (6.76, 6.97) |
| NS | Urban | 2 | 1.51 (1.44, 1.58) | 1.25 (1.19, 1.32) | 4.61 (4.54, 4.67) | 6.97 (6.89, 7.05) |
| NS | Urban | 3 | 1.49 (1.43, 1.56) | 1.41 (1.35, 1.47) | 4.77 (4.71, 4.83) | 6.60 (6.52, 6.68) |
| NS | Rural | 1 | 1.25 (1.09, 1.42) | 1.25 (1.08, 1.41) | 4.62 (4.46, 4.79) | 7.08 (6.87, 7.29) |
| NS | Rural | 2 | 1.41 (1.26, 1.55) | 1.20 (1.06, 1.33) | 4.55 (4.41, 4.69) | 7.10 (6.93, 7.28) |
| NS | Rural | 3 | 1.40 (1.27, 1.54) | 1.39 (1.25, 1.52) | 4.79 (4.65, 4.93) | 6.73 (6.55, 6.91) |
| MCS | Urban | 1 | 1.69 (1.60, 1.78) | 1.82 (1.73, 1.91) | 4.83 (4.74, 4.92) | 6.49 (6.36, 6.61) |
| MCS | Urban | 2 | 2.01 (1.92, 2.10) | 1.62 (1.54, 1.71) | 4.88 (4.79, 4.97) | 6.73 (6.62, 6.83) |
| MCS | Urban | 3 | 2.02 (1.94, 2.10) | 1.96 (1.88, 2.04) | 4.92 (4.84, 5.00) | 6.27 (6.17, 6.37) |
| MCS | Rural | 1 | 1.57 (1.42, 1.73) | 1.70 (1.55, 1.84) | 4.74 (4.59, 4.90) | 6.84 (6.65, 7.03) |
| MCS | Rural | 2 | 1.90 (1.75, 2.06) | 1.63 (1.48, 1.79) | 4.92 (4.76, 5.08) | 6.82 (6.64, 7.01) |
| MCS | Rural | 3 | 2.04 (1.90, 2.18) | 1.75 (1.63, 1.87) | 4.97 (4.83, 5.11) | 6.47 (6.31, 6.63) |

*Note.* Adjusted models included birth sex, highest qualification achieved, pre-pandemic self-reported health, pre-pandemic psychological distress, and household composition as covariates. BCS: British Cohort Study, 1970 birth cohort; GAD-2: 2-item General Anxiety Disorder questionnaire; MCS: Millennium Cohort Study, 2000 birth cohort; NCDS: National Child and Development Study, 1958 birth cohort; NS: Next Steps, 1990 cohort; NSHD: National Survey of Health and Development, 1946 birth cohort; ONS: UK Office for National Statistics; PHQ-2: 2-item Patient Health Questionnaire; UCLA-3: 3-item UCLA loneliness scale. Survey wave 1: May 2020; survey wave 2: September/October 2020; survey wave 3: February/March 2021.

#### Figure S10.1. Unadjusted and adjusted (by birth sex, highest qualification achieved, pre-pandemic self-reported health, pre-pandemic psychological distress, and household composition) anxiety symptomatology (GAD-2) marginal mean estimates and 95% confidence intervals by urbanicity.

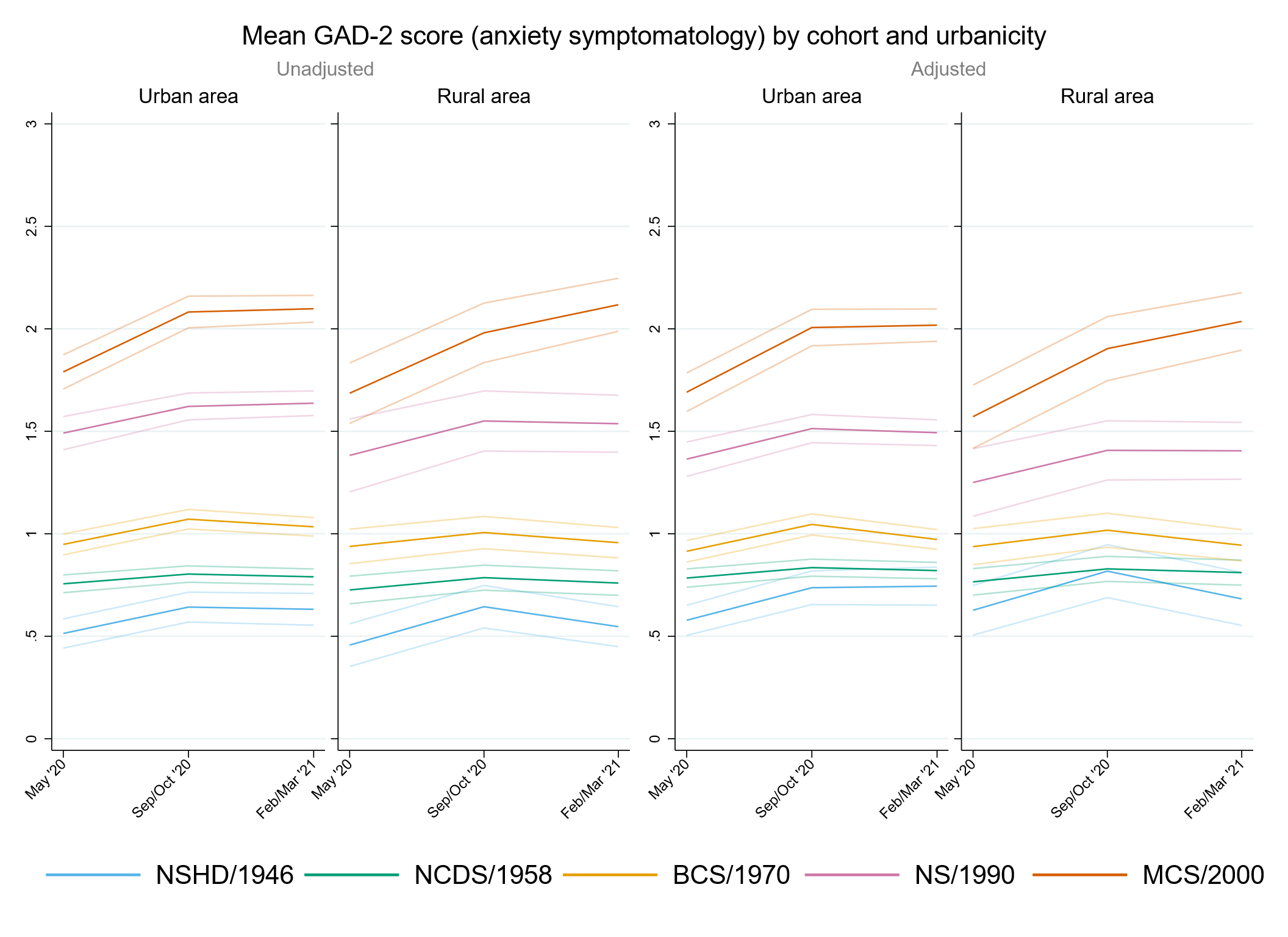

#### Figure S10.2. Unadjusted and adjusted (by birth sex, highest qualification achieved, pre-pandemic self-reported health, pre-pandemic psychological distress, and household composition) depressive symptomatology (PHQ-2) marginal mean estimates and 95% confidence intervals by urbanicity.

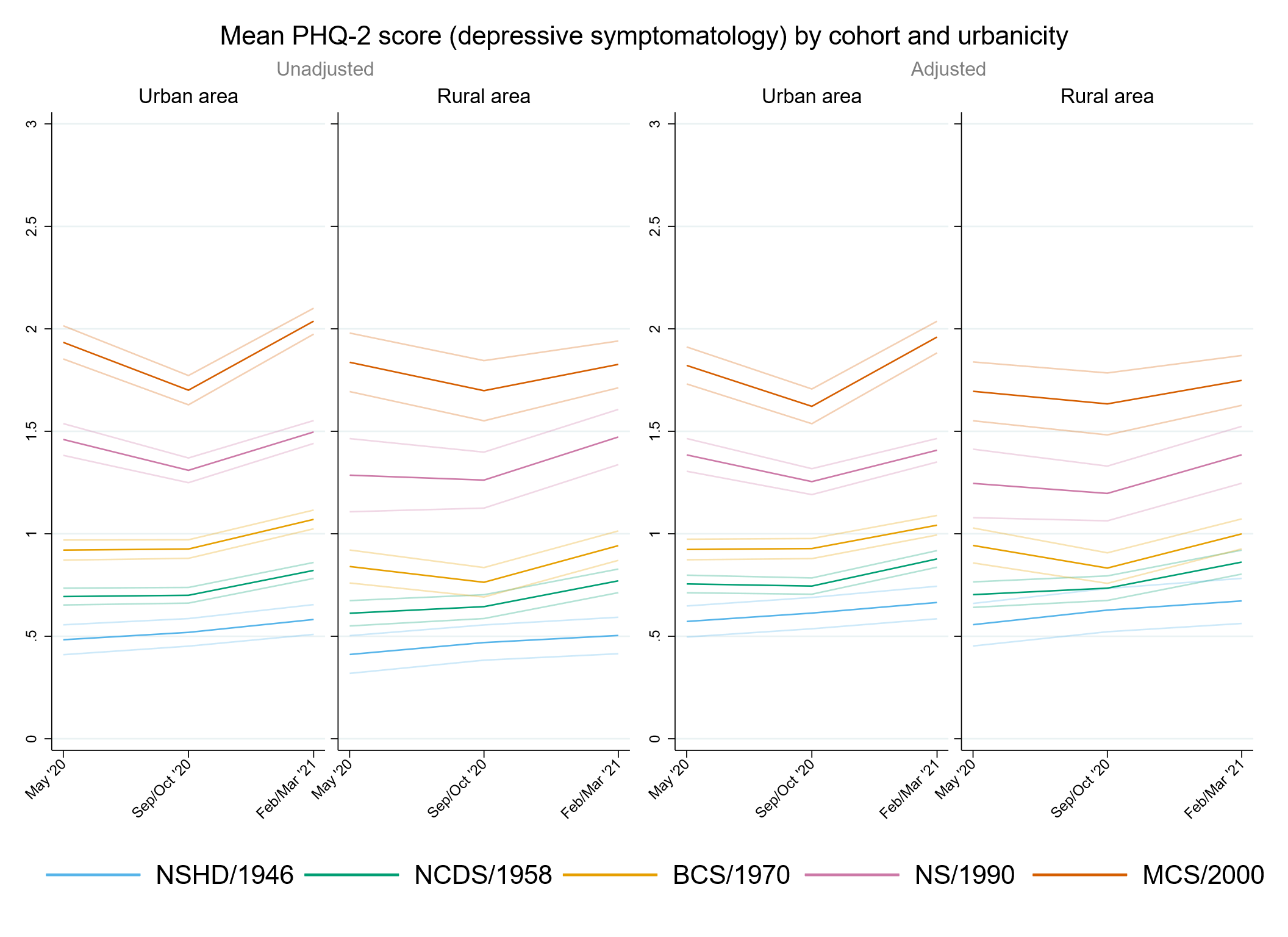

#### Figure S10.3. Unadjusted and adjusted (by birth sex, highest qualification achieved, pre-pandemic self-reported health, pre-pandemic psychological distress, and household composition) loneliness (UCLA-3) marginal mean estimates and 95% confidence intervals by urbanicity.

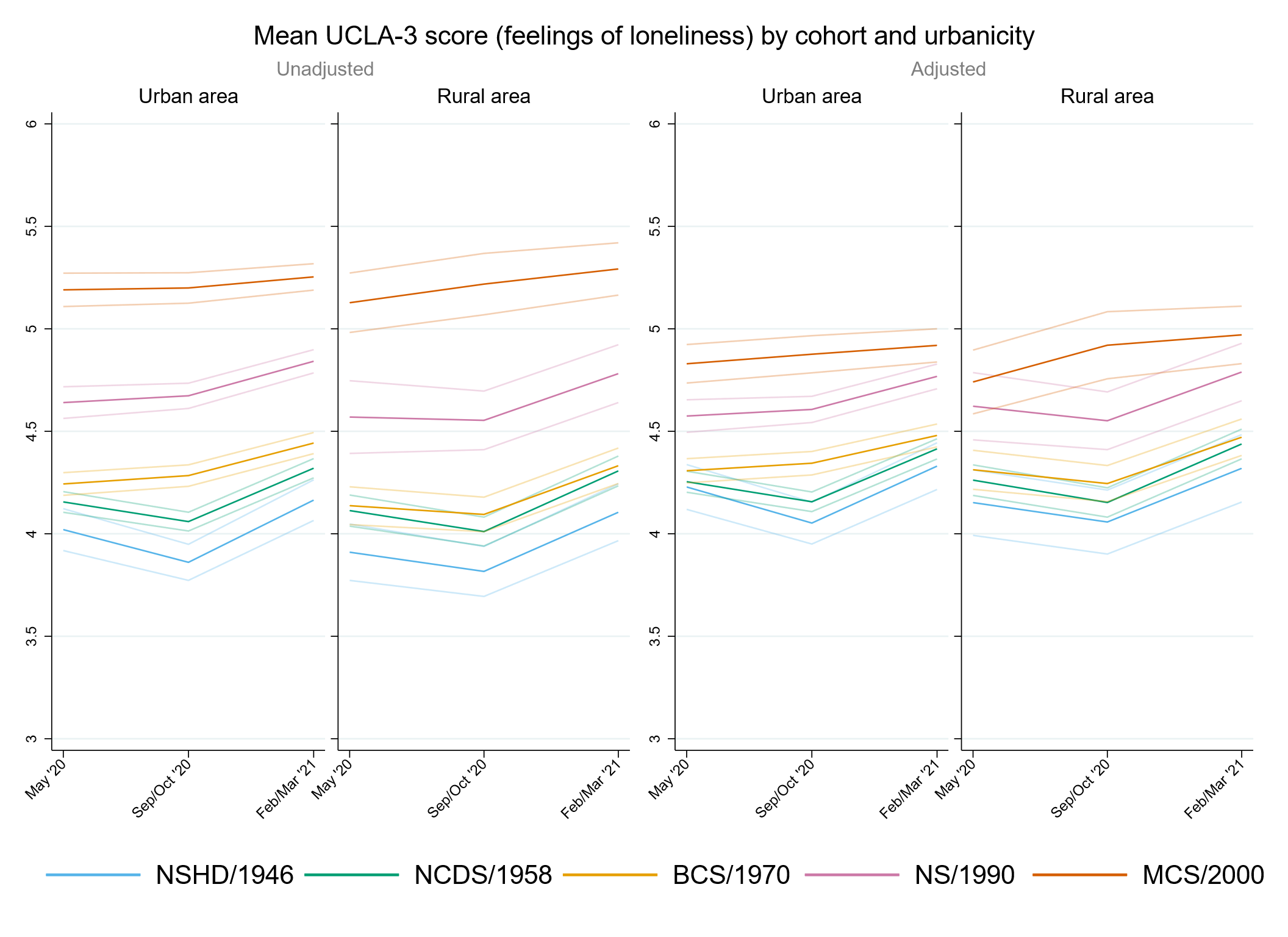

#### Figure S10.4. Unadjusted and adjusted (by birth sex, highest qualification achieved, pre-pandemic self-reported health, pre-pandemic psychological distress, and household composition) life satisfaction marginal mean estimates and 95% confidence intervals by urbanicity.

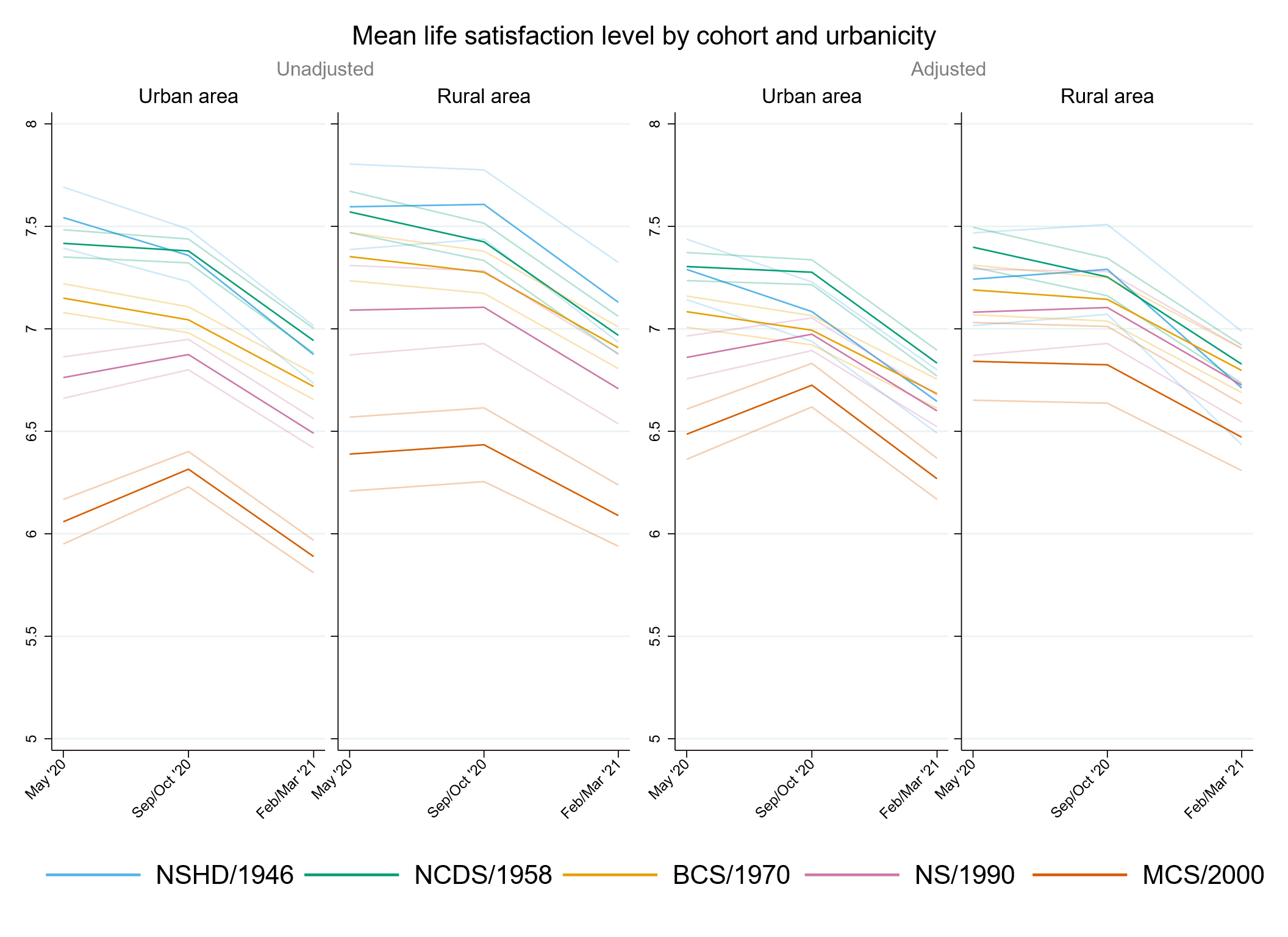

### Appendix S11. Results by UK country of residence.

#### Table S11.1. Results of multilevel growth curve models by UK country of residence.

|  | **Anxiety symptomatology (GAD-2)** | | | **Depressive symptomatology (PHQ-2)** | | | **Feelings of loneliness (UCLA-3)** | | | **Life satisfaction (ONS single question)** | | |
| --- | --- | --- | --- | --- | --- | --- | --- | --- | --- | --- | --- | --- |
| **Unadjusted models** |  | | |  | | |  | | |  | | |
| N participants | 24,739 |  |  | 24,738 |  |  | 24,762 |  |  | 24,815 |  |  |
| N observations | 52,557 |  |  | 52,541 |  |  | 52,619 |  |  | 52,844 |  |  |
|  | ***B* (95% CI)** | **χ2** | ***p*** | ***B* (95% CI)** | **χ2** | ***p*** | ***B* (95% CI)** | **χ2** | ***p*** | ***B* (95% CI)** | **χ2** | ***p*** |
| Time (linear) | 0.10 (0.04, 0.17) |  | 0.001 | -0.05 (-0.10, 0.01) |  | 0.106 | -0.29 (-0.36, -0.22) |  | <0.001 | 0.15 (0.05, 0.24) |  | 0.002 |
| Time (quad) | -0.04 (-0.07, -0.01) |  | 0.006 | 0.06 (0.03, 0.08) |  | <0.001 | 0.19 (0.16, 0.22) |  | <0.001 | -0.20 (-0.24, -0.16) |  | <0.001 |
| Cohort (ref. NCDS) |  | 846.8 | <0.001 |  | 1127.2 | <0.001 |  | 602.7 | <0.001 |  | 524.4 | <0.001 |
| NSHD | -0.25 (-0.33, -0.18) |  | <0.001 | -0.21 (-0.28, -0.14) |  | <0.001 | -0.16 (-0.26, -0.06) |  | 0.002 | 0.15 (0.00, 0.29) |  | 0.045 |
| BCS | 0.21 (0.15, 0.27) |  | <0.001 | 0.23 (0.17, 0.29) |  | <0.001 | 0.08 (0.01, 0.15) |  | 0.018 | -0.23 (-0.32, -0.14) |  | <0.001 |
| NS | 0.73 (0.65, 0.81) |  | <0.001 | 0.76 (0.68, 0.84) |  | <0.001 | 0.48 (0.40, 0.57) |  | <0.001 | -0.61 (-0.73, -0.50) |  | <0.001 |
| MCS | 0.99 (0.90, 1.09) |  | <0.001 | 1.22 (1.13, 1.31) |  | <0.001 | 1.02 (0.93, 1.11) |  | <0.001 | -1.30 (-1.42, -1.18) |  | <0.001 |
| Linear change * cohort (ref. NCDS) |  | 17.1 | 0.002 |  | 39.0 | <0.001 |  | 31.9 | <0.001 |  | 26.1 | <0.001 |
| NSHD | 0.14 (0.02, 0.26) |  | 0.026 | 0.09 (-0.03, 0.21) |  | 0.138 | -0.06 (-0.21, 0.09) |  | 0.448 | -0.06 (-0.29, 0.17) |  | 0.619 |
| BCS | 0.06 (-0.05, 0.16) |  | 0.277 | -0.06 (-0.16, 0.03) |  | 0.186 | 0.21 (0.11, 0.32) |  | <0.001 | -0.13 (-0.27, 0.01) |  | 0.078 |
| NS | 0.10 (-0.04, 0.24) |  | 0.179 | -0.25 (-0.38, -0.11) |  | <0.001 | 0.21 (0.08, 0.34) |  | 0.002 | 0.21 (0.03, 0.39) |  | 0.020 |
| MCS | 0.32 (0.16, 0.49) |  | <0.001 | -0.40 (-0.57, -0.23) |  | <0.001 | 0.31 (0.15, 0.48) |  | <0.001 | 0.36 (0.15, 0.58) |  | 0.001 |
| Quad. change * cohort (ref. NCDS) |  | 6.5 | 0.166 |  | 41.7 | <0.001 |  | 39.4 | <0.001 |  | 19.1 | <0.001 |
| NSHD | -0.05 (-0.11, 0.01) |  | 0.096 | -0.05 (-0.11, 0.00) |  | 0.065 | 0.02 (-0.04, 0.09) |  | 0.483 | 0.00 (-0.11, 0.11) |  | 0.960 |
| BCS | -0.03 (-0.07, 0.02) |  | 0.269 | 0.03 (-0.01, 0.07) |  | 0.182 | -0.10 (-0.15, -0.06) |  | <0.001 | 0.08 (0.02, 0.15) |  | 0.013 |
| NS | -0.02 (-0.09, 0.04) |  | 0.464 | 0.11 (0.05, 0.17) |  | 0.001 | -0.10 (-0.16, -0.04) |  | 0.001 | -0.05 (-0.13, 0.03) |  | 0.191 |
| MCS | -0.09 (-0.16, -0.01) |  | 0.030 | 0.19 (0.11, 0.27) |  | <0.001 | -0.18 (-0.26, -0.10) |  | <0.001 | -0.11 (-0.20, -0.01) |  | 0.032 |
| UK country of residence (ref. England) |  | 2.2 | 0.537 |  | 6.7 | 0.082 |  | 1.6 | 0.655 |  | 21.4 | <0.001 |
| Northern Ireland | 0.10 (-0.16, 0.37) |  | 0.433 | -0.27 (-0.52, -0.02) |  | 0.034 | -0.14 (-0.40, 0.12) |  | 0.290 | 0.49 (0.15, 0.83) |  | 0.004 |
| Scotland | 0.00 (-0.13, 0.13) |  | 0.947 | -0.05 (-0.17, 0.07) |  | 0.408 | -0.05 (-0.20, 0.10) |  | 0.496 | 0.20 (0.02, 0.39) |  | 0.033 |
| Wales | 0.13 (-0.07, 0.33) |  | 0.214 | 0.10 (-0.07, 0.27) |  | 0.240 | -0.02 (-0.21, 0.16) |  | 0.810 | 0.36 (0.13, 0.58) |  | 0.002 |
| Linear change * UK country of residence (ref. England) |  | 2.9 | 0.414 |  | 4.6 | 0.202 |  | 2.4 | 0.493 |  | 3.3 | 0.342 |
| Northern Ireland | 0.10 (-0.36, 0.56) |  | 0.675 | 0.23 (-0.25, 0.71) |  | 0.345 | -0.22 (-0.74, 0.30) |  | 0.401 | 0.15 (-0.44, 0.74) |  | 0.624 |
| Scotland | -0.06 (-0.26, 0.14) |  | 0.578 | 0.16 (-0.05, 0.37) |  | 0.139 | 0.13 (-0.10, 0.36) |  | 0.262 | -0.25 (-0.54, 0.05) |  | 0.098 |
| Wales | -0.28 (-0.63, 0.07) |  | 0.117 | -0.15 (-0.41, 0.11) |  | 0.251 | -0.09 (-0.38, 0.21) |  | 0.559 | -0.13 (-0.49, 0.22) |  | 0.455 |
| Quad. change * UK country of residence (ref. England) |  | 2.2 | 0.536 |  | 3.2 | 0.356 |  | 3.1 | 0.378 |  | 4.7 | 0.199 |
| Northern Ireland | -0.10 (-0.31, 0.11) |  | 0.353 | -0.07 (-0.29, 0.15) |  | 0.515 | 0.14 (-0.10, 0.38) |  | 0.252 | -0.15 (-0.42, 0.11) |  | 0.261 |
| Scotland | 0.02 (-0.08, 0.11) |  | 0.732 | -0.06 (-0.16, 0.04) |  | 0.236 | -0.06 (-0.16, 0.05) |  | 0.283 | 0.13 (-0.01, 0.27) |  | 0.066 |
| Wales | 0.09 (-0.07, 0.24) |  | 0.266 | 0.07 (-0.05, 0.19) |  | 0.264 | 0.05 (-0.09, 0.19) |  | 0.474 | 0.01 (-0.15, 0.17) |  | 0.919 |
| Cohort (ref. NCDS) * UK country of residence (ref. England) |  | 4.6 | 0.601 |  | 3.3 | 0.775 |  | 6.5 | 0.368 |  | 8.9 | 0.181 |
| NSHD * Scotland | -0.05 (-0.30, 0.20) |  | 0.704 | 0.00 (-0.25, 0.25) |  | 0.978 | 0.04 (-0.34, 0.42) |  | 0.826 | -0.19 (-0.66, 0.27) |  | 0.416 |
| BCS * Scotland | -0.12 (-0.32, 0.07) |  | 0.220 | 0.01 (-0.19, 0.21) |  | 0.929 | -0.16 (-0.38, 0.06) |  | 0.161 | -0.12 (-0.40, 0.16) |  | 0.405 |
| BCS * Wales | -0.07 (-0.35, 0.22) |  | 0.648 | 0.00 (-0.25, 0.26) |  | 0.974 | 0.07 (-0.22, 0.36) |  | 0.655 | -0.45 (-0.80, -0.11) |  | 0.010 |
| MCS * Scotland | 0.14 (-0.12, 0.39) |  | 0.304 | 0.18 (-0.07, 0.42) |  | 0.160 | 0.13 (-0.13, 0.39) |  | 0.321 | -0.20 (-0.54, 0.15) |  | 0.266 |
| MCS * Wales | 0.00 (-0.28, 0.29) |  | 0.974 | -0.10 (-0.36, 0.17) |  | 0.464 | -0.04 (-0.31, 0.24) |  | 0.784 | -0.26 (-0.62, 0.09) |  | 0.146 |
| Linear change * cohort (ref. NCDS) * UK country of residence (ref. England) |  | 5.7 | 0.46 |  | 5.3 | 0.501 |  | 7.3 | 0.291 |  | 2.2 | 0.895 |
| NSHD * Scotland | 0.21 (-0.25, 0.67) |  | 0.377 | -0.16 (-0.64, 0.31) |  | 0.500 | -0.49 (-1.10, 0.13) |  | 0.122 | -0.17 (-0.98, 0.64) |  | 0.686 |
| BCS * Scotland | 0.21 (-0.15, 0.57) |  | 0.252 | -0.08 (-0.45, 0.29) |  | 0.683 | 0.04 (-0.35, 0.43) |  | 0.834 | 0.24 (-0.24, 0.72) |  | 0.330 |
| BCS * Wales | 0.49 (-0.05, 1.03) |  | 0.076 | 0.23 (-0.17, 0.62) |  | 0.262 | 0.11 (-0.37, 0.58) |  | 0.657 | 0.22 (-0.34, 0.77) |  | 0.445 |
| MCS * Scotland | 0.05 (-0.39, 0.50) |  | 0.823 | -0.35 (-0.79, 0.08) |  | 0.112 | -0.10 (-0.54, 0.35) |  | 0.670 | 0.20 (-0.37, 0.77) |  | 0.498 |
| MCS * Wales | 0.27 (-0.26, 0.81) |  | 0.321 | 0.29 (-0.22, 0.79) |  | 0.267 | -0.10 (-0.61, 0.41) |  | 0.704 | 0.16 (-0.53, 0.84) |  | 0.658 |
| Quad. change * cohort (ref. NCDS) * UK country of residence (ref. England) |  | 3.8 | 0.704 |  | 3.2 | 0.782 |  | 8.1 | 0.23 |  | 3.1 | 0.794 |
| NSHD * Scotland | -0.12 (-0.33, 0.08) |  | 0.246 | 0.07 (-0.16, 0.29) |  | 0.555 | 0.25 (-0.03, 0.52) |  | 0.080 | 0.10 (-0.27, 0.47) |  | 0.588 |
| BCS * Scotland | -0.08 (-0.24, 0.08) |  | 0.335 | 0.04 (-0.13, 0.20) |  | 0.659 | 0.00 (-0.18, 0.18) |  | 0.983 | -0.14 (-0.36, 0.08) |  | 0.217 |
| BCS * Wales | -0.15 (-0.40, 0.09) |  | 0.209 | -0.06 (-0.25, 0.12) |  | 0.510 | 0.00 (-0.22, 0.22) |  | 0.986 | -0.07 (-0.34, 0.19) |  | 0.582 |
| MCS * Scotland | -0.05 (-0.25, 0.15) |  | 0.628 | 0.13 (-0.07, 0.33) |  | 0.205 | 0.03 (-0.17, 0.23) |  | 0.779 | -0.12 (-0.38, 0.14) |  | 0.357 |
| MCS * Wales | -0.08 (-0.33, 0.17) |  | 0.525 | -0.12 (-0.36, 0.11) |  | 0.310 | 0.08 (-0.16, 0.32) |  | 0.495 | -0.02 (-0.33, 0.30) |  | 0.925 |
| **Adjusted models** |  | | |  | | |  | | |  | | |
| N participants | 21,277 |  |  | 21,276 |  |  | 21,296 |  |  | 21,337 |  |  |
| N observations | 46,238 |  |  | 46,227 |  |  | 46,284 |  |  | 46,478 |  |  |
|  | ***B* (95% CI)** | **χ2** | ***p*** | ***B* (95% CI)** | **χ2** | ***p*** | ***B* (95% CI)** | **χ2** | ***p*** | ***B* (95% CI)** | **χ2** | ***p*** |
| Time (linear) | 0.09 (0.03, 0.16) |  | 0.005 | -0.07 (-0.13, -0.01) |  | 0.024 | -0.29 (-0.36, -0.22) |  | <0.001 | 0.15 (0.05, 0.25) |  | 0.003 |
| Time (quad) | -0.04 (-0.07, 0.00) |  | 0.024 | 0.07 (0.04, 0.10) |  | <0.001 | 0.19 (0.15, 0.22) |  | <0.001 | -0.20 (-0.24, -0.15) |  | <0.001 |
| Cohort (ref. NCDS) |  | 424.5 | <0.001 |  | 591.3 | <0.001 |  | 133.2 | <0.001 |  | 123.8 | <0.001 |
| NSHD | -0.19 (-0.26, -0.11) |  | <0.001 | -0.17 (-0.24, -0.09) |  | <0.001 | -0.05 (-0.16, 0.05) |  | 0.326 | -0.03 (-0.18, 0.11) |  | 0.680 |
| BCS | 0.15 (0.09, 0.21) |  | <0.001 | 0.19 (0.13, 0.25) |  | <0.001 | 0.06 (-0.01, 0.13) |  | 0.103 | -0.18 (-0.28, -0.09) |  | <0.001 |
| NS | 0.57 (0.48, 0.65) |  | <0.001 | 0.62 (0.54, 0.71) |  | <0.001 | 0.33 (0.24, 0.41) |  | <0.001 | -0.41 (-0.52, -0.29) |  | <0.001 |
| MCS | 0.86 (0.75, 0.97) |  | <0.001 | 1.03 (0.93, 1.14) |  | <0.001 | 0.54 (0.43, 0.65) |  | <0.001 | -0.73 (-0.87, -0.58) |  | <0.001 |
| Linear change * cohort (ref. NCDS) |  | 22.4 | <0.001 |  | 21.6 | <0.001 |  | 32.8 | <0.001 |  | 19.1 | <0.001 |
| NSHD | 0.18 (0.04, 0.32) |  | 0.013 | 0.12 (-0.02, 0.25) |  | 0.088 | -0.04 (-0.21, 0.13) |  | 0.631 | -0.06 (-0.33, 0.21) |  | 0.686 |
| BCS | 0.11 (0.00, 0.22) |  | 0.060 | -0.03 (-0.14, 0.07) |  | 0.556 | 0.22 (0.10, 0.33) |  | <0.001 | -0.13 (-0.28, 0.03) |  | 0.105 |
| NS | 0.14 (-0.01, 0.30) |  | 0.063 | -0.19 (-0.33, -0.04) |  | 0.010 | 0.20 (0.06, 0.34) |  | 0.005 | 0.20 (0.01, 0.39) |  | 0.038 |
| MCS | 0.39 (0.21, 0.57) |  | <0.001 | -0.29 (-0.47, -0.11) |  | 0.001 | 0.41 (0.23, 0.60) |  | <0.001 | 0.32 (0.09, 0.55) |  | 0.007 |
| Quad. change * cohort (ref. NCDS) |  | 11.2 | 0.024 |  | 23 | <0.001 |  | 38.3 | <0.001 |  | 15.8 | 0.003 |
| NSHD | -0.06 (-0.13, 0.01) |  | 0.071 | -0.06 (-0.13, 0.00) |  | 0.052 | 0.01 (-0.07, 0.10) |  | 0.725 | -0.01 (-0.14, 0.13) |  | 0.943 |
| BCS | -0.06 (-0.11, 0.00) |  | 0.032 | 0.01 (-0.04, 0.06) |  | 0.800 | -0.11 (-0.16, -0.06) |  | <0.001 | 0.09 (0.02, 0.16) |  | 0.017 |
| NS | -0.05 (-0.12, 0.02) |  | 0.145 | 0.07 (0.01, 0.14) |  | 0.027 | -0.09 (-0.16, -0.03) |  | 0.003 | -0.05 (-0.13, 0.04) |  | 0.282 |
| MCS | -0.11 (-0.20, -0.03) |  | 0.007 | 0.15 (0.06, 0.23) |  | 0.001 | -0.22 (-0.30, -0.13) |  | <0.001 | -0.10 (-0.21, 0.00) |  | 0.053 |
| UK country of residence (ref. England) |  | 0.5 | 0.925 |  | 5.9 | 0.115 |  | 2.0 | 0.578 |  | 16.3 | 0.001 |
| Northern Ireland | 0.09 (-0.16, 0.34) |  | 0.498 | -0.27 (-0.52, -0.03) |  | 0.030 | -0.14 (-0.39, 0.11) |  | 0.268 | 0.43 (0.08, 0.78) |  | 0.017 |
| Scotland | 0.00 (-0.12, 0.12) |  | 0.989 | -0.04 (-0.16, 0.08) |  | 0.517 | -0.06 (-0.19, 0.07) |  | 0.388 | 0.15 (-0.03, 0.33) |  | 0.100 |
| Wales | 0.01 (-0.15, 0.17) |  | 0.914 | 0.07 (-0.09, 0.23) |  | 0.391 | 0.00 (-0.18, 0.18) |  | 0.982 | 0.33 (0.11, 0.55) |  | 0.003 |
| Linear change * UK country of residence (ref. England) | 0.3 | 0.967 | <0.001 |  | 2.4 | 0.487 |  | 3.2 | 0.356 |  | 1.8 | 0.618 |
| Northern Ireland | 0.07 (-0.44, 0.58) |  | 0.792 | 0.17 (-0.35, 0.69) |  | 0.518 | -0.34 (-0.93, 0.24) |  | 0.247 | 0.15 (-0.50, 0.80) |  | 0.646 |
| Scotland | -0.05 (-0.26, 0.17) |  | 0.677 | 0.14 (-0.09, 0.36) |  | 0.231 | 0.14 (-0.08, 0.36) |  | 0.218 | -0.18 (-0.50, 0.14) |  | 0.264 |
| Wales | -0.02 (-0.33, 0.28) |  | 0.875 | -0.10 (-0.40, 0.19) |  | 0.486 | -0.08 (-0.40, 0.23) |  | 0.598 | -0.12 (-0.49, 0.24) |  | 0.509 |
| Quad. change * UK country of residence (ref. England) |  | 0.8 | 0.847 |  | 1.1 | 0.784 |  | 3.6 | 0.309 |  | 2.6 | 0.465 |
| Northern Ireland | -0.08 (-0.32, 0.15) |  | 0.485 | -0.04 (-0.28, 0.20) |  | 0.733 | 0.20 (-0.07, 0.48) |  | 0.147 | -0.14 (-0.43, 0.15) |  | 0.331 |
| Scotland | 0.02 (-0.08, 0.12) |  | 0.707 | -0.04 (-0.15, 0.06) |  | 0.418 | -0.06 (-0.16, 0.05) |  | 0.286 | 0.10 (-0.05, 0.25) |  | 0.204 |
| Wales | -0.03 (-0.17, 0.11) |  | 0.688 | 0.04 (-0.11, 0.18) |  | 0.617 | 0.04 (-0.11, 0.19) |  | 0.602 | 0.01 (-0.16, 0.17) |  | 0.953 |
| Cohort (ref. NCDS) * UK country of residence (ref. England) |  | 3.6 | 0.731 |  | 3.9 | 0.69 |  | 10.3 | 0.111 |  | 8.0 | 0.236 |
| NSHD * Scotland | -0.04 (-0.31, 0.23) |  | 0.755 | 0.00 (-0.23, 0.24) |  | 0.973 | 0.12 (-0.29, 0.52) |  | 0.576 | -0.10 (-0.52, 0.31) |  | 0.627 |
| BCS * Scotland | -0.03 (-0.23, 0.16) |  | 0.744 | 0.06 (-0.14, 0.26) |  | 0.565 | -0.13 (-0.35, 0.08) |  | 0.215 | -0.15 (-0.43, 0.13) |  | 0.306 |
| BCS * Wales | -0.14 (-0.41, 0.13) |  | 0.307 | -0.07 (-0.31, 0.18) |  | 0.584 | 0.10 (-0.19, 0.39) |  | 0.504 | -0.45 (-0.80, -0.10) |  | 0.013 |
| MCS * Scotland | 0.13 (-0.13, 0.39) |  | 0.338 | 0.19 (-0.05, 0.43) |  | 0.119 | 0.20 (-0.05, 0.45) |  | 0.124 | -0.20 (-0.55, 0.14) |  | 0.244 |
| MCS * Wales | 0.08 (-0.17, 0.32) |  | 0.545 | -0.06 (-0.31, 0.19) |  | 0.645 | -0.07 (-0.35, 0.20) |  | 0.592 | -0.29 (-0.65, 0.07) |  | 0.114 |
| Linear change * cohort (ref. NCDS) * UK country of residence (ref. England) |  | 3.0 | 0.805 |  | 5.3 | 0.507 |  | 10.0 | 0.126 |  | 3.9 | 0.685 |
| NSHD * Scotland | 0.16 (-0.39, 0.71) |  | 0.571 | -0.09 (-0.53, 0.36) |  | 0.706 | -0.74 (-1.44, -0.05) |  | 0.037 | -0.25 (-0.94, 0.44) |  | 0.475 |
| BCS * Scotland | -0.01 (-0.39, 0.37) |  | 0.955 | -0.25 (-0.64, 0.15) |  | 0.219 | -0.03 (-0.41, 0.35) |  | 0.879 | 0.30 (-0.20, 0.81) |  | 0.240 |
| BCS * Wales | 0.37 (-0.20, 0.94) |  | 0.199 | 0.27 (-0.17, 0.71) |  | 0.222 | 0.12 (-0.40, 0.65) |  | 0.647 | 0.33 (-0.29, 0.96) |  | 0.296 |
| MCS * Scotland | -0.04 (-0.52, 0.44) |  | 0.874 | -0.40 (-0.87, 0.07) |  | 0.091 | -0.17 (-0.64, 0.31) |  | 0.486 | 0.18 (-0.44, 0.80) |  | 0.562 |
| MCS * Wales | -0.09 (-0.62, 0.44) |  | 0.732 | 0.07 (-0.47, 0.61) |  | 0.800 | -0.27 (-0.82, 0.28) |  | 0.343 | 0.27 (-0.46, 0.99) |  | 0.469 |
| Quad. change * cohort (ref. NCDS) * UK country of residence (ref. England) |  | 2.7 | 0.848 |  | 4.0 | 0.682 |  | 11.2 | 0.084 |  | 4.8 | 0.564 |
| NSHD * Scotland | -0.10 (-0.37, 0.17) |  | 0.479 | 0.01 (-0.20, 0.22) |  | 0.930 | 0.32 (0.01, 0.63) |  | 0.040 | 0.19 (-0.16, 0.53) |  | 0.291 |
| BCS * Scotland | 0.01 (-0.16, 0.18) |  | 0.886 | 0.12 (-0.06, 0.29) |  | 0.193 | 0.02 (-0.16, 0.21) |  | 0.807 | -0.17 (-0.40, 0.06) |  | 0.157 |
| BCS * Wales | -0.11 (-0.37, 0.14) |  | 0.384 | -0.09 (-0.29, 0.12) |  | 0.409 | -0.03 (-0.27, 0.21) |  | 0.797 | -0.10 (-0.40, 0.20) |  | 0.511 |
| MCS * Scotland | -0.01 (-0.23, 0.21) |  | 0.924 | 0.16 (-0.06, 0.37) |  | 0.162 | 0.05 (-0.17, 0.27) |  | 0.679 | -0.10 (-0.38, 0.19) |  | 0.505 |
| MCS * Wales | 0.09 (-0.16, 0.34) |  | 0.472 | -0.01 (-0.27, 0.25) |  | 0.933 | 0.17 (-0.09, 0.43) |  | 0.196 | -0.06 (-0.39, 0.27) |  | 0.719 |
| **Sensitivity models** |  | | |  | | |  | | |  | | |
| N participants | 21,277 |  |  | 21,276 |  |  | 21,296 |  |  | 21,337 |  |  |
| N observations | 46,238 |  |  | 46,227 |  |  | 46,284 |  |  | 46,478 |  |  |
|  | ***B* (95% CI)** | **χ2** | ***p*** | ***B* (95% CI)** | **χ2** | ***p*** | ***B* (95% CI)** | **χ2** | ***p*** | ***B* (95% CI)** | **χ2** | ***p*** |
| Time (linear) | 0.10 (0.03, 0.17) |  | 0.003 | -0.06 (-0.12, 0.00) |  | 0.047 | -0.28 (-0.35, -0.21) |  | <0.001 | 0.14 (0.04, 0.23) |  | 0.007 |
| Time (quad) | -0.04 (-0.07, -0.01) |  | 0.021 | 0.06 (0.04, 0.09) |  | <0.001 | 0.18 (0.15, 0.22) |  | <0.001 | -0.20 (-0.24, -0.15) |  | <0.001 |
| Cohort (ref. NCDS) |  | 741.9 | <0.001 |  | 948.6 | <0.001 |  | 509.6 | <0.001 |  | 436.5 | <0.001 |
| NSHD | -0.24 (-0.32, -0.16) |  | <0.001 | -0.20 (-0.28, -0.12) |  | <0.001 | -0.14 (-0.24, -0.03) |  | 0.014 | 0.09 (-0.06, 0.25) |  | 0.219 |
| BCS | 0.19 (0.13, 0.26) |  | <0.001 | 0.21 (0.15, 0.27) |  | <0.001 | 0.05 (-0.02, 0.12) |  | 0.175 | -0.20 (-0.29, -0.11) |  | <0.001 |
| NS | 0.73 (0.64, 0.82) |  | <0.001 | 0.74 (0.66, 0.83) |  | <0.001 | 0.49 (0.40, 0.57) |  | <0.001 | -0.60 (-0.72, -0.49) |  | <0.001 |
| MCS | 0.99 (0.90, 1.09) |  | <0.001 | 1.18 (1.08, 1.27) |  | <0.001 | 0.99 (0.89, 1.08) |  | <0.001 | -1.24 (-1.36, -1.11) |  | <0.001 |
| Linear change * cohort (ref. NCDS) |  | 20.1 | <0.001 |  | 22.3 | <0.001 |  | 30.8 | <0.001 |  | 18.3 | 0.001 |
| NSHD | 0.17 (0.03, 0.32) |  | 0.017 | 0.12 (-0.02, 0.25) |  | 0.098 | -0.05 (-0.22, 0.13) |  | 0.605 | -0.04 (-0.32, 0.23) |  | 0.745 |
| BCS | 0.10 (-0.01, 0.21) |  | 0.078 | -0.04 (-0.15, 0.07) |  | 0.463 | 0.21 (0.09, 0.32) |  | <0.001 | -0.11 (-0.27, 0.04) |  | 0.154 |
| NS | 0.14 (-0.02, 0.29) |  | 0.082 | -0.19 (-0.34, -0.05) |  | 0.009 | 0.21 (0.06, 0.35) |  | 0.005 | 0.21 (0.01, 0.40) |  | 0.035 |
| MCS | 0.38 (0.19, 0.56) |  | <0.001 | -0.31 (-0.49, -0.13) |  | 0.001 | 0.40 (0.22, 0.59) |  | <0.001 | 0.33 (0.10, 0.56) |  | 0.006 |
| Quad. change * cohort (ref. NCDS) |  | 10.5 | 0.033 |  | 23.1 | <0.001 |  | 37.3 | <0.001 |  | 14.2 | 0.007 |
| NSHD | -0.06 (-0.13, 0.01) |  | 0.075 | -0.06 (-0.13, 0.00) |  | 0.049 | 0.01 (-0.07, 0.10) |  | 0.744 | 0.00 (-0.14, 0.13) |  | 0.945 |
| BCS | -0.05 (-0.11, 0.00) |  | 0.043 | 0.01 (-0.04, 0.06) |  | 0.688 | -0.11 (-0.16, -0.05) |  | <0.001 | 0.08 (0.01, 0.15) |  | 0.025 |
| NS | -0.05 (-0.12, 0.02) |  | 0.168 | 0.07 (0.01, 0.14) |  | 0.026 | -0.10 (-0.16, -0.03) |  | 0.003 | -0.05 (-0.13, 0.04) |  | 0.282 |
| MCS | -0.11 (-0.20, -0.03) |  | 0.009 | 0.15 (0.07, 0.24) |  | 0.001 | -0.22 (-0.30, -0.13) |  | <0.001 | -0.10 (-0.20, 0.01) |  | 0.066 |
| UK country of residence (ref. England) |  | 0.5 | 0.921 |  | 5.9 | 0.116 |  | 1.7 | 0.632 |  | 13.8 | 0.003 |
| Northern Ireland | 0.09 (-0.18, 0.36) |  | 0.512 | -0.27 (-0.54, -0.01) |  | 0.040 | -0.12 (-0.38, 0.15) |  | 0.392 | 0.39 (0.03, 0.75) |  | 0.035 |
| Scotland | -0.01 (-0.14, 0.12) |  | 0.827 | -0.05 (-0.18, 0.07) |  | 0.389 | -0.07 (-0.22, 0.07) |  | 0.324 | 0.17 (-0.02, 0.36) |  | 0.085 |
| Wales | 0.01 (-0.16, 0.18) |  | 0.916 | 0.08 (-0.09, 0.25) |  | 0.359 | 0.01 (-0.19, 0.20) |  | 0.945 | 0.32 (0.08, 0.55) |  | 0.008 |
| Linear change * UK country of residence (ref. England) |  | 0.4 | 0.932 |  | 2.2 | 0.528 |  | 2.9 | 0.403 |  | 1.8 | 0.608 |
| Northern Ireland | 0.10 (-0.42, 0.62) |  | 0.699 | 0.22 (-0.32, 0.75) |  | 0.422 | -0.33 (-0.91, 0.25) |  | 0.264 | 0.17 (-0.49, 0.83) |  | 0.610 |
| Scotland | -0.06 (-0.27, 0.16) |  | 0.594 | 0.12 (-0.10, 0.34) |  | 0.290 | 0.14 (-0.09, 0.36) |  | 0.229 | -0.18 (-0.49, 0.14) |  | 0.279 |
| Wales | 0.01 (-0.30, 0.31) |  | 0.970 | -0.09 (-0.38, 0.20) |  | 0.540 | -0.06 (-0.38, 0.25) |  | 0.691 | -0.14 (-0.50, 0.23) |  | 0.471 |
| Quad. change * UK country of residence (ref. England) |  | 1.4 | 0.714 |  | 1.2 | 0.76 |  | 3.3 | 0.343 |  | 2.5 | 0.474 |
| Northern Ireland | -0.11 (-0.35, 0.13) |  | 0.364 | -0.07 (-0.32, 0.17) |  | 0.557 | 0.19 (-0.09, 0.46) |  | 0.179 | -0.14 (-0.43, 0.16) |  | 0.358 |
| Scotland | 0.02 (-0.08, 0.12) |  | 0.680 | -0.04 (-0.14, 0.06) |  | 0.459 | -0.06 (-0.16, 0.04) |  | 0.263 | 0.10 (-0.05, 0.25) |  | 0.198 |
| Wales | -0.04 (-0.18, 0.10) |  | 0.562 | 0.03 (-0.11, 0.17) |  | 0.630 | 0.03 (-0.12, 0.19) |  | 0.656 | 0.01 (-0.16, 0.18) |  | 0.908 |
| Cohort (ref. NCDS) * UK country of residence (ref. England) |  | 4.8 | 0.567 |  | 5.6 | 0.466 |  | 12.4 | 0.054 |  | 9.4 | 0.151 |
| NSHD * Scotland | -0.02 (-0.28, 0.25) |  | 0.888 | 0.00 (-0.23, 0.24) |  | 0.989 | 0.11 (-0.30, 0.52) |  | 0.594 | -0.11 (-0.54, 0.33) |  | 0.625 |
| BCS * Scotland | -0.05 (-0.26, 0.15) |  | 0.607 | 0.03 (-0.18, 0.25) |  | 0.753 | -0.15 (-0.38, 0.08) |  | 0.212 | -0.13 (-0.42, 0.17) |  | 0.392 |
| BCS * Wales | -0.07 (-0.35, 0.22) |  | 0.636 | -0.02 (-0.28, 0.25) |  | 0.906 | 0.13 (-0.18, 0.44) |  | 0.400 | -0.50 (-0.87, -0.13) |  | 0.009 |
| MCS * Scotland | 0.23 (-0.05, 0.51) |  | 0.111 | 0.27 (0.01, 0.52) |  | 0.040 | 0.26 (0.00, 0.53) |  | 0.054 | -0.29 (-0.65, 0.07) |  | 0.118 |
| MCS * Wales | 0.11 (-0.16, 0.37) |  | 0.428 | -0.03 (-0.30, 0.24) |  | 0.823 | -0.04 (-0.33, 0.24) |  | 0.764 | -0.31 (-0.69, 0.06) |  | 0.099 |
| Linear change * cohort (ref. NCDS) * UK country of residence (ref. England) |  | 1.9 | 0.926 |  | 4.4 | 0.62 |  | 8.8 | 0.186 |  | 3.8 | 0.703 |
| NSHD * Scotland | 0.15 (-0.41, 0.71) |  | 0.600 | -0.10 (-0.54, 0.35) |  | 0.675 | -0.76 (-1.46, -0.06) |  | 0.033 | -0.25 (-0.94, 0.45) |  | 0.486 |
| BCS * Scotland | 0.01 (-0.37, 0.39) |  | 0.942 | -0.21 (-0.60, 0.19) |  | 0.301 | -0.01 (-0.39, 0.38) |  | 0.975 | 0.28 (-0.23, 0.78) |  | 0.283 |
| BCS * Wales | 0.27 (-0.30, 0.85) |  | 0.350 | 0.21 (-0.24, 0.65) |  | 0.364 | 0.06 (-0.47, 0.59) |  | 0.832 | 0.40 (-0.22, 1.02) |  | 0.210 |
| MCS * Scotland | -0.04 (-0.53, 0.45) |  | 0.861 | -0.40 (-0.88, 0.07) |  | 0.098 | -0.17 (-0.65, 0.30) |  | 0.470 | 0.16 (-0.46, 0.79) |  | 0.610 |
| MCS * Wales | -0.02 (-0.56, 0.52) |  | 0.939 | 0.14 (-0.42, 0.69) |  | 0.625 | -0.20 (-0.76, 0.36) |  | 0.481 | 0.18 (-0.56, 0.91) |  | 0.632 |
| Quad. change * cohort (ref. NCDS) * UK country of residence (ref. England) |  | 1.6 | 0.954 |  | 3.2 | 0.783 |  | 9.7 | 0.139 |  | 4.7 | 0.585 |
| NSHD * Scotland | -0.09 (-0.37, 0.18) |  | 0.506 | 0.01 (-0.20, 0.23) |  | 0.901 | 0.33 (0.02, 0.63) |  | 0.036 | 0.18 (-0.16, 0.53) |  | 0.297 |
| BCS * Scotland | 0.01 (-0.17, 0.18) |  | 0.943 | 0.11 (-0.07, 0.28) |  | 0.241 | 0.02 (-0.17, 0.20) |  | 0.866 | -0.16 (-0.40, 0.07) |  | 0.175 |
| BCS * Wales | -0.08 (-0.34, 0.18) |  | 0.552 | -0.07 (-0.28, 0.14) |  | 0.517 | -0.01 (-0.25, 0.23) |  | 0.928 | -0.12 (-0.41, 0.18) |  | 0.438 |
| MCS * Scotland | -0.02 (-0.24, 0.20) |  | 0.861 | 0.15 (-0.07, 0.37) |  | 0.194 | 0.05 (-0.17, 0.27) |  | 0.659 | -0.09 (-0.37, 0.20) |  | 0.554 |
| MCS * Wales | 0.05 (-0.20, 0.30) |  | 0.700 | -0.05 (-0.31, 0.21) |  | 0.682 | 0.13 (-0.13, 0.39) |  | 0.329 | -0.02 (-0.35, 0.32) |  | 0.929 |

*Note.* Adjusted models included birth sex, highest qualification achieved, pre-pandemic self-reported health, pre-pandemic psychological distress, and household composition as covariates. Sensitivity models correspond to the unadjusted models after restricting the analytical sample to that of the adjusted models. BCS: British Cohort Study, 1970 birth cohort; GAD-2: 2-item General Anxiety Disorder questionnaire; MCS: Millennium Cohort Study, 2000 birth cohort; NCDS: National Child and Development Study, 1958 birth cohort; NS: Next Steps, 1990 cohort; NSHD: National Survey of Health and Development, 1946 birth cohort; ONS: UK Office for National Statistics; PHQ-2: 2-item Patient Health Questionnaire; UCLA-3: 3-item UCLA loneliness scale. χ2: Wald test performed to assess the overall statistical significance of the interaction terms; χ2 statistics in this table have 3 degrees of freedom in contrasts involving only country; 4 degrees of freedom in contrasts involving cohort only; and 6 degrees of freedom in contrasts involving both country and cohort. Due to differences in the samples, not all cohorts include all UK countries. Data from cohort*country combinations with small number of participants have been excluded from the analyses and tables to safeguard anonymity.

#### Table S11.2. Unadjusted and adjusted marginal mean estimates and 95% confidence intervals by UK country of residence.

|  |  |  | **Anxiety symptomatology (GAD-2)** | **Depressive symptomatology (PHQ-2)** | **Feelings of loneliness (UCLA-3)** | **Life satisfaction (ONS single question)** |
| --- | --- | --- | --- | --- | --- | --- |
| Cohort | UK country of residence | Survey wave | Unadjusted marginal mean (95% CI) | Unadjusted marginal mean (95% CI) | Unadjusted marginal mean (95% CI) | Unadjusted marginal mean (95% CI) |
| NSHD | England | 1 | 0.49 (0.43, 0.55) | 0.46 (0.40, 0.52) | 3.99 (3.91, 4.08) | 7.57 (7.44, 7.70) |
| NSHD | England | 2 | 0.64 (0.58, 0.71) | 0.51 (0.45, 0.56) | 3.86 (3.78, 3.93) | 7.46 (7.35, 7.57) |
| NSHD | England | 3 | 0.62 (0.55, 0.69) | 0.56 (0.50, 0.62) | 4.14 (4.06, 4.23) | 6.96 (6.84, 7.08) |
| NSHD | Scotland | 1 | 0.44 (0.23, 0.64) | 0.41 (0.19, 0.62) | 3.98 (3.65, 4.32) | 7.58 (7.17, 7.99) |
| NSHD | Scotland | 2 | 0.63 (0.46, 0.80) | 0.46 (0.30, 0.61) | 3.68 (3.45, 3.91) | 7.29 (6.94, 7.63) |
| NSHD | Scotland | 3 | 0.44 (0.28, 0.60) | 0.53 (0.34, 0.72) | 4.18 (3.88, 4.48) | 7.07 (6.71, 7.44) |
| NCDS | England | 1 | 0.74 (0.70, 0.78) | 0.67 (0.63, 0.71) | 4.15 (4.10, 4.19) | 7.42 (7.36, 7.49) |
| NCDS | England | 2 | 0.81 (0.77, 0.84) | 0.68 (0.64, 0.71) | 4.05 (4.01, 4.09) | 7.37 (7.32, 7.43) |
| NCDS | England | 3 | 0.79 (0.76, 0.83) | 0.80 (0.77, 0.84) | 4.32 (4.28, 4.36) | 6.92 (6.87, 6.98) |
| NCDS | Scotland | 1 | 0.74 (0.61, 0.86) | 0.62 (0.50, 0.73) | 4.10 (3.95, 4.24) | 7.63 (7.45, 7.80) |
| NCDS | Scotland | 2 | 0.76 (0.65, 0.88) | 0.73 (0.61, 0.84) | 4.07 (3.94, 4.20) | 7.46 (7.31, 7.61) |
| NCDS | Scotland | 3 | 0.74 (0.63, 0.85) | 0.83 (0.72, 0.94) | 4.30 (4.17, 4.43) | 7.16 (7.00, 7.32) |
| NCDS | Wales | 1 | 0.87 (0.67, 1.06) | 0.77 (0.61, 0.94) | 4.13 (3.95, 4.31) | 7.78 (7.57, 8.00) |
| NCDS | Wales | 2 | 0.74 (0.59, 0.89) | 0.70 (0.55, 0.85) | 3.99 (3.82, 4.16) | 7.60 (7.39, 7.81) |
| NCDS | Wales | 3 | 0.70 (0.56, 0.85) | 0.88 (0.73, 1.03) | 4.33 (4.16, 4.50) | 7.04 (6.82, 7.26) |
| BCS70 | England | 1 | 0.95 (0.91, 1.00) | 0.90 (0.85, 0.94) | 4.23 (4.18, 4.28) | 7.20 (7.13, 7.26) |
| BCS70 | England | 2 | 1.05 (1.00, 1.09) | 0.87 (0.83, 0.91) | 4.24 (4.19, 4.29) | 7.10 (7.04, 7.16) |
| BCS70 | England | 3 | 1.01 (0.97, 1.05) | 1.02 (0.98, 1.06) | 4.41 (4.36, 4.46) | 6.77 (6.71, 6.83) |
| BCS70 | Scotland | 1 | 0.83 (0.69, 0.97) | 0.86 (0.71, 1.01) | 4.02 (3.86, 4.18) | 7.28 (7.09, 7.48) |
| BCS70 | Scotland | 2 | 1.01 (0.87, 1.15) | 0.89 (0.76, 1.02) | 4.14 (3.98, 4.30) | 7.17 (6.98, 7.36) |
| BCS70 | Scotland | 3 | 0.94 (0.81, 1.06) | 1.05 (0.92, 1.18) | 4.30 (4.15, 4.46) | 6.81 (6.63, 6.99) |
| BCS70 | Wales | 1 | 1.01 (0.81, 1.21) | 1.01 (0.82, 1.19) | 4.28 (4.06, 4.49) | 7.10 (6.85, 7.36) |
| BCS70 | Wales | 2 | 1.25 (1.05, 1.45) | 1.06 (0.89, 1.24) | 4.35 (4.15, 4.55) | 7.02 (6.76, 7.28) |
| BCS70 | Wales | 3 | 1.21 (1.03, 1.39) | 1.31 (1.12, 1.49) | 4.69 (4.48, 4.90) | 6.58 (6.34, 6.82) |
| NS | England | 1 | 1.47 (1.40, 1.55) | 1.43 (1.36, 1.50) | 4.63 (4.56, 4.70) | 6.81 (6.72, 6.90) |
| NS | England | 2 | 1.61 (1.55, 1.67) | 1.30 (1.24, 1.35) | 4.64 (4.59, 4.70) | 6.92 (6.85, 6.99) |
| NS | England | 3 | 1.62 (1.57, 1.68) | 1.49 (1.44, 1.54) | 4.83 (4.78, 4.88) | 6.52 (6.45, 6.59) |
| MCS | England | 1 | 1.74 (1.65, 1.82) | 1.89 (1.81, 1.97) | 5.17 (5.09, 5.25) | 6.12 (6.02, 6.23) |
| MCS | England | 2 | 2.04 (1.96, 2.12) | 1.69 (1.62, 1.77) | 5.20 (5.13, 5.28) | 6.33 (6.24, 6.42) |
| MCS | England | 3 | 2.09 (2.02, 2.16) | 1.99 (1.92, 2.05) | 5.25 (5.18, 5.32) | 5.92 (5.84, 6.00) |
| MCS | N. Ireland | 1 | 1.84 (1.59, 2.09) | 1.62 (1.38, 1.86) | 5.03 (4.79, 5.27) | 6.62 (6.29, 6.94) |
| MCS | N. Ireland | 2 | 2.14 (1.92, 2.37) | 1.58 (1.36, 1.79) | 4.98 (4.73, 5.23) | 6.82 (6.55, 7.08) |
| MCS | N. Ireland | 3 | 2.00 (1.80, 2.20) | 1.88 (1.70, 2.07) | 5.23 (5.04, 5.42) | 6.10 (5.85, 6.35) |
| MCS | Scotland | 1 | 1.87 (1.66, 2.07) | 2.02 (1.82, 2.21) | 5.25 (5.05, 5.45) | 6.13 (5.86, 6.40) |
| MCS | Scotland | 2 | 2.13 (1.94, 2.32) | 1.69 (1.52, 1.87) | 5.29 (5.11, 5.47) | 6.29 (6.09, 6.50) |
| MCS | Scotland | 3 | 2.07 (1.91, 2.24) | 2.00 (1.85, 2.16) | 5.29 (5.13, 5.44) | 5.86 (5.67, 6.06) |
| MCS | Wales | 1 | 1.87 (1.69, 2.05) | 1.90 (1.71, 2.08) | 5.11 (4.92, 5.29) | 6.22 (5.96, 6.47) |
| MCS | Wales | 2 | 2.17 (1.97, 2.37) | 1.78 (1.59, 1.97) | 5.09 (4.90, 5.28) | 6.43 (6.19, 6.68) |
| MCS | Wales | 3 | 2.23 (2.06, 2.40) | 2.05 (1.89, 2.21) | 5.36 (5.19, 5.52) | 6.03 (5.82, 6.24) |
| Cohort | UK country of residence | Survey wave | Adjusted marginal mean (95% CI) | Adjusted marginal mean (95% CI) | Adjusted marginal mean (95% CI) | Adjusted marginal mean (95% CI) |
| NSHD | England | 1 | 0.59 (0.52, 0.66) | 0.57 (0.51, 0.64) | 4.21 (4.11, 4.31) | 7.27 (7.13, 7.40) |
| NSHD | England | 2 | 0.77 (0.69, 0.84) | 0.62 (0.55, 0.69) | 4.08 (3.98, 4.17) | 7.16 (7.02, 7.29) |
| NSHD | England | 3 | 0.74 (0.66, 0.82) | 0.68 (0.61, 0.75) | 4.35 (4.24, 4.45) | 6.64 (6.49, 6.79) |
| NSHD | Scotland | 1 | 0.55 (0.32, 0.78) | 0.54 (0.34, 0.74) | 4.27 (3.89, 4.64) | 7.31 (6.96, 7.67) |
| NSHD | Scotland | 2 | 0.76 (0.52, 0.99) | 0.61 (0.43, 0.79) | 3.79 (3.51, 4.07) | 7.06 (6.69, 7.43) |
| NSHD | Scotland | 3 | 0.61 (0.33, 0.89) | 0.61 (0.41, 0.81) | 4.25 (3.96, 4.55) | 6.96 (6.55, 7.37) |
| NCDS | England | 1 | 0.78 (0.74, 0.82) | 0.74 (0.70, 0.78) | 4.26 (4.22, 4.31) | 7.30 (7.23, 7.36) |
| NCDS | England | 2 | 0.84 (0.80, 0.88) | 0.74 (0.70, 0.77) | 4.16 (4.11, 4.20) | 7.25 (7.19, 7.30) |
| NCDS | England | 3 | 0.83 (0.79, 0.86) | 0.87 (0.83, 0.90) | 4.42 (4.38, 4.47) | 6.80 (6.75, 6.86) |
| NCDS | Scotland | 1 | 0.78 (0.66, 0.90) | 0.70 (0.59, 0.82) | 4.20 (4.08, 4.33) | 7.45 (7.28, 7.62) |
| NCDS | Scotland | 2 | 0.81 (0.70, 0.92) | 0.79 (0.68, 0.90) | 4.18 (4.06, 4.30) | 7.32 (7.17, 7.47) |
| NCDS | Scotland | 3 | 0.81 (0.71, 0.92) | 0.93 (0.82, 1.03) | 4.42 (4.29, 4.54) | 6.99 (6.83, 7.15) |
| NCDS | Wales | 1 | 0.79 (0.63, 0.94) | 0.81 (0.65, 0.97) | 4.26 (4.08, 4.44) | 7.63 (7.41, 7.84) |
| NCDS | Wales | 2 | 0.79 (0.65, 0.94) | 0.74 (0.58, 0.90) | 4.11 (3.94, 4.28) | 7.46 (7.26, 7.66) |
| NCDS | Wales | 3 | 0.68 (0.55, 0.81) | 0.87 (0.74, 1.01) | 4.42 (4.25, 4.59) | 6.91 (6.69, 7.12) |
| BCS70 | England | 1 | 0.93 (0.88, 0.98) | 0.93 (0.88, 0.98) | 4.32 (4.27, 4.38) | 7.11 (7.04, 7.18) |
| BCS70 | England | 2 | 1.04 (0.99, 1.09) | 0.90 (0.85, 0.95) | 4.32 (4.27, 4.38) | 7.02 (6.96, 7.09) |
| BCS70 | England | 3 | 0.97 (0.92, 1.01) | 1.02 (0.97, 1.06) | 4.48 (4.43, 4.53) | 6.71 (6.65, 6.78) |
| BCS70 | Scotland | 1 | 0.90 (0.75, 1.05) | 0.95 (0.79, 1.11) | 4.13 (3.97, 4.29) | 7.12 (6.91, 7.32) |
| BCS70 | Scotland | 2 | 0.98 (0.85, 1.12) | 0.88 (0.76, 1.01) | 4.21 (4.05, 4.36) | 7.08 (6.89, 7.27) |
| BCS70 | Scotland | 3 | 0.95 (0.82, 1.08) | 1.11 (0.97, 1.25) | 4.37 (4.21, 4.53) | 6.68 (6.49, 6.88) |
| BCS70 | Wales | 1 | 0.80 (0.58, 1.01) | 0.93 (0.75, 1.11) | 4.42 (4.20, 4.64) | 7.00 (6.73, 7.26) |
| BCS70 | Wales | 2 | 1.11 (0.92, 1.30) | 1.02 (0.86, 1.18) | 4.47 (4.27, 4.66) | 7.02 (6.75, 7.29) |
| BCS70 | Wales | 3 | 0.97 (0.80, 1.13) | 1.15 (0.99, 1.32) | 4.69 (4.49, 4.89) | 6.64 (6.39, 6.89) |
| NS | England | 1 | 1.35 (1.27, 1.42) | 1.36 (1.29, 1.44) | 4.59 (4.52, 4.66) | 6.89 (6.79, 6.98) |
| NS | England | 2 | 1.50 (1.44, 1.56) | 1.24 (1.19, 1.30) | 4.59 (4.53, 4.65) | 6.99 (6.92, 7.07) |
| NS | England | 3 | 1.48 (1.42, 1.54) | 1.40 (1.35, 1.46) | 4.77 (4.72, 4.83) | 6.61 (6.54, 6.68) |
| MCS | England | 1 | 1.64 (1.55, 1.74) | 1.77 (1.68, 1.86) | 4.80 (4.71, 4.90) | 6.57 (6.45, 6.69) |
| MCS | England | 2 | 1.98 (1.89, 2.07) | 1.62 (1.54, 1.71) | 4.89 (4.80, 4.99) | 6.73 (6.63, 6.84) |
| MCS | England | 3 | 2.02 (1.94, 2.10) | 1.91 (1.83, 1.98) | 4.93 (4.85, 5.01) | 6.30 (6.20, 6.40) |
| MCS | N. Ireland | 1 | 1.73 (1.49, 1.97) | 1.50 (1.26, 1.74) | 4.66 (4.42, 4.90) | 7.00 (6.66, 7.33) |
| MCS | N. Ireland | 2 | 2.05 (1.82, 2.28) | 1.48 (1.26, 1.70) | 4.61 (4.34, 4.89) | 7.17 (6.90, 7.44) |
| MCS | N. Ireland | 3 | 1.90 (1.71, 2.10) | 1.81 (1.63, 1.99) | 4.91 (4.72, 5.11) | 6.46 (6.21, 6.71) |
| MCS | Scotland | 1 | 1.77 (1.55, 1.99) | 1.92 (1.73, 2.12) | 4.94 (4.74, 5.14) | 6.52 (6.24, 6.79) |
| MCS | Scotland | 2 | 2.03 (1.85, 2.22) | 1.62 (1.44, 1.79) | 4.99 (4.81, 5.18) | 6.69 (6.47, 6.90) |
| MCS | Scotland | 3 | 2.01 (1.84, 2.18) | 1.97 (1.81, 2.13) | 4.97 (4.80, 5.13) | 6.26 (6.06, 6.46) |
| MCS | Wales | 1 | 1.73 (1.56, 1.90) | 1.78 (1.60, 1.97) | 4.73 (4.54, 4.92) | 6.61 (6.34, 6.88) |
| MCS | Wales | 2 | 2.01 (1.81, 2.21) | 1.63 (1.44, 1.81) | 4.68 (4.48, 4.88) | 6.86 (6.62, 7.10) |
| MCS | Wales | 3 | 2.12 (1.95, 2.30) | 1.95 (1.78, 2.12) | 5.00 (4.83, 5.17) | 6.40 (6.19, 6.62) |

*Note.* Adjusted models included birth sex, highest qualification achieved, pre-pandemic self-reported health, pre-pandemic psychological distress, and household composition as covariates. BCS: British Cohort Study, 1970 birth cohort; GAD-2: 2-item General Anxiety Disorder questionnaire; MCS: Millennium Cohort Study, 2000 birth cohort; NCDS: National Child and Development Study, 1958 birth cohort; NS: Next Steps, 1990 cohort; NSHD: National Survey of Health and Development, 1946 birth cohort; ONS: UK Office for National Statistics; PHQ-2: 2-item Patient Health Questionnaire; UCLA-3: 3-item UCLA loneliness scale. Survey wave 1: May 2020; survey wave 2: September/October 2020; survey wave 3: February/March 2021. Due to differences in the samples, not all cohorts include all UK countries. Data from cohort*country combinations with small number of participants have been excluded from the analyses and tables to safeguard anonymity.

#### Figure S11.1. Unadjusted and adjusted (by birth sex, highest qualification achieved, pre-pandemic self-reported health, pre-pandemic psychological distress, and household composition) anxiety symptomatology (GAD-2) marginal mean estimates and 95% confidence intervals by UK country of residence.

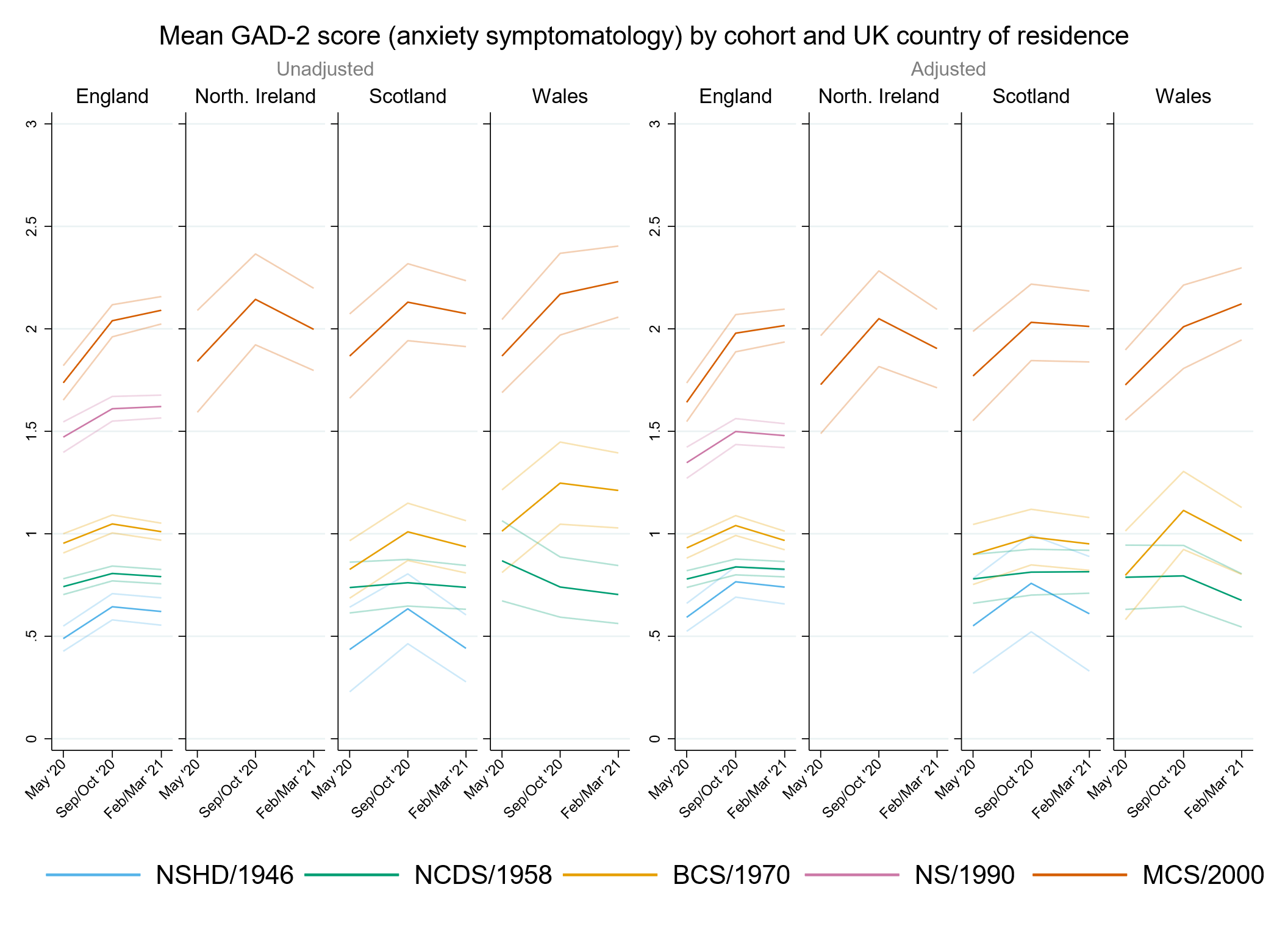

#### Figure S11.2. Unadjusted and adjusted (by birth sex, highest qualification achieved, pre-pandemic self-reported health, pre-pandemic psychological distress, and household composition) depressive symptomatology (PHQ-2) marginal mean estimates and 95% confidence intervals by UK country of residence.

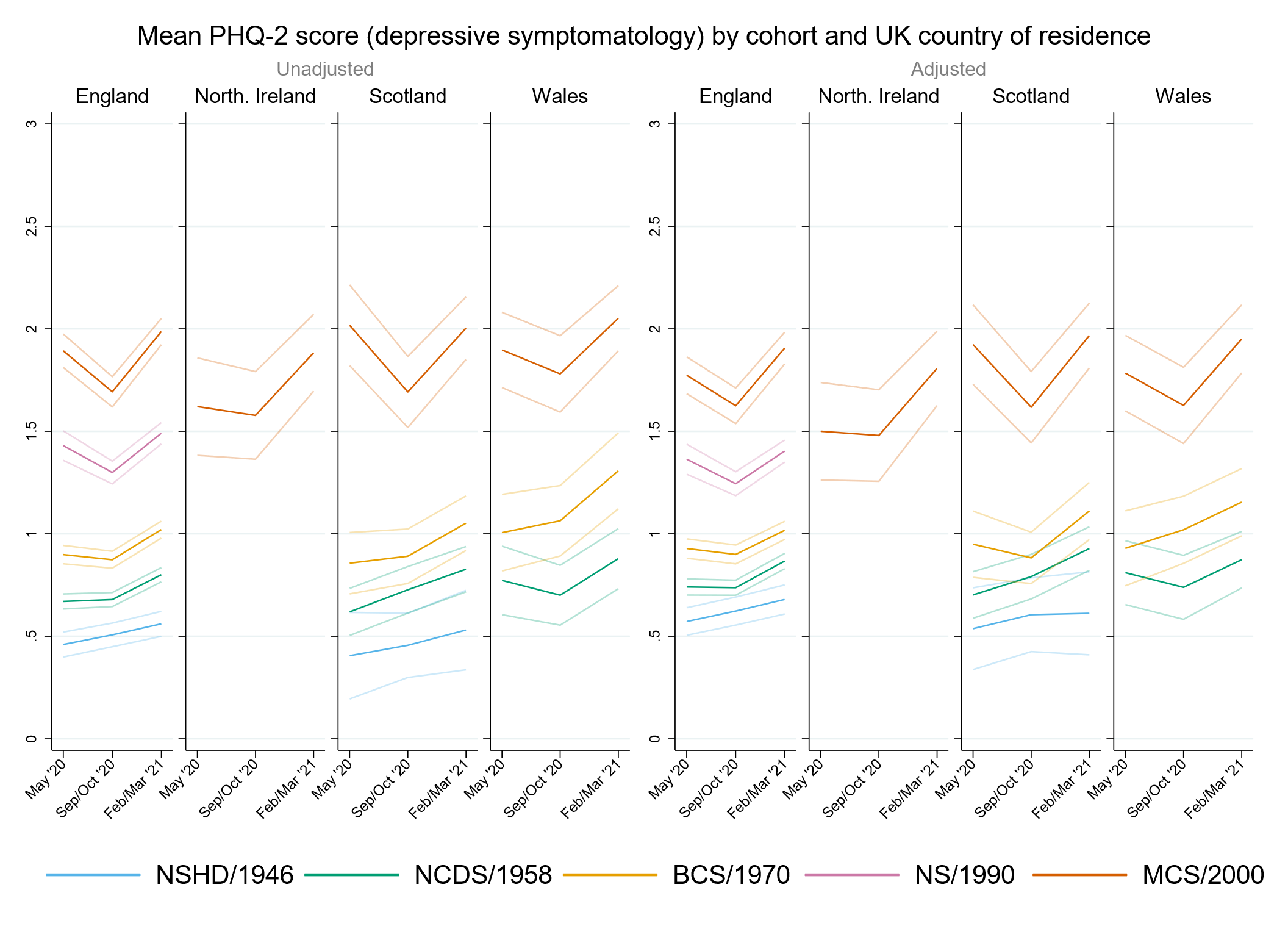

#### Figure S11.3. Unadjusted and adjusted (by birth sex, highest qualification achieved, pre-pandemic self-reported health, pre-pandemic psychological distress, and household composition) loneliness (UCLA-3) marginal mean estimates and 95% confidence intervals by UK country of residence.

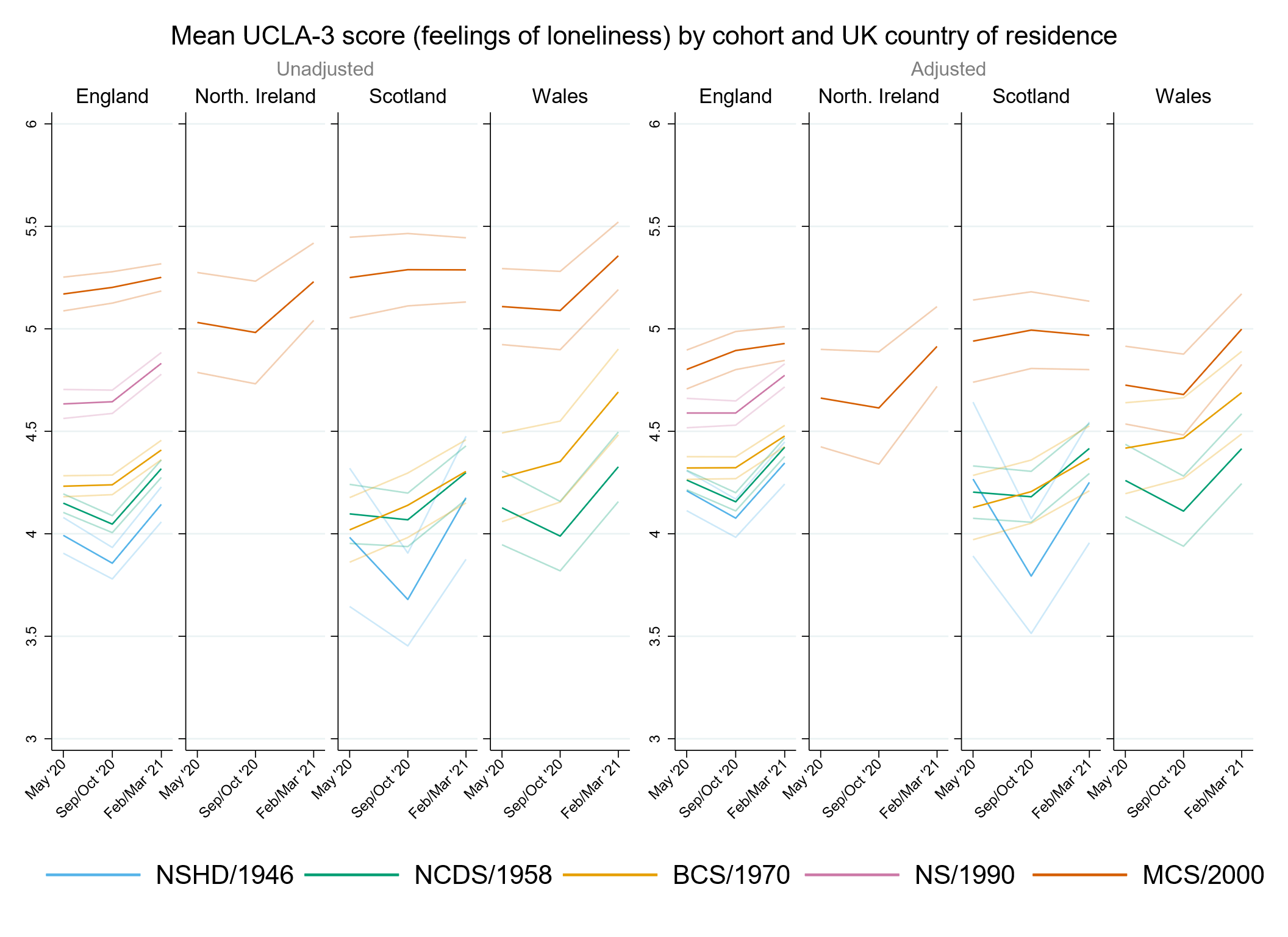

#### Figure S11.4. Unadjusted and adjusted (by birth sex, highest qualification achieved, pre-pandemic self-reported health, pre-pandemic psychological distress, and household composition) life satisfaction marginal mean estimates and 95% confidence intervals by UK country of residence.

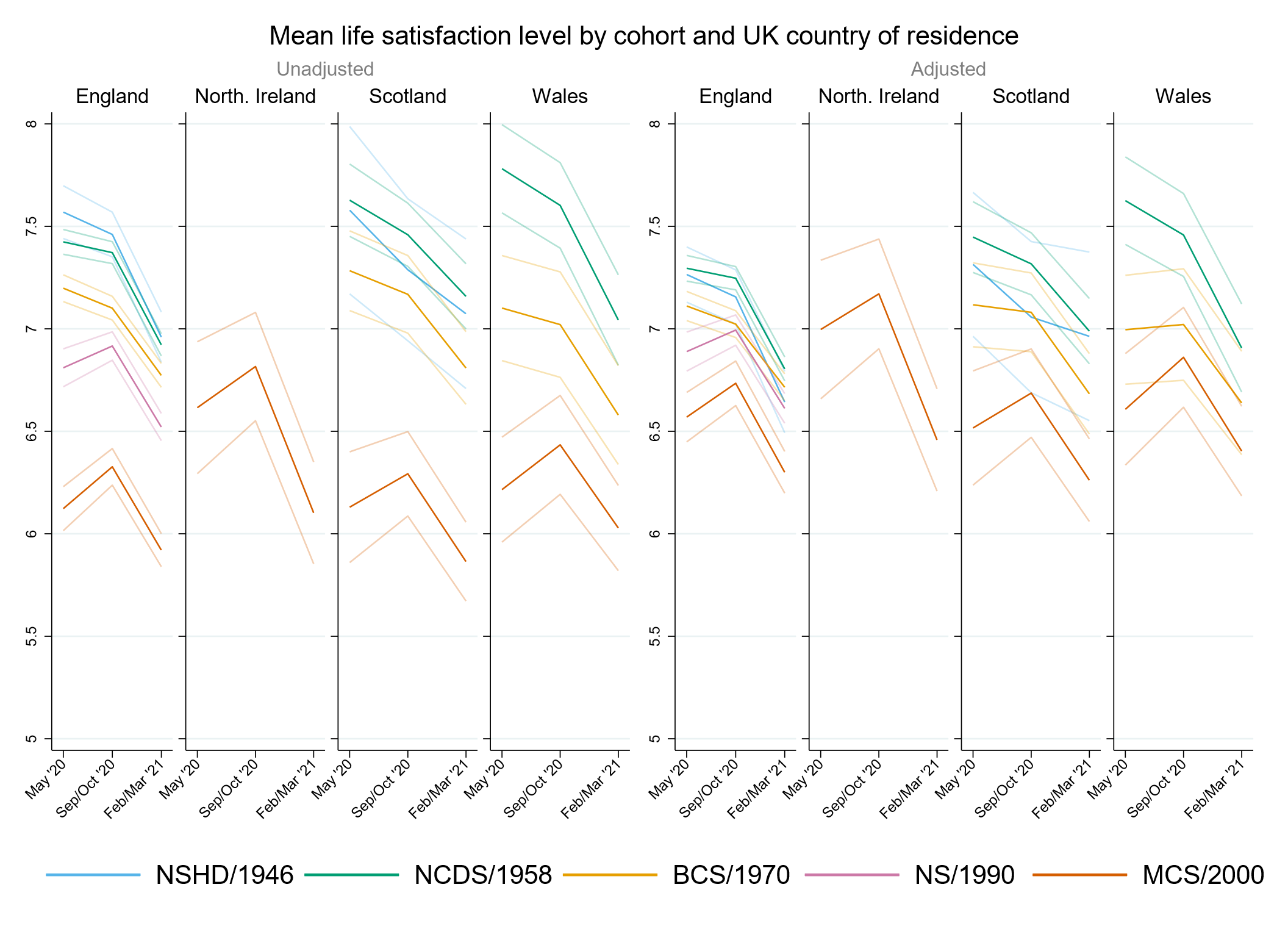

### Appendix S12. Results by ethnicity.

#### Table S12.1. Results of multilevel growth curve models by ethnicity.

|  | **Anxiety symptomatology (GAD-2)** | | | **Depressive symptomatology (PHQ-2)** | | | **Feelings of loneliness (UCLA-3)** | | | **Life satisfaction (ONS single question)** | | |
| --- | --- | --- | --- | --- | --- | --- | --- | --- | --- | --- | --- | --- |
| **Unadjusted models** |  | | |  | | |  | | |  | | |
| N participants | 9,902 |  |  | 9,900 |  |  | 9,910 |  |  | 9,950 |  |  |
| N observations | 18,557 |  |  | 18,551 |  |  | 18,610 |  |  | 18,691 |  |  |
|  | ***B* (95% CI)** | **χ2** | ***p*** | ***B* (95% CI)** | **χ2** | ***p*** | ***B* (95% CI)** | **χ2** | ***p*** | ***B* (95% CI)** | **χ2** | ***p*** |
| Time (linear) | 0.20 (0.06, 0.33) |  | 0.005 | -0.30 (-0.43, -0.17) |  | 0.000 | -0.09 (-0.22, 0.03) |  | 0.154 | 0.34 (0.18, 0.50) |  | 0.000 |
| Time (quadratic) | -0.06 (-0.12, 0.00) |  | 0.060 | 0.17 (0.12, 0.23) |  | 0.000 | 0.10 (0.04, 0.15) |  | 0.001 | -0.24 (-0.31, -0.17) |  | 0.000 |
| MCS cohort (ref. NS) | 0.29 (0.18, 0.40) |  | 0.000 | 0.47 (0.36, 0.57) |  | 0.000 | 0.57 (0.46, 0.67) |  | 0.000 | -0.66 (-0.80, -0.52) |  | 0.000 |
| Linear change * MCS cohort (ref. NS) | 0.23 (0.04, 0.43) |  | 0.021 | -0.19 (-0.38, 0.01) |  | 0.060 | 0.08 (-0.11, 0.27) |  | 0.409 | 0.21 (-0.03, 0.45) |  | 0.089 |
| Quadratic change * MCS cohort (ref. NS) | -0.08 (-0.17, 0.01) |  | 0.102 | 0.09 (0.00, 0.18) |  | 0.043 | -0.07 (-0.15, 0.02) |  | 0.128 | -0.09 (-0.20, 0.02) |  | 0.101 |
| Ethnicity (ref. White) |  | 8.5 | 0.075 |  | 0.7 | 0.949 |  | 3.1 | 0.549 |  | 3.1 | 0.543 |
| Mixed | -0.16 (-0.51, 0.18) |  | 0.355 | 0.04 (-0.26, 0.35) |  | 0.780 | 0.26 (-0.08, 0.60) |  | 0.136 | -0.18 (-0.65, 0.29) |  | 0.460 |
| Indian/Pakistani/Bangladeshi | -0.18 (-0.41, 0.05) |  | 0.116 | -0.04 (-0.26, 0.18) |  | 0.730 | 0.07 (-0.17, 0.30) |  | 0.568 | -0.23 (-0.52, 0.06) |  | 0.119 |
| Black Caribbean/Black African | -0.40 (-0.72, -0.08) |  | 0.014 | -0.07 (-0.51, 0.38) |  | 0.775 | 0.11 (-0.25, 0.48) |  | 0.546 | -0.15 (-0.61, 0.30) |  | 0.516 |
| Other | -0.15 (-0.65, 0.35) |  | 0.560 | -0.20 (-0.81, 0.40) |  | 0.506 | 0.15 (-0.27, 0.58) |  | 0.475 | -0.08 (-0.76, 0.60) |  | 0.824 |
| Linear change * ethnicity (ref. White) |  | 0.4 | 0.983 |  | 1.9 | 0.760 |  | 2.1 | 0.723 |  | 2.8 | 0.588 |
| Mixed | 0.01 (-0.64, 0.67) |  | 0.970 | -0.02 (-0.59, 0.54) |  | 0.935 | -0.01 (-0.58, 0.56) |  | 0.979 | -0.09 (-0.82, 0.64) |  | 0.808 |
| Indian/Pakistani/Bangladeshi | -0.06 (-0.47, 0.35) |  | 0.780 | 0.06 (-0.34, 0.45) |  | 0.786 | 0.05 (-0.35, 0.45) |  | 0.797 | 0.23 (-0.26, 0.72) |  | 0.357 |
| Black Caribbean/Black African | -0.16 (-0.73, 0.41) |  | 0.575 | -0.10 (-0.90, 0.70) |  | 0.806 | 0.18 (-0.45, 0.81) |  | 0.574 | -0.36 (-1.17, 0.45) |  | 0.383 |
| Other | -0.10 (-1.18, 0.99) |  | 0.862 | 0.80 (-0.39, 1.99) |  | 0.189 | 0.45 (-0.21, 1.12) |  | 0.183 | -0.57 (-1.68, 0.55) |  | 0.318 |
| Quadratic change * ethnicity (ref. White) |  | 0.2 | 0.995 |  | 2.4 | 0.663 |  | 2.0 | 0.74 |  | 1.9 | 0.759 |
| Mixed | 0.02 (-0.26, 0.30) |  | 0.887 | 0.01 (-0.24, 0.26) |  | 0.935 | -0.01 (-0.25, 0.23) |  | 0.944 | 0.03 (-0.28, 0.33) |  | 0.867 |
| Indian/Pakistani/Bangladeshi | 0.02 (-0.16, 0.20) |  | 0.820 | -0.07 (-0.24, 0.11) |  | 0.456 | -0.04 (-0.21, 0.13) |  | 0.619 | -0.06 (-0.27, 0.15) |  | 0.547 |
| Black Caribbean/Black African | 0.05 (-0.21, 0.30) |  | 0.721 | -0.01 (-0.35, 0.34) |  | 0.962 | -0.11 (-0.39, 0.16) |  | 0.422 | 0.16 (-0.19, 0.51) |  | 0.372 |
| Other | 0.05 (-0.42, 0.52) |  | 0.835 | -0.34 (-0.84, 0.15) |  | 0.169 | -0.17 (-0.48, 0.13) |  | 0.264 | 0.19 (-0.29, 0.67) |  | 0.437 |
| Cohort (ref. NS) * ethnicity (ref. White) |  | 1.5 | 0.835 |  | 1.7 | 0.797 |  | 7.3 | 0.121 |  | 0.4 | 0.986 |
| Mixed * MCS | 0.16 (-0.32, 0.65) |  | 0.513 | 0.19 (-0.27, 0.65) |  | 0.418 | -0.04 (-0.52, 0.45) |  | 0.884 | -0.03 (-0.63, 0.57) |  | 0.930 |
| Indian/Pakistani/Bangladeshi * MCS | -0.15 (-0.50, 0.21) |  | 0.418 | -0.14 (-0.49, 0.21) |  | 0.440 | -0.46 (-0.80, -0.11) |  | 0.009 | 0.08 (-0.43, 0.60) |  | 0.750 |
| Black Caribbean/Black African * MCS | 0.06 (-0.51, 0.64) |  | 0.830 | -0.16 (-0.80, 0.47) |  | 0.617 | -0.27 (-0.88, 0.34) |  | 0.387 | -0.03 (-0.87, 0.82) |  | 0.953 |
| Other * MCS | 0.18 (-0.55, 0.90) |  | 0.637 | 0.10 (-0.69, 0.90) |  | 0.799 | -0.05 (-0.73, 0.63) |  | 0.887 | -0.24 (-1.21, 0.73) |  | 0.632 |
| Linear change * cohort (ref. NS) * ethnicity (ref. White) |  | 0.6 | 0.961 |  | 3.6 | 0.471 |  | 2.6 | 0.629 |  | 2.0 | 0.742 |
| Mixed * MCS | -0.05 (-1.08, 0.97) |  | 0.917 | -0.34 (-1.25, 0.57) |  | 0.466 | 0.02 (-0.91, 0.95) |  | 0.965 | 0.05 (-0.88, 0.98) |  | 0.917 |
| Indian/Pakistani/Bangladeshi * MCS | 0.12 (-0.53, 0.77) |  | 0.717 | 0.54 (-0.12, 1.20) |  | 0.108 | 0.41 (-0.24, 1.06) |  | 0.220 | -0.60 (-1.64, 0.43) |  | 0.254 |
| Black Caribbean/Black African * MCS | -0.32 (-1.41, 0.76) |  | 0.560 | 0.39 (-0.97, 1.75) |  | 0.572 | -0.22 (-1.35, 0.92) |  | 0.707 | -0.43 (-2.01, 1.15) |  | 0.593 |
| Other * MCS | -0.24 (-1.70, 1.21) |  | 0.742 | 0.07 (-1.51, 1.64) |  | 0.931 | -0.56 (-1.77, 0.65) |  | 0.367 | 0.46 (-1.11, 2.02) |  | 0.567 |
| Quadratic change * cohort (ref. NS) * ethnicity (ref. White) |  | 1.1 | 0.889 |  | 2.9 | 0.567 |  | 1.7 | 0.79 |  | 2.4 | 0.66 |
| Mixed * MCS | 0.01 (-0.47, 0.49) |  | 0.966 | 0.16 (-0.27, 0.60) |  | 0.459 | -0.02 (-0.43, 0.39) |  | 0.930 | -0.03 (-0.45, 0.38) |  | 0.878 |
| Indian/Pakistani/Bangladeshi * MCS | -0.05 (-0.34, 0.24) |  | 0.736 | -0.22 (-0.52, 0.07) |  | 0.138 | -0.15 (-0.44, 0.15) |  | 0.330 | 0.30 (-0.15, 0.75) |  | 0.197 |
| Black Caribbean/Black African * MCS | 0.24 (-0.24, 0.73) |  | 0.327 | -0.07 (-0.71, 0.57) |  | 0.836 | 0.14 (-0.38, 0.66) |  | 0.593 | 0.25 (-0.44, 0.95) |  | 0.473 |
| Other * MCS | 0.05 (-0.61, 0.70) |  | 0.889 | -0.07 (-0.77, 0.63) |  | 0.845 | 0.17 (-0.37, 0.71) |  | 0.539 | -0.15 (-0.85, 0.54) |  | 0.668 |
| **Adjusted models** |  | | |  | | |  | | |  | | |
| N participants | 8,766 |  |  | 8,762 |  |  | 8,775 |  |  | 8,806 |  |  |
| N observations | 16,801 |  |  | 16,795 |  |  | 16,849 |  |  | 16,919 |  |  |
|  | ***B* (95% CI)** | **χ2** | ***p*** | ***B* (95% CI)** | **χ2** | ***p*** | ***B* (95% CI)** | **χ2** | ***p*** | ***B* (95% CI)** | **χ2** | ***p*** |
| Time (linear) | 0.21 (0.07, 0.35) |  | 0.004 | -0.28 (-0.42, -0.15) |  | 0 | -0.13 (-0.26, 0.00) |  | 0.050 | 0.37 (0.20, 0.54) |  | 0.000 |
| Time (quadratic) | -0.07 (-0.13, 0.00) |  | 0.038 | 0.16 (0.10, 0.22) |  | 0.000 | 0.11 (0.05, 0.17) |  | 0.000 | -0.25 (-0.33, -0.17) |  | 0.000 |
| MCS cohort (ref. NS) | 0.22 (0.09, 0.35) |  | 0.001 | 0.34 (0.22, 0.47) |  | 0.000 | 0.19 (0.06, 0.32) |  | 0.003 | -0.21 (-0.37, -0.04) |  | 0.015 |
| Linear change * MCS cohort (ref. NS) | 0.28 (0.07, 0.48) |  | 0.008 | -0.16 (-0.36, 0.04) |  | 0.119 | 0.16 (-0.04, 0.35) |  | 0.115 | 0.18 (-0.07, 0.43) |  | 0.157 |
| Quadratic change * MCS cohort (ref. NS) | -0.08 (-0.18, 0.01) |  | 0.081 | 0.09 (0.00, 0.19) |  | 0.042 | -0.09 (-0.18, 0.00) |  | 0.046 | -0.09 (-0.20, 0.02) |  | 0.109 |
| Ethnicity (ref. White) |  | 14.9 | 0.005 |  | 2.0 | 0.744 |  | 2.0 | 0.729 |  | 0.6 | 0.962 |
| Mixed | -0.32 (-0.68, 0.05) |  | 0.092 | -0.08 (-0.39, 0.22) |  | 0.591 | 0.06 (-0.28, 0.40) |  | 0.737 | -0.07 (-0.62, 0.48) |  | 0.812 |
| Indian/Pakistani/Bangladeshi | -0.19 (-0.43, 0.05) |  | 0.128 | -0.03 (-0.26, 0.20) |  | 0.815 | -0.11 (-0.34, 0.12) |  | 0.337 | 0.03 (-0.25, 0.31) |  | 0.850 |
| Black Caribbean/Black African | -0.55 (-0.87, -0.22) |  | 0.001 | -0.24 (-0.70, 0.21) |  | 0.295 | -0.20 (-0.61, 0.21) |  | 0.329 | 0.18 (-0.33, 0.69) |  | 0.483 |
| Other | -0.28 (-0.84, 0.28) |  | 0.330 | -0.28 (-0.95, 0.40) |  | 0.418 | -0.08 (-0.52, 0.36) |  | 0.717 | 0.07 (-0.67, 0.81) |  | 0.857 |
| Linear change * ethnicity (ref. White) |  | 0.3 | 0.993 |  | 3.3 | 0.514 |  | 5.0 | 0.292 |  | 1.3 | 0.868 |
| Mixed | 0.05 (-0.65, 0.76) |  | 0.881 | -0.04 (-0.64, 0.56) |  | 0.899 | -0.05 (-0.64, 0.54) |  | 0.866 | 0.01 (-0.87, 0.88) |  | 0.987 |
| Indian/Pakistani/Bangladeshi | 0.03 (-0.42, 0.48) |  | 0.904 | 0.11 (-0.32, 0.55) |  | 0.613 | 0.30 (-0.11, 0.71) |  | 0.157 | -0.07 (-0.58, 0.44) |  | 0.785 |
| Black Caribbean/Black African | -0.08 (-0.67, 0.51) |  | 0.781 | -0.47 (-1.21, 0.28) |  | 0.218 | 0.42 (-0.28, 1.13) |  | 0.242 | -0.45 (-1.38, 0.48) |  | 0.344 |
| Other | -0.22 (-1.38, 0.95) |  | 0.715 | 0.78 (-0.52, 2.07) |  | 0.239 | 0.54 (-0.22, 1.30) |  | 0.165 | -0.37 (-1.61, 0.86) |  | 0.555 |
| Quadratic change * ethnicity (ref. White) |  | 0.2 | 0.997 |  | 4.8 | 0.314 |  | 5.6 | 0.232 |  | 1.4 | 0.853 |
| Mixed | 0.00 (-0.30, 0.30) |  | 1.000 | 0.01 (-0.25, 0.28) |  | 0.914 | 0.04 (-0.21, 0.28) |  | 0.769 | -0.04 (-0.39, 0.32) |  | 0.839 |
| Indian/Pakistani/Bangladeshi | -0.02 (-0.22, 0.18) |  | 0.859 | -0.10 (-0.29, 0.09) |  | 0.312 | -0.15 (-0.32, 0.03) |  | 0.109 | 0.06 (-0.17, 0.28) |  | 0.625 |
| Black Caribbean/Black African | 0.02 (-0.25, 0.28) |  | 0.910 | 0.21 (-0.09, 0.51) |  | 0.176 | -0.22 (-0.53, 0.09) |  | 0.160 | 0.21 (-0.19, 0.61) |  | 0.308 |
| Other | 0.08 (-0.42, 0.59) |  | 0.743 | -0.34 (-0.86, 0.17) |  | 0.193 | -0.20 (-0.55, 0.14) |  | 0.250 | 0.07 (-0.45, 0.59) |  | 0.785 |
| Cohort (ref. NS) * ethnicity (ref. White) |  | 4.5 | 0.348 |  | 2.5 | 0.644 |  | 5.6 | 0.233 |  | 1.2 | 0.883 |
| Mixed * MCS | 0.40 (-0.11, 0.90) |  | 0.123 | 0.36 (-0.09, 0.81) |  | 0.118 | 0.07 (-0.39, 0.53) |  | 0.769 | -0.05 (-0.71, 0.62) |  | 0.884 |
| Indian/Pakistani/Bangladeshi * MCS | 0.02 (-0.37, 0.41) |  | 0.916 | 0.01 (-0.36, 0.39) |  | 0.951 | -0.35 (-0.68, -0.02) |  | 0.038 | -0.13 (-0.69, 0.42) |  | 0.636 |
| Black Caribbean/Black African * MCS | 0.25 (-0.37, 0.87) |  | 0.427 | 0.08 (-0.58, 0.74) |  | 0.811 | 0.12 (-0.52, 0.76) |  | 0.712 | -0.22 (-1.07, 0.63) |  | 0.608 |
| Other * MCS | 0.56 (-0.29, 1.40) |  | 0.195 | 0.09 (-0.86, 1.04) |  | 0.853 | 0.33 (-0.44, 1.09) |  | 0.404 | -0.49 (-1.61, 0.63) |  | 0.389 |
| Linear change * cohort (ref. NS) * ethnicity (ref. White) |  | 1.6 | 0.801 |  | 1.4 | 0.837 |  | 2.1 | 0.72 |  | 1.7 | 0.792 |
| Mixed * MCS | -0.09 (-1.20, 1.02) |  | 0.878 | -0.18 (-1.18, 0.83) |  | 0.732 | 0.16 (-0.82, 1.13) |  | 0.754 | -0.05 (-1.13, 1.04) |  | 0.932 |
| Indian/Pakistani/Bangladeshi * MCS | -0.25 (-1.00, 0.49) |  | 0.508 | 0.31 (-0.41, 1.04) |  | 0.395 | 0.39 (-0.29, 1.07) |  | 0.257 | -0.43 (-1.58, 0.72) |  | 0.465 |
| Black Caribbean/Black African * MCS | -0.51 (-1.68, 0.65) |  | 0.388 | 0.47 (-0.93, 1.86) |  | 0.514 | -0.39 (-1.63, 0.85) |  | 0.538 | -0.66 (-2.38, 1.06) |  | 0.451 |
| Other * MCS | -0.58 (-2.10, 0.93) |  | 0.451 | -0.37 (-2.26, 1.52) |  | 0.702 | -0.40 (-1.95, 1.16) |  | 0.618 | 0.72 (-1.16, 2.60) |  | 0.451 |
| Quadratic change * cohort (ref. NS) * ethnicity (ref. White) |  | 2.4 | 0.664 |  | 1.3 | 0.855 |  | 1.8 | 0.767 |  | 2.0 | 0.743 |
| Mixed * MCS | -0.03 (-0.54, 0.49) |  | 0.922 | 0.06 (-0.43, 0.54) |  | 0.817 | -0.10 (-0.54, 0.35) |  | 0.678 | 0.03 (-0.44, 0.50) |  | 0.913 |
| Indian/Pakistani/Bangladeshi * MCS | 0.08 (-0.25, 0.41) |  | 0.627 | -0.17 (-0.49, 0.16) |  | 0.315 | -0.16 (-0.48, 0.15) |  | 0.306 | 0.26 (-0.24, 0.77) |  | 0.308 |
| Black Caribbean/Black African * MCS | 0.37 (-0.15, 0.88) |  | 0.162 | -0.11 (-0.77, 0.54) |  | 0.735 | 0.21 (-0.34, 0.76) |  | 0.455 | 0.33 (-0.43, 1.10) |  | 0.395 |
| Other * MCS | 0.18 (-0.48, 0.85) |  | 0.590 | 0.15 (-0.68, 0.99) |  | 0.716 | 0.01 (-0.70, 0.71) |  | 0.988 | -0.20 (-1.05, 0.65) |  | 0.647 |
| **Sensitivity models** |  | | |  | | |  | | |  | | |
| N participants | 8,766 |  |  | 8,762 |  |  | 8,775 |  |  | 8,806 |  |  |
| N observations | 16,801 |  |  | 16,795 |  |  | 16,849 |  |  | 16,919 |  |  |
|  | ***B* (95% CI)** | **χ2** | ***p*** | ***B* (95% CI)** | **χ2** | ***p*** | ***B* (95% CI)** | **χ2** | ***p*** | ***B* (95% CI)** | **χ2** | ***p*** |
| Time (linear) | 0.19 (-0.51, 0.89) |  | 0.602 | -0.36 (-0.98, 0.26) |  | 0.257 | -0.19 (-0.75, 0.37) |  | 0.510 | 0.34 (-0.51, 1.19) |  | 0.428 |
| Time (quadratic) | -0.03 (-0.33, 0.26) |  | 0.825 | 0.19 (-0.08, 0.46) |  | 0.166 | 0.15 (-0.09, 0.39) |  | 0.217 | -0.28 (-0.62, 0.07) |  | 0.118 |
| MCS cohort (ref. NS) | 0.54 (0.02, 1.05) |  | 0.040 | 0.71 (0.23, 1.19) |  | 0.004 | 0.50 (0.04, 0.95) |  | 0.032 | -0.60 (-1.26, 0.06) |  | 0.076 |
| Linear change * MCS cohort (ref. NS) | 0.25 (-0.86, 1.36) |  | 0.664 | -0.33 (-1.35, 0.68) |  | 0.517 | 0.28 (-0.67, 1.23) |  | 0.561 | 0.22 (-0.83, 1.27) |  | 0.679 |
| Quadratic change * MCS cohort (ref. NS) | -0.13 (-0.65, 0.39) |  | 0.620 | 0.15 (-0.33, 0.63) |  | 0.536 | -0.17 (-0.61, 0.27) |  | 0.449 | -0.10 (-0.56, 0.36) |  | 0.670 |
| Ethnicity (ref. White) |  | 6.1 | 0.194 |  | 0.6 | 0.965 |  | 2.2 | 0.694 |  | 3.2 | 0.533 |
| Mixed | 0.19 (-0.19, 0.57) |  | 0.317 | -0.02 (-0.36, 0.33) |  | 0.918 | -0.19 (-0.52, 0.13) |  | 0.244 | 0.20 (-0.36, 0.75) |  | 0.487 |
| Indian/Pakistani/Bangladeshi | 0.07 (-0.37, 0.50) |  | 0.763 | 0.04 (-0.36, 0.44) |  | 0.849 | -0.12 (-0.52, 0.28) |  | 0.554 | -0.03 (-0.63, 0.58) |  | 0.930 |
| Black Caribbean/Black African | -0.19 (-0.69, 0.31) |  | 0.463 | -0.10 (-0.67, 0.48) |  | 0.743 | -0.07 (-0.58, 0.44) |  | 0.793 | 0.01 (-0.74, 0.75) |  | 0.985 |
| Other | 0.07 (-0.57, 0.70) |  | 0.837 | -0.18 (-0.93, 0.58) |  | 0.645 | -0.03 (-0.59, 0.53) |  | 0.910 | -0.03 (-0.95, 0.90) |  | 0.953 |
| Linear change * ethnicity (ref. White) |  | 0.5 | 0.974 |  | 3.9 | 0.42 |  | 3.7 | 0.451 |  | 1.5 | 0.819 |
| Mixed | 0.04 (-0.68, 0.75) |  | 0.923 | 0.10 (-0.54, 0.73) |  | 0.765 | 0.09 (-0.49, 0.67) |  | 0.761 | -0.01 (-0.87, 0.86) |  | 0.989 |
| Indian/Pakistani/Bangladeshi | -0.03 (-0.86, 0.79) |  | 0.936 | 0.09 (-0.65, 0.84) |  | 0.804 | 0.27 (-0.42, 0.97) |  | 0.439 | 0.12 (-0.86, 1.11) |  | 0.810 |
| Black Caribbean/Black African | -0.16 (-1.08, 0.75) |  | 0.725 | -0.47 (-1.42, 0.49) |  | 0.340 | 0.38 (-0.51, 1.27) |  | 0.403 | -0.35 (-1.60, 0.90) |  | 0.583 |
| Other | -0.06 (-1.43, 1.30) |  | 0.926 | 0.94 (-0.52, 2.40) |  | 0.208 | 0.70 (-0.25, 1.65) |  | 0.146 | -0.55 (-2.03, 0.94) |  | 0.471 |
| Quadratic change * ethnicity (ref. White) |  | 0.3 | 0.989 |  | 4.9 | 0.295 |  | 3.9 | 0.415 |  | 1.1 | 0.903 |
| Mixed | -0.04 (-0.35, 0.26) |  | 0.789 | -0.04 (-0.32, 0.23) |  | 0.767 | -0.05 (-0.30, 0.19) |  | 0.663 | 0.04 (-0.31, 0.40) |  | 0.823 |
| Indian/Pakistani/Bangladeshi | -0.02 (-0.37, 0.33) |  | 0.922 | -0.09 (-0.42, 0.23) |  | 0.580 | -0.16 (-0.45, 0.14) |  | 0.299 | 0.02 (-0.39, 0.43) |  | 0.922 |
| Black Caribbean/Black African | 0.02 (-0.38, 0.42) |  | 0.921 | 0.20 (-0.19, 0.60) |  | 0.306 | -0.23 (-0.62, 0.16) |  | 0.250 | 0.21 (-0.32, 0.74) |  | 0.431 |
| Other | 0.00 (-0.60, 0.60) |  | 0.996 | -0.40 (-0.99, 0.20) |  | 0.191 | -0.28 (-0.69, 0.14) |  | 0.195 | 0.17 (-0.44, 0.79) |  | 0.582 |
| Cohort (ref. NS) * ethnicity (ref. White) |  | 2.5 | 0.641 |  | 1.5 | 0.82 |  | 9.9 | 0.041 |  | 0.6 | 0.962 |
| Mixed * MCS | -0.24 (-0.77, 0.28) |  | 0.368 | -0.25 (-0.74, 0.25) |  | 0.332 | 0.06 (-0.41, 0.53) |  | 0.806 | -0.05 (-0.73, 0.63) |  | 0.880 |
| Indian/Pakistani/Bangladeshi * MCS | -0.35 (-0.99, 0.29) |  | 0.284 | -0.35 (-0.96, 0.25) |  | 0.255 | -0.50 (-1.08, 0.07) |  | 0.083 | 0.09 (-0.77, 0.95) |  | 0.839 |
| Black Caribbean/Black African * MCS | -0.24 (-1.07, 0.58) |  | 0.562 | -0.39 (-1.22, 0.43) |  | 0.352 | -0.19 (-0.99, 0.62) |  | 0.652 | 0.08 (-1.06, 1.22) |  | 0.889 |
| Other * MCS | 0.26 (-0.72, 1.23) |  | 0.604 | -0.16 (-1.20, 0.89) |  | 0.768 | 0.24 (-0.66, 1.14) |  | 0.595 | -0.35 (-1.67, 0.97) |  | 0.604 |
| Linear change * cohort (ref. NS) * ethnicity (ref. White) |  | 1.9 | 0.757 |  | 1.9 | 0.759 |  | 2.7 | 0.618 |  | 2.1 | 0.726 |
| Mixed * MCS | 0.01 (-1.12, 1.14) |  | 0.981 | 0.16 (-0.88, 1.19) |  | 0.767 | -0.15 (-1.12, 0.82) |  | 0.762 | -0.03 (-1.11, 1.05) |  | 0.958 |
| Indian/Pakistani/Bangladeshi * MCS | -0.16 (-1.49, 1.17) |  | 0.813 | 0.56 (-0.66, 1.79) |  | 0.369 | 0.33 (-0.83, 1.48) |  | 0.582 | -0.59 (-2.14, 0.97) |  | 0.459 |
| Black Caribbean/Black African * MCS | -0.55 (-2.19, 1.09) |  | 0.512 | 0.60 (-1.14, 2.34) |  | 0.498 | -0.54 (-2.10, 1.02) |  | 0.497 | -0.61 (-2.68, 1.47) |  | 0.566 |
| Other * MCS | -0.76 (-2.63, 1.12) |  | 0.431 | -0.25 (-2.42, 1.93) |  | 0.824 | -0.60 (-2.44, 1.23) |  | 0.519 | 0.82 (-1.35, 2.98) |  | 0.460 |
| Quadratic change * cohort (ref. NS) * ethnicity (ref. White) |  | 2.5 | 0.647 |  | 1.6 | 0.805 |  | 2.1 | 0.718 |  | 2.1 | 0.717 |
| Mixed * MCS | 0.05 (-0.48, 0.58) |  | 0.857 | -0.05 (-0.54, 0.44) |  | 0.829 | 0.09 (-0.36, 0.53) |  | 0.710 | 0.01 (-0.46, 0.48) |  | 0.972 |
| Indian/Pakistani/Bangladeshi * MCS | 0.10 (-0.51, 0.71) |  | 0.745 | -0.25 (-0.82, 0.33) |  | 0.397 | -0.10 (-0.64, 0.43) |  | 0.707 | 0.31 (-0.37, 0.98) |  | 0.375 |
| Black Caribbean/Black African * MCS | 0.43 (-0.31, 1.17) |  | 0.252 | -0.16 (-0.98, 0.66) |  | 0.700 | 0.30 (-0.41, 1.01) |  | 0.403 | 0.31 (-0.60, 1.23) |  | 0.503 |
| Other * MCS | 0.28 (-0.56, 1.13) |  | 0.511 | 0.07 (-0.91, 1.06) |  | 0.887 | 0.08 (-0.76, 0.91) |  | 0.856 | -0.21 (-1.18, 0.77) |  | 0.674 |

*Note.* Adjusted models included birth sex, highest qualification achieved, pre-pandemic self-reported health, pre-pandemic psychological distress, and household composition as covariates. Sensitivity models correspond to the unadjusted models after restricting the analytical sample to that of the adjusted models. GAD-2: 2-item General Anxiety Disorder questionnaire; MCS: Millennium Cohort Study, 2000 birth cohort; NS: Next Steps, 1990 cohort; ONS: UK Office for National Statistics; PHQ-2: 2-item Patient Health Questionnaire; UCLA-3: 3-item UCLA loneliness scale. χ2: Wald test performed to assess the overall statistical significance of the interaction terms; all χ2 statistics in this table have 4 degrees of freedom.

#### Table S12.2. Unadjusted and adjusted marginal mean estimates and 95% confidence intervals by ethnicity.

|  |  |  | **Anxiety symptomatology (GAD-2)** | **Depressive symptomatology (PHQ-2)** | **Feelings of loneliness (UCLA-3)** | **Life satisfaction (ONS single question)** |
| --- | --- | --- | --- | --- | --- | --- |
| **Cohort** | **Ethnicity** | **Survey wave** | **Unadjusted marginal mean (95% CI)** | **Unadjusted marginal mean (95% CI)** | **Unadjusted marginal mean (95% CI)** | **Unadjusted marginal mean (95% CI)** |
| NS | White | 1 | 1.36 (1.03, 1.70) | 1.48 (1.19, 1.78) | 4.87 (4.55, 5.20) | 6.68 (6.22, 7.14) |
| NS | White | 2 | 1.53 (1.28, 1.79) | 1.34 (1.10, 1.58) | 4.86 (4.59, 5.13) | 6.71 (6.40, 7.03) |
| NS | White | 3 | 1.63 (1.37, 1.88) | 1.57 (1.32, 1.81) | 5.02 (4.77, 5.27) | 6.31 (6.02, 6.61) |
| NS | Mixed | 1 | 1.34 (1.13, 1.56) | 1.40 (1.19, 1.60) | 4.69 (4.46, 4.91) | 6.62 (6.35, 6.90) |
| NS | Mixed | 2 | 1.44 (1.28, 1.60) | 1.26 (1.10, 1.41) | 4.70 (4.54, 4.85) | 6.89 (6.70, 7.08) |
| NS | Mixed | 3 | 1.47 (1.33, 1.60) | 1.34 (1.21, 1.46) | 4.82 (4.68, 4.96) | 6.54 (6.36, 6.72) |
| NS | Indian/Pakistani/Bangladeshi | 1 | 1.12 (0.82, 1.43) | 1.37 (0.93, 1.81) | 4.73 (4.37, 5.09) | 6.70 (6.26, 7.15) |
| NS | Indian/Pakistani/Bangladeshi | 2 | 1.15 (0.92, 1.37) | 1.14 (0.85, 1.42) | 4.80 (4.52, 5.08) | 6.60 (6.25, 6.96) |
| NS | Indian/Pakistani/Bangladeshi | 3 | 1.14 (0.91, 1.38) | 1.23 (0.99, 1.47) | 4.83 (4.57, 5.10) | 6.34 (6.02, 6.66) |
| NS | Black Caribbean/Black African | 1 | 1.38 (0.89, 1.87) | 1.23 (0.63, 1.83) | 4.77 (4.36, 5.19) | 6.78 (6.10, 7.45) |
| NS | Black Caribbean/Black African | 2 | 1.47 (1.07, 1.87) | 1.56 (1.15, 1.97) | 5.06 (4.69, 5.43) | 6.50 (5.94, 7.06) |
| NS | Black Caribbean/Black African | 3 | 1.54 (1.20, 1.89) | 1.54 (1.19, 1.90) | 5.19 (4.82, 5.55) | 6.12 (5.69, 6.55) |
| NS | Other | 1 | 1.81 (1.74, 1.89) | 1.90 (1.83, 1.97) | 5.18 (5.11, 5.26) | 6.19 (6.10, 6.29) |
| NS | Other | 2 | 2.11 (2.04, 2.18) | 1.68 (1.61, 1.74) | 5.20 (5.13, 5.27) | 6.41 (6.33, 6.49) |
| NS | Other | 3 | 2.14 (2.07, 2.20) | 1.98 (1.93, 2.04) | 5.28 (5.21, 5.34) | 5.97 (5.90, 6.04) |
| MCS | White | 1 | 1.81 (1.48, 2.15) | 2.13 (1.80, 2.47) | 5.40 (5.06, 5.75) | 5.99 (5.63, 6.35) |
| MCS | White | 2 | 2.10 (1.71, 2.48) | 1.72 (1.38, 2.06) | 5.41 (5.06, 5.75) | 6.16 (5.82, 6.50) |
| MCS | White | 3 | 2.18 (1.89, 2.46) | 2.19 (1.91, 2.46) | 5.41 (5.16, 5.67) | 5.66 (5.34, 5.97) |
| MCS | Mixed | 1 | 1.48 (1.23, 1.74) | 1.73 (1.46, 1.99) | 4.79 (4.55, 5.03) | 6.05 (5.63, 6.46) |
| MCS | Mixed | 2 | 1.81 (1.59, 2.04) | 1.81 (1.58, 2.04) | 5.08 (4.84, 5.32) | 6.13 (5.80, 6.45) |
| MCS | Mixed | 3 | 1.81 (1.63, 2.00) | 1.84 (1.66, 2.02) | 5.05 (4.86, 5.23) | 6.01 (5.76, 6.25) |
| MCS | Indian/Pakistani/Bangladeshi | 1 | 1.47 (1.00, 1.94) | 1.67 (1.23, 2.12) | 5.03 (4.55, 5.51) | 6.02 (5.31, 6.72) |
| MCS | Indian/Pakistani/Bangladeshi | 2 | 1.57 (1.15, 2.00) | 1.67 (1.15, 2.18) | 5.03 (4.58, 5.49) | 5.86 (5.32, 6.39) |
| MCS | Indian/Pakistani/Bangladeshi | 3 | 1.98 (1.64, 2.32) | 2.04 (1.64, 2.43) | 5.16 (4.82, 5.50) | 5.87 (5.40, 6.33) |
| MCS | Black Caribbean/Black African | 1 | 1.84 (1.31, 2.37) | 1.80 (1.29, 2.31) | 5.29 (4.77, 5.81) | 5.88 (5.20, 6.56) |
| MCS | Black Caribbean/Black African | 2 | 1.89 (1.43, 2.36) | 2.03 (1.54, 2.53) | 5.20 (4.76, 5.63) | 6.03 (5.52, 6.53) |
| MCS | Black Caribbean/Black African | 3 | 1.87 (1.51, 2.24) | 1.96 (1.61, 2.31) | 5.16 (4.83, 5.50) | 5.59 (5.10, 6.08) |
| MCS | Other | 1 | 1.36 (1.03, 1.70) | 1.48 (1.19, 1.78) | 4.87 (4.55, 5.20) | 6.68 (6.22, 7.14) |
| MCS | Other | 2 | 1.53 (1.28, 1.79) | 1.34 (1.10, 1.58) | 4.86 (4.59, 5.13) | 6.71 (6.40, 7.03) |
| MCS | Other | 3 | 1.63 (1.37, 1.88) | 1.57 (1.32, 1.81) | 5.02 (4.77, 5.27) | 6.31 (6.02, 6.61) |
| **Cohort** | **Ethnicity** | **Survey wave** | **Adjusted marginal mean (95% CI)** | **Adjusted marginal mean (95% CI)** | **Adjusted marginal mean (95% CI)** | **Adjusted marginal mean (95% CI)** |
| NS | White | 1 | 1.47 (1.38, 1.57) | 1.47 (1.38, 1.55) | 4.81 (4.72, 4.90) | 6.62 (6.50, 6.73) |
| NS | White | 2 | 1.62 (1.54, 1.70) | 1.34 (1.27, 1.41) | 4.79 (4.71, 4.87) | 6.74 (6.64, 6.83) |
| NS | White | 3 | 1.62 (1.54, 1.70) | 1.53 (1.46, 1.60) | 4.99 (4.91, 5.06) | 6.36 (6.26, 6.45) |
| NS | Mixed | 1 | 1.16 (0.80, 1.52) | 1.38 (1.08, 1.68) | 4.87 (4.54, 5.20) | 6.55 (6.01, 7.10) |
| NS | Mixed | 2 | 1.35 (1.09, 1.62) | 1.23 (0.98, 1.48) | 4.83 (4.57, 5.09) | 6.64 (6.31, 6.97) |
| NS | Mixed | 3 | 1.41 (1.16, 1.66) | 1.43 (1.16, 1.70) | 5.09 (4.83, 5.35) | 6.16 (5.82, 6.50) |
| NS | Indian/Pakistani/Bangladeshi | 1 | 1.29 (1.06, 1.51) | 1.44 (1.22, 1.66) | 4.70 (4.48, 4.92) | 6.64 (6.38, 6.91) |
| NS | Indian/Pakistani/Bangladeshi | 2 | 1.44 (1.26, 1.61) | 1.33 (1.16, 1.50) | 4.83 (4.66, 5.00) | 6.75 (6.54, 6.95) |
| NS | Indian/Pakistani/Bangladeshi | 3 | 1.42 (1.28, 1.56) | 1.33 (1.21, 1.46) | 4.89 (4.74, 5.04) | 6.46 (6.27, 6.66) |
| NS | Black Caribbean/Black African | 1 | 0.93 (0.61, 1.24) | 1.22 (0.77, 1.67) | 4.61 (4.20, 5.01) | 6.80 (6.30, 7.30) |
| NS | Black Caribbean/Black African | 2 | 1.00 (0.75, 1.25) | 0.84 (0.61, 1.07) | 4.79 (4.49, 5.08) | 6.68 (6.31, 7.05) |
| NS | Black Caribbean/Black African | 3 | 0.97 (0.70, 1.24) | 1.18 (0.93, 1.44) | 4.74 (4.44, 5.05) | 6.48 (6.11, 6.86) |
| NS | Other | 1 | 1.20 (0.64, 1.75) | 1.19 (0.51, 1.86) | 4.73 (4.30, 5.16) | 6.69 (5.95, 7.42) |
| NS | Other | 2 | 1.21 (0.83, 1.58) | 1.49 (1.11, 1.88) | 5.04 (4.66, 5.43) | 6.50 (5.94, 7.06) |
| NS | Other | 3 | 1.25 (0.85, 1.65) | 1.43 (1.07, 1.79) | 5.17 (4.79, 5.55) | 5.97 (5.55, 6.39) |
| MCS | White | 1 | 1.70 (1.62, 1.78) | 1.81 (1.73, 1.88) | 5.00 (4.93, 5.08) | 6.41 (6.31, 6.51) |
| MCS | White | 2 | 2.04 (1.96, 2.11) | 1.62 (1.55, 1.69) | 5.05 (4.97, 5.13) | 6.62 (6.53, 6.71) |
| MCS | White | 3 | 2.07 (2.00, 2.14) | 1.94 (1.87, 2.00) | 5.13 (5.06, 5.20) | 6.15 (6.06, 6.23) |
| MCS | Mixed | 1 | 1.78 (1.44, 2.12) | 2.09 (1.76, 2.41) | 5.13 (4.82, 5.44) | 6.30 (5.93, 6.66) |
| MCS | Mixed | 2 | 2.06 (1.67, 2.45) | 1.75 (1.38, 2.13) | 5.22 (4.83, 5.61) | 6.45 (6.10, 6.80) |
| MCS | Mixed | 3 | 1.98 (1.70, 2.26) | 2.07 (1.79, 2.35) | 5.23 (4.97, 5.50) | 5.91 (5.60, 6.21) |
| MCS | Indian/Pakistani/Bangladeshi | 1 | 1.53 (1.23, 1.83) | 1.79 (1.50, 2.08) | 4.54 (4.31, 4.77) | 6.30 (5.84, 6.77) |
| MCS | Indian/Pakistani/Bangladeshi | 2 | 1.71 (1.46, 1.95) | 1.77 (1.51, 2.02) | 4.96 (4.70, 5.22) | 6.33 (5.97, 6.70) |
| MCS | Indian/Pakistani/Bangladeshi | 3 | 1.71 (1.51, 1.91) | 1.71 (1.51, 1.91) | 4.81 (4.60, 5.02) | 6.31 (6.02, 6.60) |
| MCS | Black Caribbean/Black African | 1 | 1.40 (0.88, 1.92) | 1.64 (1.17, 2.12) | 4.92 (4.43, 5.41) | 6.37 (5.70, 7.04) |
| MCS | Black Caribbean/Black African | 2 | 1.52 (1.12, 1.93) | 1.55 (1.05, 2.05) | 4.99 (4.53, 5.45) | 6.02 (5.48, 6.55) |
| MCS | Black Caribbean/Black African | 3 | 2.11 (1.72, 2.49) | 2.14 (1.69, 2.60) | 5.07 (4.73, 5.41) | 6.06 (5.56, 6.57) |
| MCS | Other | 1 | 1.98 (1.35, 2.61) | 1.62 (0.95, 2.29) | 5.25 (4.62, 5.88) | 5.99 (5.15, 6.82) |
| MCS | Other | 2 | 1.78 (1.33, 2.24) | 1.65 (1.07, 2.22) | 5.24 (4.70, 5.78) | 6.42 (5.77, 7.07) |
| MCS | Other | 3 | 1.82 (1.41, 2.23) | 1.80 (1.42, 2.18) | 4.87 (4.49, 5.25) | 5.92 (5.33, 6.50) |

*Note.* Adjusted models included birth sex, highest qualification achieved, pre-pandemic self-reported health, pre-pandemic psychological distress, and household composition as covariates. BCS: British Cohort Study, 1970 birth cohort; GAD-2: 2-item General Anxiety Disorder questionnaire; MCS: Millennium Cohort Study, 2000 birth cohort; NCDS: National Child and Development Study, 1958 birth cohort; NS: Next Steps, 1990 cohort; NSHD: National Survey of Health and Development, 1946 birth cohort; ONS: UK Office for National Statistics; PHQ-2: 2-item Patient Health Questionnaire; UCLA-3: 3-item UCLA loneliness scale. Survey wave 1: May 2020; survey wave 2: September/October 2020; survey wave 3: February/March 2021.

#### Figure S12.1. Unadjusted and adjusted (by birth sex, highest qualification achieved, pre-pandemic self-reported health, pre-pandemic psychological distress, and household composition) anxiety symptomatology (GAD-2) marginal mean estimates and 95% confidence intervals by ethnicity.

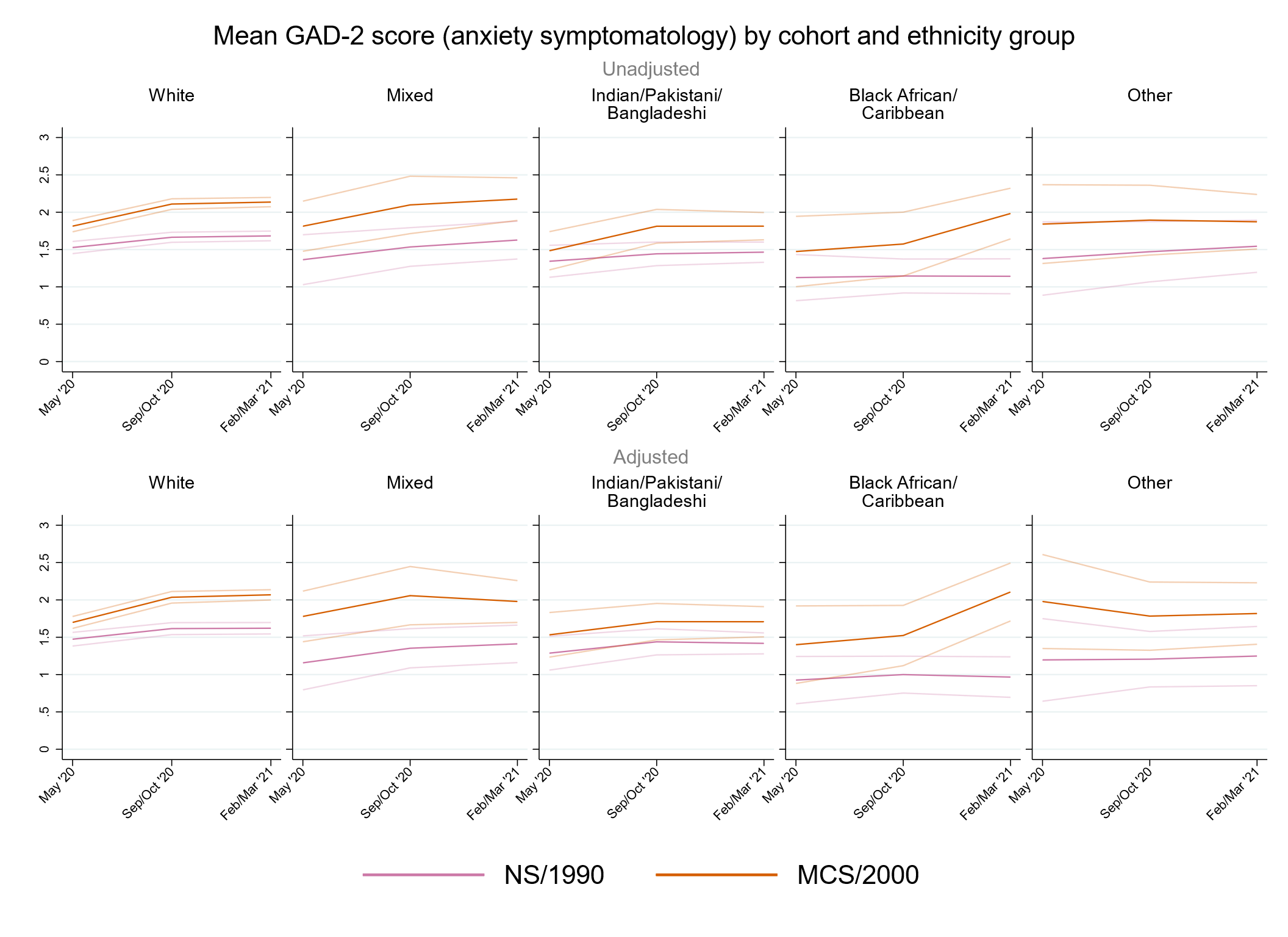

#### Figure S12.2. Unadjusted and adjusted (by birth sex, highest qualification achieved, pre-pandemic self-reported health, pre-pandemic psychological distress, and household composition) depressive symptomatology (PHQ-2) marginal mean estimates and 95% confidence intervals by ethnicity.

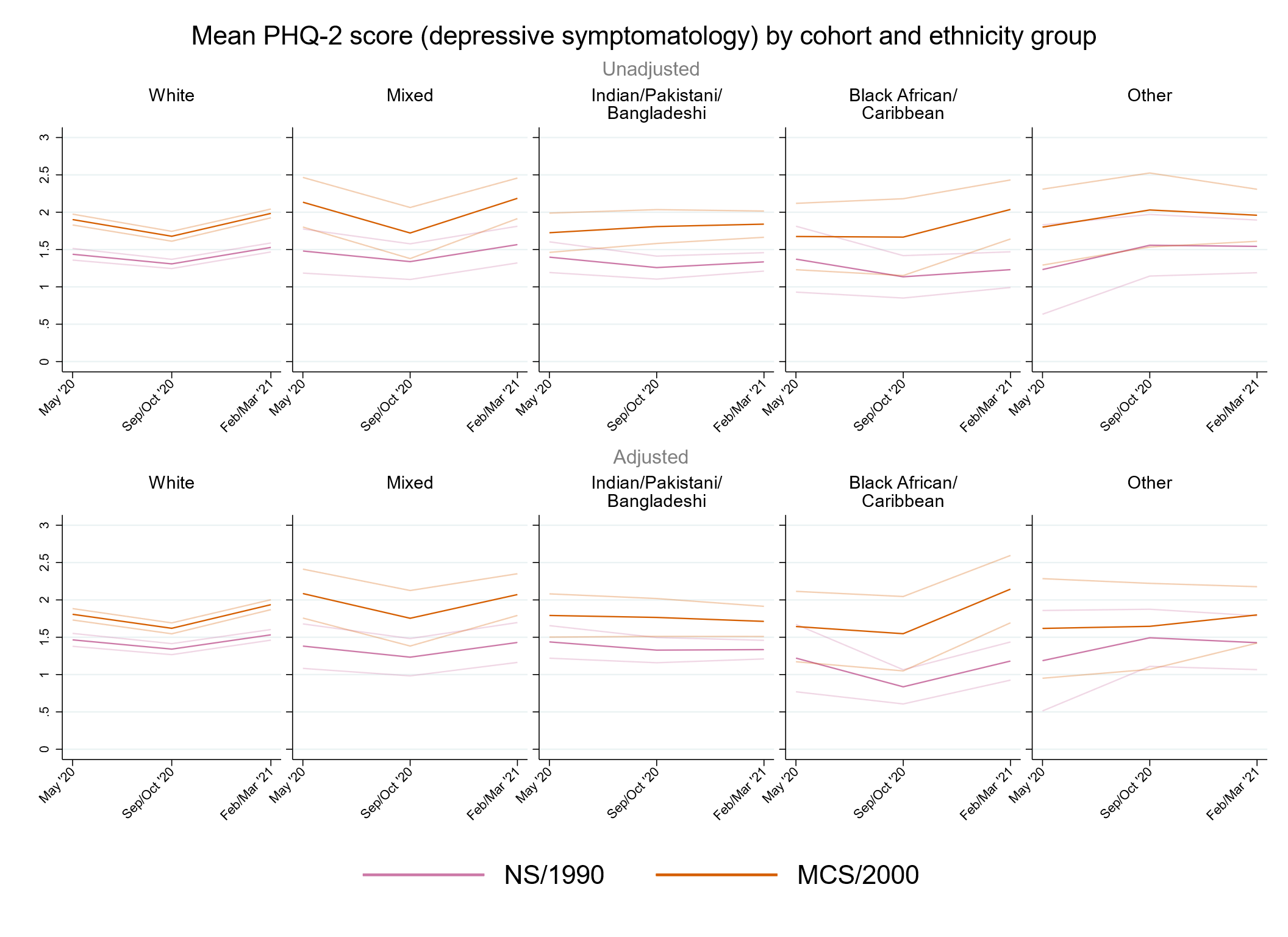

#### Figure S12.3. Unadjusted and adjusted (by birth sex, highest qualification achieved, pre-pandemic self-reported health, pre-pandemic psychological distress, and household composition) loneliness (UCLA-3) marginal mean estimates and 95% confidence intervals by ethnicity.

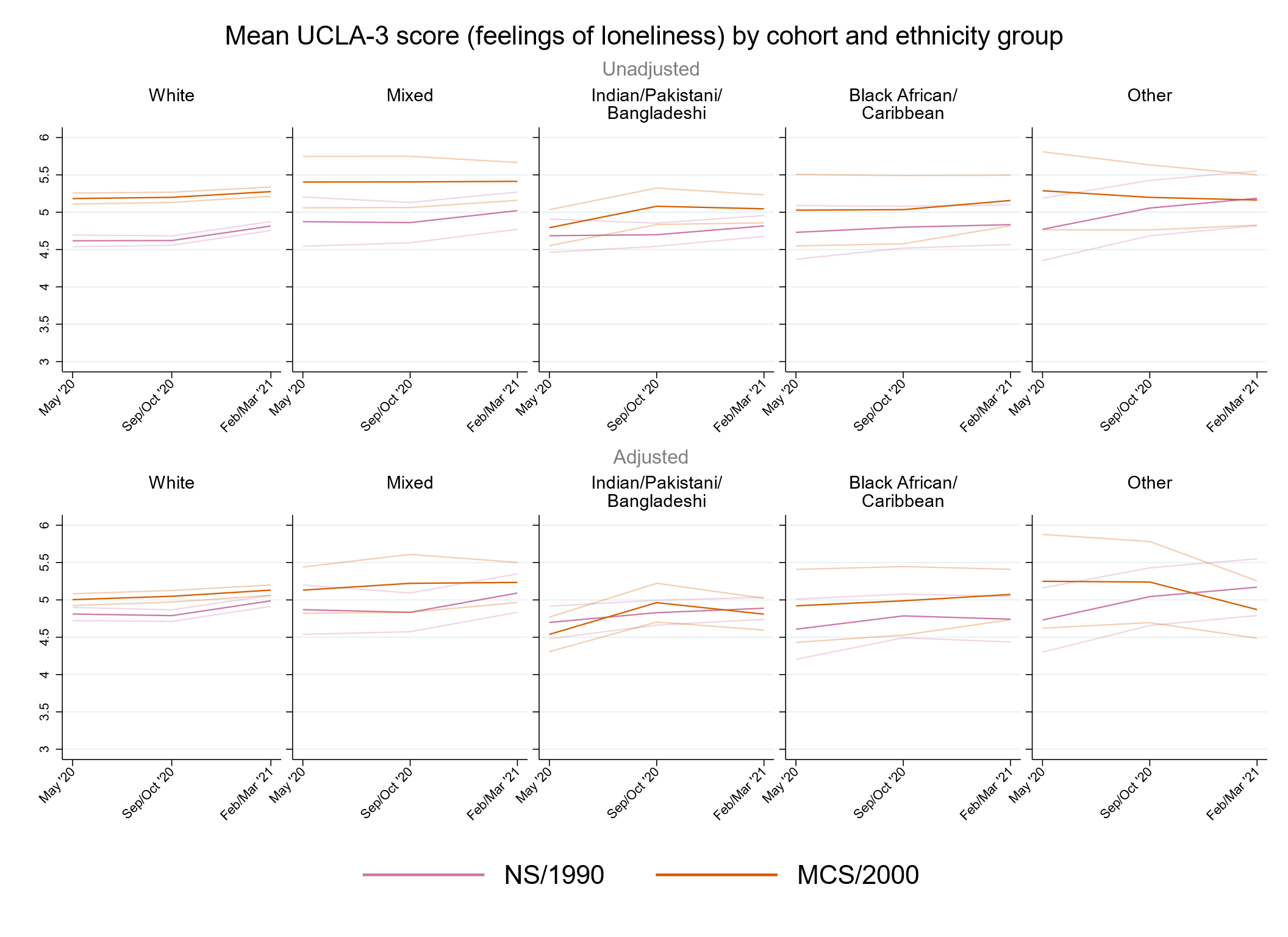

#### Figure S12.4. Unadjusted and adjusted (by birth sex, highest qualification achieved, pre-pandemic self-reported health, pre-pandemic psychological distress, and household composition) life satisfaction marginal mean estimates and 95% confidence intervals by ethnicity.

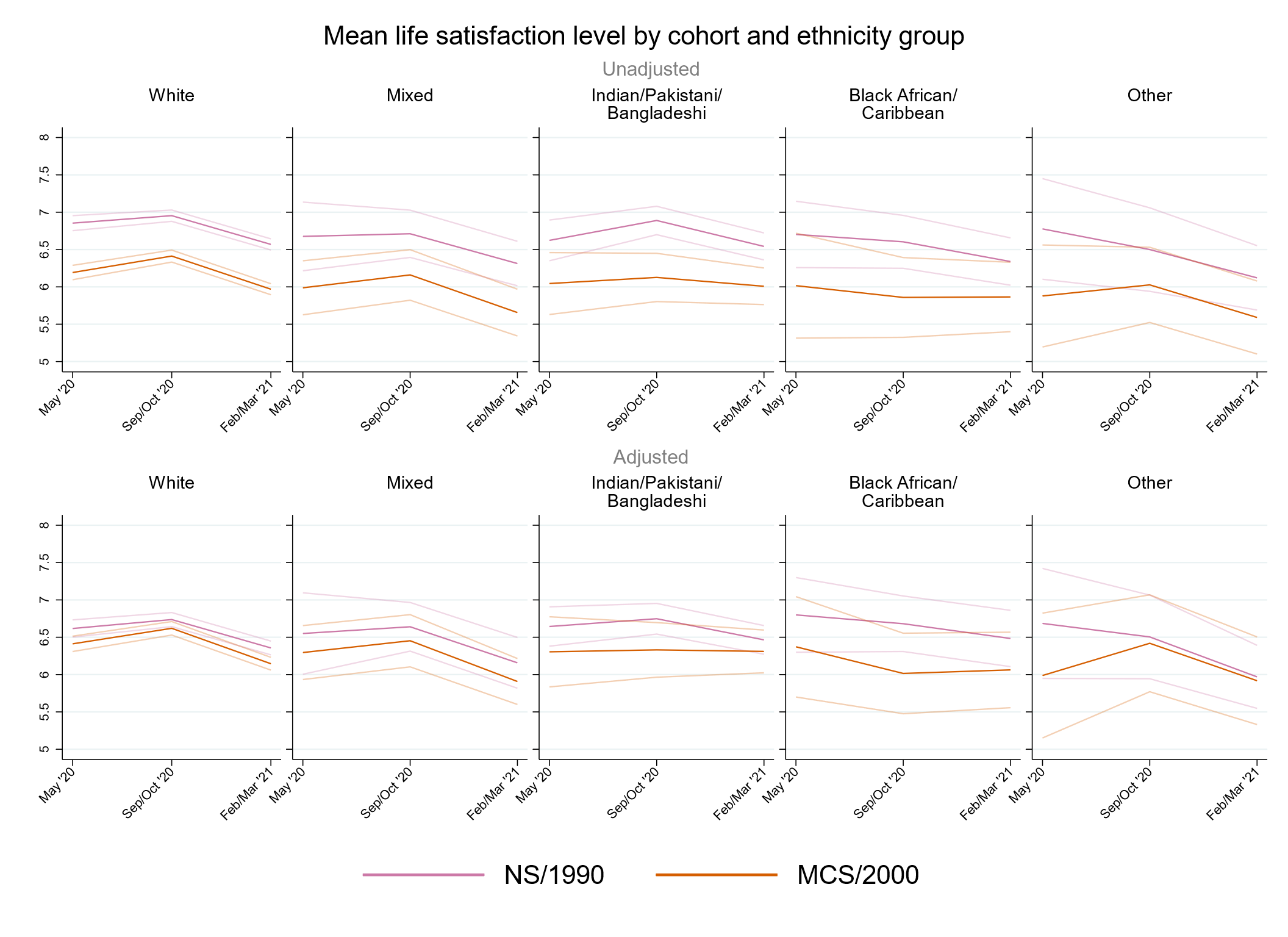

### Supplementary References

1.	Brown M, Goodman A, Peters A, et al. *COVID-19 Survey in Five National Longitudinal Studies: Waves 1, 2 and 3 User Guide (Version 3).* UCL Centre for Longitudinal Studies and MRC Unit for Lifelong Health and Ageing; 2021.

2.	Dodgeon B, Parsons S. *Deriving highest qualification in NCDS and BCS70*. Centre for Longitudinal Studies, Institute of Education; 2011.

3.	McElroy E, Richards M, Fitzsimons E, et al. *Feasibility of retrospectively harmonising cognitive measures in five British birth cohort studies*. CLOSER; 2021.

4.	Dodgeon B, Morris T, Crawford C, et al. *CLOSER work package 2: Harmonised socio-economic measures user guide (revised)*. CLOSER; 2011.
